# Mapping the human genetic architecture of COVID-19 by worldwide meta-analysis

**DOI:** 10.1101/2021.03.10.21252820

**Authors:** The COVID-19 Host Genetics Initiative, Andrea Ganna

## Abstract

The genetic makeup of an individual contributes to susceptibility and response to viral infection. While environmental, clinical and social factors play a role in exposure to SARS-CoV-2 and COVID-19 disease severity, host genetics may also be important. Identifying host-specific genetic factors indicate biological mechanisms of therapeutic relevance and clarify causal relationships of modifiable environmental risk factors for SARS-CoV-2 infection and outcomes. We formed a global network of researchers to investigate the role of human genetics in SARS-COV-2 infection and COVID-19 severity. We describe the results of three genome-wide association meta-analyses comprising up to 49,562 COVID-19 patients from 46 studies across 19 countries worldwide. We reported 13 genome-wide significant loci that are associated with SARS-CoV-2 infection or severe manifestations of COVID-19. Several of these loci correspond to previously documented associations to lung or autoimmune and inflammatory diseases. They also represent potentially actionable mechanisms in response to infection. We further identified smoking and body mass index as causal risk factors for severe COVID-19. The identification of novel host genetic factors associated with COVID-19, with unprecedented speed, was enabled by prioritization of shared resources and analytical frameworks. This working model of international collaboration provides a blue-print for future genetic discoveries in the event of pandemics or for any complex human disease.

## INTRODUCTION

The coronavirus disease 2019 (COVID-19) pandemic, caused by infections with severe acute respiratory syndrome coronavirus 2 (SARS-CoV-2), has resulted in enormous health and economic burden worldwide. One of the most remarkable features of SARS-CoV-2 infection is that a large proportion of individuals ^1^ are asymptomatic while others experience progressive, even life-threatening, viral pneumonia and acute respiratory distress syndrome. While established host factors contribute to disease severity (e.g., increasing age, male gender, and higher body mass index ^2^), these risk factors alone do not explain all variability in disease severity observed among individuals.

The contribution of host genetics to susceptibility and severity of infectious disease, including respiratory viruses, is well-documented, and encompasses rare inborn errors of immunity ^3,4^ as well as common genetic variation ^5–10^. Characterizing which genetic factors contribute to COVID-19 susceptibility and severity may uncover novel biological insights into disease pathogenesis and identify mechanistic targets for therapeutic development or drug repurposing, as treating the disease remains a highly important goal despite the recent development of vaccines. For example, rare loss-of-function variants in genes involved in type I interferon (*IFN*) response may be involved in severe forms of COVID-19 ^11–14^. At the same time, several genome-wide association studies (GWAS) that investigate the contribution of common genetic variation ^15–18^ to COVID-19 have provided support for the involvement of several genomic loci associated with COVID-19 severity and susceptibility, with the strongest and most robust finding for severity being at locus 3p21.31^15–19^. However, much remains unknown about the genetic basis of susceptibility to SARS-CoV-2 and severity of COVID-19.

The COVID-19 Host Genetics Initiative (COVID-19 HGI) (https://www.covid19hg.org/) ^20^ is an international, open-science collaboration to share scientific methods and resources with research groups across the world with the goal to robustly map the host genetic determinants of SARS-CoV-2 infection and severity of the resulting COVID-19 disease. We have carefully aligned phenotype definitions and incorporated variable ascertainment strategies to achieve greater statistical confidence in our results. We openly and continuously share updated results to the research community. Here, we report the latest results of meta-analyses of 46 studies from 19 countries (**Fig. 1**) for COVID-19 host genetic effects.

**Figure 1.**
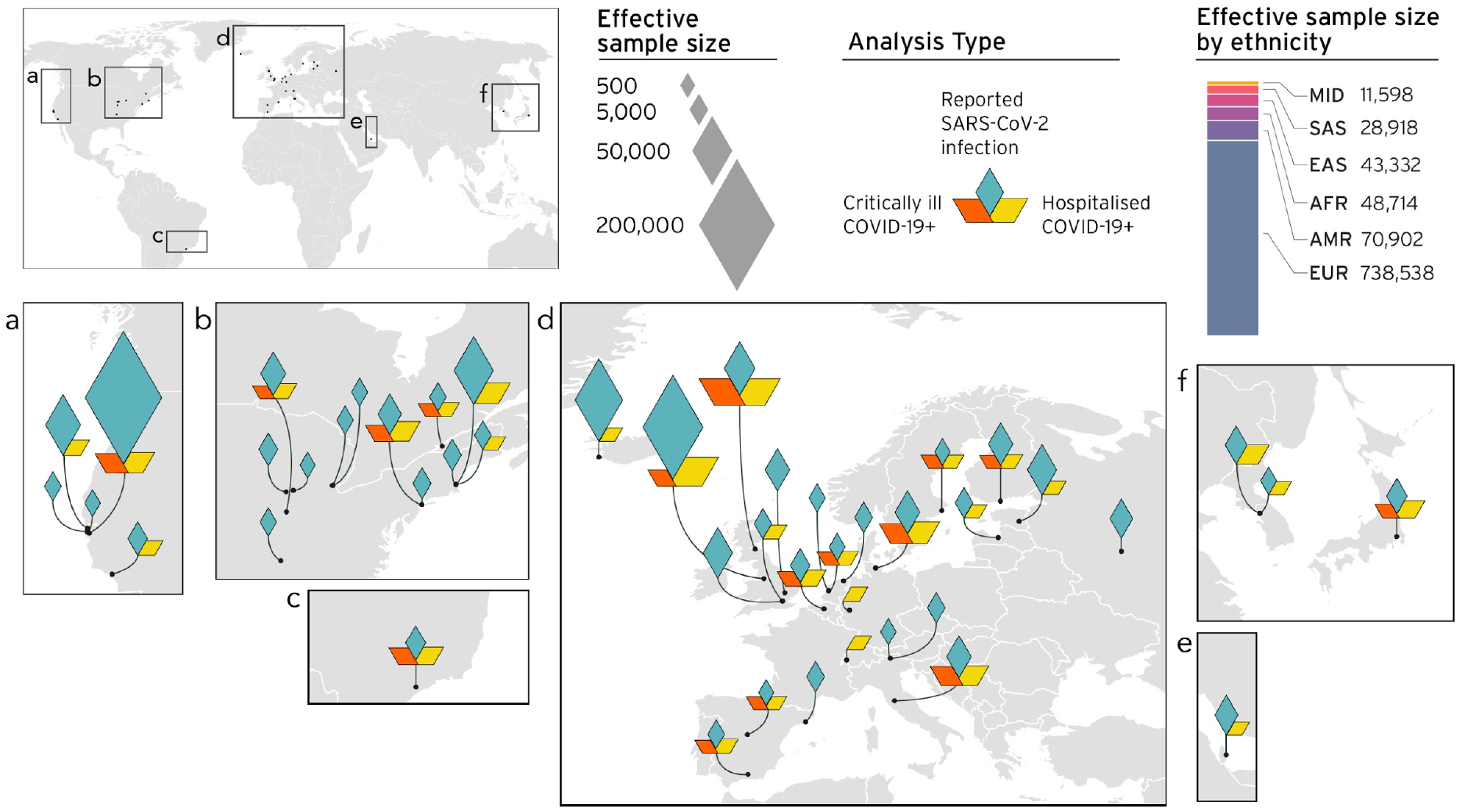
Geographical overview of the contributing studies to the COVID-19 HGI and composition by major ancestry groups. Middle Eastern (MID), South Asian (SAS), East Asian (EAS), African (AFR), Admixed American (AMR), European (EUR).

## RESULTS

### Worldwide meta-analyses of COVID-19

Overall, the COVID-19 Host Genetics Initiative combined genetic data for up to 49,562 cases and two million controls across 46 distinct studies (**Fig. 1**). The data included studies from populations of different genetic ancestries, including European, Admixed American, African, Middle Eastern, South Asian and East Asian individuals (**Supplementary Table 1**). An overview of the study design is provided in **Extended Data Figure 1**. We performed case-control meta-analyses in three main categories of COVID-19 disease according to predefined and partially overlapping phenotypic criteria. These were (1) critically ill COVID-19 cases defined as those who required respiratory support in hospital or who were deceased due to the disease, (2) cases with moderate or severe COVID-19 defined as those hospitalized due to symptoms associated with the infection, and (3) all cases with reported SARS-CoV-2 infection with or without symptoms of any severity (**Methods**). Controls for all three analyses were selected as genetically ancestry-matched samples without known SARS-CoV-2 infection, if that information was available (**Methods**). Each individual study that contributed data to a particular analysis met a minimum threshold of 50 cases, as defined by the aforementioned phenotypic criteria, for statistical robustness. Where more detailed demographic data was available, the average age of COVID-19 cases was 55 years (**Supplementary Table 1**). We report quantile-quantile plots and ancestry principal component plots for contributing studies in **Extended Data Figures 2** and **3**.

Across our three analyses, we reported a total of 13 independent genome-wide significant loci associated with COVID-19 (*P* < 1.67 × 10^−8^; threshold adjusted for multiple trait testing) (**Table 1, Supplementary Table 2**), most of which were shared between two or more COVID-19 phenotypes. Two of these loci are in very close proximity within the 3p21.31 region, which was previously reported as one single locus associated with COVID-19 severity ^15–19^ (**Extended Data Figure 5**). Overall, we reported six genome-wide significant associations for critical illness due to COVID-19, using data for 6,179 cases and 1,483,780 controls from 16 studies (**Extended Data Figure 4**). Nine genome-wide significant loci were detected for moderate to severe hospitalized COVID-19, from an analysis of 13,641 COVID-19 cases and 2,070,709 controls, across 29 studies (**Fig. 2 top panel**). Finally, seven loci reached genome-wide significance in the analysis using data for all available 49,562 reported cases of SARS-CoV-2 infection and 1,770,206 controls, using data from a total of 44 studies (**Fig. 2 bottom panel**). The proportion of cases with non-European genetic ancestry for each of the three analyses was 23%, 29% and 22%, respectively.

**Figure 2.**
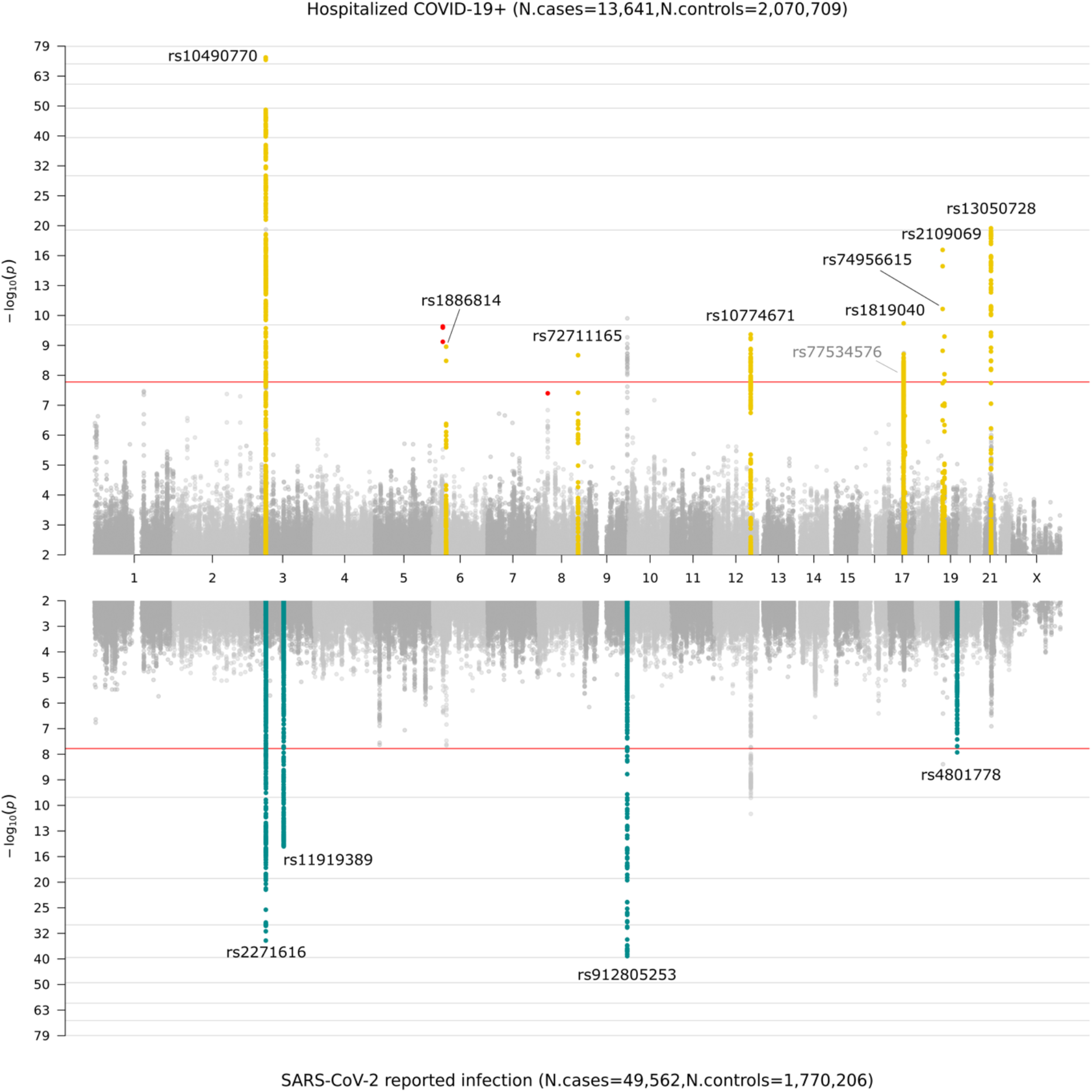
Genome-wide association results for COVID-19. Top panel shows results of genome-wide association study of hospitalized COVID-19 and controls, and bottom panel the results of reported SARS-CoV-2 infection and controls. Loci highlighted in yellow (top panel) represent regions associated with severity of COVID-19 manifestation i.e. increasing odds for more severe COVID-19 phenotypes, where loci highlighted in green (bottom panel) are regions associated with reported SARS-CoV-2 infection, i.e. the effect is the same across mild and severe COVID-19 phenotypes. A window of +/- 500kb region from the lead variant (**Table 1**) was used to highlight these loci. We highlight only loci that were specific to each analysis. That is, some genome-wide significant loci are not colored in yellow or green if they are not specific to severity (top panel) or susceptibility (bottom panel). Variants highlighted in red represent genome-wide significant variants that had high heterogeneity across studies that contributed data, and which were therefore not considered in the final analyses. Note that top panel regions on chromosomes 17 and 19 contain two statistically independent loci within proximity of each other, and on the bottom panel the first signal on chromosome 3 contains two peaks within the same region. The locus tagged by lead variant rs77534576 on chromosome 17 was discovered in the critically ill COVID-19 analysis (**see Table 1**).

**Table 1:**
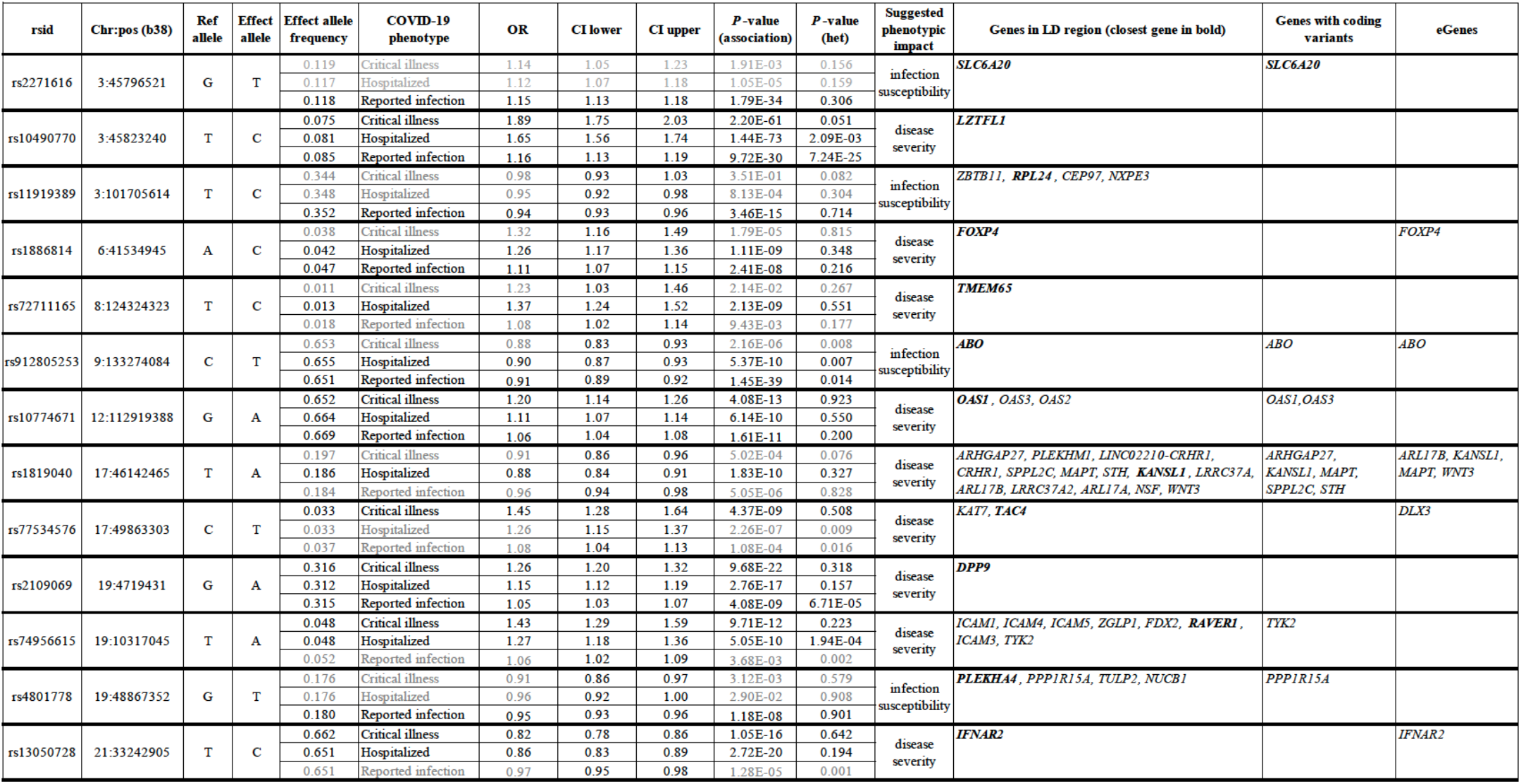
Genome-wide significant results for each worldwide meta-analysis. Meta-analysis results that reached a genome-wide significance threshold of P < 1.67 × 10^−8^ (adjusted for multiple testing for three phenotypes) are coloured in black; associations not reaching significance are coloured in grey; association was not performed for variants that were missing in more than ⅔ of studies contributing to the meta-analysis. Effect allele frequency is the sample size weighted frequency across studies included in each meta-analysis. P-value is reported for meta-analysis variant association with trait (P-value association) and heterogeneity (P-value het) between studies included in each meta-analysis. Suggested phenotypic impact of the locus was inferred using a test comparing variant effects across analyses (see **Methods**). Closest gene: A gene with a minimum distance from each lead variant to gene body. Genes in linkage disequilibrium (LD) region: Genes that overlap with a genomic range that contains any variants in LD (r^2^ > 0.6) with each lead variant. Genes with coding variants: Genes with a loss-of-function or missense variant in LD with a lead variant (r^2^ > 0.6). eGenes: Genes with a fine-mapped cis-eQTL variant (PIP > 0.1) in GTEx Lung that is in LD with a lead variant (r^2^ > 0.6).

### Comparison of effect for genome-wide significant results across studies and phenotype definitions

We found no genome-wide significant sex-specific effects at the 13 loci. However, we did identify significant heterogeneous effects (*P* <0.003) across studies for 3 out of the 13 loci, likely reflecting heterogeneous ascertainment of cases across studies contributing data to these analyses (**Table 1**). There was minor sample overlap (*n* = 8,380 EUR; *n* = 745 EAS) between controls from the genOMICC and the UK Biobank studies, but leave-one-out sensitivity analyses did not reveal any bias in the corresponding effect sizes or *P*-values (**Extended Data Fig. 6**).

We next wanted to better understand whether the 13 significant loci were acting through mechanisms increasing susceptibility to infection or by affecting the progression of symptoms towards more severe disease. For all 13 loci, we compared the lead variant (strongest association *P*-value) odds ratios (ORs) for the risk-increasing allele across our different COVID-19 phenotype definitions.

We first noted that four loci had consistent ORs between the two larger and better powered analyses; all cases with reported infection and all cases hospitalized due to COVID-19 (**Methods**) (**Table 1, Supplementary Table 2**). Such consistency implied that these four loci were likely associated with overall susceptibility to SARS-CoV-2 infection, but not with the progression to more severe COVID-19 phenotypes. These susceptibility loci included the previously reported *ABO* locus ^15,16,18,19^. The lead variant rs912805253 at this locus reached genome-wide significance in both our reported infection (OR [95%CI] = 0.90 [0.89, 0.92]; *P* =1.4 × 10^−39^) and hospitalized COVID-19 (OR [95%CI] = 0.90 [0.87, 0.93]; *P* =5.4 × 10^−10^) analyses, but the odds of becoming hospitalized were no different from the odds of becoming infected when carrying this allele. Interestingly, and in agreement with the report by Robert and colleagues ^18^, we reported a locus within the 3p21.31 region that was more strongly associated with susceptibility to SARS-CoV-2 than progression to more severe COVID-19 phenotypes. Rs2271616 showed a stronger association with reported infection (*P*=1.79×10^−34^; OR[95%CI]= 1.15 [1.13-1.18]) than hospitalization (*P*=1.05×10^−5^; OR[95%CI]=1.12[1.06-1.19]). For this locus, which contains additional independent signals, the linkage-disequilibrium pattern is discordant with the *P*-value expectation (**Supplementary Notes**), pointing to a key missing causal variant or to a potentially complicated structural variation in this locus. More research into understanding the biology of these signals is needed.

In contrast, nine out of the 13 loci were associated with increased risk of severe symptoms with significantly larger ORs for hospitalized COVID-19 compared to the mildest phenotype of reported infection (eight loci below threshold *P* <0.003 (0.05/15) test for effect size difference, and additionally lead variant rs10774671 had a clear increase in ORs despite not passing this threshold) (**Table 1, Supplementary Table 2)**. We further compared the ORs for these nine loci for critical illness due to COVID-19 *vs*. hospitalized due to COVID-19, and found that these loci exhibited a general increase in effect risk for critical illness (**Methods**) (**Extended Data Fig. 7A, Supplementary Table 3)**, but we note that due to the lower power for association in the critically ill analysis these results should be considered as suggestive. Overall, these results indicated that these nine loci were more likely associated with progression of the disease and worse outcome from SARS-CoV-2 infection compared to being associated with susceptibility to SARS-CoV-2 infection. We noted that two loci, tagged by lead variants rs1886814 and rs72711165, had higher allele frequencies in South East Asian (rs1886814, 15%) and East Asian genetic ancestry (rs72711165, 8%) whilst the minor allele frequencies in European populations were < 3%. This highlights the value of including data from diverse populations for genetic discovery, but challenges over inter-cohort heterogeneity and lack of appropriate LD reference for multi-ancestry analysis remain, therefore hindering proper fine-mapping and conditional analysis in these data.

Our phenotype definitions include population controls without known SARS-CoV-2 infection. This is not an optimal control group because some individuals, if exposed to SARS-CoV-2 could develop a severe form of COVID-19 disease and should be classified as cases. To better understand the effect of such potential misclassification, we conducted a new meta-analysis, including only the studies that compared hospitalized COVID-19 cases with controls with laboratory-confirmed SARS-CoV-2 infection but who had mild symptoms or were asymptomatic (*n* = 5,773 cases and *n* = 15,497 controls). We then compared the effect sizes obtained from this analysis with those from the main phenotype definition (hospitalized cases *vs*. controls without known SARS-CoV-2 infection, if that information was available) using only studies that reported results for both analyses (**Methods**). We found that across the nine loci that had reached genome-wide significance in our main hospitalized COVID-19 analysis, the ORs were not significantly different in the analysis with better refined controls (**Extended Data Fig. 7B, Supplementary Table 3**). These results indicate that using population controls can be a valid and powerful strategy for host genetic discovery of infectious disease.

### Gene prioritization and association with other diseases or traits

To better understand the potential biological mechanism of each locus, we applied several approaches to prioritize candidate causal genes and explore additional associations with other complex diseases and traits. For gene prioritization, we first identified genes within each COVID-19 associated region by distance or linkage disequilibrium (LD) to a lead variant, and then prioritized those with protein-alternating variants, lung eQTLs, or having the highest prioritization score in the OpenTargets V2G (Variant-to-Gene) algorithm ^21^ (see **Methods, Supplementary Tables 2 and 4**). For reporting PheWAS associations (**Supplementary Table 5**), we only considered phenotypes for which the lead variants were in high LD (*r*^2^ > 0.8) with the 13 genome-wide significant lead variants from our main COVID-19 meta-analysis. This conservative approach allowed spurious signals primarily driven by proximity rather than actual colocalization to be removed (see **Methods**).

Of the 13 genome-wide significant loci, we found nine loci to have a distinct candidate gene(s), including biologically plausible genes (**Table 1**). Protein-altering variants in LD with lead variants implicated genes at six loci, including *TYK2* (19p13.2) and *PPP1R15A* (19q13.33). The COVID-19 lead variant rs74956615:T>A in *TYK2*, which confers risk for critical illness (OR[95%] = 1.43 [1.29, 1.59]; *P* = 9.71 × 10^−12^) and hospitalization (OR [95%CI] = 1.27 [1.18, 1.36]; *P* = 5.05 × 10^−10^) due to COVID-19, is correlated with the missense variant rs34536443:G>C (p.Pro1104Ala; *r*^2^ = 0.82). This is consistent with the primary immunodeficiency described with complete *TYK2* loss of function ^22^. In contrast, this missense variant was previously reported to be protective against autoimmune diseases, including rheumatoid arthritis (OR = 0.74; *P* = 3.0 × 10^−8^; UKB SAIGE), and hypothyroidism (OR = 0.84; *P* = 1.8 × 10^−10^; UK Biobank) (**Fig. 3**). An additional independent missense variant rs2304256:C>A (p.Val362Phe; *r*^2^ = 0.08 with rs34536443) in *TYK2* was also associated with critical illness (OR [95%] = 1.17 [1.11, 1.23]; *P* = 2.4 × 10^−10^). At the 19q13.33 locus, the lead variant rs4801778, that was significantly associated with reported infection (OR [95%CI] = 0.95 [0.93, 0.96]; *P* = 2.1 × 10^−8^), is in LD (*r*^2^ = 0.93) with a missense variant rs11541192:G>A (p.Gly312Ser) in *PPP1R15A*. Further evidence for this missense variant shows significant association with increased reticulocyte fraction and strong correlation with a reticulocyte lead variant rs56104184:C>T (OR [95%CI] = 1.03 [1.02, 1.04]; *P* = 4.1 × 10^−13^)^23^.

**Figure 3.**
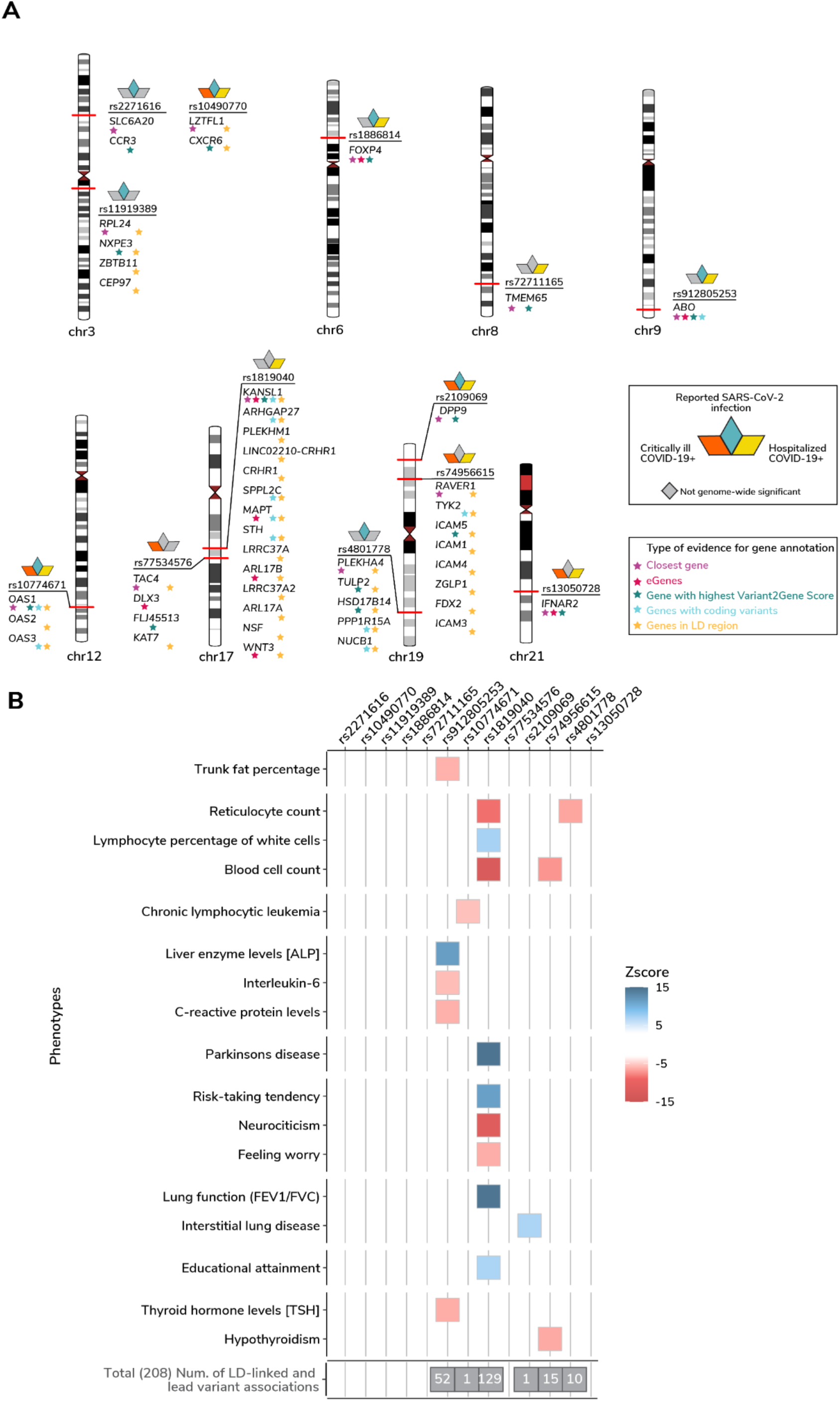
Gene prioritization and PheWas. **A)** Gene prioritization using different evidence measures of gene annotation. The ideogram shows the position of the COVID-19 index SNPs and associated genes with type of gene annotation highlighted as stars. The squares above the SNP rsid represent COVID-19 sub-phenotypes for which the SNP is genome-wide significant. Closest gene: A gene with a minimum distance from each lead variant to gene body. Genes in linkage disequilibrium (LD) region: Genes that overlap with a genomic range that contains any variants in LD (r^2^ > 0.6) with each lead variant. Genes with coding variants: Genes with a loss-of-function or missense variant in LD with a lead variant (r^2^ > 0.6). eGenes: Genes with a fine-mapped cis-eQTL variant (PIP > 0.1) in GTEx Lung that is in LD with a lead variant (r^2^> 0.6) (see **Supplementary Table 4** for a complete list). V2G: Highest gene prioritized by OpenTargetGenetics’ V2G score. **B)** Selected phenotypes associated with genome-wide significant COVID-19 variants (see **Supplementary Table 5** for a complete list). We report those associations for which a lead variant from a prior GWAS results was in high LD (r^2^ > 0.8) with the index COVID-19 variants. The colour represents the Z-scores of correlated risk increasing alleles for the trait. The total number of associations for each COVID-19 variant is highlighted in the grey box.

Lung-specific *cis*-eQTL from GTEx v8 ^24^ (*n* = 515) and the Lung eQTL Consortium ^25^ (*n* = 1,103) provided further support for a subset of loci, including *FOXP4* (6p21.1) and *ABO* (9q34.2), *OAS1*/*OAS3*/*OAS2* (12q24.13), and *IFNAR2/IL10RB* (21q22.11), where the COVID-19 associated variants modifies gene expression in lung. Furthermore, our PheWAS analysis implicated three additional loci related to lung function, with modest lung eQTL evidence, i.e. the lead variant was not fine-mapped but significantly associated. An intronic variant rs2109069:G>A in *DPP9* (19p13.3), positively associated with critical illness, was previously reported to be risk-increasing for interstitial lung disease (tag lead variant rs12610495:A>G [p.Leu8Pro], OR = 1.29, *P* =2.0 × 10^−12^) ^26^. The COVID-19 lead variant rs1886814:A>C in *FOXP4* locus is modestly LD-linked (*r*^2^ = 0.64) with a lead variant of lung adenocarcinoma (tag variant=rs7741164; OR=1.2, P=6.0 × 10^−13^) ^27^. We also found that intronic variants (1q22) and rs1819040:T>A in *KANSL1* (17q21.31), associated protectively against hospitalization due to COVID-19, were previously reported for reduced lung function (e.g. tag lead variant rs141942982:G>T, OR [95%CI] = 0.96 [0.95, 0.97], *P* = 1.00 × 10^−20^) ^28^. Notably, the 17q21.31 locus is a well-known locus for structural variants containing a megabase inversion polymorphism (H1 and inverted H2 forms) and complex copy-number variations, where the inverted H2 forms were shown to be positively selected in Europeans ^29,30^.

Lastly, there are two loci in the 3p21.31 region with varying genes prioritized by different methods for different independent signals. For the severity lead variant rs10490770:T>C, we prioritized *CXCR6* with the Variant2Gene (V2G) algorithm ^21^, while *LZTFL1* is the closest gene. The *CXCR6* plays a role in chemokine signaling ^31^, and *LZTFL1* has been implicated in lung cancer ^32^. Rs2271616:G>T, associated with susceptibility, tags a complex region including several independent signals (**Supplementary Notes**) all located within a gene body of *SLC6A20* which is known to functionally interact with the SARS-CoV-2 receptor ACE2 ^33^. However, none of the lead variants in the 3p21.31 region has been previously associated with other traits or diseases in our PheWAS analysis. While these results provide supporting *in-silico* evidence for candidate causal gene prioritization, further functional characterization is strongly needed. Detailed locus descriptions and LocusZoom plots are provided in **Extended Data Fig. 8**.

### Polygenic architecture of COVID-19

To further investigate the genetic architecture of COVID-19, we used results from meta-analyses including only European ancestry samples (sample sizes described in **Methods and Supplementary Table 1**). We applied linkage disequilibrium (LD) score regression ^34^ to the summary statistics to estimate SNP heritability, i.e. proportion of variation in the two phenotypes that was attributable to common genetic variants, and to determine whether heritability for COVID-19 phenotypes was enriched in genes specifically expressed in certain tissues ^35^ from GTEx dataset ^36^. We detected a low, but significant heritability across all three analyses (<1% on observed scale, all *P*-values < 0.0001, LDSC intercept range 1.0024-1.0137; **Supplementary Table 6**). Despite these low values, which interpretation is complicated by the use of population controls and variation in the disease prevalence estimates, we found that heritability for reported infection was significantly enriched in genes specifically expressed in the lung (*P* = 5.0 × 10^−4^) (**Supplementary Table 7**). These findings, together with genome-wide significant loci identified in the meta-analyses, illustrate that there is a significant polygenic or oligogenic architecture that can be better leveraged with future, larger, sample sizes.

### Genetic correlation and causal relationship between COVID-19 and other traits

Genetic correlations (*rg*) between the three COVID-19 phenotypes was high, though lower correlations were observed between hospitalized COVID-19 and reported infection (critical illness *vs*. hospitalized: *rg* [95%CI] = 1.37 [1.08, 1.65], *P* = 2.9 × 10^−21^; critical illness *vs*. reported infection, *rg* [95%CI] = 0.96 [0.71, 1.20], *P* = 1.1 × 10^−14^; hospitalized *vs*. reported infection: *rg* [95%CI] = 0.85 [0.68, 1.02], *P* = 1.1 × 10^−22^). To better understand which traits are genetically correlated and/or potentially causally associated with COVID-19 hospitalization, critical illness and SARS-CoV-2 reported infection, we chose a set of 38 disease, health and neuropsychiatric phenotypes as potential COVID-19 risk factors based on their putative relevance to the disease susceptibility, severity, or mortality (**Extended Data Figure 9, Supplementary Table 8**).

We found evidence (FDR<0.05) of significant genetic correlations between 9 traits and hospitalized COVID-19 and SARS-CoV-2 reported infection (**Fig. 4; Supplementary Table 9**). Genetic correlation results for COVID-19 hospitalization and critical illness partially overlap with reported SARS-CoV-2 infection, with genetic liability to BMI, type 2 diabetes, smoking, and attention deficit hyperactivity disorder showing significant positive correlations (*rg* range between 0.16 - 0.26). However some results were significantly different between COVID-19 hospitalization and critical illness versus reported infection. For example, genetic liability to ischemic stroke, was only significantly positively correlated with critical illness or hospitalization due to COVID-19, but not with a higher likelihood of reported SARS-CoV-2 infection (infection *r g*= 0.019 *vs*. hospitalization *rg* = 0.41, z = 2.7, *P* = 0.006; infection *rg* = 0.019 *vs*. critical illness *rg* = 0.40, z = 2.49, *P* = 0.013). In addition, coronary artery disease, and systemic lupus erythematosus showed positive genetic correlations with critical illness or hospitalization due to COVID-19. Genetic liability to risk tolerance, on the hand, was the only trait specifically associated with SARS-CoV-2 infection. This potentially reflects that risk taking behavior could be associated with a higher chance of infection, but is not, *per-se*, impacting the chances to develop a severe form of COVID-19. With improved phenotyping of cases and controls, methods to deconvolute the effects specific to SARS-CoV-2 infection - a proxy for disease susceptibility - and those specific for progression to severe disease can be applied to better interpret these results.

**Figure 4.**
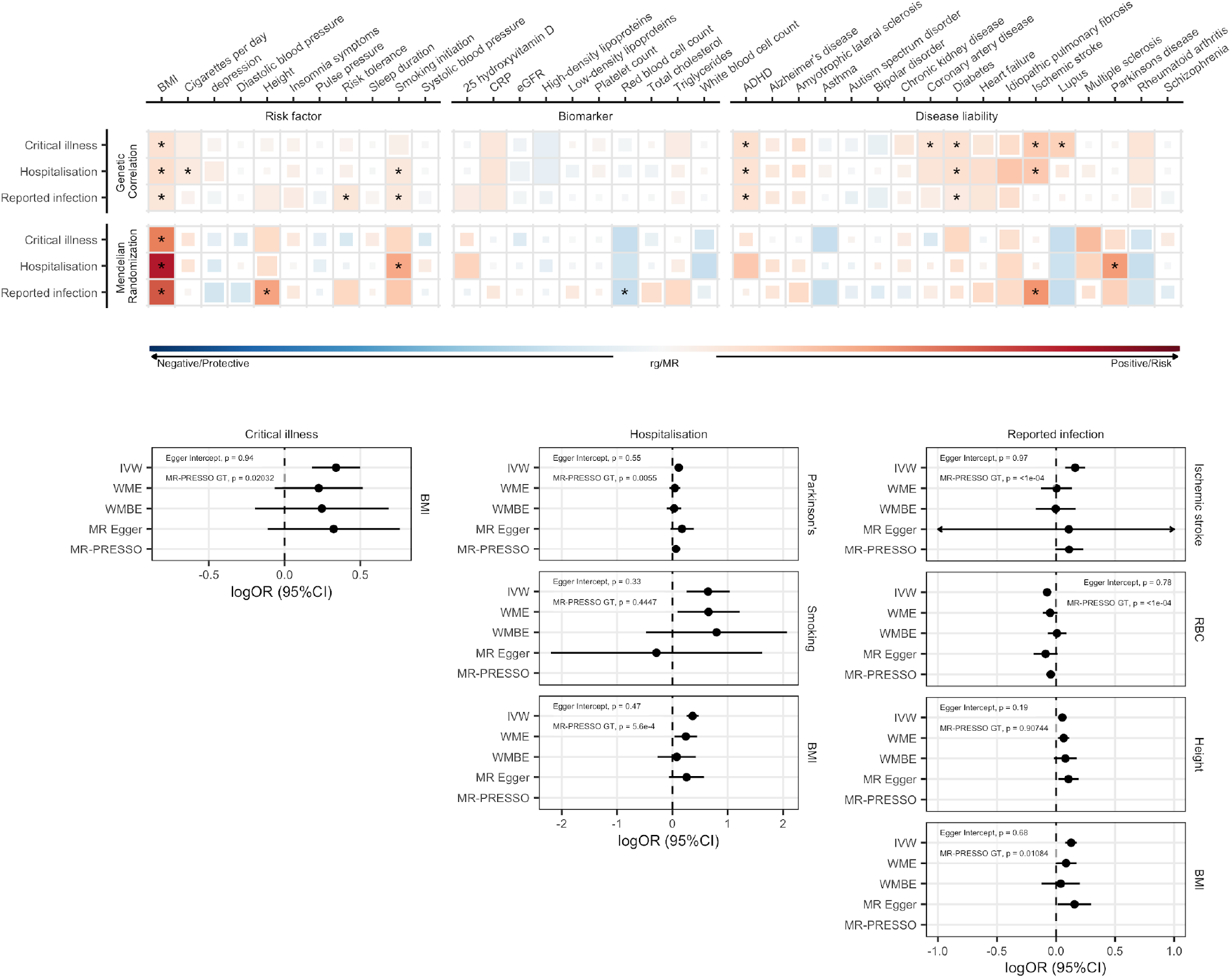
Genetic correlations and Mendelian randomization causal estimates between 38 traits and COVID-19 hospitalization, critical illness and SARS-CoV-2 reported infection. Blue, negative genetic correlation and protective Mendelian randomization (MR) causal estimates; red, positive genetic correlation and risk MR causal estimates. Larger squares correspond to more significant P values, with genetic correlations or MR causal estimates significantly different from zero at a P < 0.05 shown as a full-sized square. Genetic correlations or causal estimates that are significantly different from zero at a false discovery rate (FDR) of 5% are marked with an asterisk. Forest plots display the causal estimates for each of the sensitivity analyses used in the MR analysis for trait pairs that were significant at an FDR of 5%. Individual scatter and funnel plots for each pair of traits are available in **Extended Data Fig. 10**. IVW: Inverse variance weighted analysis; WME: Weighted median estimator; WMBE: weighted mode based estimator; MR-PRESSO: Mendelian Randomization Pleiotropy RESidual Sum and Outlier. RBC: Red blood cell count

We next used two-sample Mendelian randomization (MR) to infer potentially causal relationships between these traits. Fixed-effects IVW analysis was used as the primary analysis ^37^, with weighted median estimator (WME) ^38^, weighted mode based estimator (WMBE) ^39^, MR Egger regression ^40^ and MR-PRESSO ^41^ outlier corrected estimates used as additional sensitivity analyses.

After correcting for multiple testing (FDR < 0.05), 8 exposure — COVID-19 trait-pairs showed suggestive evidence of a causal association (**Fig. 4; Supplementary Table 10a**). Five of these associations were robust to potential violations of the underlying assumptions of MR. Corroborating our genetic correlation results and evidence from traditional epidemiological studies, genetically predicted higher BMI (OR [95%CI] 1.4 [1.3, 1.6], *P* = 8.5 × 10^−11^) and smoking (OR [95%CI] = 1.9 [1.3, 2.8], *P* = 0.0012) were associated with increased risk of COVID-19 hospitalization, with BMI also being associated with increased risk of SARS-CoV-2 infection (OR [95%CI] = 1.1 [1.1, 1.2], *P* = 4.8 × 10^−7^). Genetically predicted increased height (OR [95%CI] = 1.1 [1, 1.1]), *P* = 8.9 × 10^−4^) was associated with an increased risk of reported infection, and genetically predicted higher red blood cell count (OR [95%CI] = 0.93 [0.89, 0.96], *P* = 5.7 × 10^−5^) was associated with a reduced risk of reported infection.

We noted that there was sample overlap between some datasets used to generate exposures used in the previous analysis, and the samples contributing to our meta-analysis of hospitalized COVID-19, as a result of inclusion of samples from the UK Biobank. We therefore conducted an additional sensitivity analysis, using new hospitalized COVID-19 summary statistics in which the UK Biobank study had been removed (**Supplementary Table 10b**). In this analysis, genetically predicted BMI, height, and red blood cell counts remained significantly associated with COVID-19 outcomes (unadjusted *P-value* < 0.05).

## DISCUSSION

The COVID-19 Host Genetics Initiative has brought together investigators from across the world to advance genetic discovery for SARS-CoV-2 infection and severe COVID-19 disease. We report 15 genome-wide significant loci associated with some aspect of SARS-CoV-2 infection or COVID-19. Many of these loci overlap with previously reported associations with lung-related phenotypes or autoimmune/inflammatory diseases, but some loci have no obvious candidate gene as yet.

Four out of the 13 genome-wide significant loci showed similar effects in the reported infection analysis (a proxy for disease susceptibility) and all-hospitalized COVID-19 (a proxy for disease severity). Of these, one locus was in close proximity, but yet independent, to the major genetic signal for COVID-19 severity at 3p21.31. Surprisingly, this locus was associated with COVID-19 susceptibility rather than severity. The locus, which comprises at least three independent signals all associated with susceptibility, overlaps SLC6A20, which encodes an amino acid transporter that interacts with ACE2. Nonetheless we caution that more data is needed to resolve the structure of the locus. In particular, the physical proximity and the lack of expected relationship between *P*-value and linkage disequilibrium structure raises the suspicion that untagged genetic variation might be underlying this cluster of loci. Our findings support the notion that some genetic variants, most notably at *ABO* and *PPP1R15A* loci, in addition to the aforementioned SLC6A20, might indeed impact susceptibility to infection rather than progression to a severe form of the disease once infected. Whilst our ability to draw definitive conclusions is impaired by incomplete capture of who has been infected with SARS-CoV-2, a recent study based on self-reported exposure to COVID-19 positive housemate and consequent development of the disease, support our findings ^42^.

Several of the loci reported here, as noted in previous publications ^15,17^, intersect with well-known genetic variants that have established genetic associations. Examples of these include variants at *DPP9* which show prior evidence of increasing risk for interstitial lung disease ^26^, and missense variants within *TYK2* that show a protective effect on several autoimmune-related diseases ^43–46^. Variants overlapping the well-known structural variants-rich 17q21.31 locus have been previously associated with pulmonary function. Together with the heritability enrichment observed in genes expressed in lung tissues, these results highlight the involvement of lung-related biological pathways in developing severe COVID-19. Several other loci show no prior documented genome-wide significant associations, even despite the high significance and attractive candidate genes for COVID-19 (e.g., *CXCR6, LZTFL1, IFNAR2* and *OAS1/2/3* loci). The previously reported associations for the strongest association for COVID-19 severity at 3p21.31 and monocytes count are likely to be due to proximity and not a true co-localization.

Increasing the global representation in genetic studies enhances the ability to detect novel associations. Two of the loci affecting disease severity were only discovered by including the four studies of individuals with East Asian ancestry. One of these loci, close to *FOXP4*, is common particularly in East Asian (32%) as well as Admixed American in the Americas (20%) and Middle Eastern samples (7%), but has a low frequency in most European populations (2-3%) in our data. Previous studies have reported association between this locus and lung cancer ^27,47^ and interstitial lung disease ^48^. Although we cannot be certain of the mechanism of action of this association *FOXP4* is an attractive biological target, as it is expressed in the proximal and distal airway epithelium ^49^, and has been shown to play a role in controlling epithelial cell fate during lung development ^50^. The COVID-19 Host Genetics Initiative continues to pursue expansion of the datasets included in the consortium’s analyses to populations from underrepresented populations in upcoming data releases.

A central challenge for the COVID-19 HGI was to define the phenotypes and analytic pipelines from the outset of the Initiative so that these would be applicable to cohorts with extremely heterogeneous designs, sample ascertainment and control populations. Large-scale biobanks with existing genotype resources and connections to medical systems, newly enrolled hospital-based studies (particularly well-powered to study the extremes of severity by through the recruitment of individuals from intensive care units), and direct-to-consumer genetics studies with customer surveys each contributed different aspects to understanding the genetic basis of susceptibility and severity traits. Indeed, working together through aligning phenotype definitions and sharing results accelerated progress and has enhanced the robustness of the reported findings.

Nevertheless, the differences in study sample size, ascertainment and phenotyping of COVID-19 cases are unavoidable and care should be taken when interpreting the results from a meta-analysis. First, studies enriched with severe cases or studies with antibody-tested controls may disproportionately contribute to genetic discovery despite potentially smaller sample sizes. Second, differences in genomic profiling technology, imputation, and sample size across the constituent studies can have dramatic impacts on replication and downstream analyses (particularly fine-mapping where differential missing patterns in the reported results can muddy the signal). Third, the use of population controls with no complete information about SARS-CoV-2 exposure might result in cases of misclassification or reflect ascertainment biases in testing and reporting rather than true susceptibility to infection. Genotyping large numbers of control samples who have been exposed to the virus but remained asymptomatic or experienced only mild symptoms is challenging. Therefore many studies prefer to use pre-existing datasets of genetically ancestry-matched samples as their controls, protecting against population stratification, but potentially introducing some of these biases. Our analysis comparing the discovery meta-analysis effects to one where controls were phenotypically refined, indicated that, for genome-wide significant variants, such bias was limited. Finally, sociodemographic factors may influence an individual’s susceptibility to SARS-CoV-2 infection and COVID-19 severity. In particular, lower socioeconomic level is associated with a higher risk of infection and hospitalization ^51^. This can result in collider bias which distorts the relationship between genetic variation and the phenotypes being examined ^52^. Additionally, other factors such as time of infection, which affects mortality and critical care admissions ^53^), or differences in vaccination schemes may change the sociodemographic characteristics of COVID-19 positive participants.

Drawing a comprehensive and reproducible map of the host genetics factors associated with COVID-19 severity and SARS-CoV-2 requires a sustained international effort to include diverse ancestries and study designs. The number of COVID-19 study participants and studies contributing data to this study illustrate the benefits of worldwide international collaboration, open governance and planning, and sharing of technological and analytical resources. To expedite downstream scientific research and therapeutic discovery, the COVID-19 Host Genetic Initiative regularly publishes meta-analysis results from periodic data freezes on the website www.covid19hg.org as new data are included in the study. We also provide an interactive explorer where researchers can browse the results and the genomic loci in more detail. Future work will be required to better understand the biological and clinical value of these findings. Continued efforts to collect more samples and detailed phenotypic data should be endorsed globally; allowing for more thorough investigation of variable, heritable symptoms ^54,55^, particularly in the light of newly emerging strains of SARS-CoV-2 virus, which may provoke different host responses leading to disease.

## METHODS

### Contributing studies

In total 16 studies contributed data to analysis of critical illness due to COVID-19, 29 studies contributed data to hospitalized COVID-19 analysis, and 44 studies contributed to the analysis of all COVID-19 cases. The effective sample sizes for each ancestry group shown in Figure 1 were calculated for display using the formula: ((4 × N_cases × N_controls) / (N_cases + N_controls)). Details of contributing research groups are described in **Supplementary Table 1**. All subjects were recruited following protocols approved by local Institutional Review Boards (IRBs). All protocols followed local ethics recommendations and informed consent was obtained when required.

### Phenotype Definitions

COVID-19 disease status (critical illness, hospitalization status) was assessed following the Diagnosis and Treatment Protocol for Novel Coronavirus Pneumonia ^56^. The critically ill COVID-19 group included patients who were hospitalized due to symptoms associated with laboratory-confirmed SARS-CoV-2 infection and who required respiratory support or whose cause of death was associated with COVID-19. The hospitalized COVID-19 group included patients who were hospitalized due to symptoms associated with laboratory-confirmed SARS-CoV-2 infection.

The reported infection cases group included individuals with laboratory-confirmed SARS-CoV-2 infection or electronic health record, ICD coding or clinically confirmed COVID-19, or self-reported COVID-19 (e.g. by questionnaire), with or without symptoms of any severity. Genetic ancestry-matched controls for the three case definitions were sourced from population-based cohorts, including individuals whose exposure status to SARS-CoV-2 was either unknown or infection-negative for questionnaire/electronic health record based cohorts. Additional information regarding individual studies contributing to the consortium are described in **Supplementary Table 1**.

### GWAS and meta-analysis

Each contributing study genotyped the samples and performed quality controls, data imputation and analysis independently, but following consortium recommendations (information available at www.covid19hg.org). We recommended to run GWAS analysis using Scalable and Accurate Implementation of GEneralized mixed model (SAIGE) ^57^ on chromosomes 1-22 and X. The recommended analysis tool was SAIGE, but studies also used other software such as PLINK ^58^. The suggested covariates were age, age2, sex, age*sex, and 20 first principal components. Any other study-specific covariates to account for known technical artefacts could be added. SAIGE automatically accounts for sample relatedness and case-control imbalances. Individual study quality control and analysis approaches are reported in **Supplementary Table 1**.

Study-specific summary statistics were then processed for meta-analysis. Potential false positives, inflation, and deflation were examined for each submitted GWAS. Standard error values as a function of effective sample size was used to find studies which deviated from the expected trend (**Extended Data Figure 11**). Summary statistics passing this manual quality control were included in the meta-analysis. Variants with allele frequency of >0.1% and imputation INFO>0.6 were carried forward from each study. Variants and alleles were lifted over to genome build GRCh38, if needed, and harmonized to gnomAD 3.0 genomes ^59^ by finding matching variants by strand flipping or switching ordering of alleles. If multiple matching variants, the best match was chosen by minimum absolute allele frequency fold change. Meta-analysis was performed using the inverse-variance weighted method on variants that were present in at least ⅔ of studies contributing to the phenotype analysis. The method summarizes effect sizes across the multiple studies by computing the mean of the effect sizes weighted by the inverse variance in each individual study.

We report 15 variants that pass genome-wide significance threshold after adjusting the threshold for multiple traits tested (*P* < 5 × 10^−8^ / 3) and three additional which pass genome-wide significance before threshold adjusting (*P* < 5 × 10^−8^). Two loci reached genome-wide significance but were excluded from **Table 1** significant results due to heterogeneity and missingness between studies at chr6:31057940-31380334 and chr7:54671568-54759789; however these regions are not excluded from the corresponding summary statistics data release. For each of the total 15 independent lead variants reported in **Table 1**, we tested whether there was heterogeneity between the effect sizes associated with hospitalized COVID-19 (progression to severe disease) and reported SARS-CoV-2 infection. We used Cochran’s Q measure ^60,61^, which is calculated for each variant as the weighted sum of squared differences between the two analysis effects sizes and their meta-analysis effect, the weights being the inverse variance of the effect size. Q is distributed as a chi-square statistic with k (number of studies) minus 1 degrees of freedom. A significant *P*-value <0.003 (0.05/15 for multiple tests) indicates that the effect sizes for a particular variant are significantly different in the two analyses (**Supplementary Table 2**). For the 9 loci, where the lead variant effect size was significantly higher for hospitalized COVID-19, we carried out the same test again but comparing effect sizes from hospitalized COVID-19 with critically ill COVID-19 (**Supplementary Table 3**). Further, we carried out the same test comparing meta-analyzed hospitalized COVID-19 (population as controls) and hospitalized COVID-19 (SARS-CoV-2 positive but non-hospitalized as controls) (**Supplementary Table 3)**. For these pairs of phenotype comparisons, we generated new meta-analysis summary statistics to use; including only those studies that could contribute data to both phenotypes that were under comparison.

### PC projection

To project every GWAS participant into the same PC space, we used pre-computed PC loadings and reference allele frequencies. For reference, we used unrelated samples from the 1000 Genomes Project and the Human Genome Diversity Project (HGDP) and computed PC loadings and allele frequencies for the 117,221 SNPs that are i) available in every cohort, ii) MAF > 0.1% in the reference, and iii) LD pruned (r2 < 0.8; 500kb window). We then asked each cohort to project their samples using our automated script provided here (https://github.com/covid19-hg/pca_projection). It internally uses PLINK2 ^62^ --score function with variance-standardize option and reference allele frequencies (--read-freq); so that each cohort-specific genotype/dosage matrix is mean-centered and variance-standardized with regards to reference allele frequencies, not cohort-specific allele frequencies. We further normalized the projected PC scores by dividing by a square root of the number of variants used for projection to account for a subtle difference due to missing variants.

### Gene prioritization

To prioritize candidate causal genes, we employed various gene prioritization approaches using both locus-based and similarity-based methods. Because we only referred *in-silico* gene prioritization results without characterizing actual functional activity *in-vitro/vivo*, we aimed to provide a conservative list of any potential causal genes in a locus using the following criteria:

1. Closest gene: a gene that is closest to a lead variant by distance to the gene body
2. Genes in LD region: genes that overlap with a genomic range containing any variants in LD (*r*^*2*^ > 0.6) with a lead variant. For LD computation, we retrieved LD matrices provided by the gnomAD v2.1.1 ^59^ for each population analyzed in this study (except for Admixed American, Middle Eastern, and South Asian that are not available). We then constructed a weighted-average LD matrix by per-population sample sizes in each meta-analysis, which we used as a LD reference.
3. Genes with coding variants: genes with at least one loss of function or missense variant (annotated by VEP ^63^ v95 with GENCODE v29) that is in LD with a lead variant (*r*^2^ > 0.6).
4. eGenes: genes with at least one fine-mapped *cis*-eQTL variant (PIP > 0.1) that is in LD with a lead variant (*r*^2^ > 0.6) (**Supplementary Table 4**). We retrieved fine-mapped variants from the GTEx v8^24^ (https://www.finucanelab.org/) and eQTL catalogue^64^. In addition, we looked up significant associations in the Lung eQTL Consortium ^25^ (*n* = 1,103) to further support findings in lung with a larger sample size (**Supplementary Table 11**). We note that, unlike the GTEx or eQTL catalogue, we only looked at associations and didn’t finemap in the Lung eQTL Consortium data.
5. V2G: a gene with the highest overall Variant-to-Gene (V2G) score based on the Open Targets Genetics (OTG) ^21^. For each variant, the overall V2G score aggregates differentially weighted evidence of variant-gene association from several data sources, including molecular cis-QTL data (e.g., cis-pQTLs from ^65^., cis-eQTLs from GTEx v7 etc.), interaction-based datasets (e.g., Promoter Capture Hi-C), genomic distance, and variant effect predictions (VEP) from Ensembl. A detailed description of the evidence sources and weights used is provided in the OTG documentation (https://genetics-docs.opentargets.org/our-approach/data-pipeline) ^21,66^.

### Phenome-wide association study

To investigate the evidence of shared effects of 15 index variants for COVID-19 and previously reported phenotypes, we performed a phenome-wide association study. We considered phenotypes in (Open Target)

OTG obtained from the GWAS catalog (this included studies with and without full summary statistics, n = 300 and 14,013, respectively) ^67^, and from UK Biobank. Summary statistics for UK Biobank traits were extracted from SAIGE ^57^ for binary outcomes (*n* = 1,283 traits), and Neale v2 (*n* = 2,139 traits) for both binary and quantitative traits (http://www.nealelab.is/uk-biobank/)and FinnGen Freeze 4 cohort (https://www.finngen.fi/en/access_results). To remove plausible spurious associations, we retrieved phenotypes for GWAS lead variants that were in LD (*r*^*2*^>0.8) with COVID-19 index variants.

### Heritability

LD score regression v 1.0.1 ^34^ was used to estimate SNP heritability of the phenotypes from the meta-analysis summary statistic files. As this method depends on matching the linkage disequilibrium (LD) structure of the analysis sample to a reference panel, the European-only summary statistics were used. Sample sizes were *n* = 5,101 critically ill COVID-19 cases and *n* = 1,383,241 controls, *n* = 9,986 hospitalized COVID-19 cases and *n* = 1,877,672 controls, and *n* = 38,984 cases and *n* = 1,644,784 controls for all cases analysis, all including the 23andMe cohort. Pre-calculated LD scores from the 1000 Genomes European reference population were obtained online (https://data.broadinstitute.org/alkesgroup/LDSCORE/). Analyses were conducted using the standard program settings for variant filtering (removal of non-HapMap3 SNPs, the HLA region on chromosome 6, non-autosomal, chi-square > 30, MAF < 1%, or allele mismatch with reference). We additionally report SNP heritability estimates for the all-ancestries meta-analyses, calculated using European panel LD scores, in **Supplementary Table 6**.

### Partitioned heritability

We used partitioned LD score regression ^68^ to partition COVID-19 SNP heritability in cell types in our European-only summary statistics. We ran the analysis using the baseline model LD scores calculated for European populations and regression weights that are available online. We used the COVID-19 European only summary statistics for the analysis.

### Genome-wide association summary statistics

We obtained genome-wide association summary statistics for 43 complex disease, neuropsychiatric, behavioural, or biomarker phenotypes (**Supplementary Table 8**). These phenotypes were selected based on their putative relevance to COVID-19 susceptibility, severity, or mortality, with 19 selected based on the Centers for Disease Control list of underlying medical conditions associated with COVID-19 severity ^69^ or traits reported to be associated with increased risk of COVID-19 mortality by OpenSafely ^70^. Summary statistics generated from GWAS using individuals of European ancestry were preferentially selected if available. These summary statistics were used in subsequent genetic correlation and Mendelian randomization analyses.

### Genetic Correlation

LD score regression ^68^ was also used to estimate genetic correlations between our COVID-19 meta-analysis phenotypes reported using European-only samples, and between these and the curated set of 38 summary statistics. Genetic correlations were estimated using the same LD score regression settings as for heritability calculations. Differences between the observed genetic correlations of SARS-CoV-2 infection and COVID-19 severity were compared using a z score method ^71^.

### Mendelian Randomization

Two-sample Mendelian randomization was employed to evaluate the causal association of the 38 traits on COVID-19 hospitalization, on COVID-19 severity and SARS-CoV-2 reported infection using European-only samples. Independent genome-wide significant SNPs robustly associated with the exposures of interest (*P* < 5 × 10^−8^) were selected as genetic instruments by performing LD clumping using PLINK ^58^. We used a strict *r*^*2*^ threshold of 0.001, a 10MB clumping window, and the European reference panel from the 1000 Genomes project ^72^ to discard SNPs in linkage disequilibrium with another variant with smaller p-value association. For genetic variants that were not present in the hospitalized COVID analysis, PLINK was used to identify proxy variants that were in LD (*r*^*2*^ > 0.8). Next, the exposure and outcome datasets were harmonized using the R-package TwoSampleMR ^73^. Namely, we ensured that the effect of a variant on the exposure and outcome corresponded to the same allele, we inferred positive strand alleles and dropped palindromes with ambiguous allele frequencies, as well as incompatible alleles. **Supplementary Table 8** includes the harmonized datasets used in the analyses.

Mendelian Randomization Pleiotropy residual sum and outlier (MR-PRESSO) Global test ^41^ was used to investigate overall horizontal pleiotropy. In short, the standard IVW meta-analytic framework was employed to calculate the average causal effect by excluding each genetic variant used to instrument the analysis. A global statistic was calculated by summing the observed residual sum of squares, i.e., the difference between the effect predicted by the IVW slope excluding the SNP, and the observed SNP-effect on the outcome. Overall horizontally pleiotropy was subsequently probed by comparing the observed residual sum of squares, with the residual sum of squares expected under the null hypothesis of no pleiotropy. The MR-PRESSO Global test was shown to perform well when the outcome and exposure GWASs are not disjoint (although the power to detect horizontal pleiotropy is slightly reduced by complete sample overlap). We also used the MR-Egger regression intercept ^40^ to evaluate potential bias due to directional pleiotropic effects. This additional check was employed in MR analyses with an 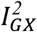 index surpassing the recommended threshold (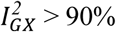; ^74^). Contingent on the MR-PRESSO Global test results we probed the causal effect of each exposure on COVID-19 hospitalization by using a fixed effect inverse-weighted (IVW) meta-analysis as the primary analysis, or, if pleiotropy was present, the MR-PRESSO outlier corrected test. The IVW approach estimates the causal effect by aggregating the single-SNP causal effects (obtained using the ratio of coefficients method, i.e., the ratio of the effect of the SNP on the outcome on the effect of the SNP on the exposure) in a fixed effects meta-analysis. The SNPs were assigned weights based on their inverse variance. The IVW method confers the greatest statistical power for estimating causal associations ^75^, but assumes that all variants are valid instruments and can produce biased estimates if the average pleiotropic effect differs from zero. Alternatively, when horizontal pleiotropy was present, we used MR-PRESSO Outlier corrected method to correct the IVW test by removing outlier SNPs. We conducted further sensitivity analyses using alternative MR methods that provide consistent estimates of the causal effect even when some instrumental variables are invalid, at the cost of reduced statistical power including: 1) Weighted Median Estimator (WME); 2) Weighted Mode Based Estimator (WMBE); 3) MR-Egger regression. Robust causal estimates were defined as those that were significant at an FDR of 5% and either 1) showed no evidence of heterogeneity (MR-PRESSO Global test *P* > 0.05) or horizontal pleiotropy (Egger Intercept *P* > 0.05), or 2) in the presence of heterogeneity or horizontal pleiotropy, either the WME, WMBE, MR-Egger or MR-PRESSO corrected estimates were significant (*P* < 0.05). All statistical analyses were conducted using R version 4.0.3. MR analysis was performed using the “TwoSampleMR” version 0.5.5 package ^73^.

### Website and data distribution

In anticipation of the need to coordinate many international partners around a single meta-analysis effort, we created the COVID-19 HGI website (https://covid19hg.org). We were able to centralize information, recruit partner studies, rapidly distribute summary statistics, and present preliminary interpretations of the results to the public. Open meetings are held on a monthly basis to discuss future plans and new results; video recordings and supporting documents are shared (https://covid19hg.org/meeting-archive). This centralized resource provides a conceptual and technological framework for organizing global academic and industry groups around a shared goal. The website source code and additional technical details are available at https://github.com/covid19-hg/covid19hg.

To recruit new international partner studies, we developed a workflow whereby new studies are registered and verified by a curation team (https://covid19hg.org/register). Users can explore the registered studies using a customized interface to find and contact studies with similar goals or approaches (https://covid19hg.org/partners). This helps to promote organic assembly around focused projects that are adjacent to the centralized effort (https://covid19hg.org/projects). Visitors can query study information, including study design and research questions. Registered studies are visualized on a world map and are searchable by institutional affiliation, city, and country.

To encourage data sharing and other forms of participation, we created a rolling acknowledgements page (https://covid19hg.org/acknowledgements) and directions on how to contribute data to the central meta-analysis effort (https://covid19hg.org/data-sharing). Upon the completion of each data freeze, we post summary statistics, plots, and sample size breakdowns for each phenotype and contributing cohort (https://covid19hg.org/results). The results can be explored using an interactive web browser (https://app.covid19hg.org). Several computational research groups carry out follow-up analyses, which are made available for download (https://covid19hg.org/in-silico). To enhance scientific communication to the public, preliminary results are described in blog posts by the scientific communications team and shared on Twitter. The first post was translated to 30 languages with the help of 85 volunteering translators. We compile publications and preprints submitted by participating groups and summarize genome-wide significant findings from these publications (https://covid19hg.org/publications).

## Data Availability

Summary statistics generated by COVID-19 HGI are available at https://www.covid19hg.org/results/r5/ and will be made available on GWAS Catalog. The analyses described here utilize the freeze 5 data. COVID-19 HGI continues to regularly release new data freezes. Summary statistics for non-European ancestry samples are not currently available due to the small individual sample sizes of these groups. Summary statistics liftover and meta-analysis and PCA pipeline code is available at https://github.com/covid19-hg/ and code for Mendelian randomization and genetic correlation pipeline at https://github.com/marcoralab/MRcovid.

https://github.com/covid19-hg/

https://github.com/marcoralab/MRcovid

## Data availability

Summary statistics generated by COVID-19 HGI are available at https://www.covid19hg.org/results/r5/ and will be made available on GWAS Catalog. The analyses described here utilize the freeze 5 data. COVID-19 HGI continues to regularly release new data freezes. Summary statistics for non-European ancestry samples are not currently available due to the small individual sample sizes of these groups.

## Code availability

Summary statistics liftover and meta-analysis and PCA pipeline code is available at https://github.com/covid19-hg/ and code for Mendelian randomization and genetic correlation pipeline at https://github.com/marcoralab/MRcovid

