## Extended data and supplement for "Mapping the human genetic architecture of COVID-19 by worldwide meta-analysis": Extended_Data_Figure_2.pdf

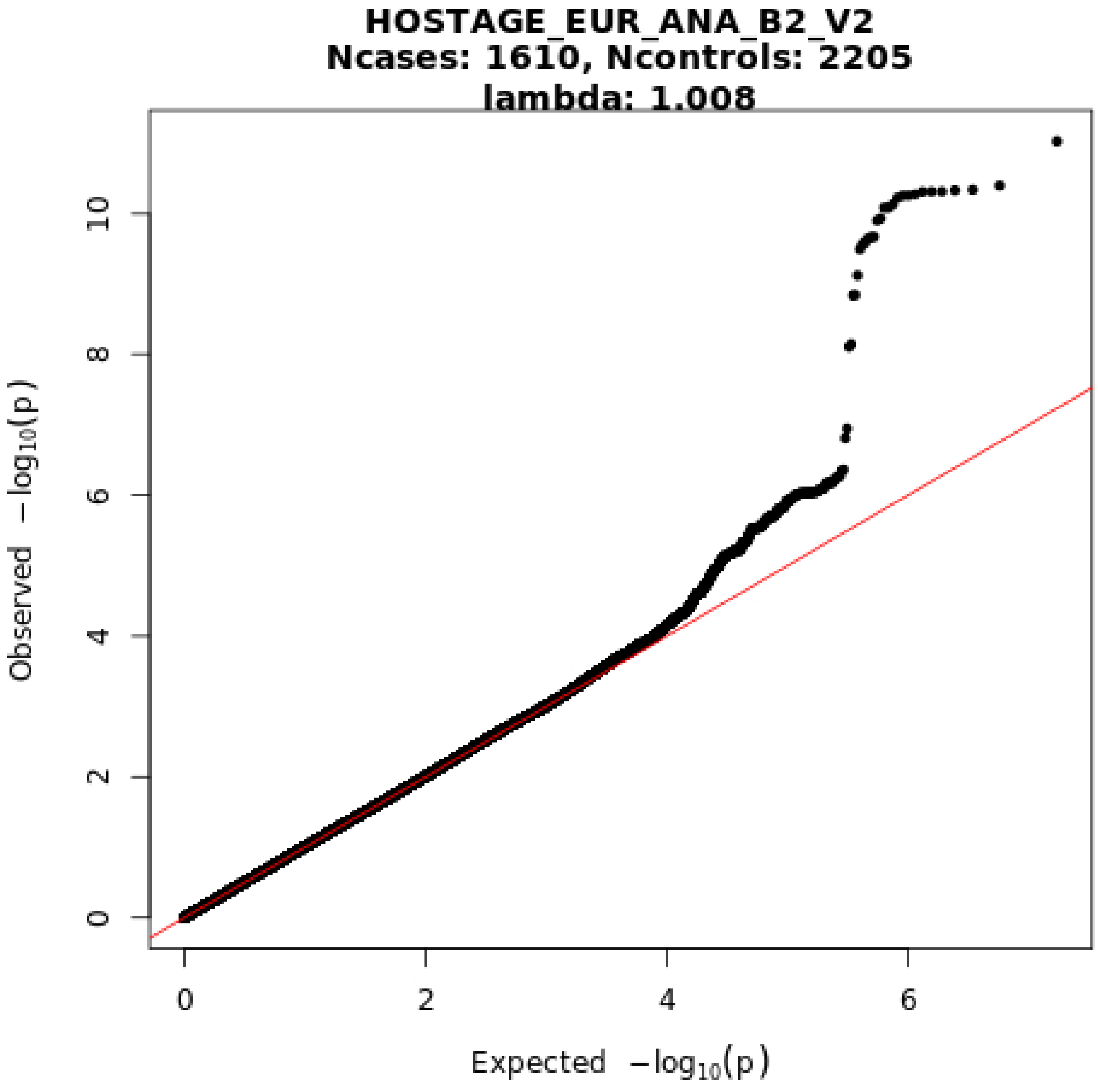
HOSTAGE_EUR_ANA_B2_V2
Ncases: 1610, Ncontrols: 2205
lambda: 1.008
Observed $-\log_{10}(p)$
Expected $-\log_{10}(p)$
0
2
4
6
8
10
0
2
4
6

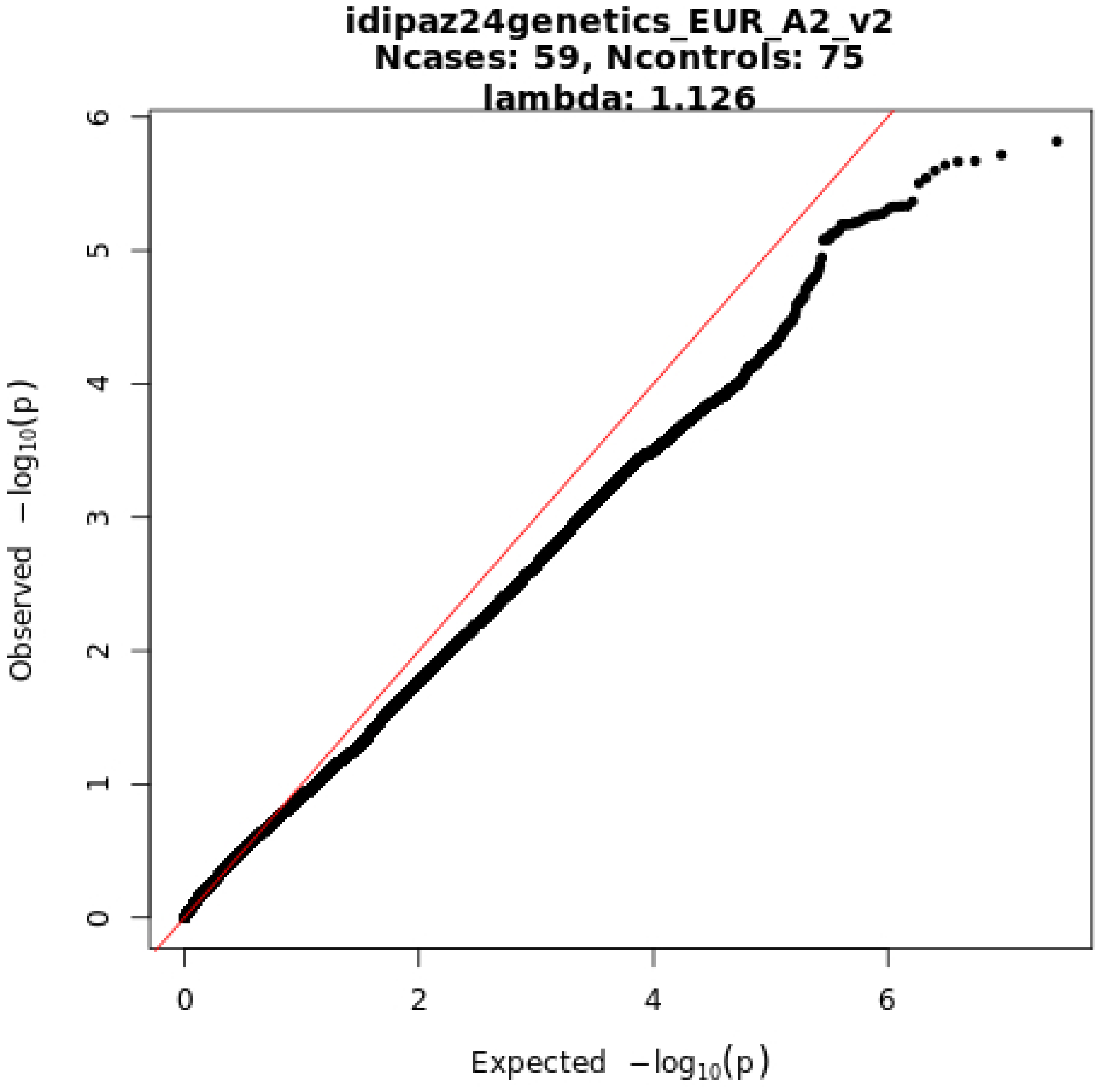
idipaz24genetics_EUR_A2_v2
Ncases: 59, Ncontrols: 75
lambda: 1.126
Observed −log10(p)
Expected −log10(p)
0
1
2
3
4
5
6
0
2
4
6

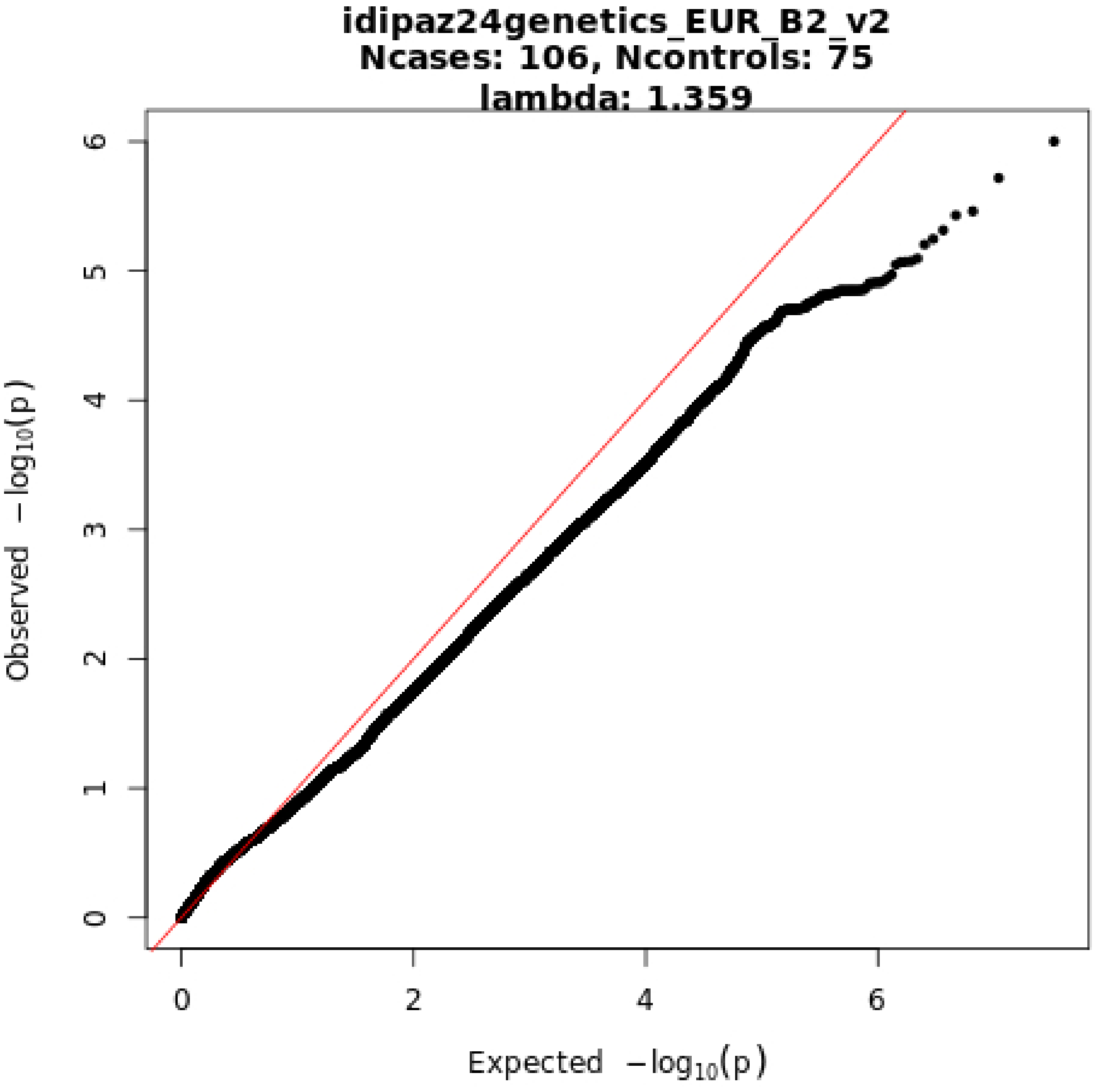

idipaz24genetics_EUR_B2_v2
Ncases: 106, Ncontrols: 75
lambda: 1.359
Observed −log10(p)
Expected −log10(p)
0
1
2
3
4
5
6
0
2
4
6

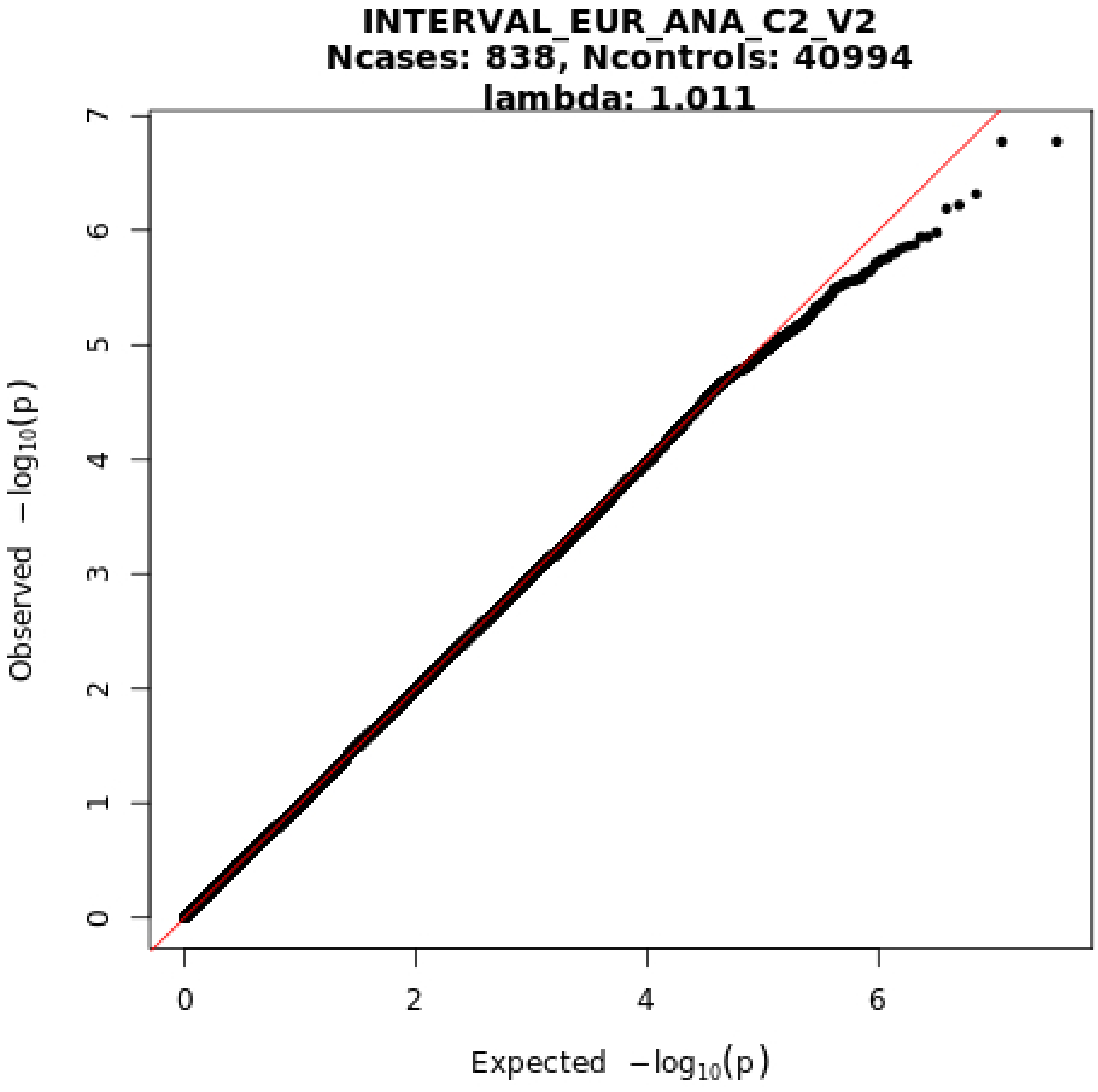
INTERVAL_EUR_ANA_C2_V2
Ncases: 838, Ncontrols: 40994
lambda: 1.011
Observed −log10(p)
Expected −log10(p)
0
1
2
3
4
5
6
7
0
2
4
6

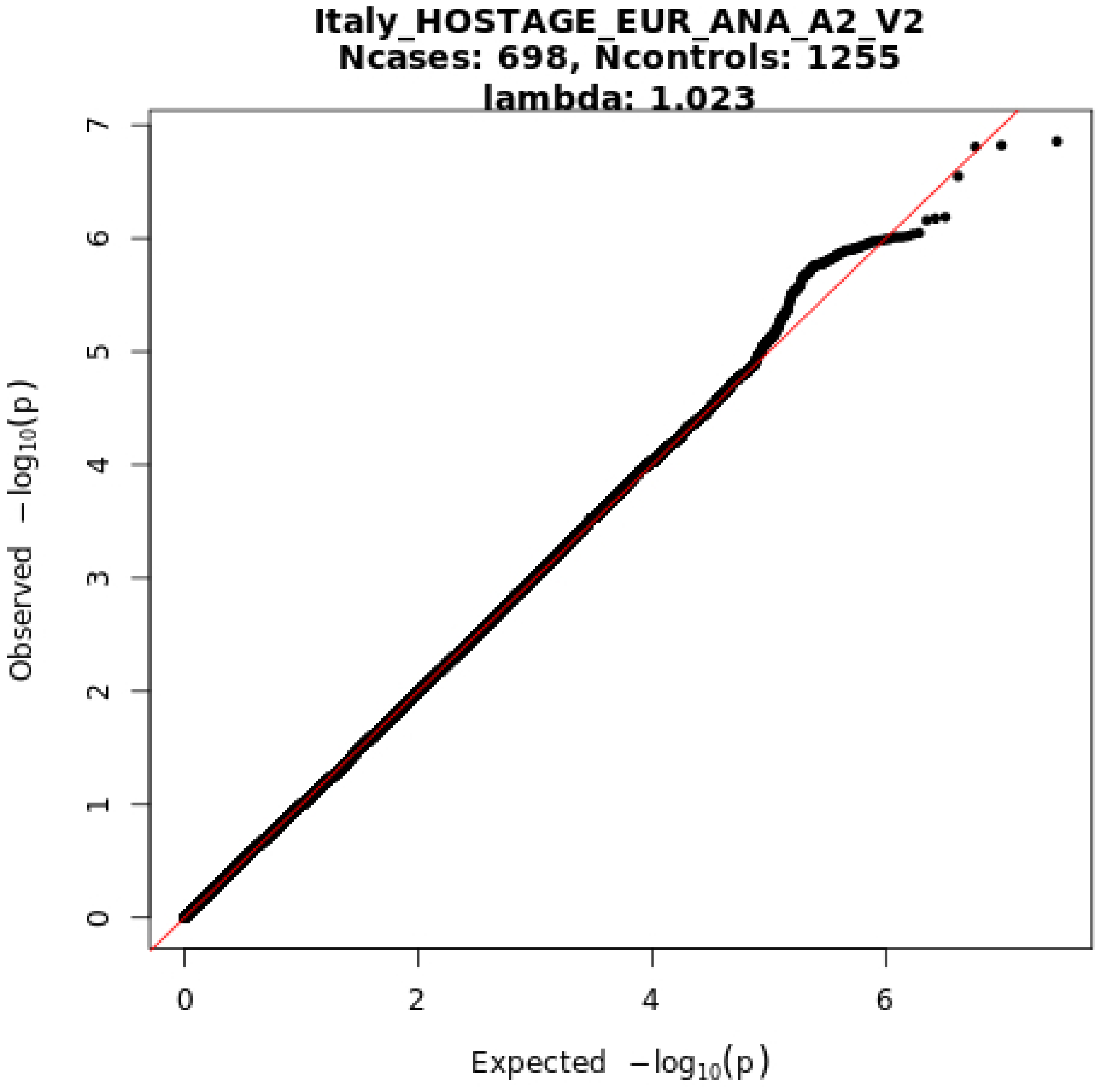

Italy_HOSTAGE_EUR_ANA_A2_V2
Ncases: 698, Ncontrols: 1255
lambda: 1.023
Observed $-\log_{10}(p)$
Expected $-\log_{10}(p)$
0
1
2
3
4
5
6
7
0
2
4
6

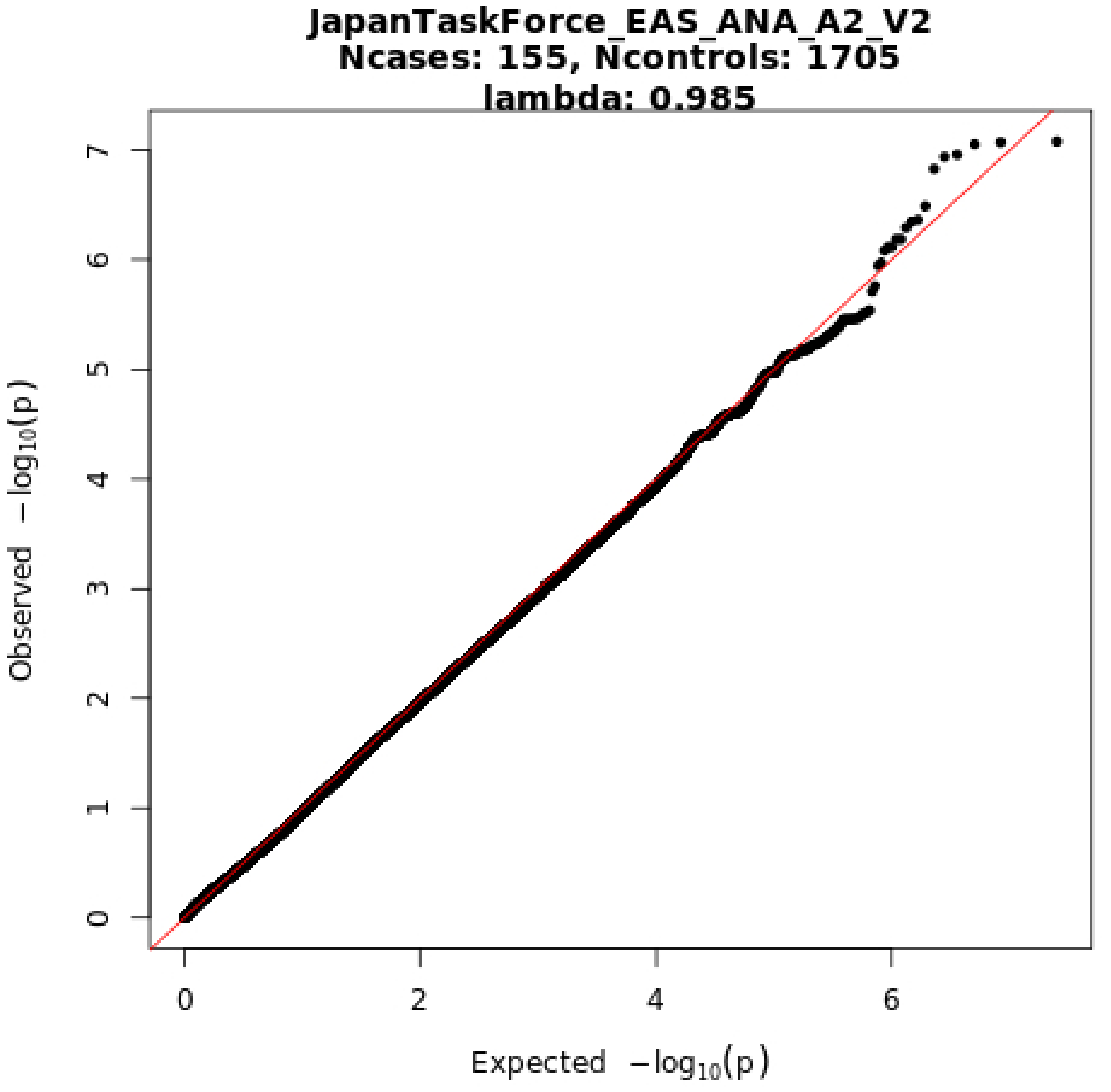
JapanTaskForce_EAS_ANA_A2_V2
Ncases: 155, Ncontrols: 1705
lambda: 0.985
Observed −log10(p)
Expected −log10(p)
0
1
2
3
4
5
6
7
0
2
4
6

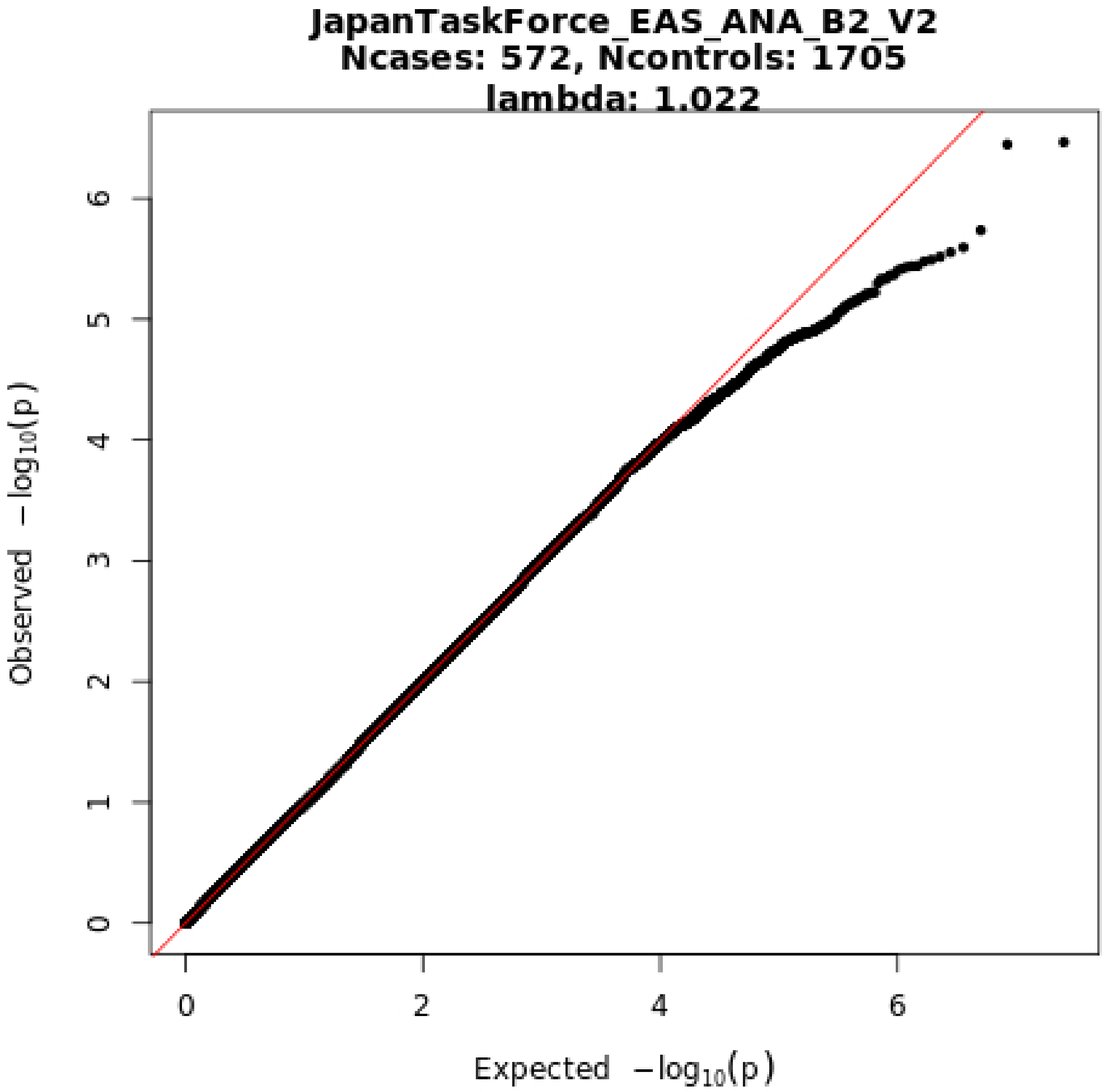
JapanTaskForce_EAS_ANA_B2_V2
Ncases: 572, Ncontrols: 1705
lambda: 1.022
Observed −log10(p)
Expected −log10(p)
0
1
2
3
4
5
6
0
2
4
6

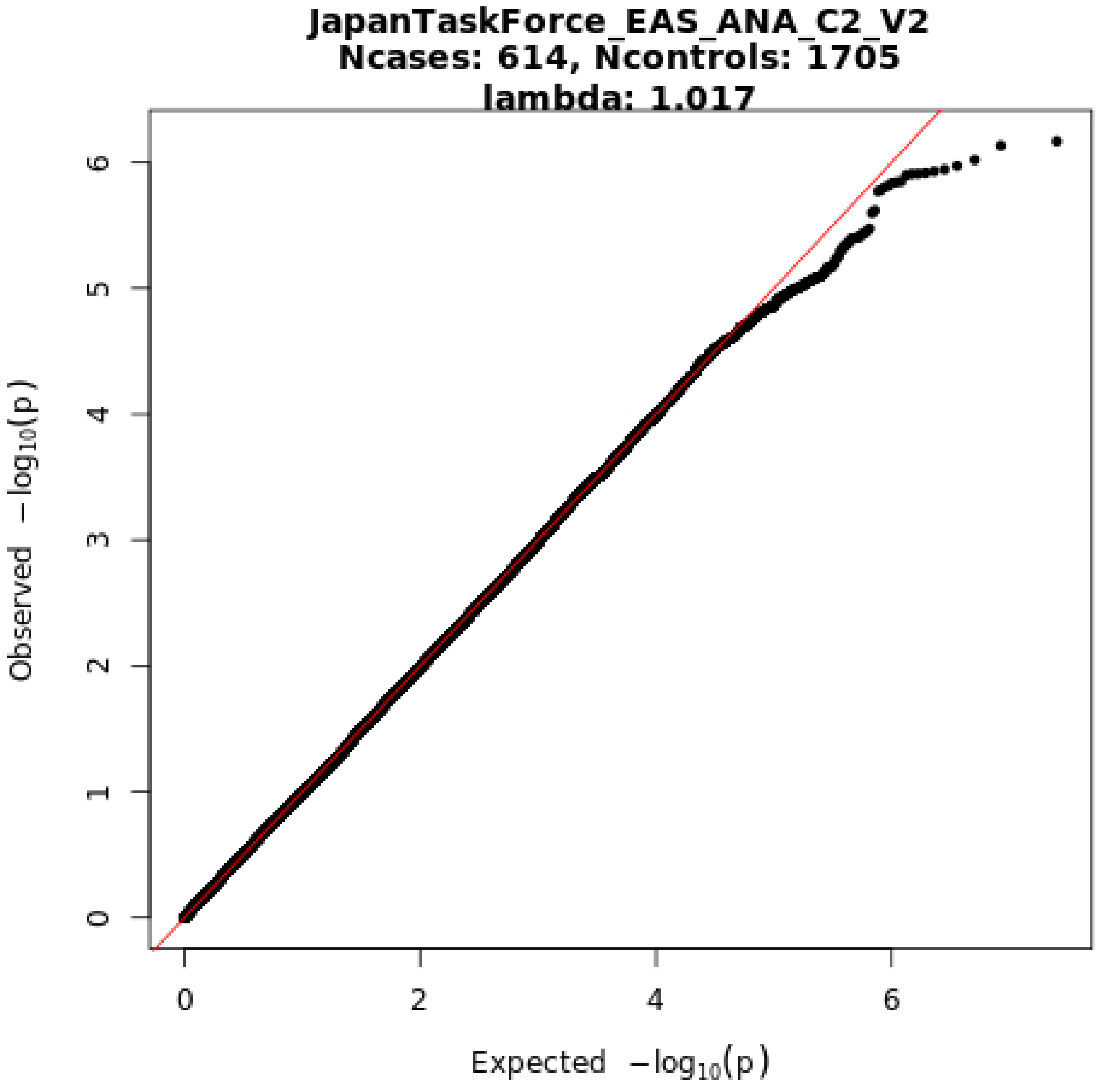
JapanTaskForce_EAS_ANA_C2_V2
Ncases: 614, Ncontrols: 1705
lambda: 1.017
Observed −log10(p)
Expected −log10(p)
0
1
2
3
4
5
6
0
2
4
6

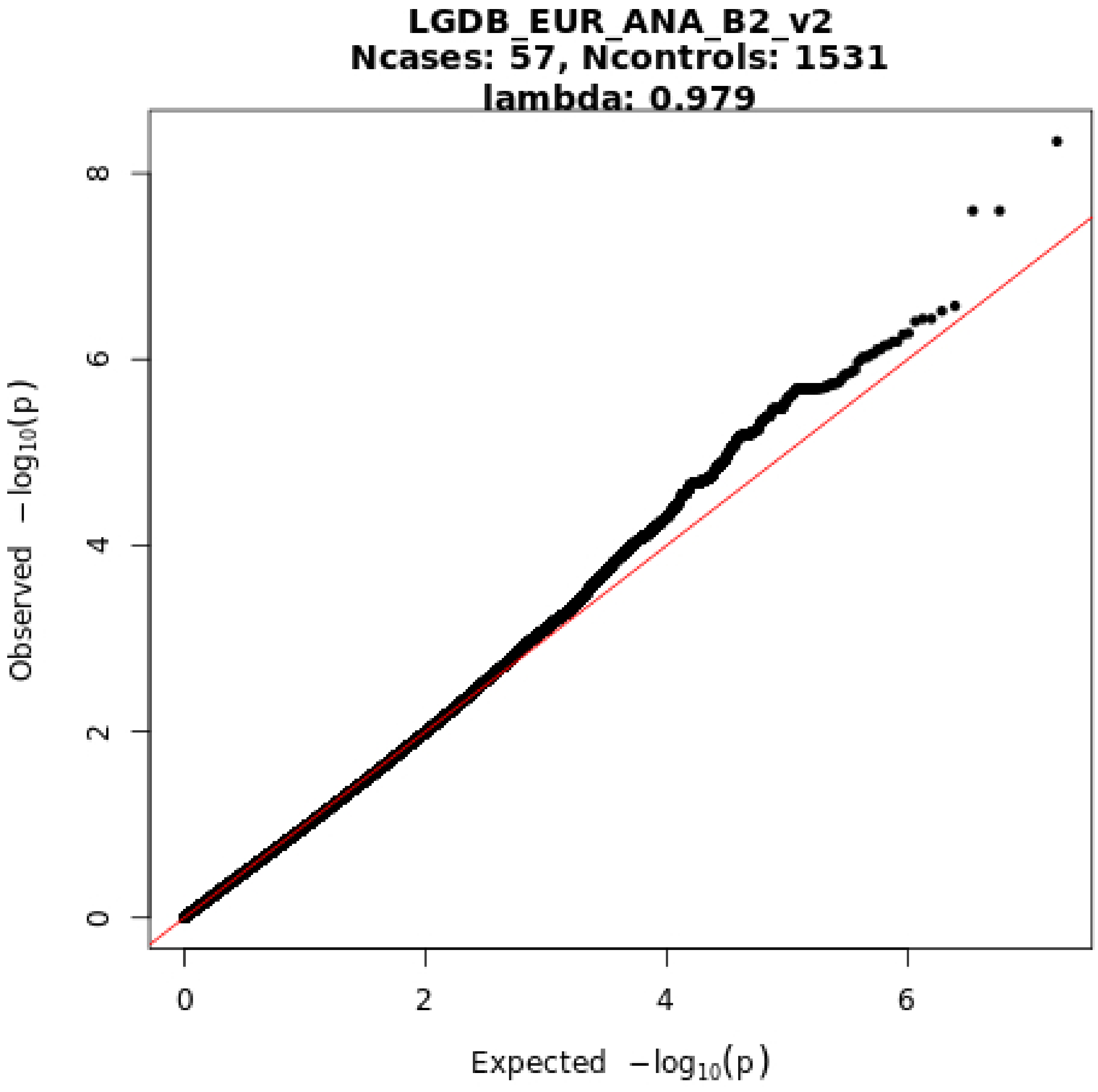

LGDB_EUR_ANA_B2_v2
Ncases: 57, Ncontrols: 1531
lambda: 0.979
0
2
4
6
8
Observed $-\log_{10}(p)$
Expected $-\log_{10}(p)$

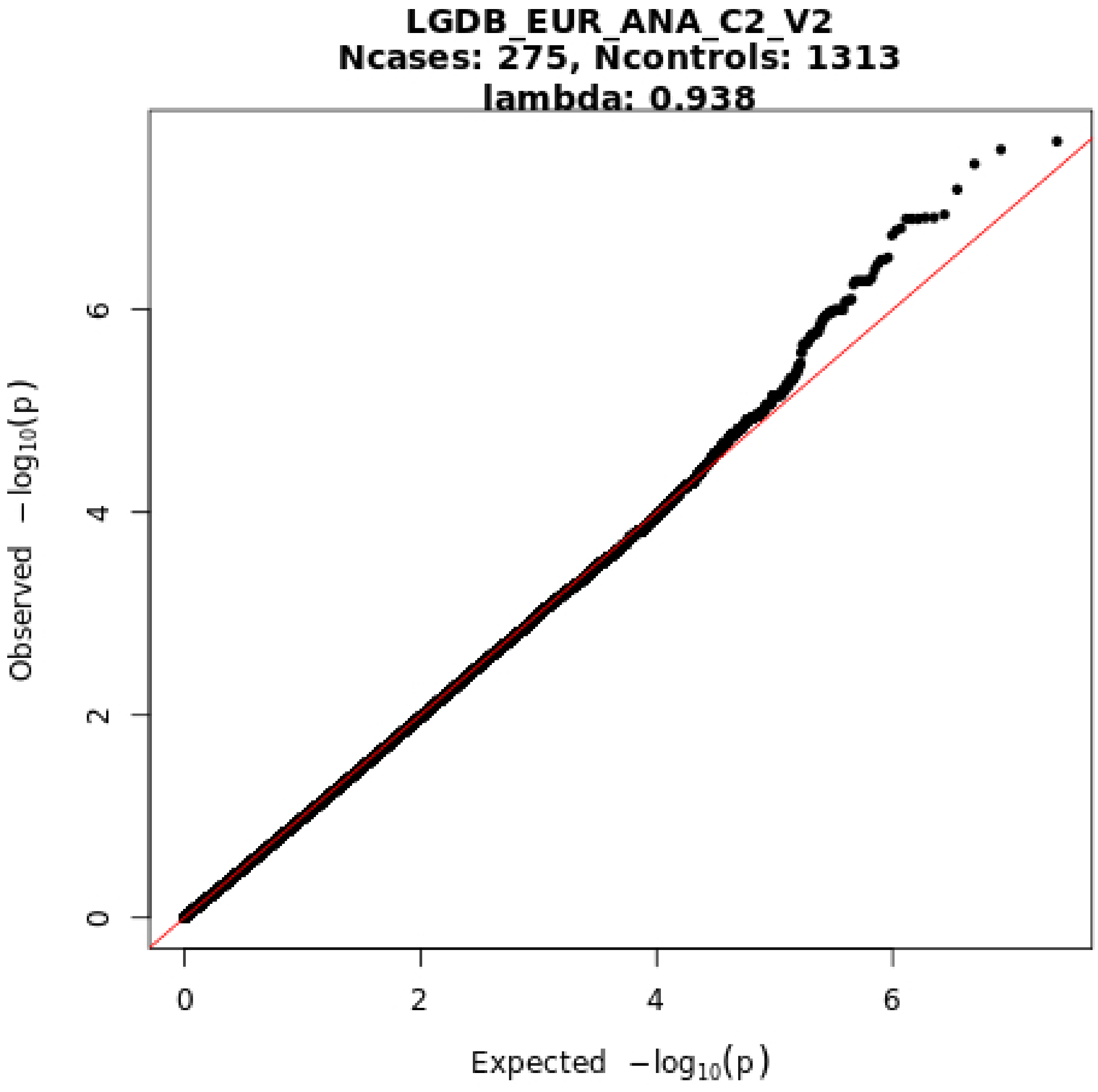
LGDB_EUR_ANA_C2_V2
Ncases: 275, Ncontrols: 1313
lambda: 0.938
Observed −log10(p)
Expected −log10(p)
0
2
4
6
0
2
4
6

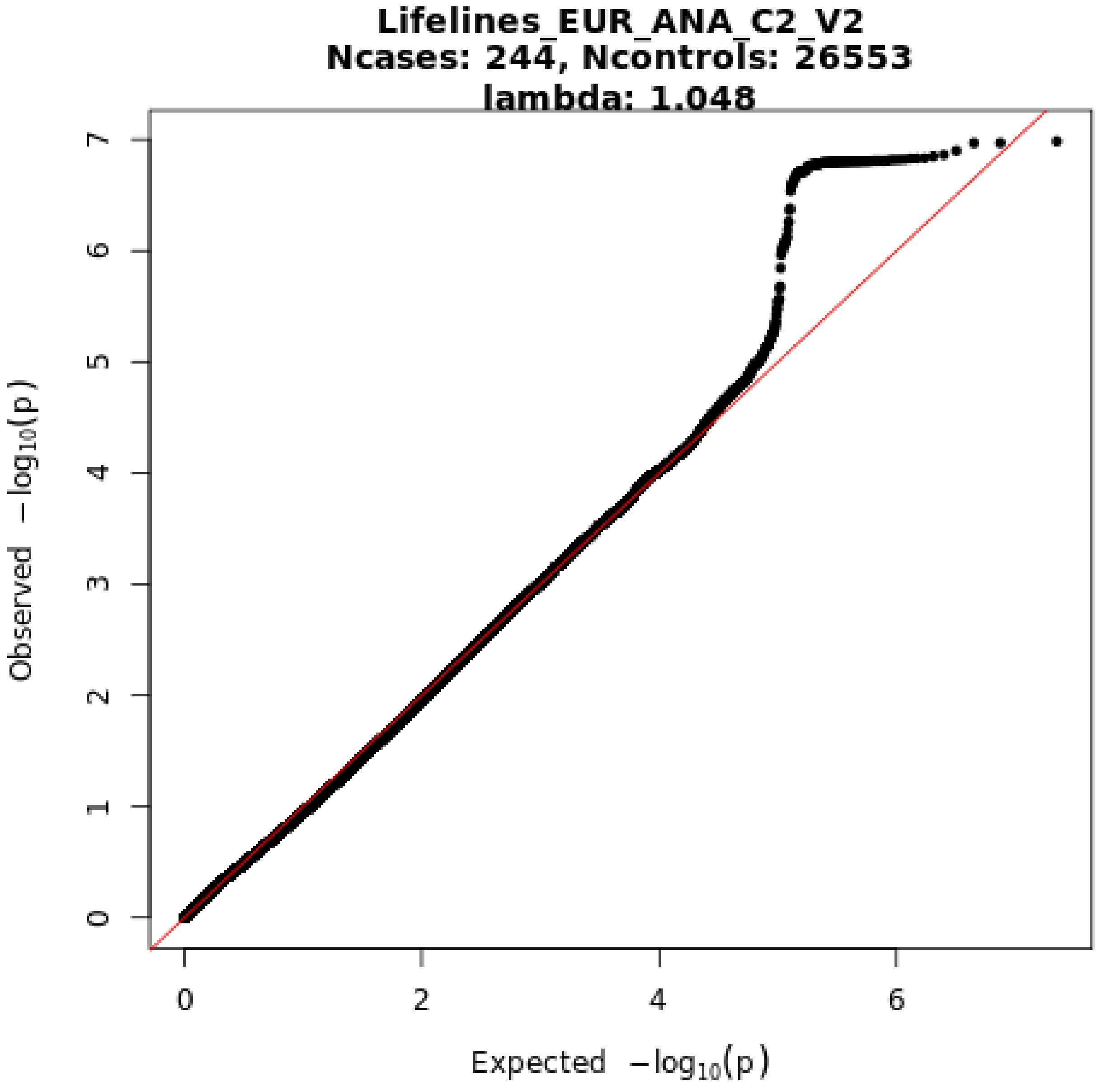

Lifelines_EUR_ANA_C2_V2
Ncases: 244, Ncontrols: 26553
lambda: 1.048
Observed −log10(p)
Expected −log10(p)
0
1
2
3
4
5
6
7
0
2
4
6

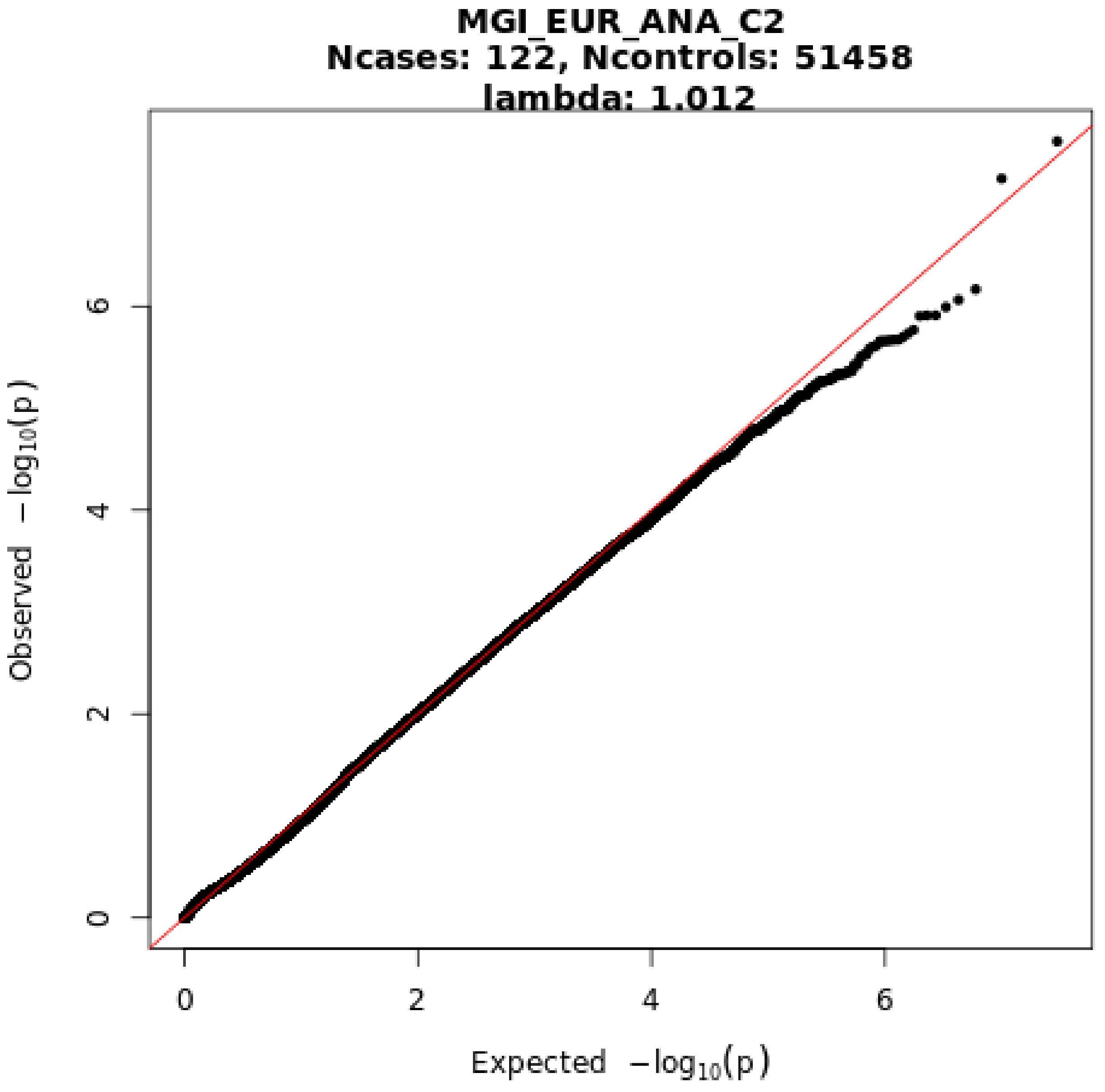
MGI_EUR_ANA_C2
Ncases: 122, Ncontrols: 51458
lambda: 1.012
Expected −log10(p)
Observed −log10(p)
0
2
4
6
0
2
4
6

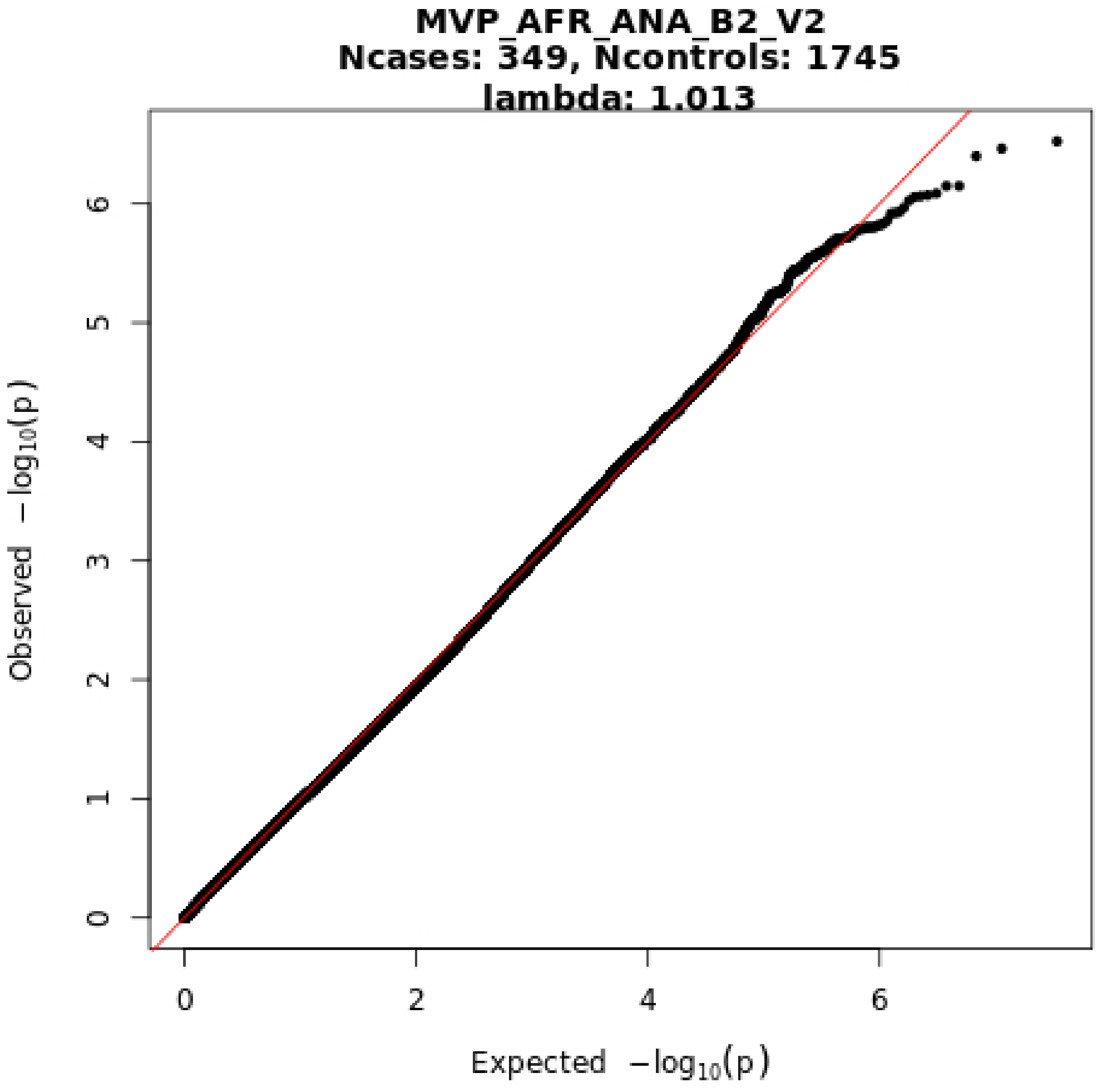
MVP_AFR_ANA_B2_V2
Ncases: 349, Ncontrols: 1745
lambda: 1.013
Observed $-\log_{10}(p)$
Expected $-\log_{10}(p)$
0
1
2
3
4
5
6
0
2
4
6

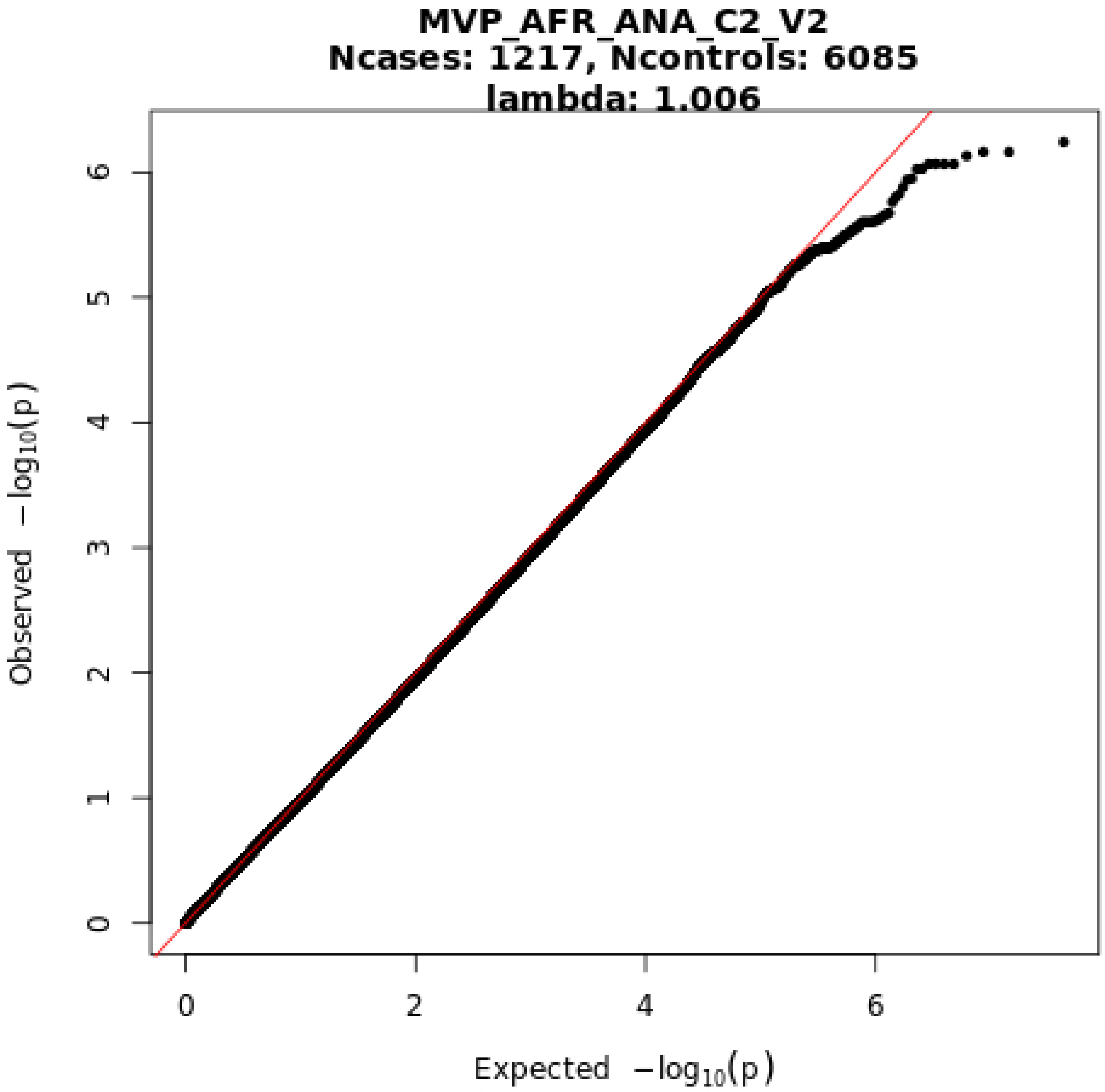

MVP_AFR_ANA_C2_V2
Ncases: 1217, Ncontrols: 6085
lambda: 1.006
Observed −log10(p)
Expected −log10(p)
0
1
2
3
4
5
6
0
2
4
6

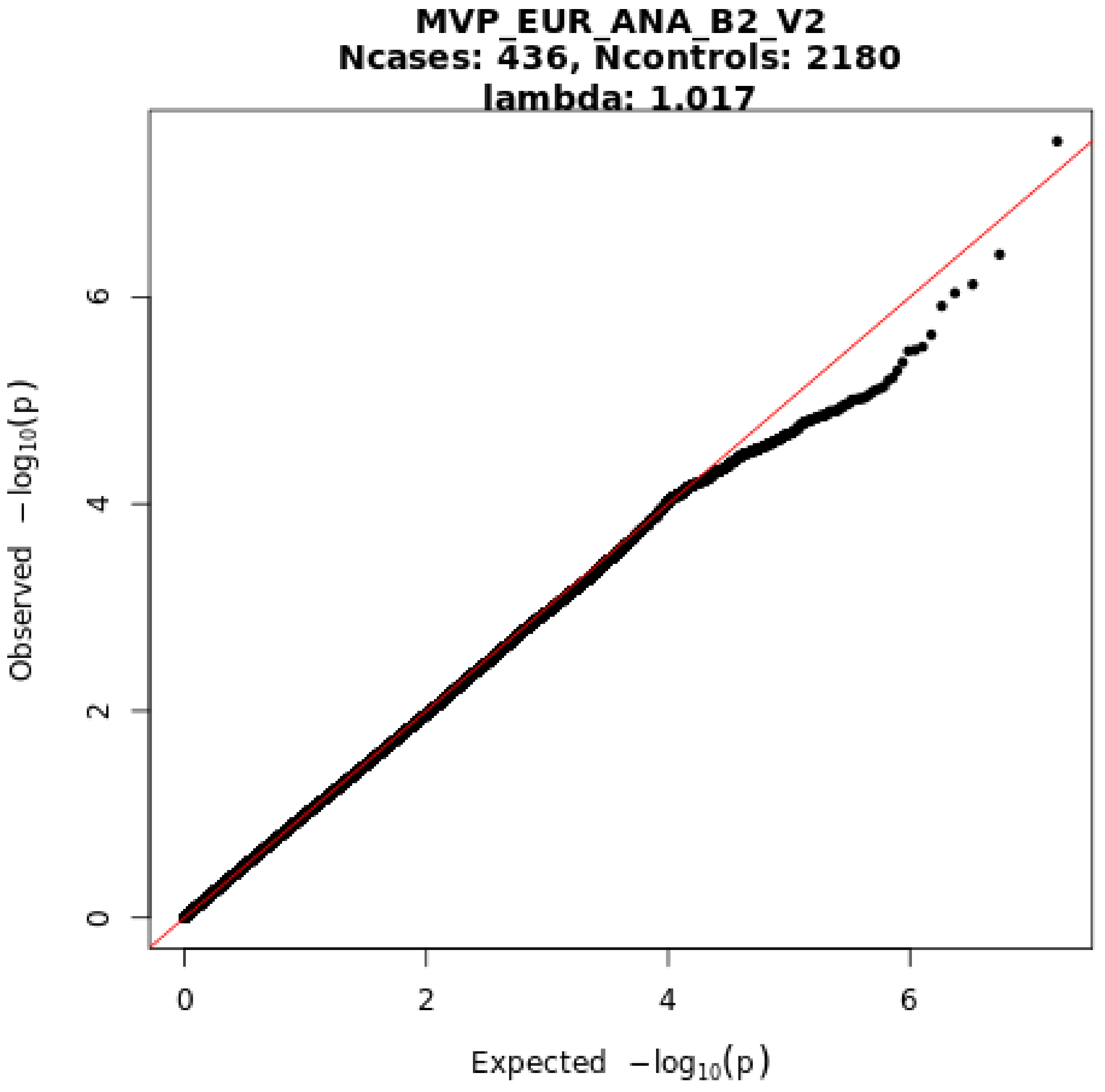

MVP_EUR_ANA_B2_V2
Ncases: 436, Ncontrols: 2180
lambda: 1.017
Observed −log10(p)
Expected −log10(p)
0
2
4
6

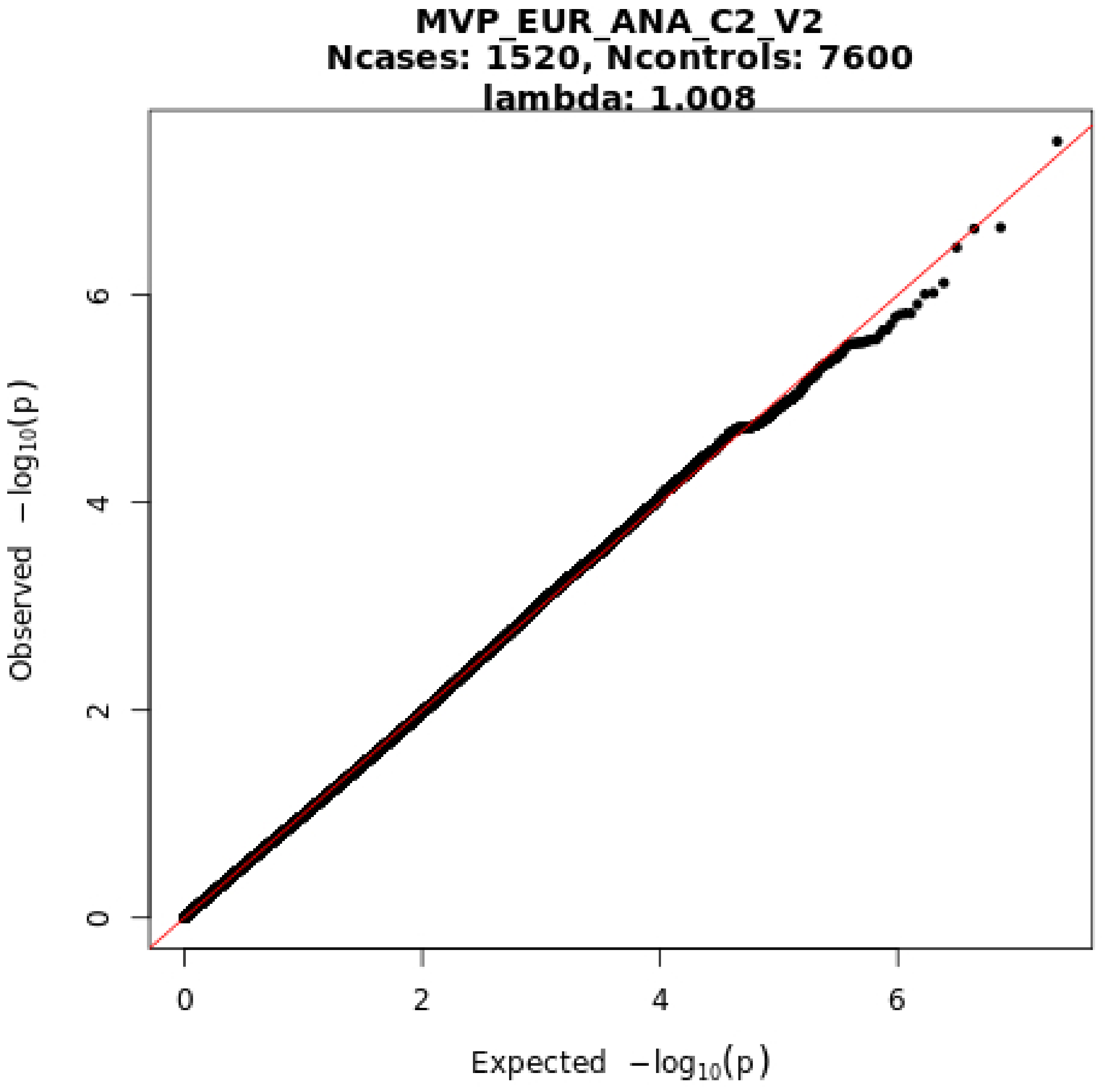
MVP_EUR_ANA_C2_V2
Ncases: 1520, Ncontrols: 7600
lambda: 1.008
Observed −log10(p)
Expected −log10(p)
0
2
4
6
0
2
4
6

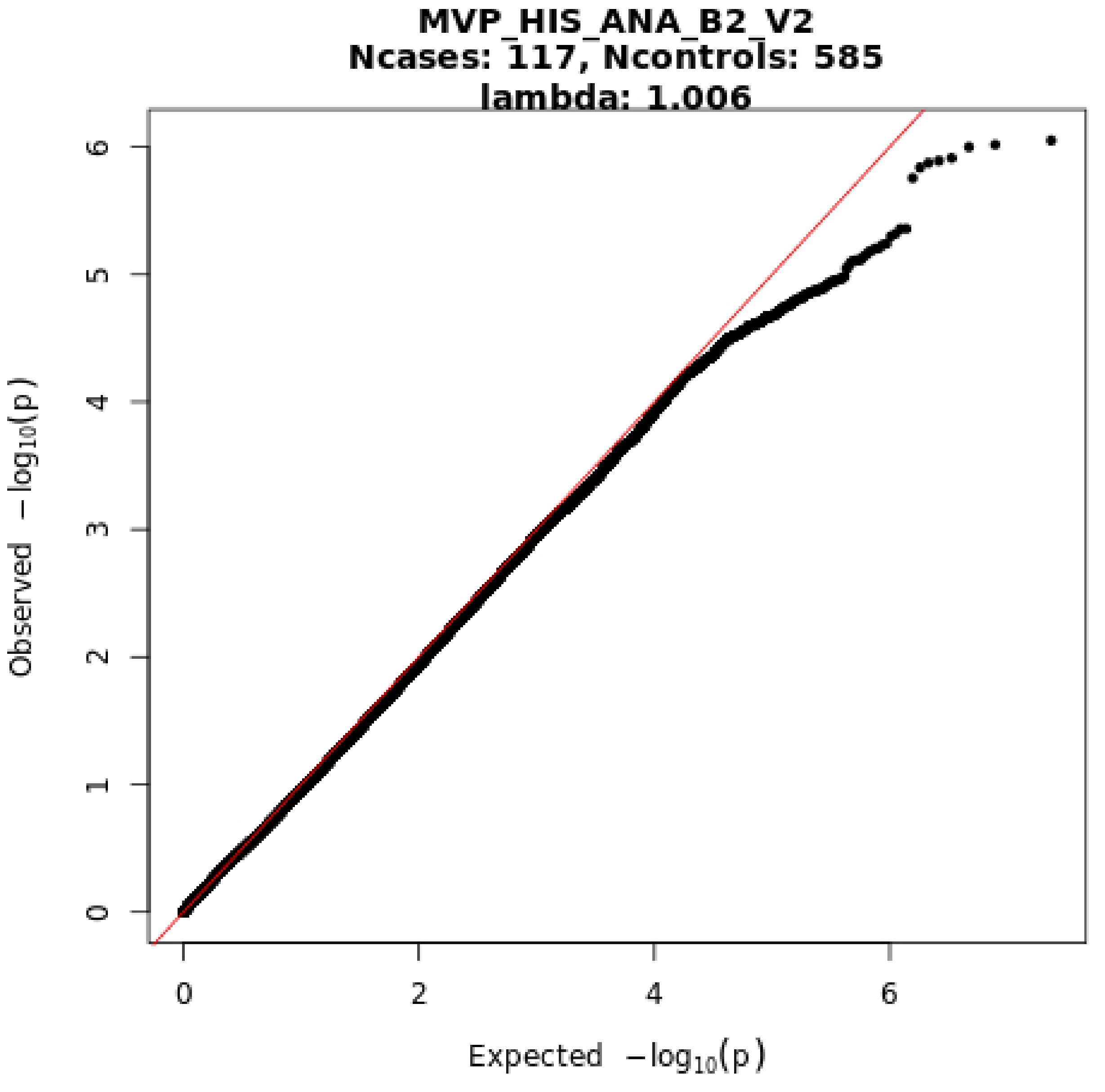
MVP_HIS_ANA_B2_V2
Ncases: 117, Ncontrols: 585
lambda: 1.006
Observed −log10(p)
Expected −log10(p)
0
1
2
3
4
5
6
0
2
4
6

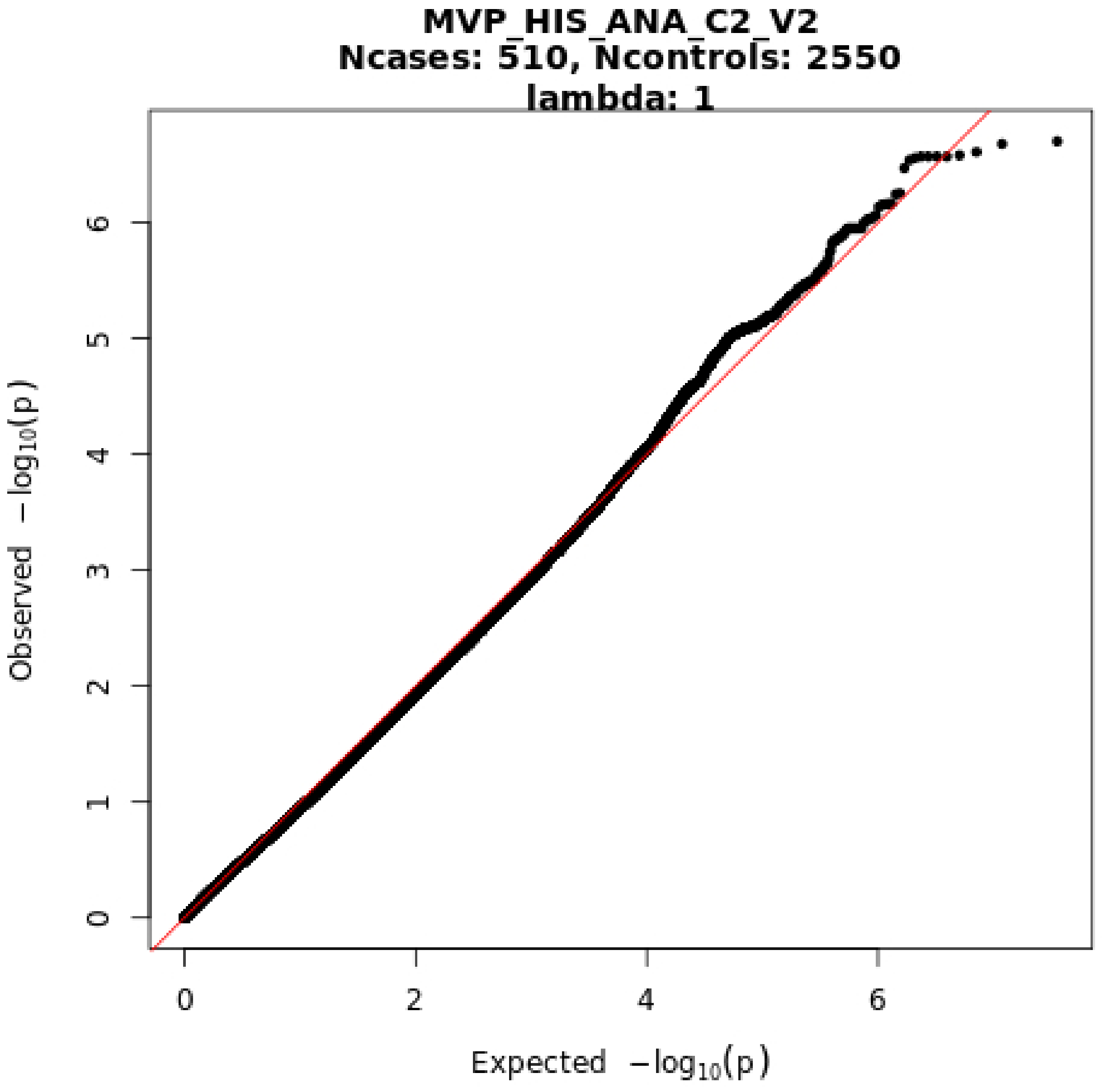
MVP_HIS_ANA_C2_V2
Ncases: 510, Ncontrols: 2550
lambda: 1
Observed −log10(p)
Expected −log10(p)
0
1
2
3
4
5
6
0
2
4
6

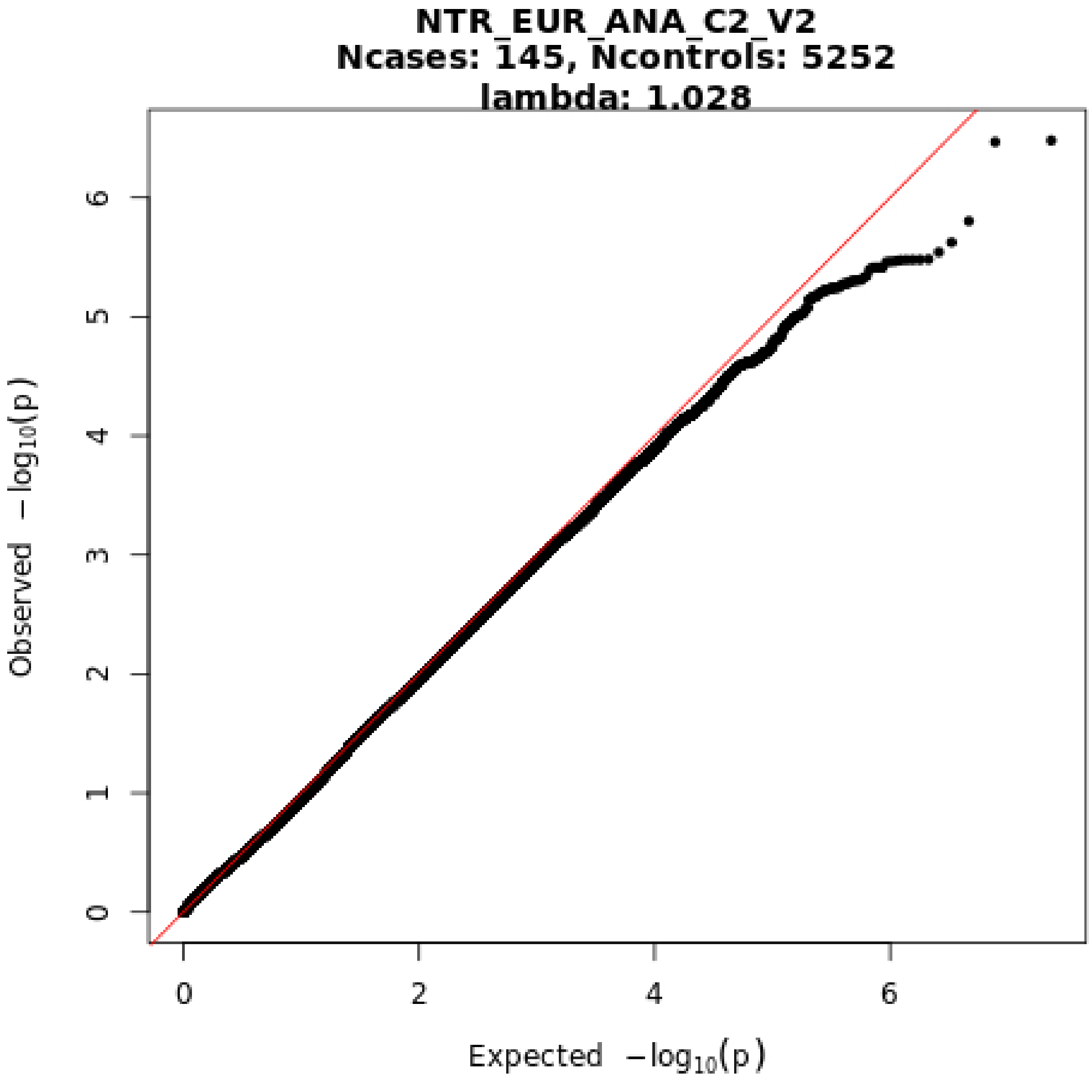
NTR_EUR_ANA_C2_V2
Ncases: 145, Ncontrols: 5252
lambda: 1.028
Observed −log10(p)
Expected −log10(p)
0
1
2
3
4
5
6
0
2
4
6

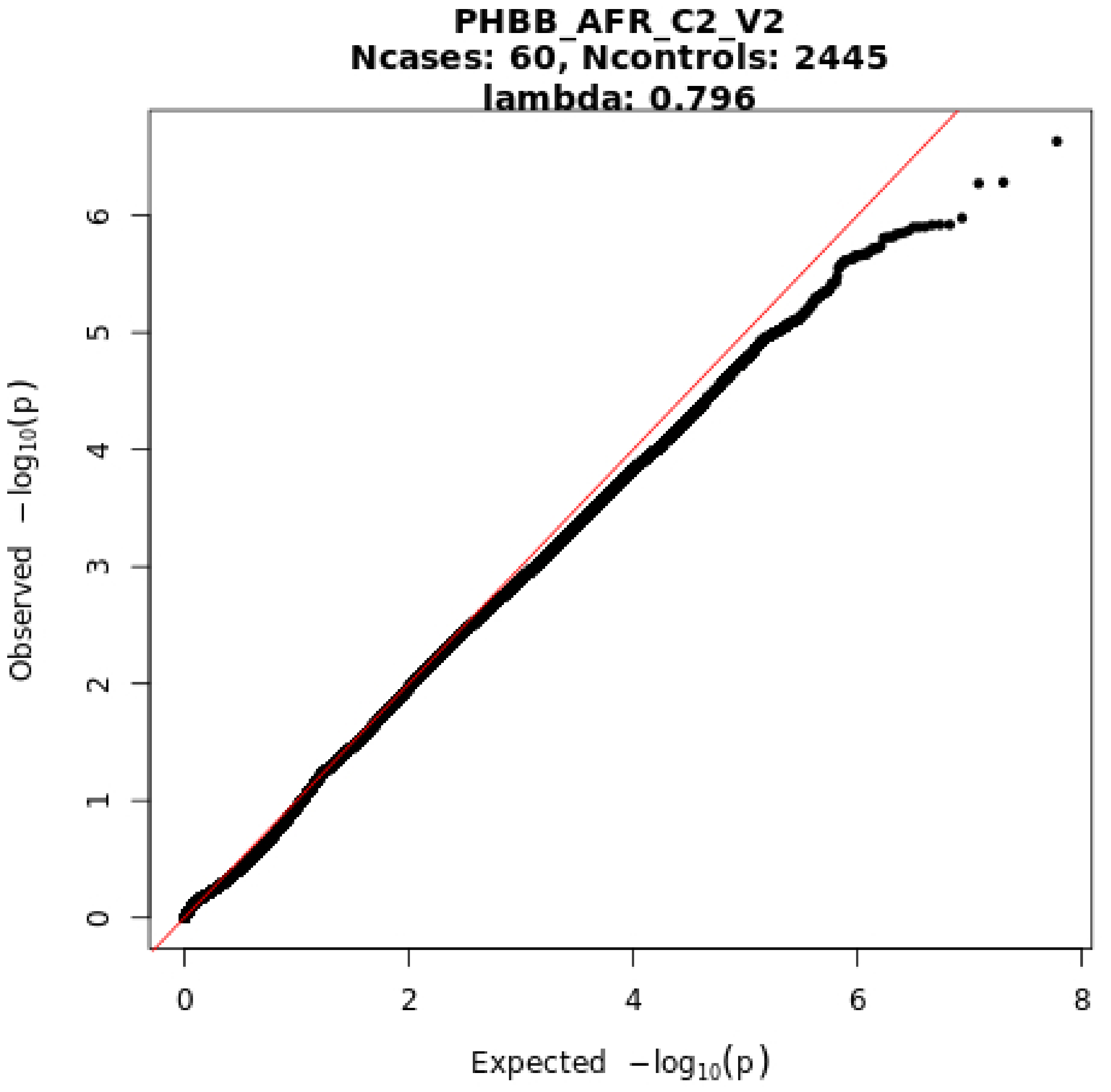
PHBB_AFR_C2_V2
Ncases: 60, Ncontrols: 2445
lambda: 0.796
Observed −log10(p)
Expected −log10(p)
0
1
2
3
4
5
6
0
2
4
6
8

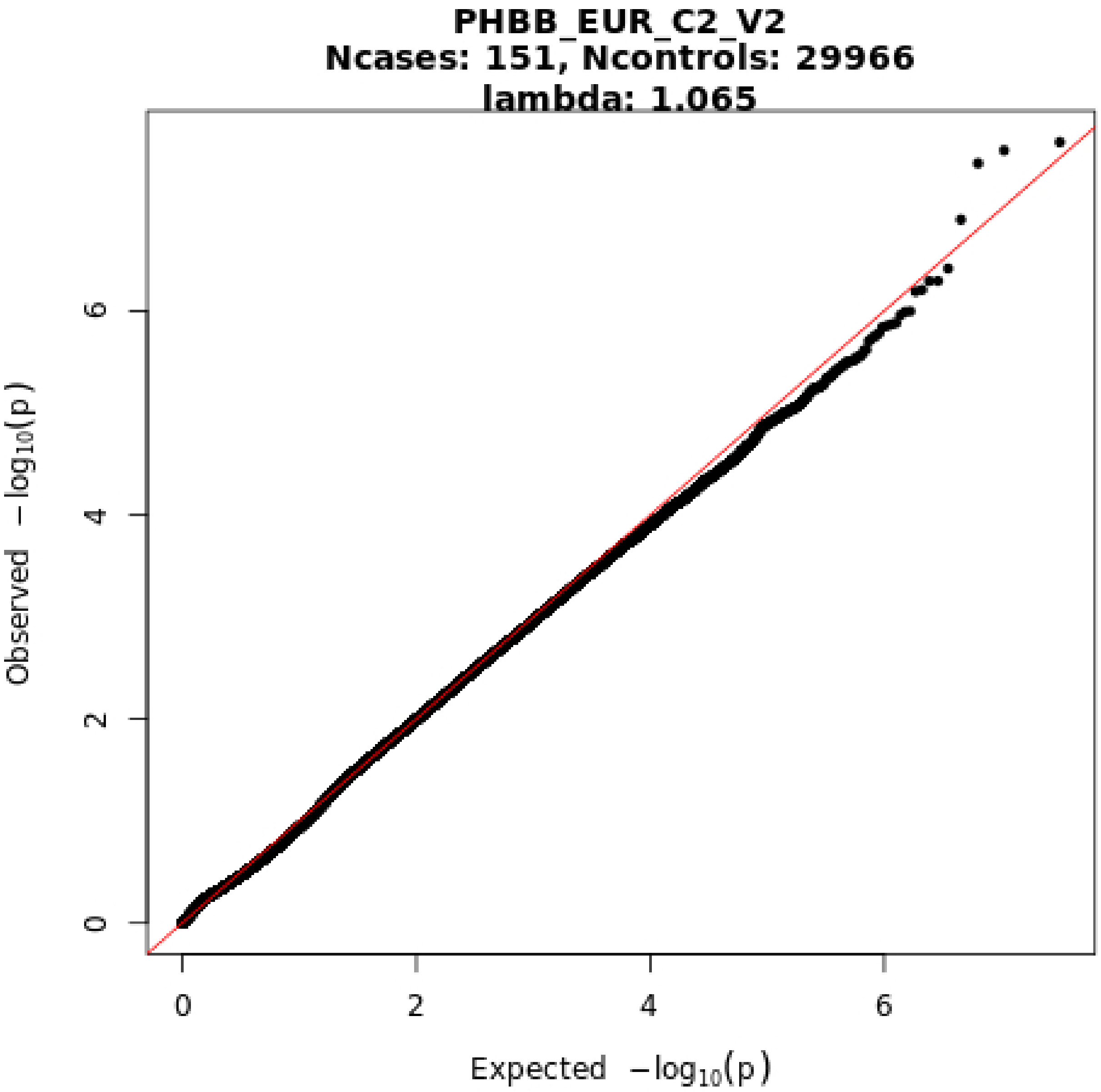
PHBB_EUR_C2_V2
Ncases: 151, Ncontrols: 29966
lambda: 1.065
Observed −log10(p)
Expected −log10(p)
0
2
4
6
0
2
4
6

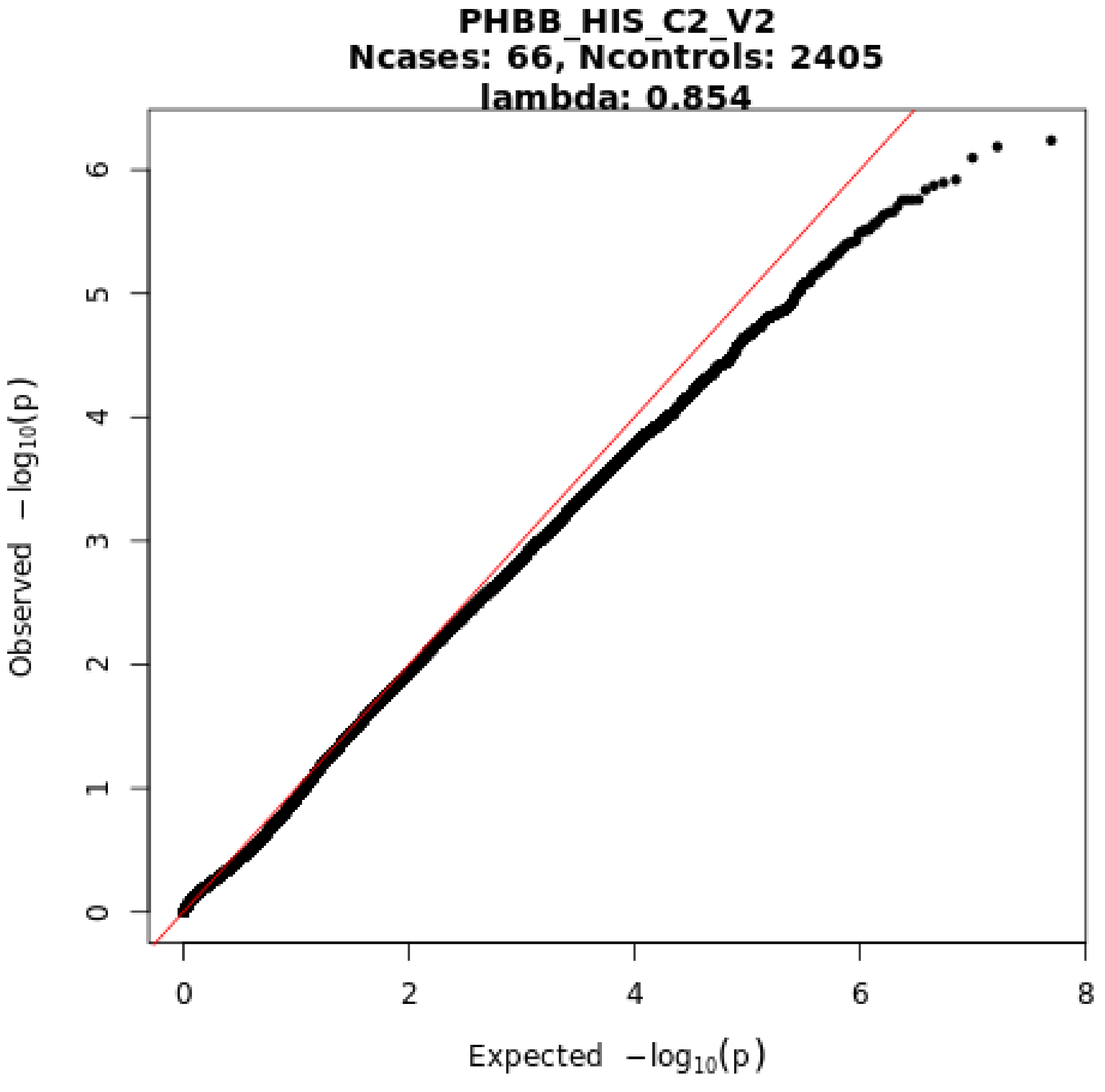
PHBB_HIS_C2_V2
Ncases: 66, Ncontrols: 2405
lambda: 0.854
Observed −log10(p)
0
1
2
3
4
5
6
Expected −log10(p)
0
2
4
6
8

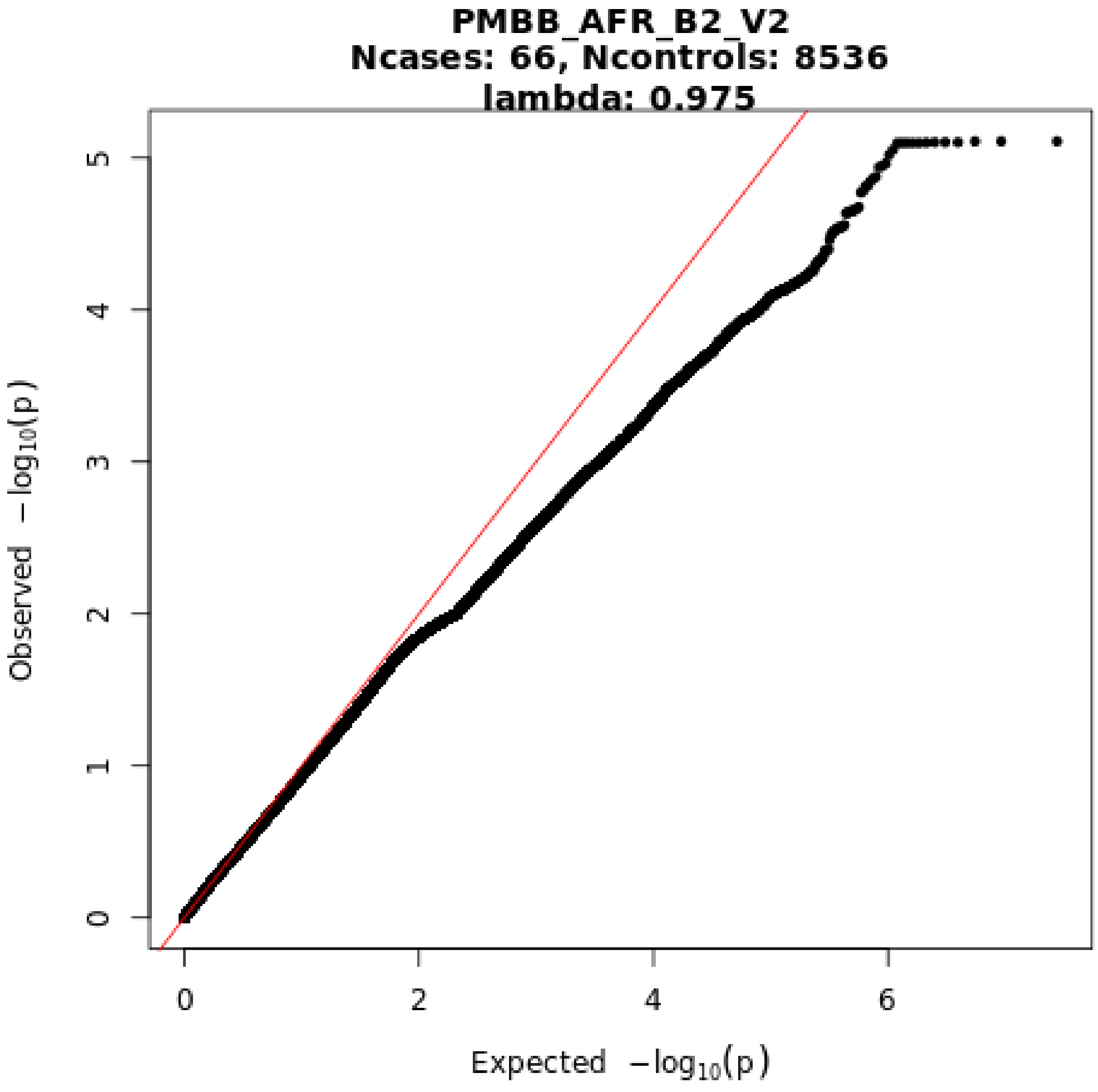
PMBB_AFR_B2_V2
Ncases: 66, Ncontrols: 8536
lambda: 0.975
Observed −log10(p)
Expected −log10(p)
0
1
2
3
4
5
0
2
4
6

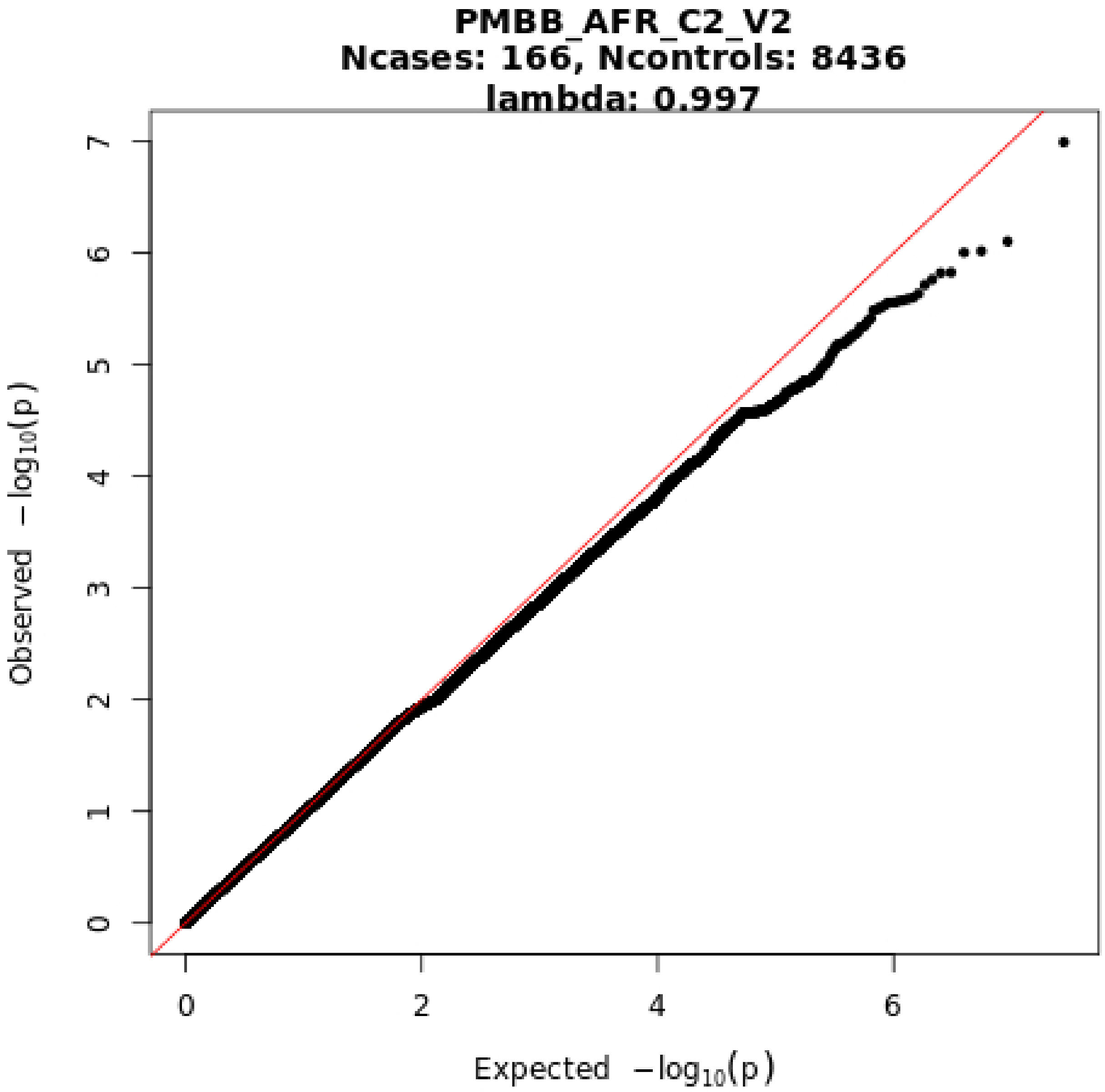
PMBB_AFR_C2_V2
Ncases: 166, Ncontrols: 8436
lambda: 0.997
Observed −log10(p)
Expected −log10(p)
0
1
2
3
4
5
6
7
0
2
4
6

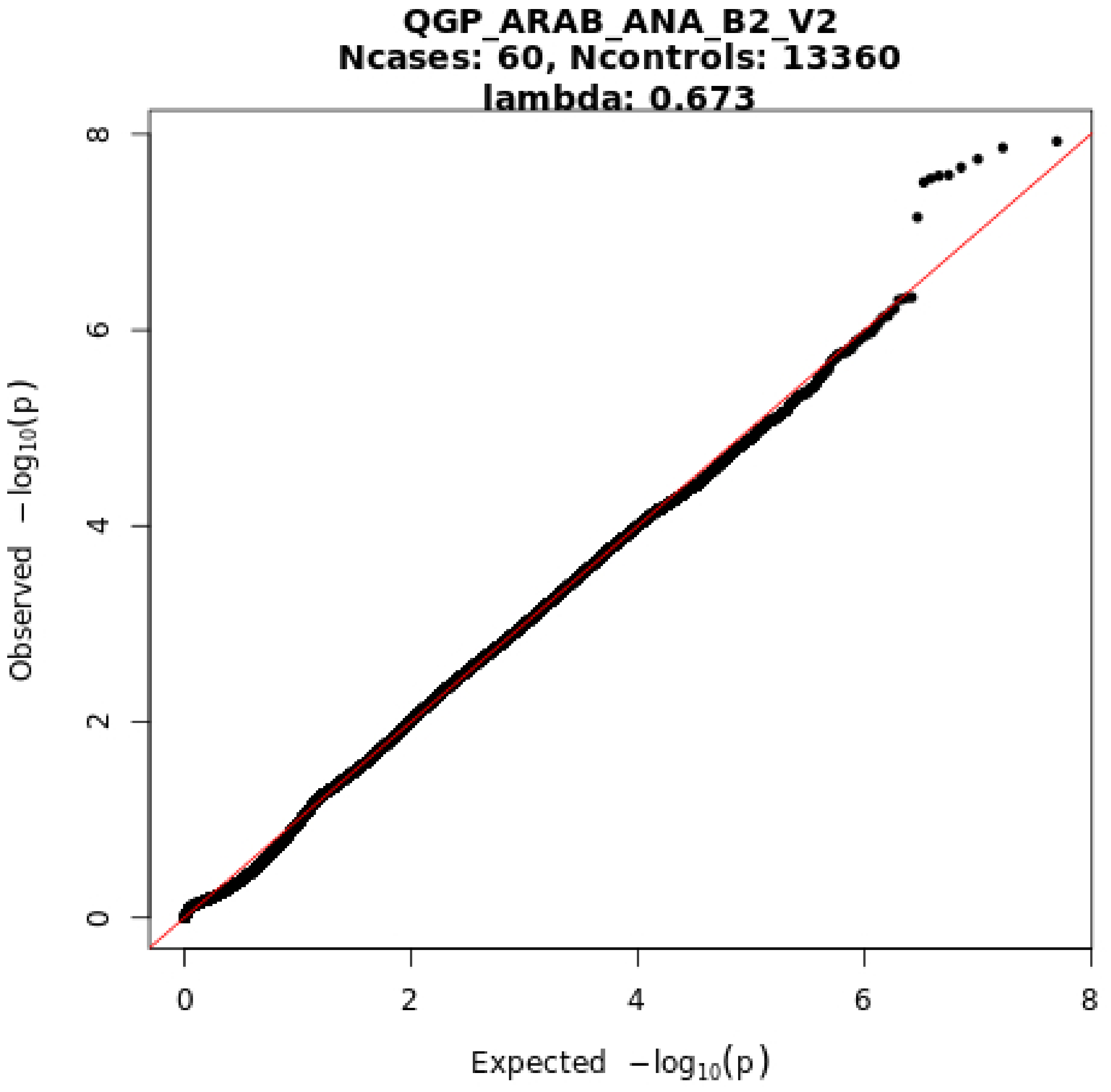
QGP_ARAB_ANA_B2_V2
Ncases: 60, Ncontrols: 13360
lambda: 0.673
Observed −log10(p)
Expected −log10(p)
0
2
4
6
8
0
2
4
6
8

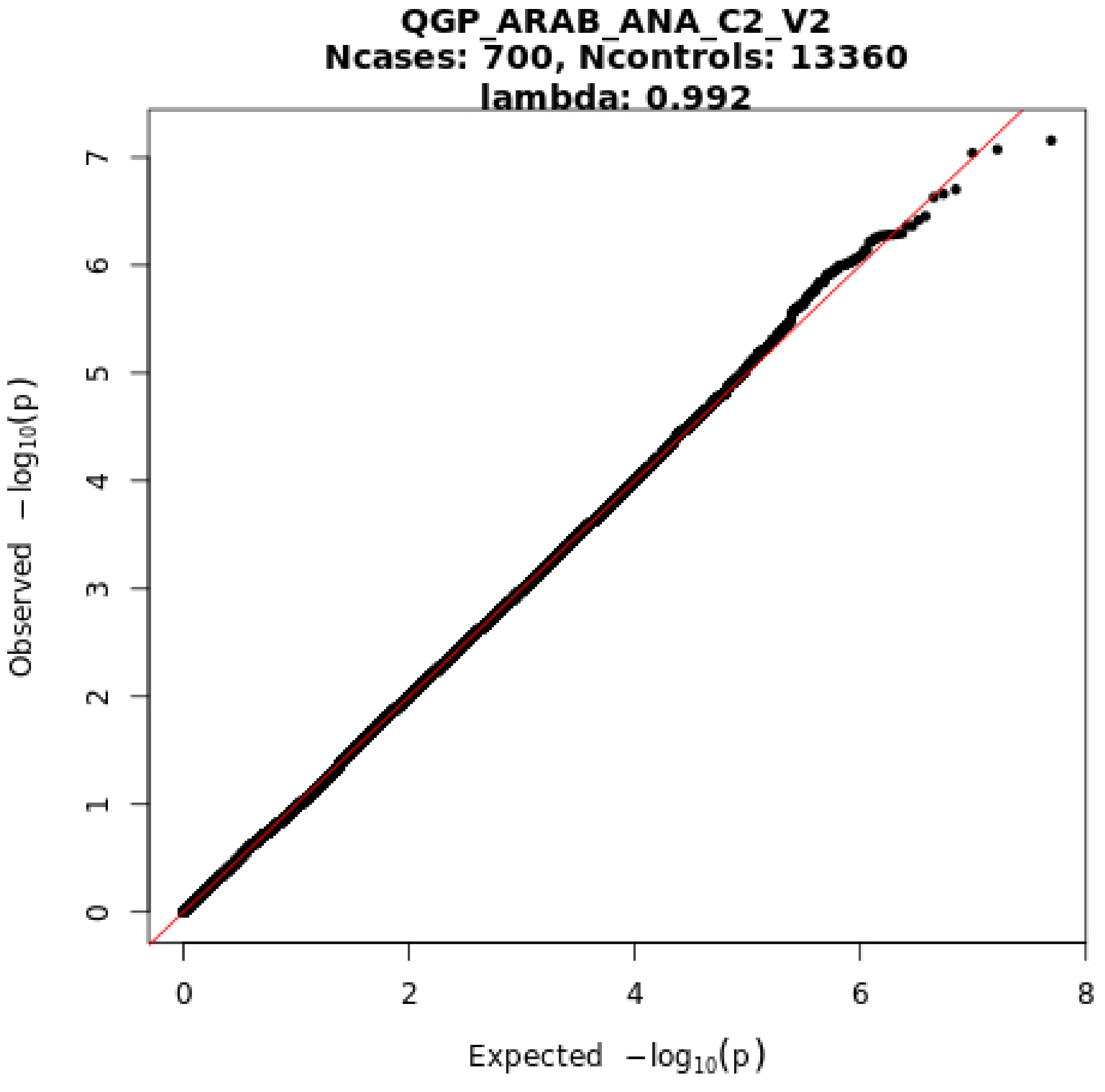
QGP_ARAB_ANA_C2_V2
Ncases: 700, Ncontrols: 13360
lambda: 0.992
Observed −log10(p)
Expected −log10(p)
0
1
2
3
4
5
6
7
0
2
4
6
8

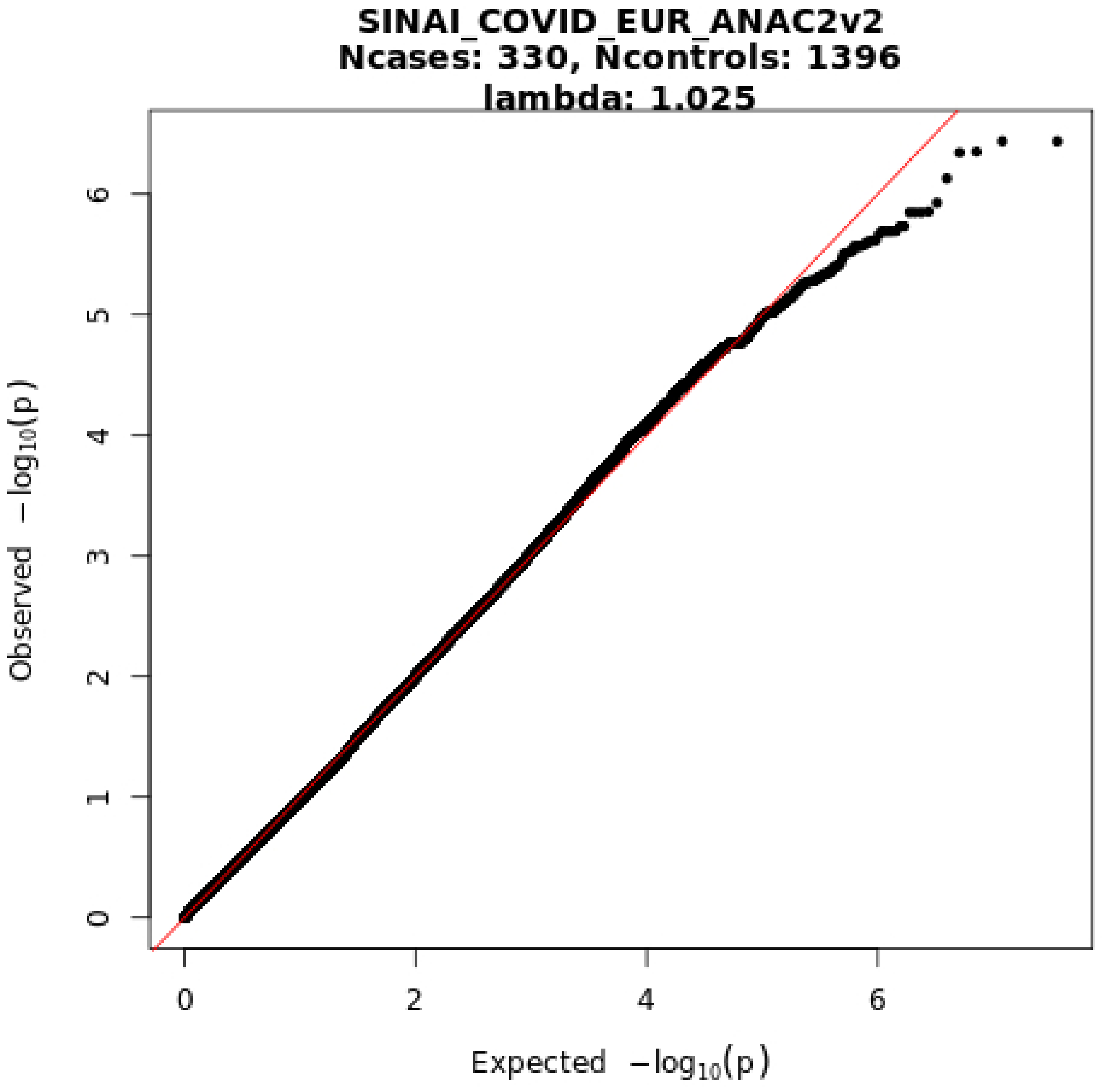
SINAI_COVID_EUR_ANAC2v2
Ncases: 330, Ncontrols: 1396
lambda: 1.025
Observed $-\log_{10}(p)$
Expected $-\log_{10}(p)$
0
1
2
3
4
5
6
0
2
4
6

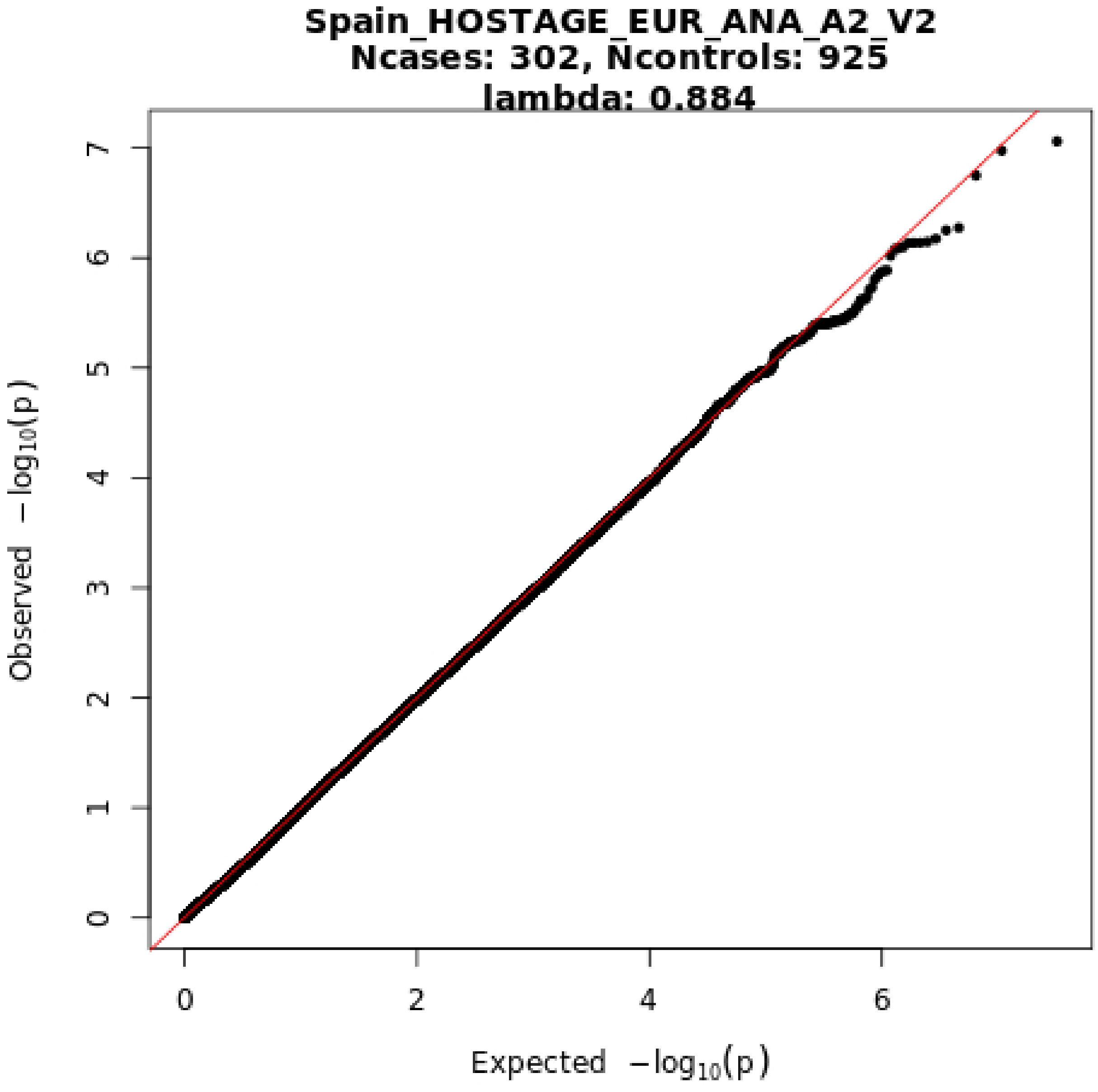

Spain_HOSTAGE_EUR_ANA_A2_V2
Ncases: 302, Ncontrols: 925
lambda: 0.884
Observed −log10(p)
Expected −log10(p)
0
1
2
3
4
5
6
7
0
2
4
6

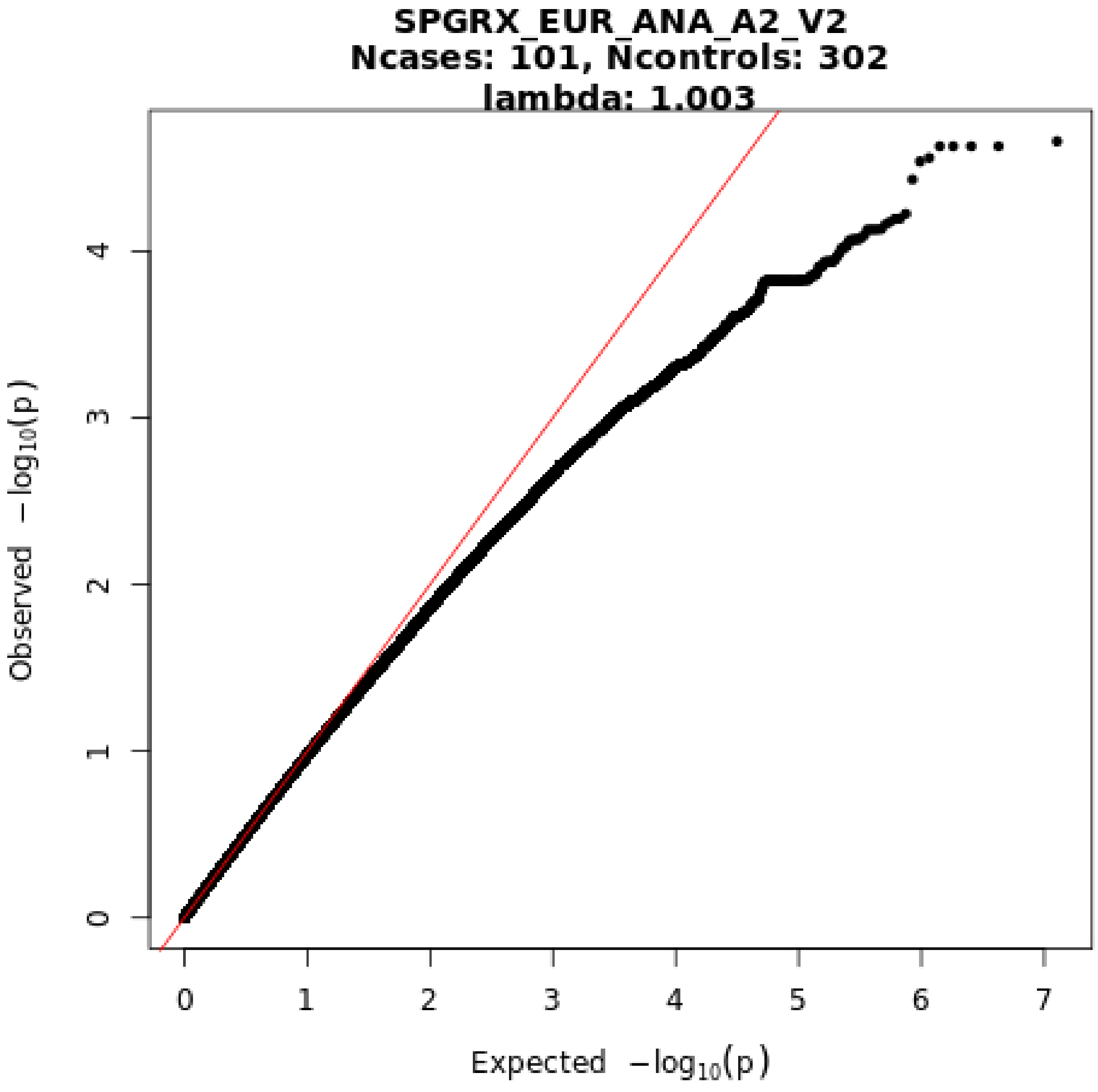
SPGRX_EUR_ANA_A2_V2
Ncases: 101, Ncontrols: 302
lambda: 1.003
Observed −log10(p)
Expected −log10(p)
0
1
2
3
4
5
6
7
0
1
2
3
4

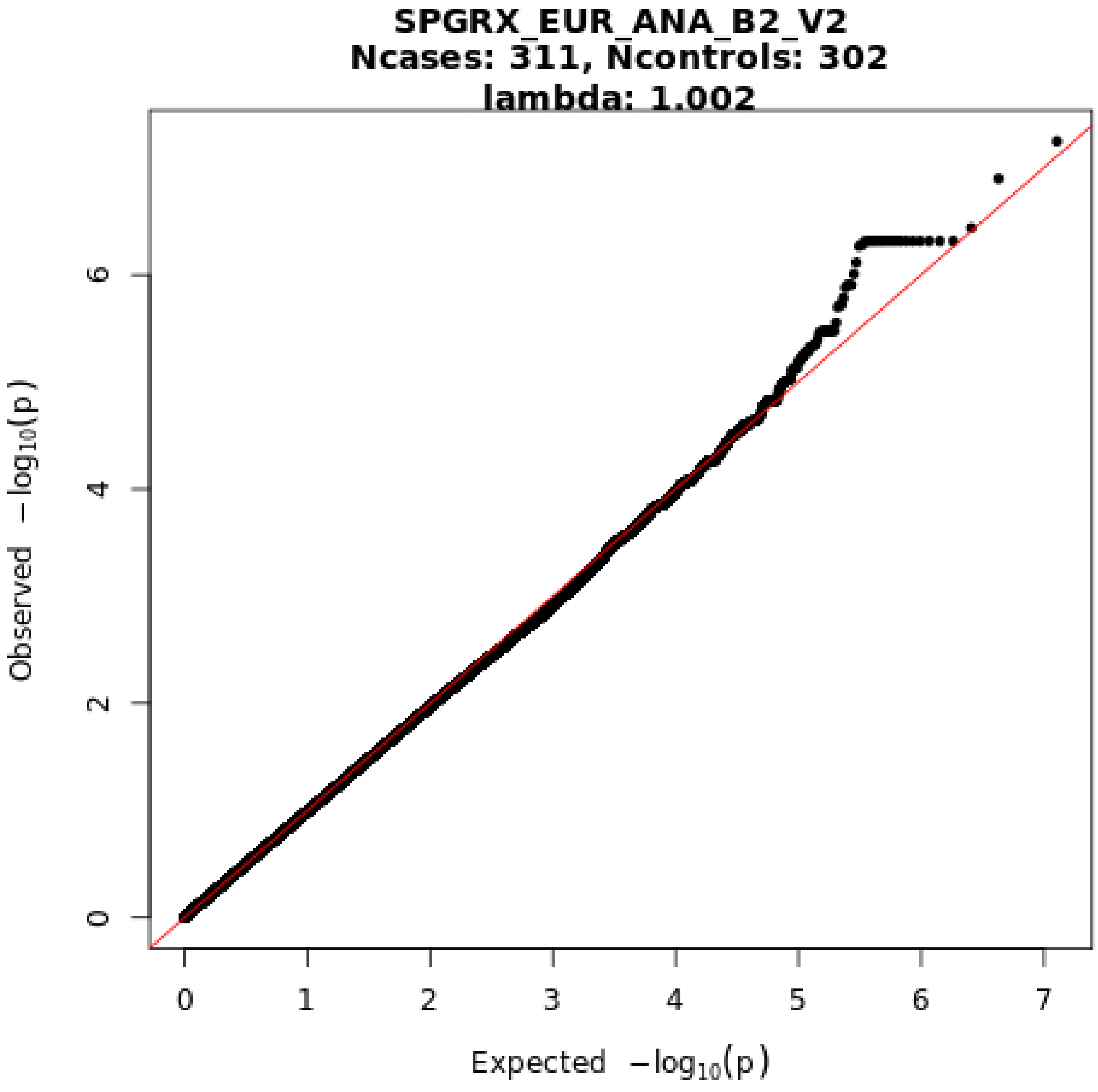
SPGRX_EUR_ANA_B2_V2
Ncases: 311, Ncontrols: 302
lambda: 1.002
Observed $-\log_{10}(p)$
Expected $-\log_{10}(p)$
0
2
4
6
0
1
2
3
4
5
6
7

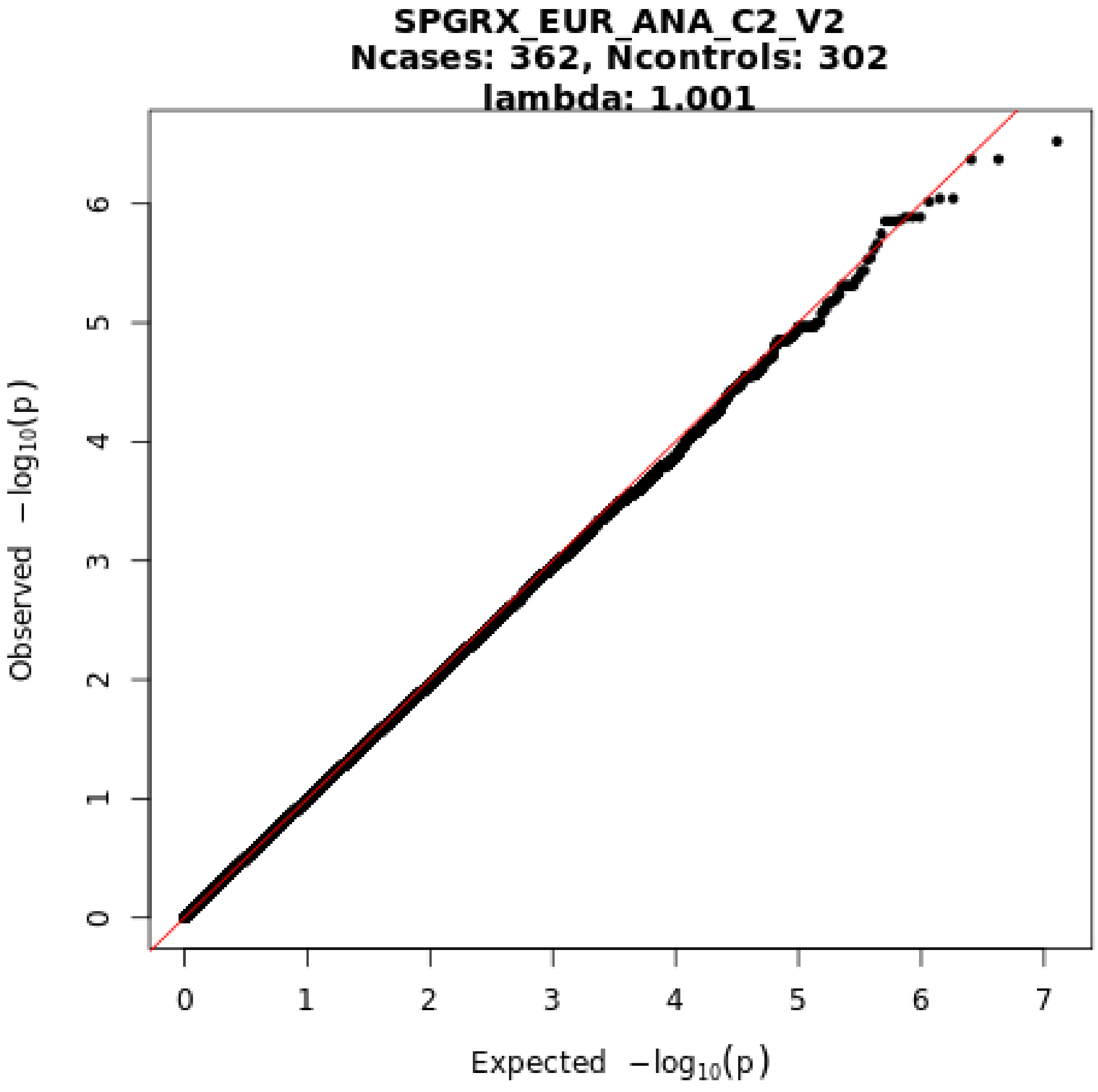

SPGRX_EUR_ANA_C2_V2
Ncases: 362, Ncontrols: 302
lambda: 1.001
Observed −log10(p)
Expected −log10(p)
0
1
2
3
4
5
6
7

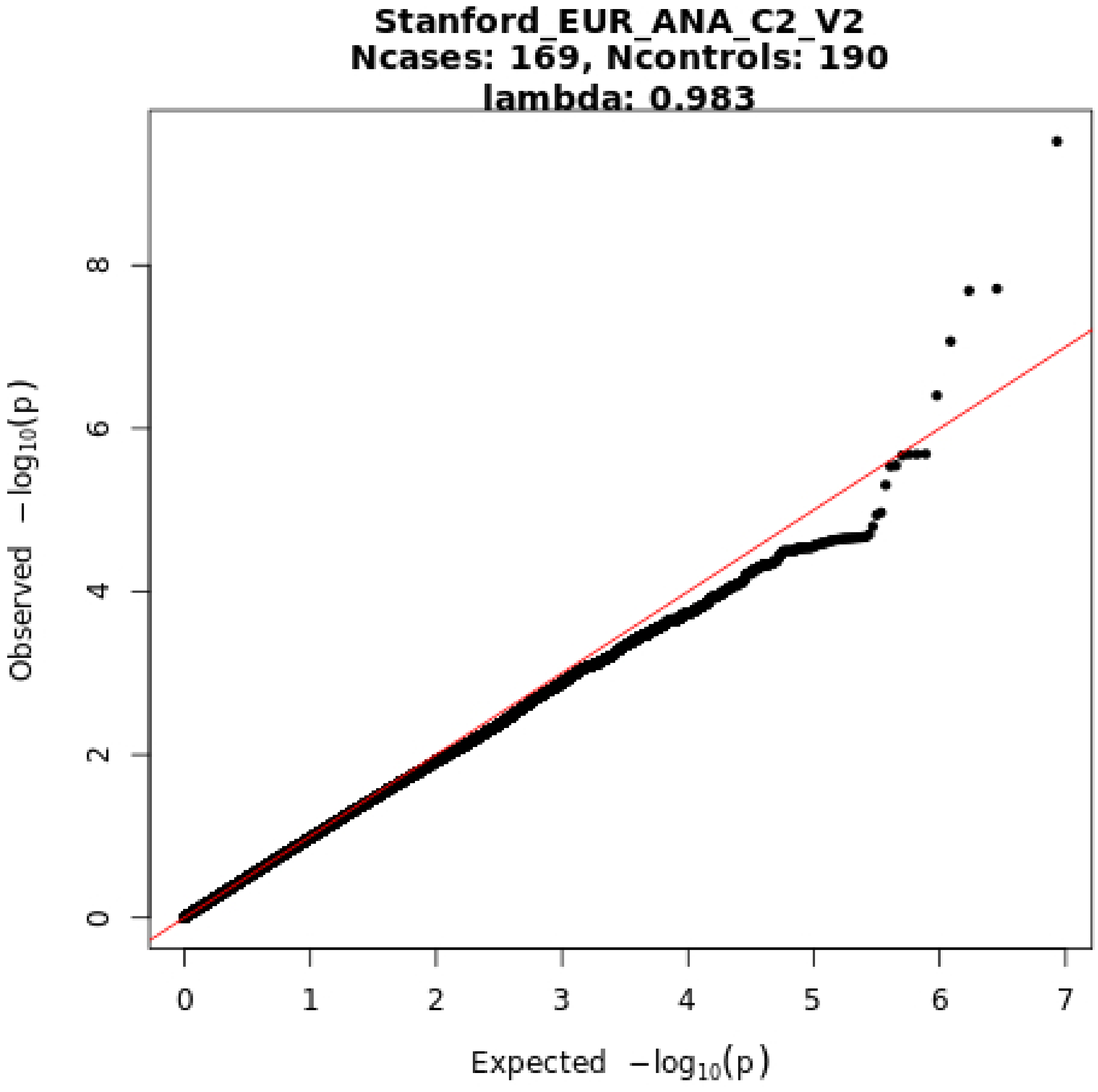
Stanford_EUR_ANA_C2_V2
Ncases: 169, Ncontrols: 190
lambda: 0.983
0
2
4
6
8
0
1
2
3
4
5
6
7
Expected −log10(p)
Observed −log10(p)

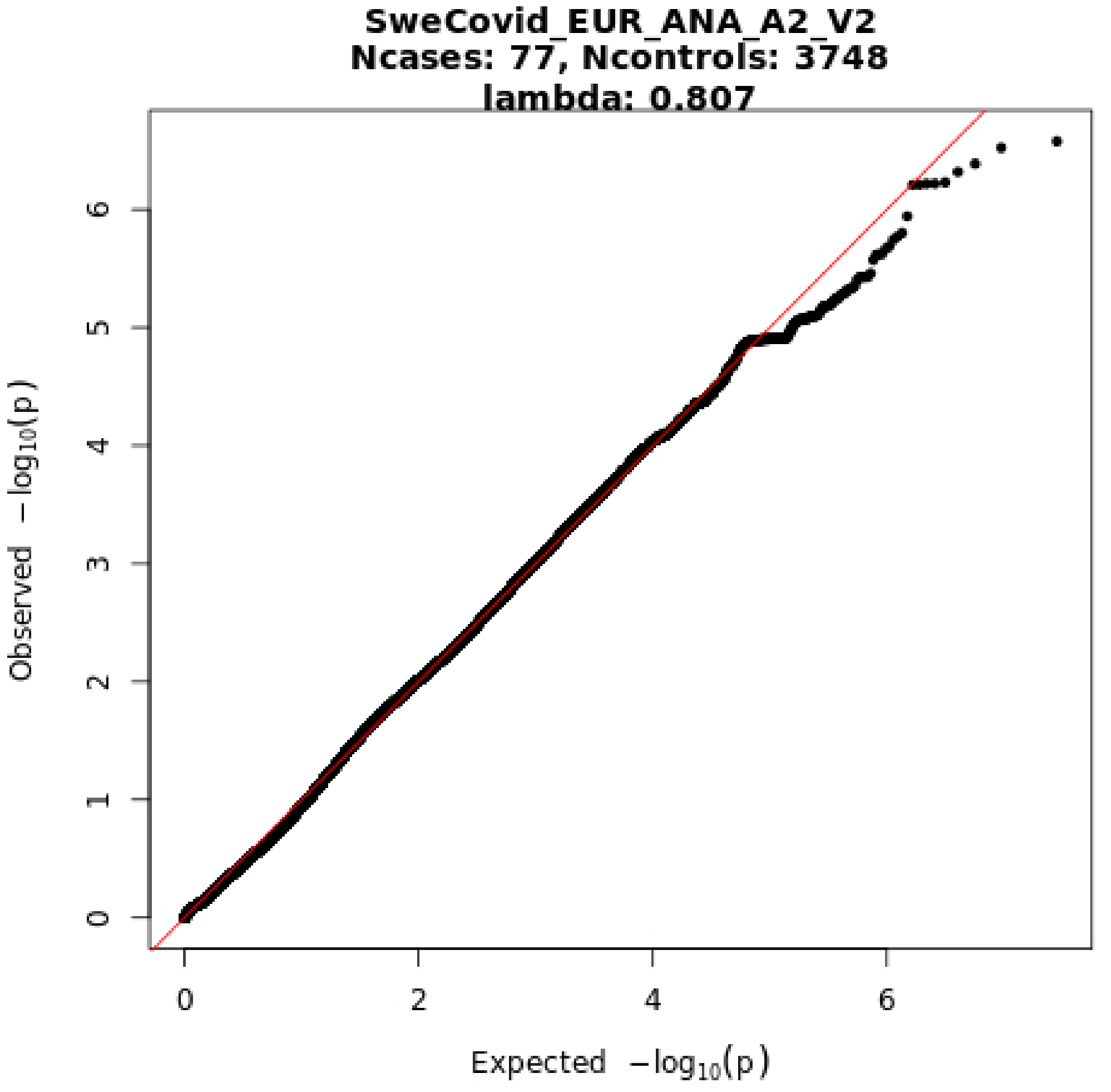
SweCovid_EUR_ANA_A2_V2
Ncases: 77, Ncontrols: 3748
lambda: 0.807
Observed −log10(p)
Expected −log10(p)
0
1
2
3
4
5
6
0
2
4
6

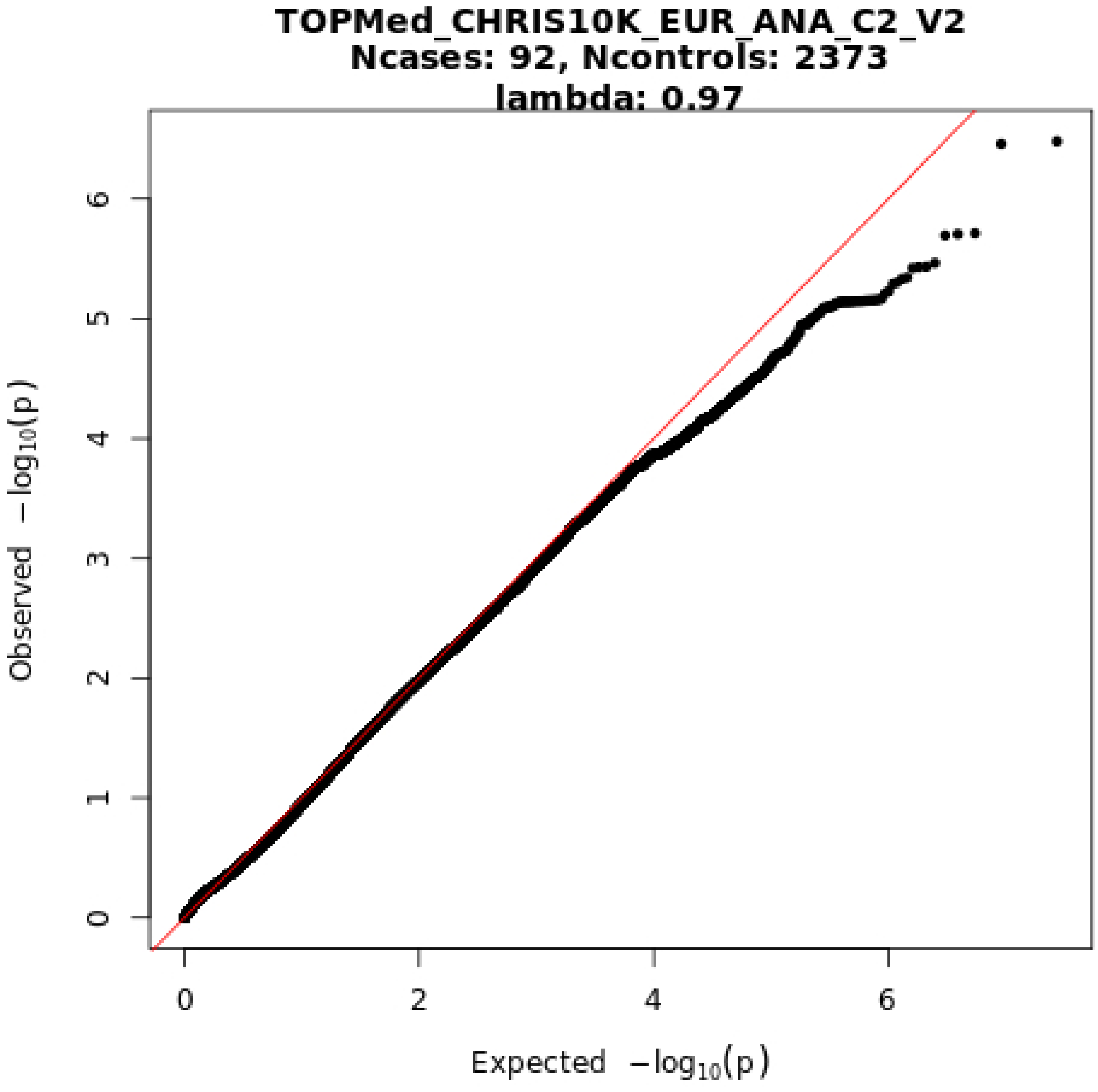
TOPMed_CHRIS10K_EUR_ANA_C2_V2
Ncases: 92, Ncontrols: 2373
lambda: 0.97
Observed −log10(p)
Expected −log10(p)
0
1
2
3
4
5
6
0
2
4
6

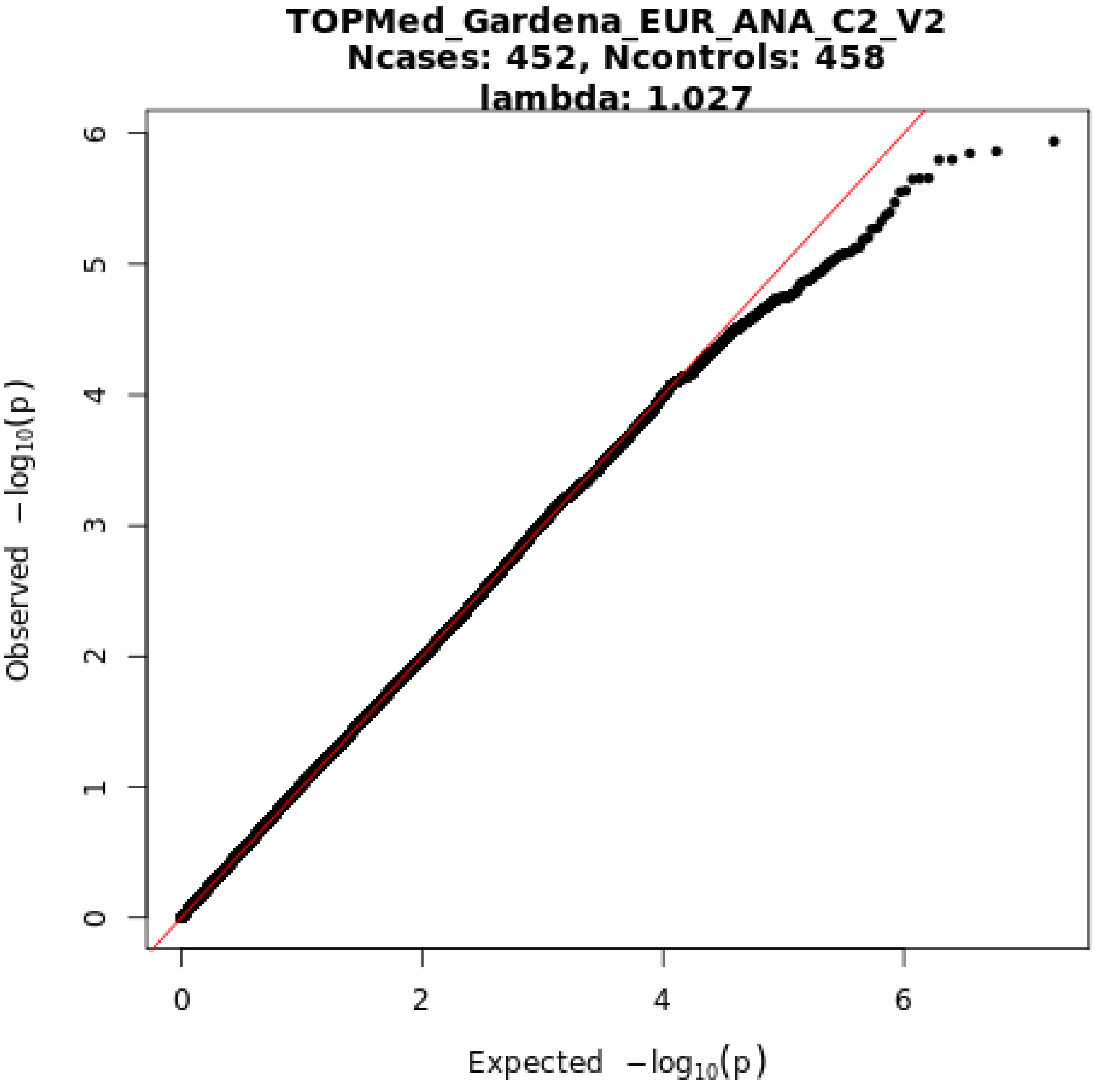

TOPMed_Gardena_EUR_ANA_C2_V2
Ncases: 452, Ncontrols: 458
lambda: 1.027
Observed −log10(p)
Expected −log10(p)
0
1
2
3
4
5
6
0
2
4
6

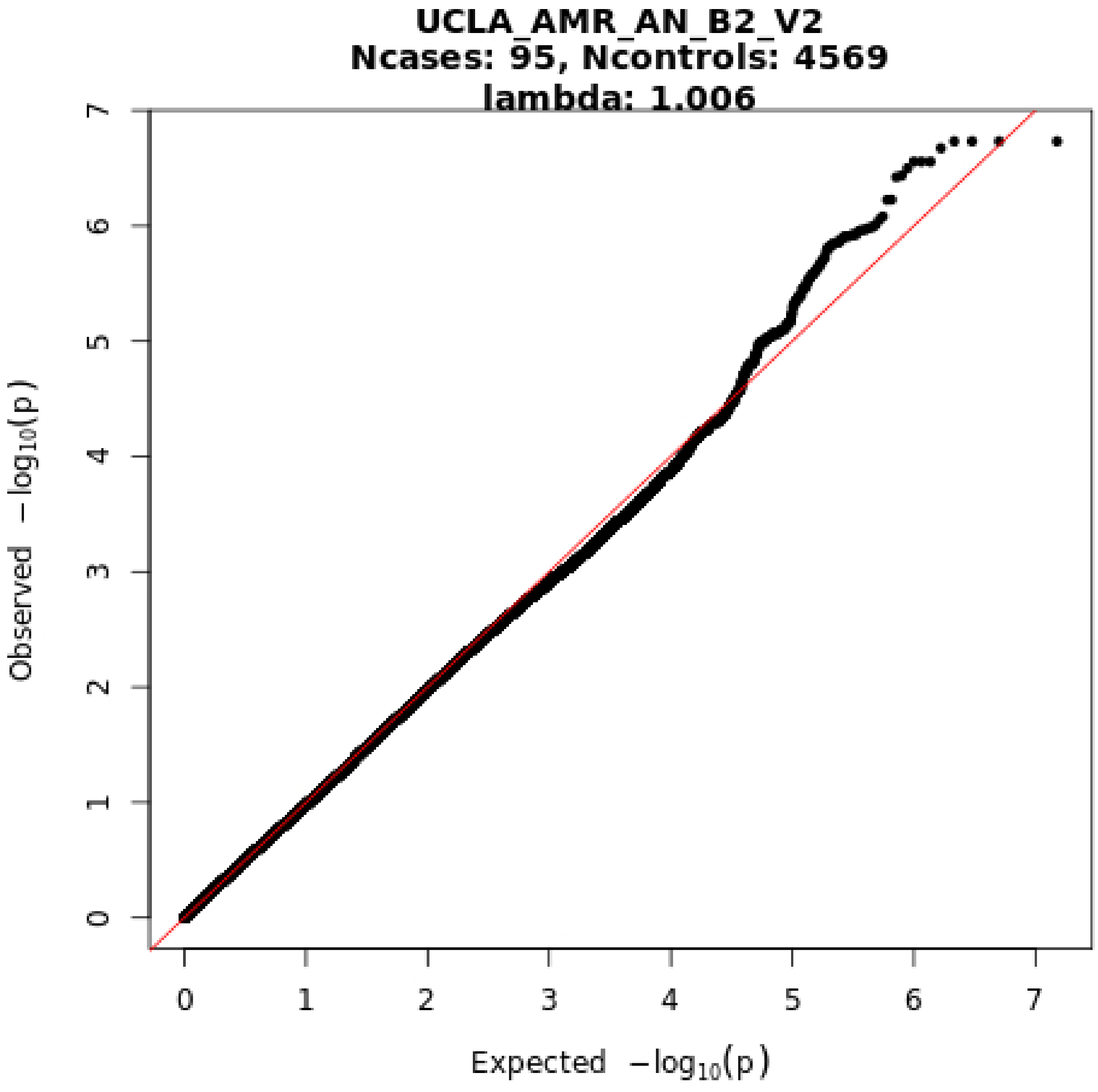

UCLA_AMR_AN_B2_V2
Ncases: 95, Ncontrols: 4569
lambda: 1.006
Observed −log10(p)
Expected −log10(p)
0
1
2
3
4
5
6
7

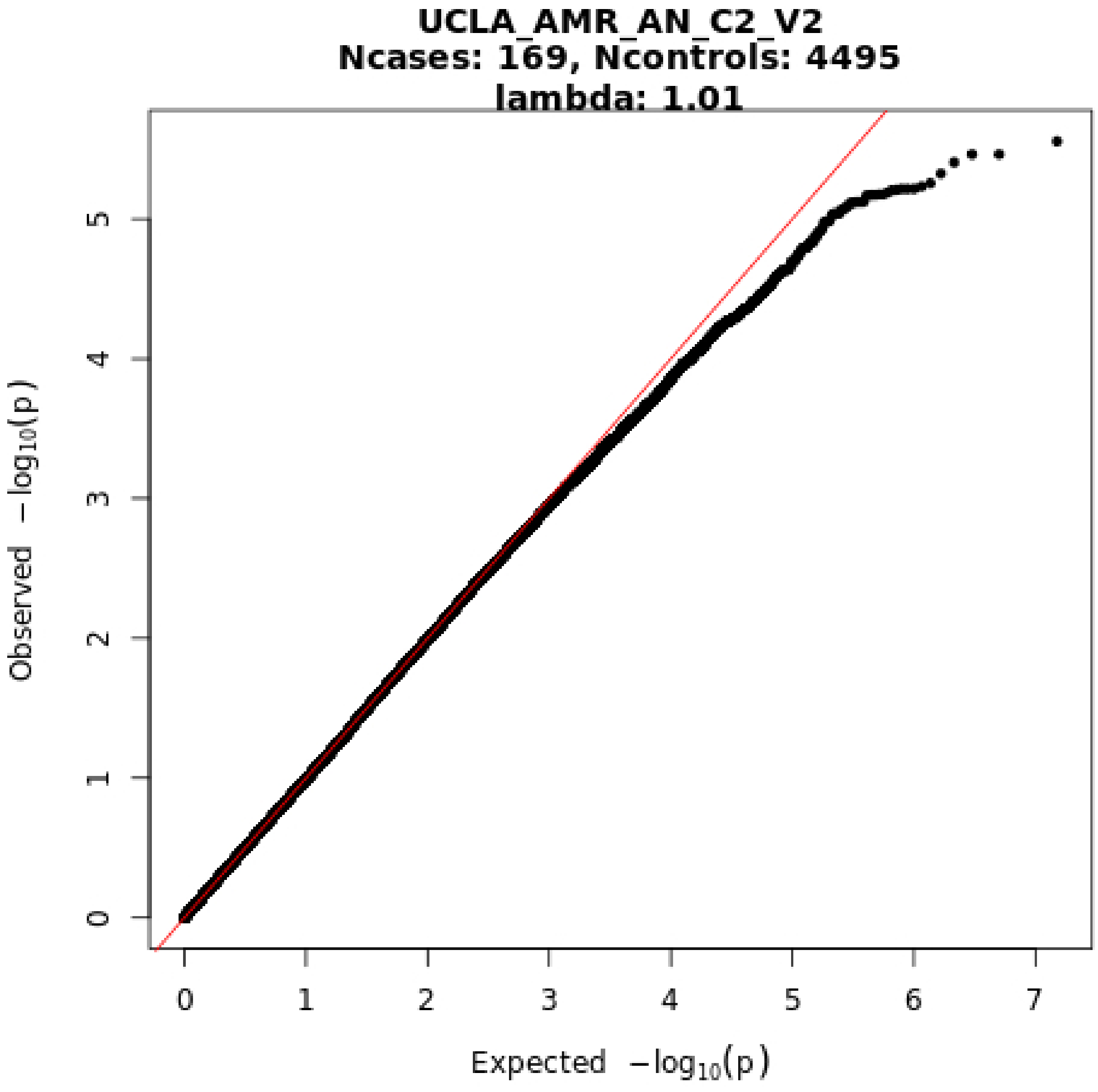
UCLA_AMR_AN_C2_V2
Ncases: 169, Ncontrols: 4495
lambda: 1.01
Observed −log10(p)
Expected −log10(p)
0
1
2
3
4
5
6
7

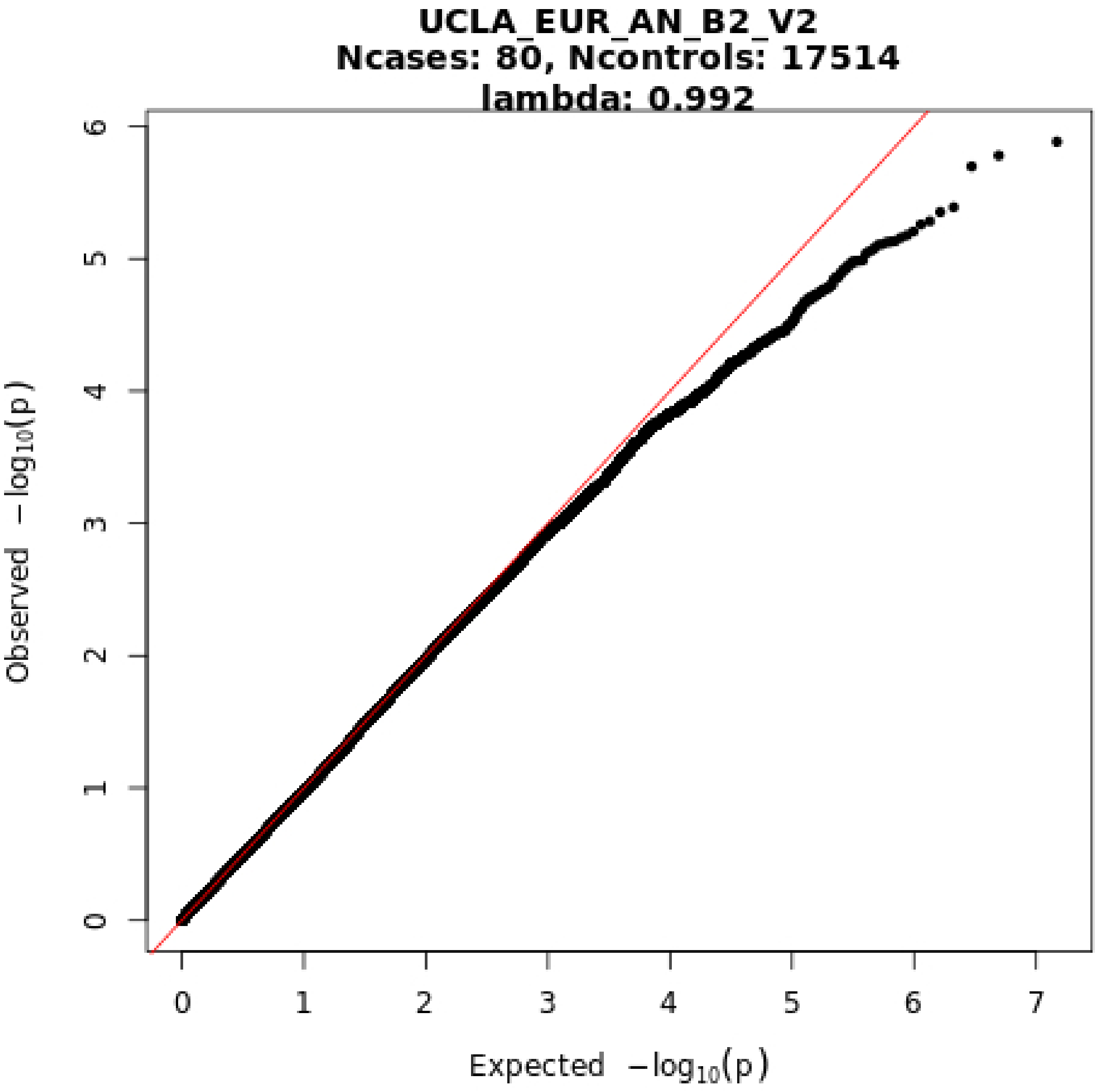
UCLA_EUR_AN_B2_V2
Ncases: 80, Ncontrols: 17514
lambda: 0.992
Observed −log10(p)
Expected −log10(p)
0
1
2
3
4
5
6
7

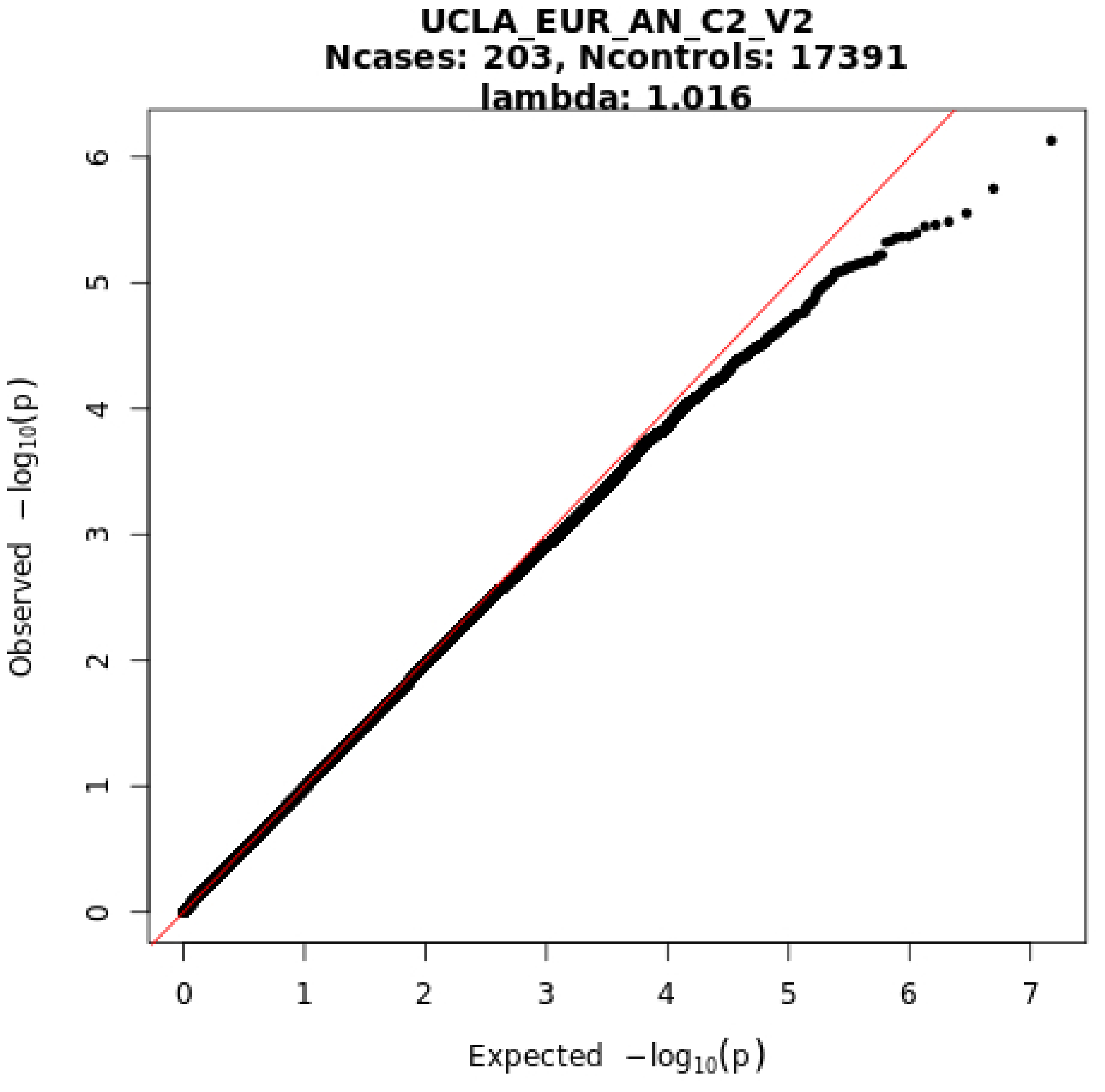

UCLA_EUR_AN_C2_V2
Ncases: 203, Ncontrols: 17391
lambda: 1.016
Observed −log10(p)
Expected −log10(p)
0
1
2
3
4
5
6
7

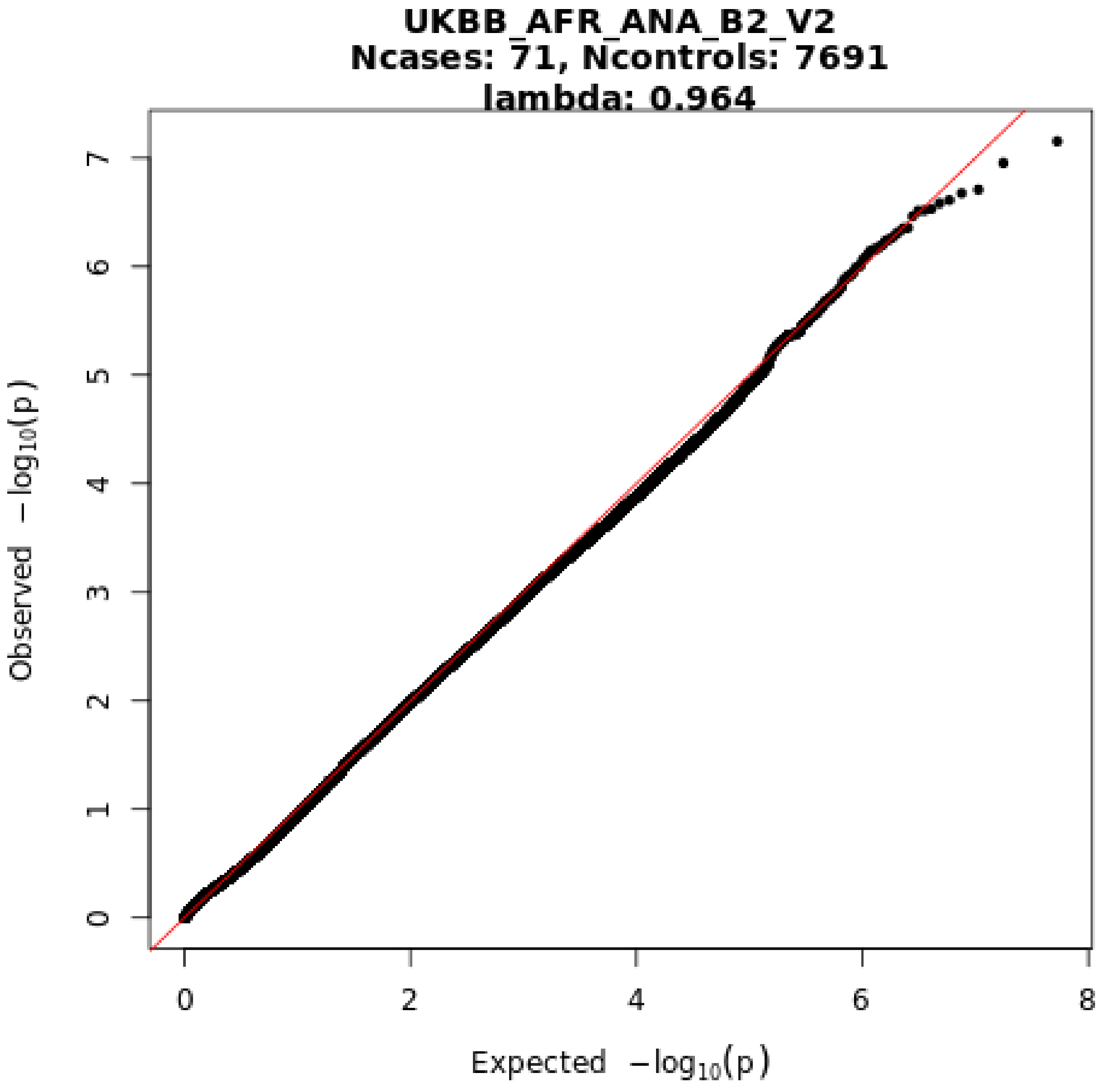
UKBB_AFR_ANA_B2_V2
Ncases: 71, Ncontrols: 7691
lambda: 0.964
Observed −log10(p)
Expected −log10(p)
0
1
2
3
4
5
6
7
8

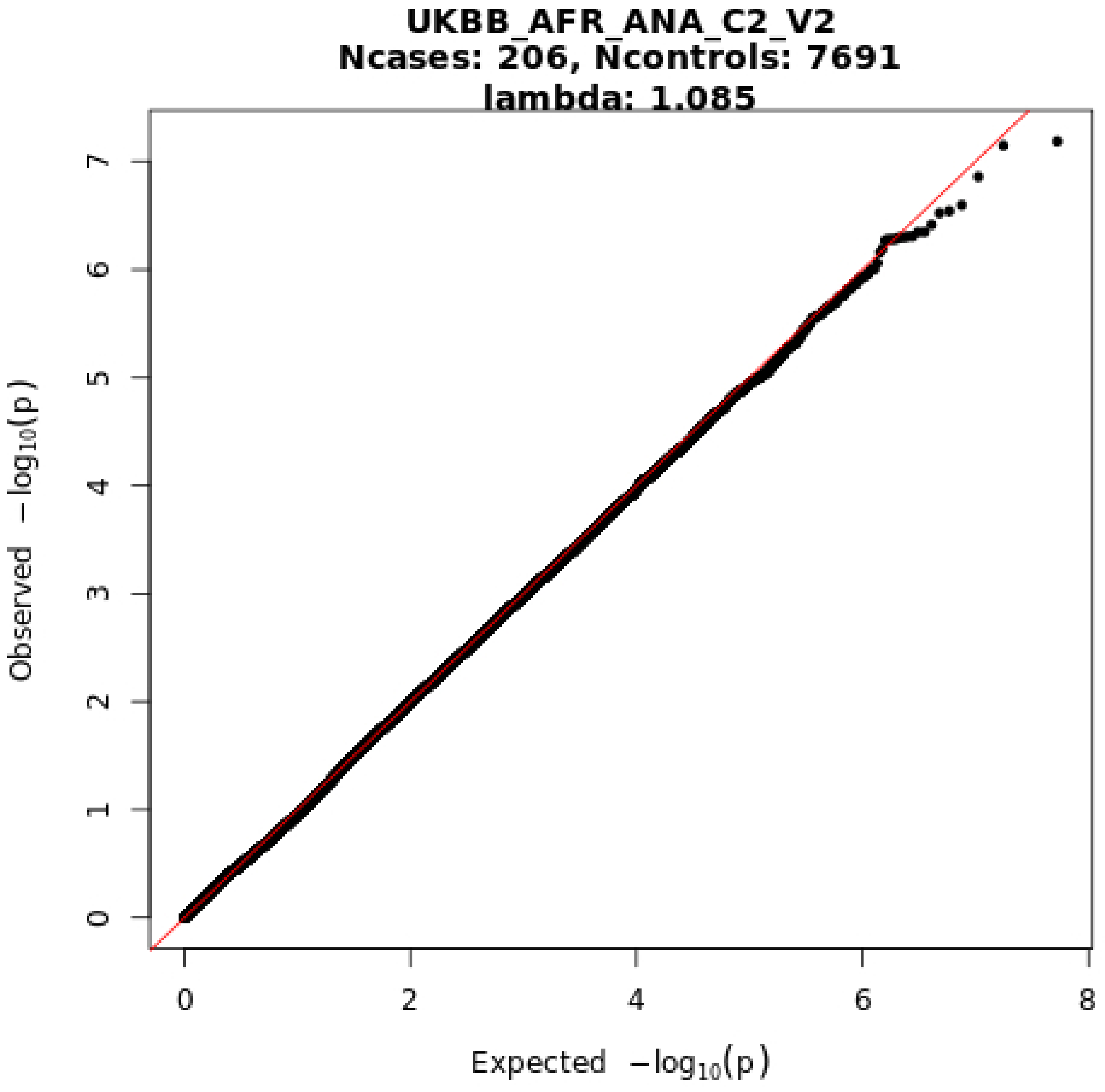

UKBB_AFR_ANA_C2_V2
Ncases: 206, Ncontrols: 7691
lambda: 1.085
Observed −log10(p)
Expected −log10(p)
0
1
2
3
4
5
6
7
0
2
4
6
8

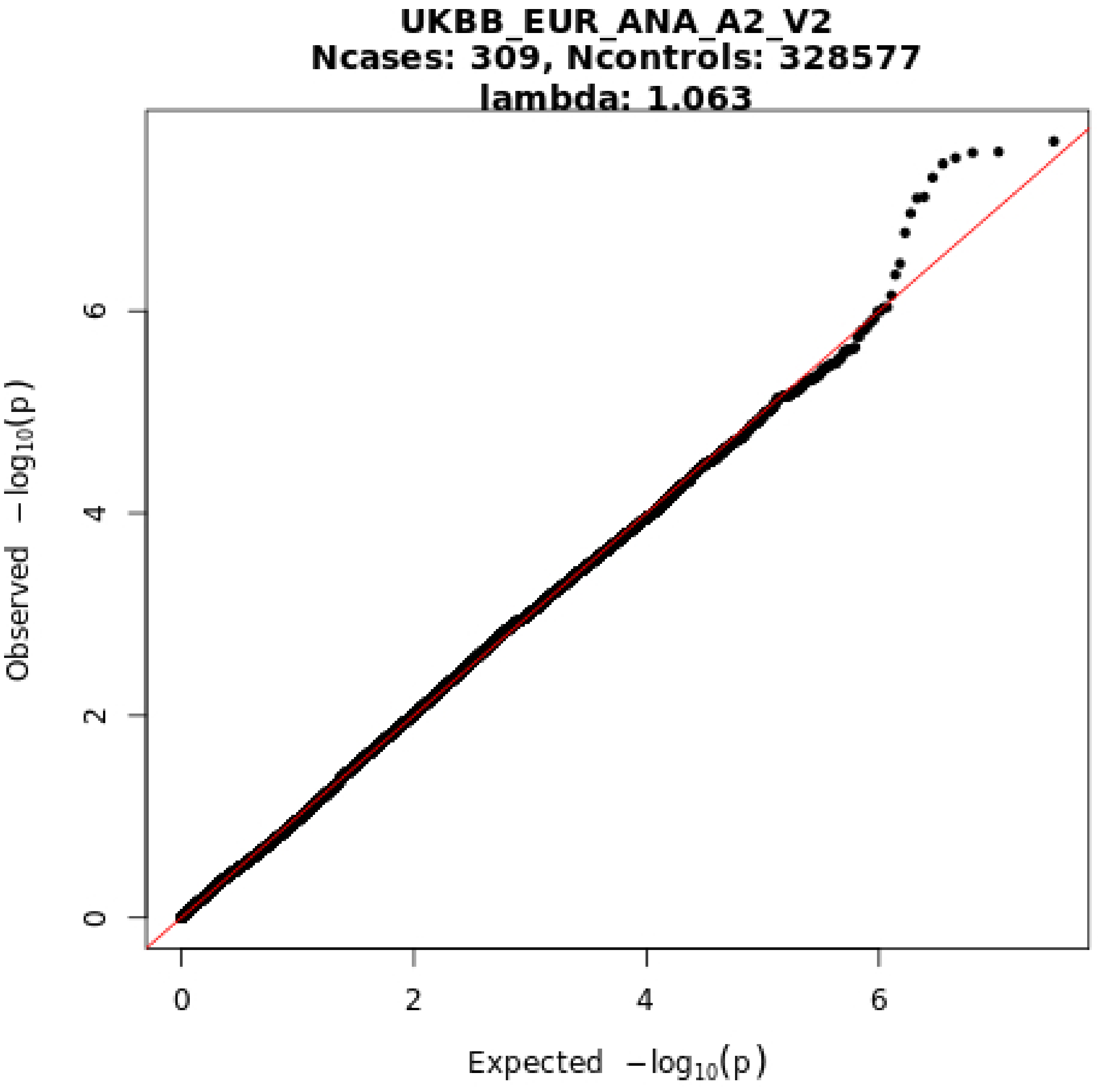
UKBB_EUR_ANA_A2_V2
Ncases: 309, Ncontrols: 328577
lambda: 1.063
Observed −log10(p)
Expected −log10(p)
0
2
4
6
0
2
4
6

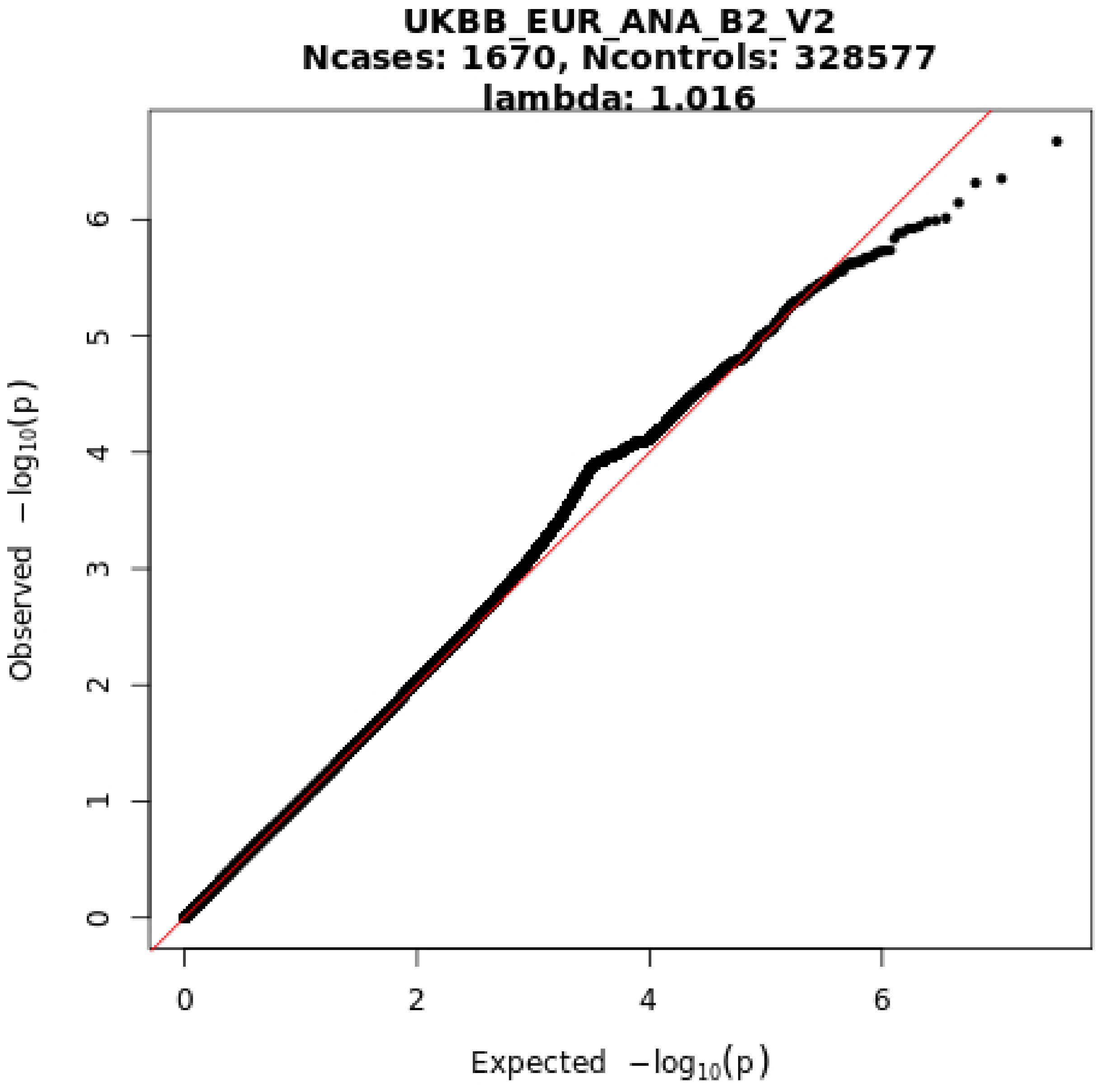
UKBB_EUR_ANA_B2_V2
Ncases: 1670, Ncontrols: 328577
lambda: 1.016
Observed −log10(p)
Expected −log10(p)
0
1
2
3
4
5
6
0
2
4
6

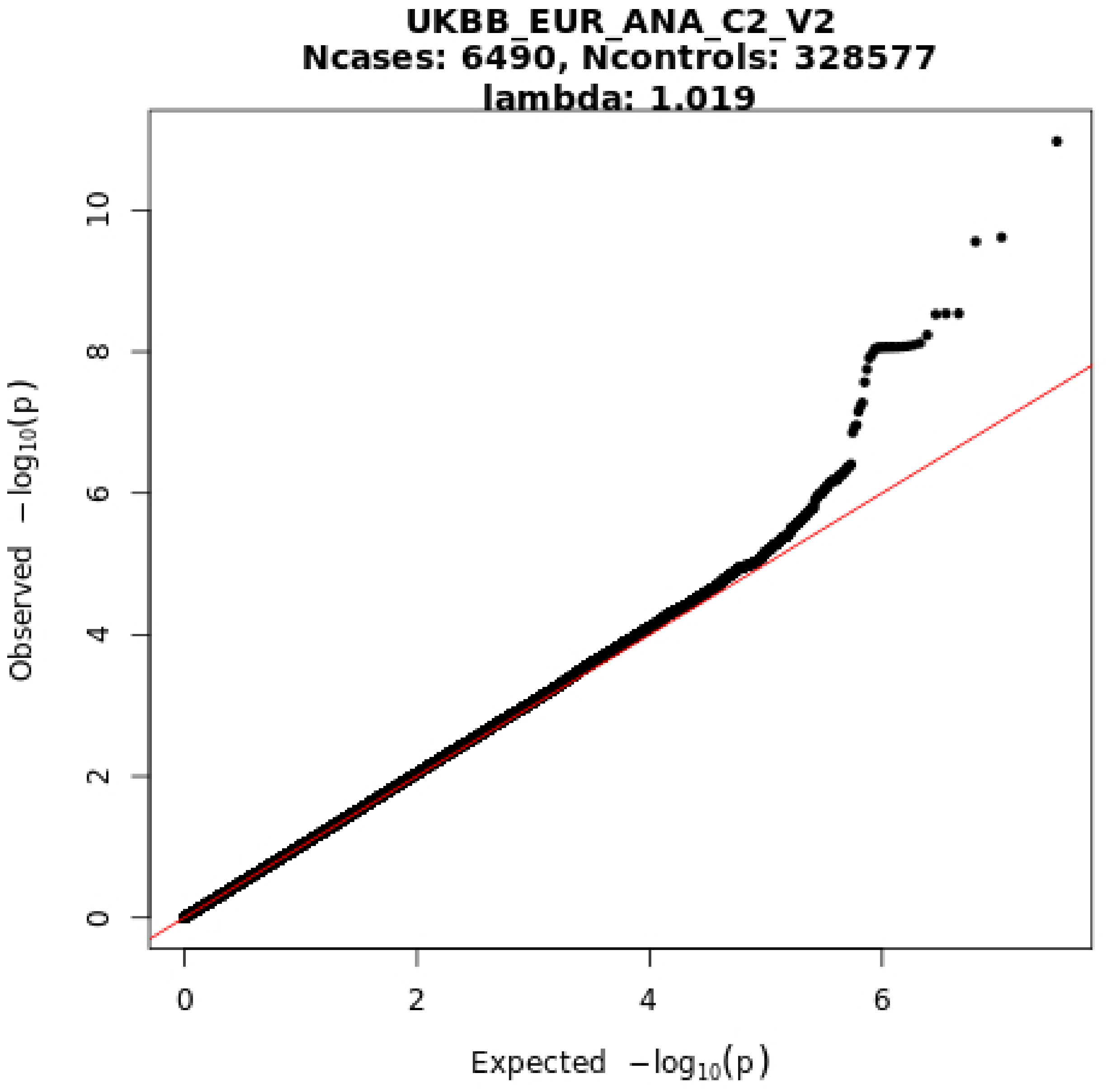
UKBB_EUR_ANA_C2_V2
Ncases: 6490, Ncontrols: 328577
lambda: 1.019
Observed −log10(p)
Expected −log10(p)
0
2
4
6
8
10
0
2
4
6

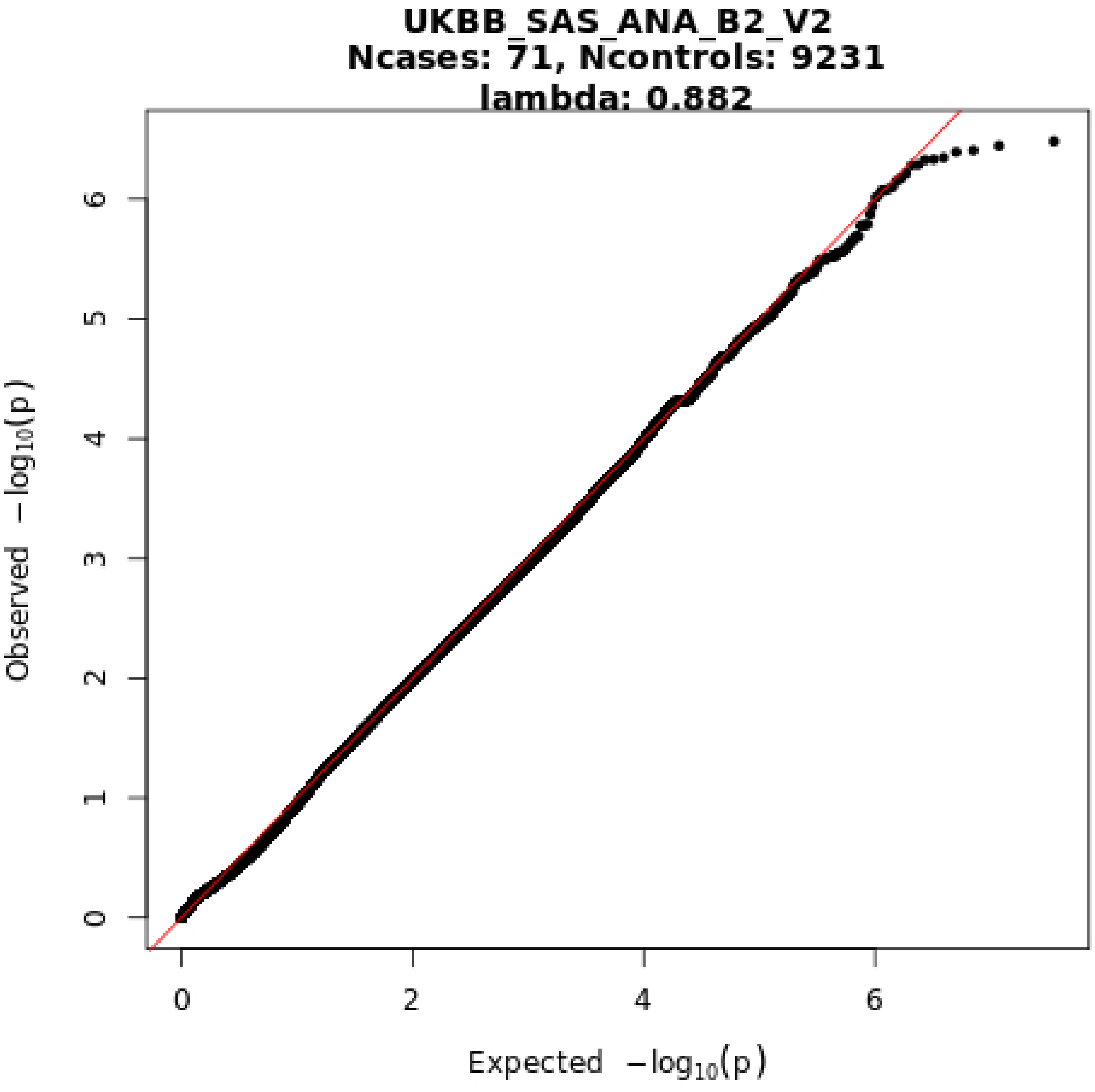
UKBB_SAS_ANA_B2_V2
Ncases: 71, Ncontrols: 9231
lambda: 0.882
Observed −log10(p)
Expected −log10(p)
0
1
2
3
4
5
6
0
2
4
6

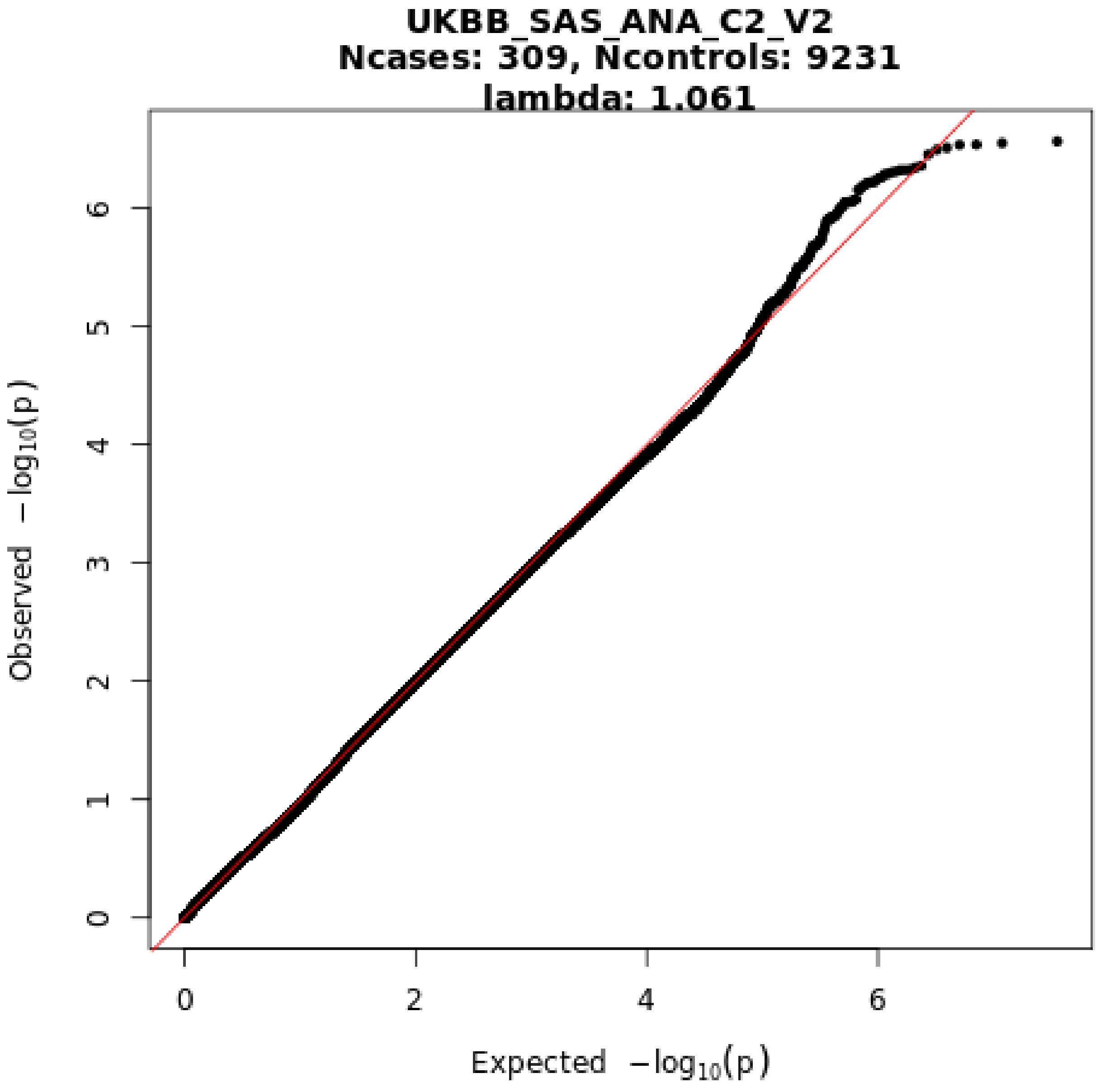
UKBB_SAS_ANA_C2_V2
Ncases: 309, Ncontrols: 9231
lambda: 1.061
Observed −log10(p)
Expected −log10(p)
0
1
2
3
4
5
6
0
2
4
6

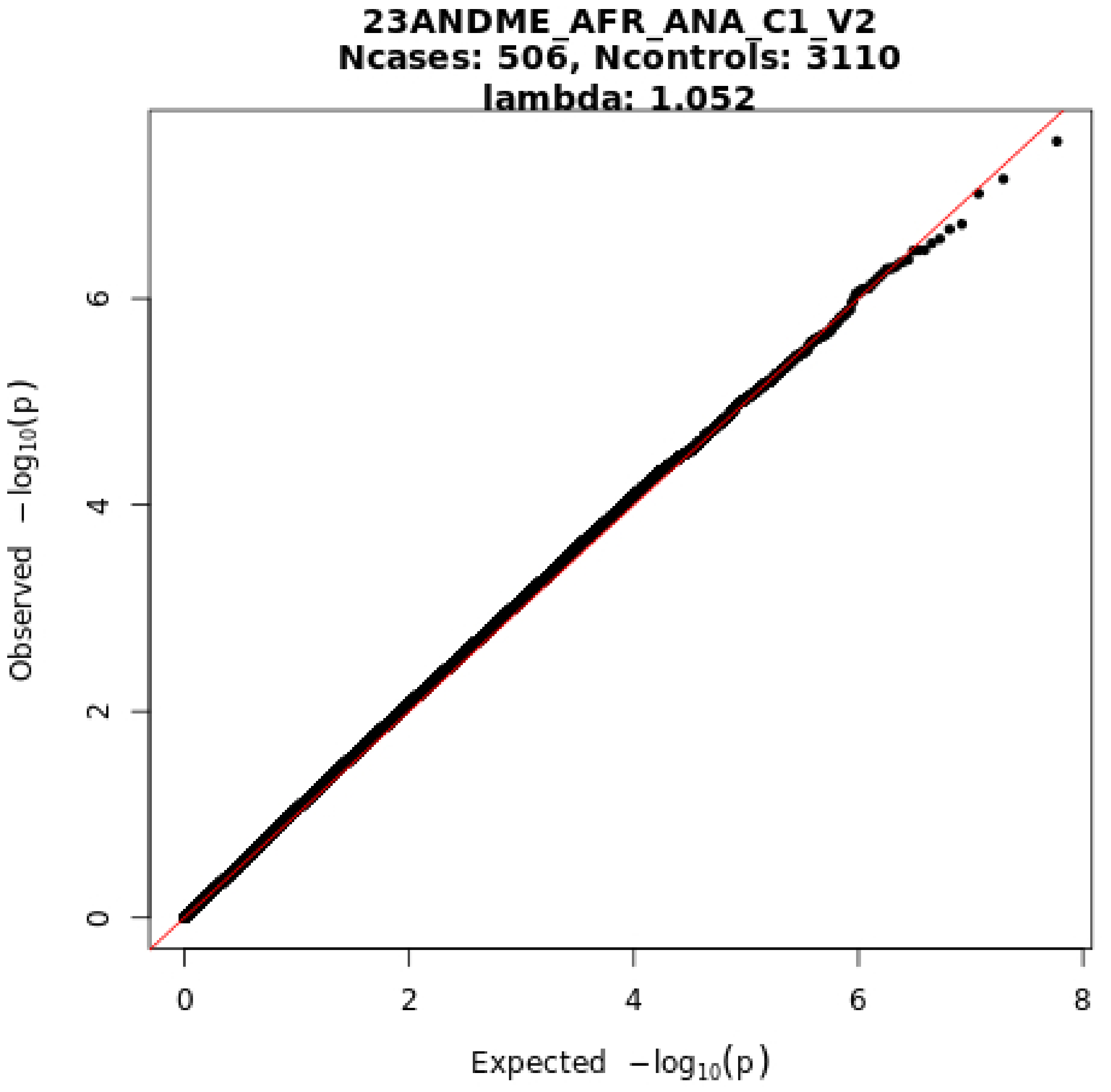

23ANDME_AFR_ANA_C1_V2
Ncases: 506, Ncontrols: 3110
lambda: 1.052
Observed −log10(p)
Expected −log10(p)
0
2
4
6
8

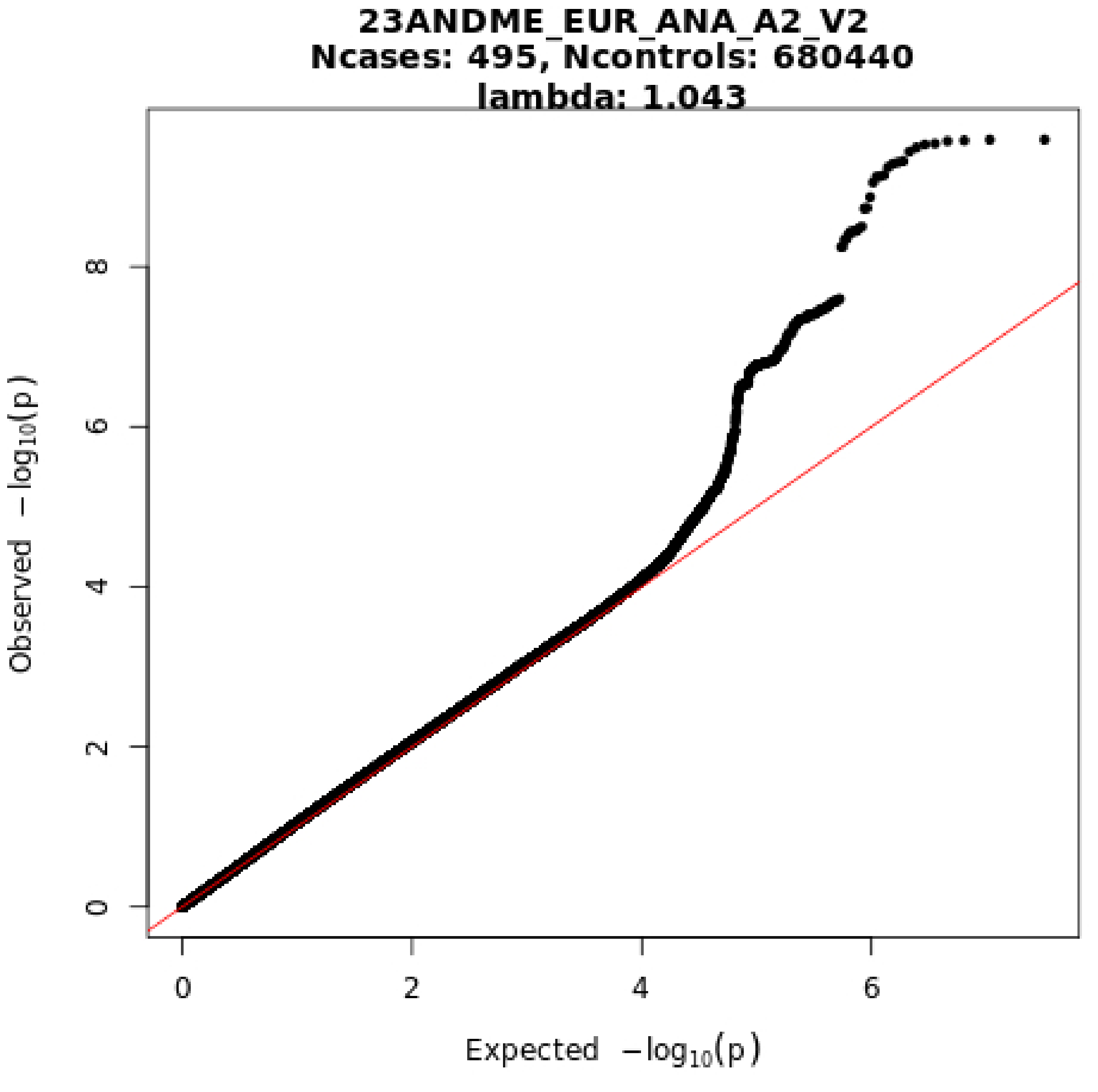

23ANDME_EUR_ANA_A2_V2
Ncases: 495, Ncontrols: 680440
lambda: 1.043
Observed −log10(p)
Expected −log10(p)
0
2
4
6
8
0
2
4
6

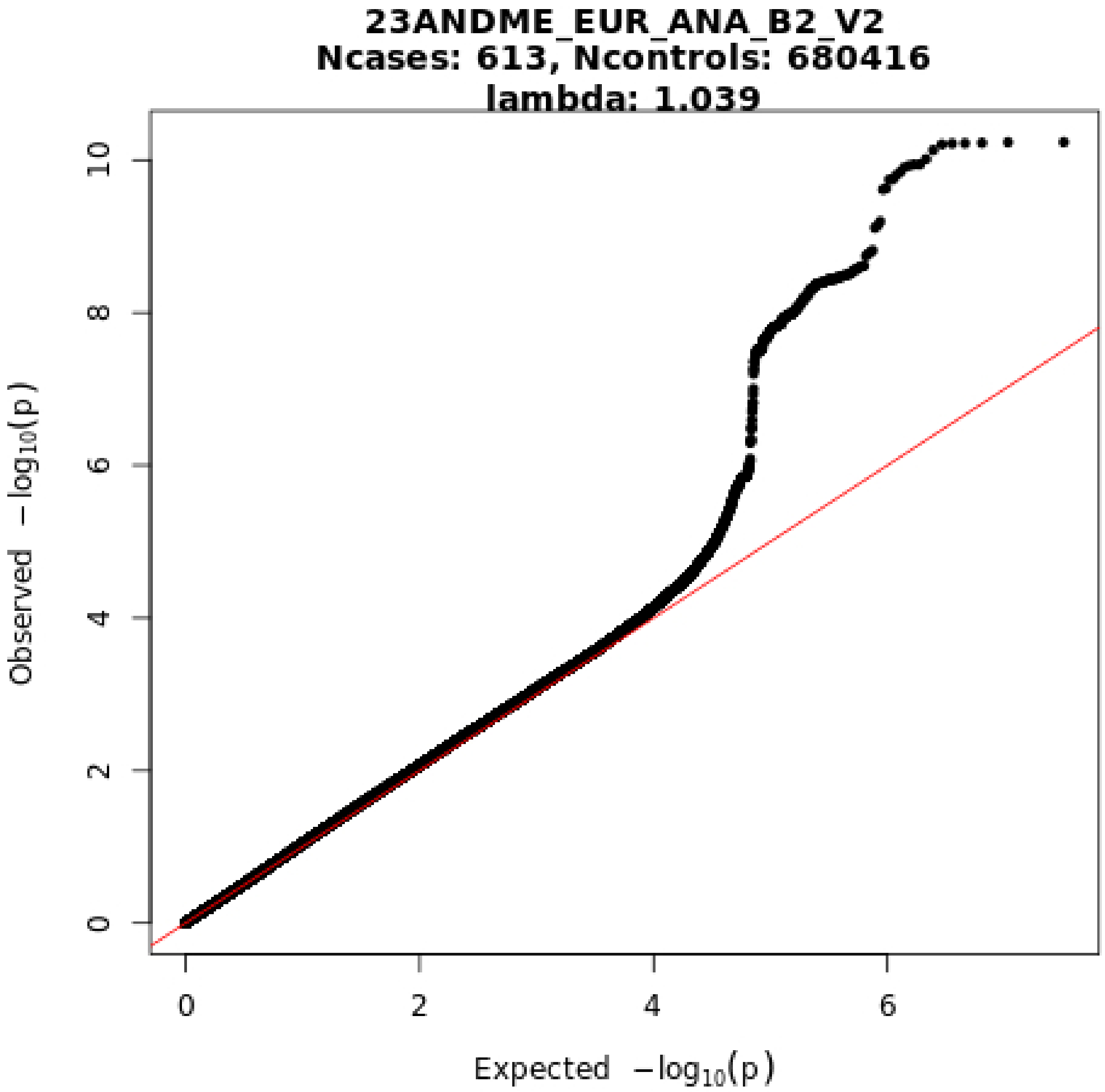

23ANDME_EUR_ANA_B2_V2
Ncases: 613, Ncontrols: 680416
lambda: 1.039
Observed −log10(p)
Expected −log10(p)
0
2
4
6
8
10
0
2
4
6

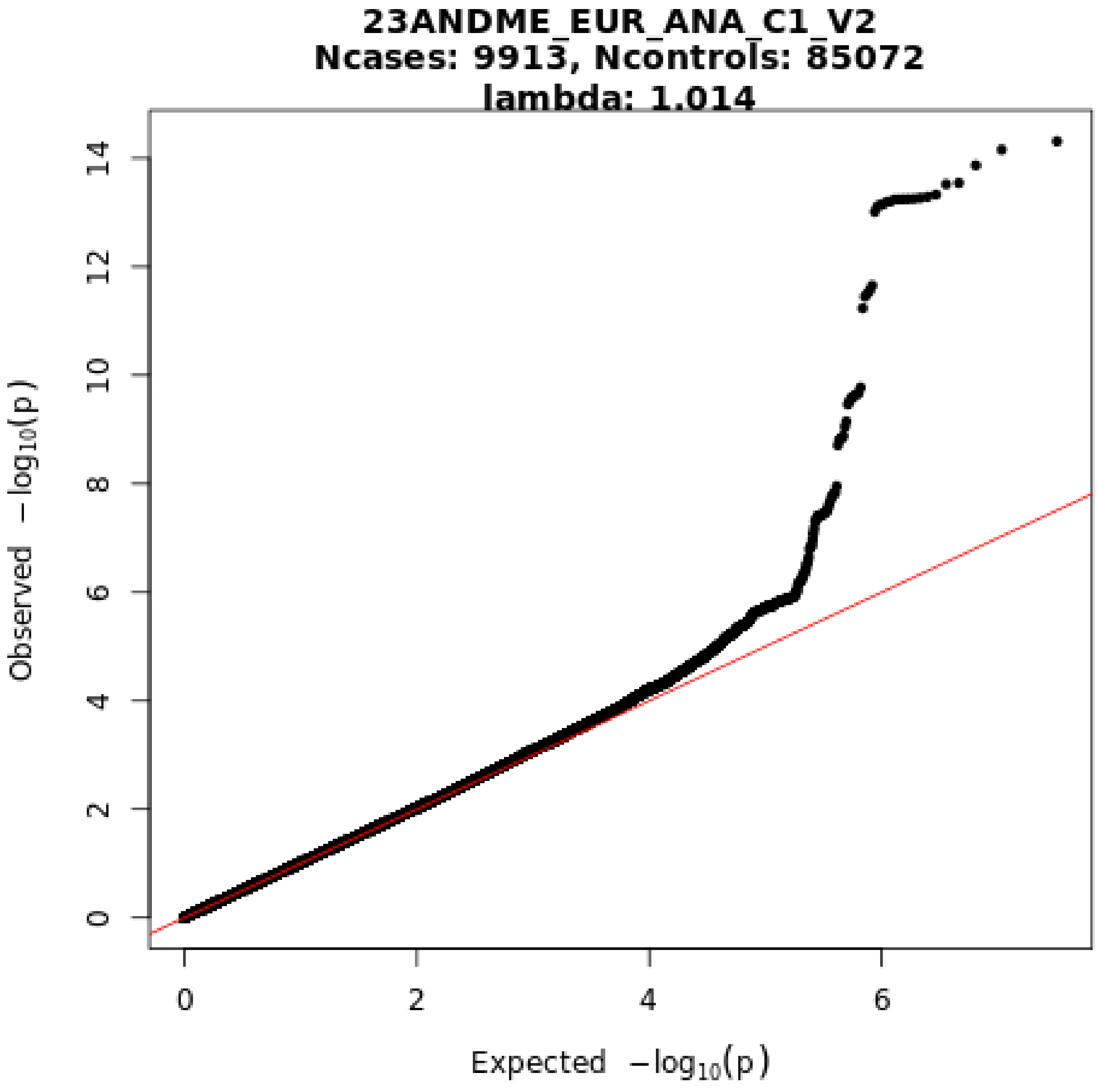
23ANDME_EUR_ANA_C1_V2
Ncases: 9913, Ncontrols: 85072
lambda: 1.014
Observed −log10(p)
Expected −log10(p)
0
2
4
6
8
10
12
14
0
2
4
6

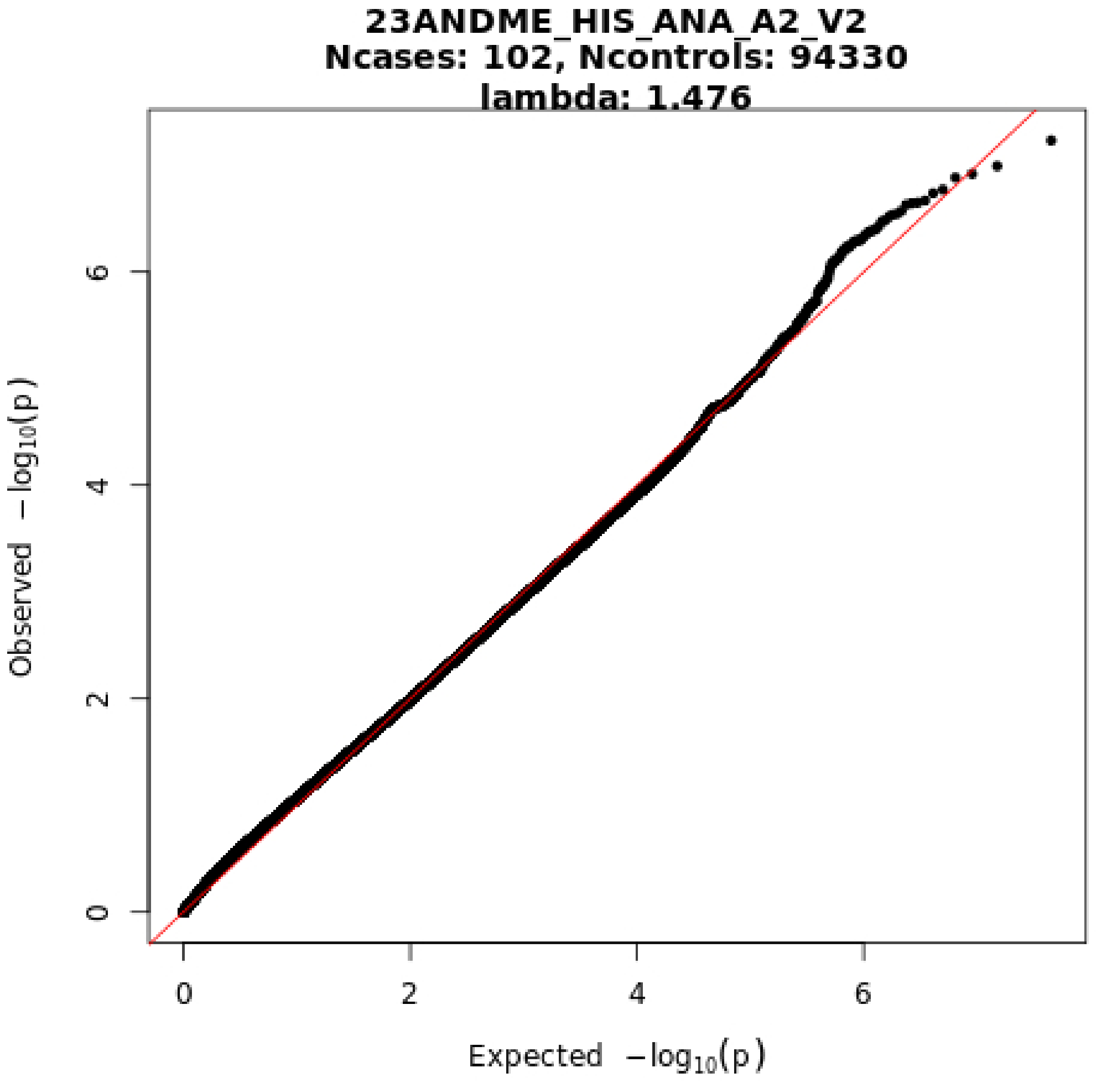
23ANDME_HIS_ANA_A2_V2
Ncases: 102, Ncontrols: 94330
lambda: 1.476
Observed −log10(p)
Expected −log10(p)
0
2
4
6
0
2
4
6

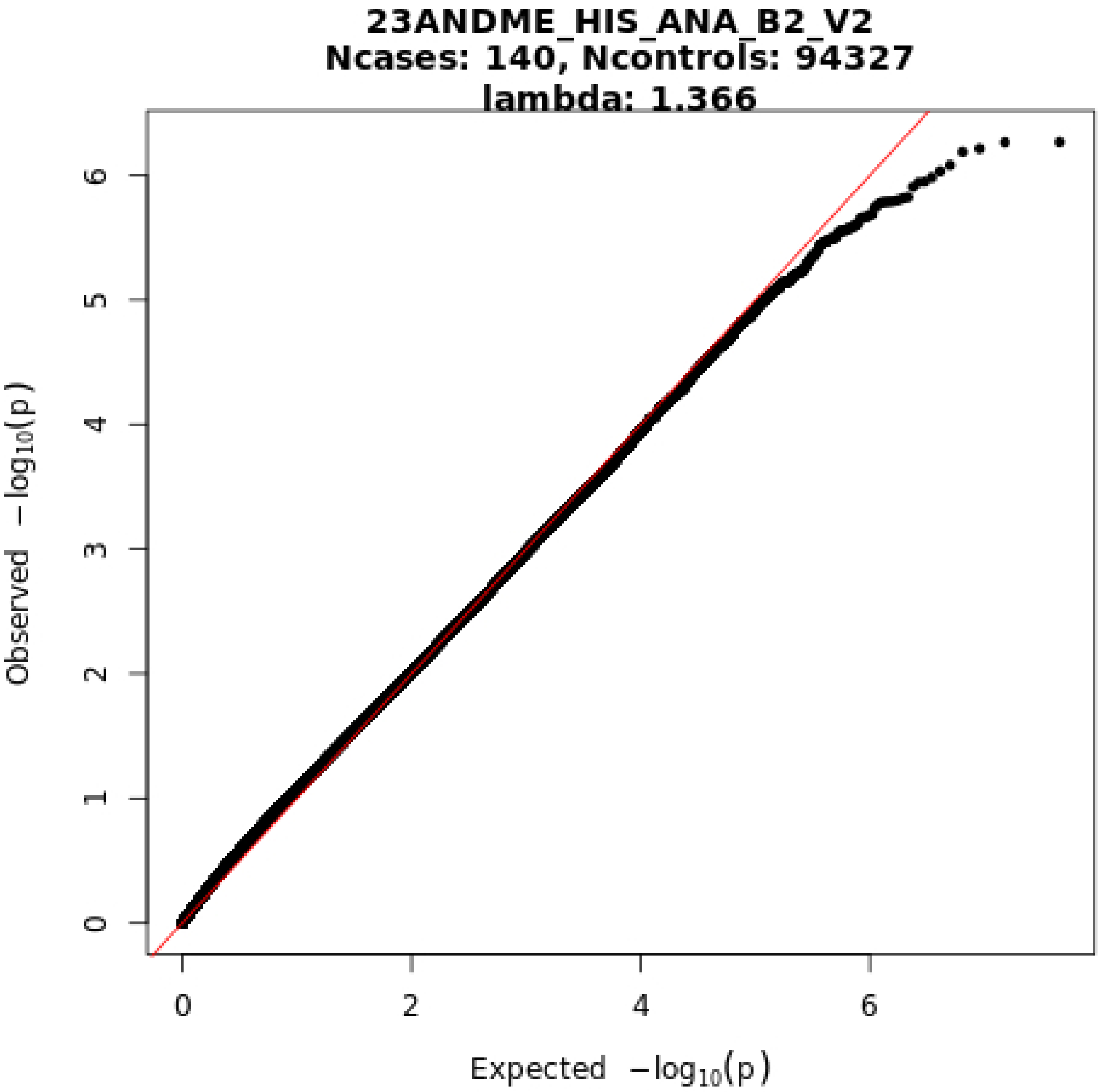

23ANDME_HIS_ANA_B2_V2
Ncases: 140, Ncontrols: 94327
lambda: 1.366
Observed −log10(p)
Expected −log10(p)
0
1
2
3
4
5
6
0
2
4
6

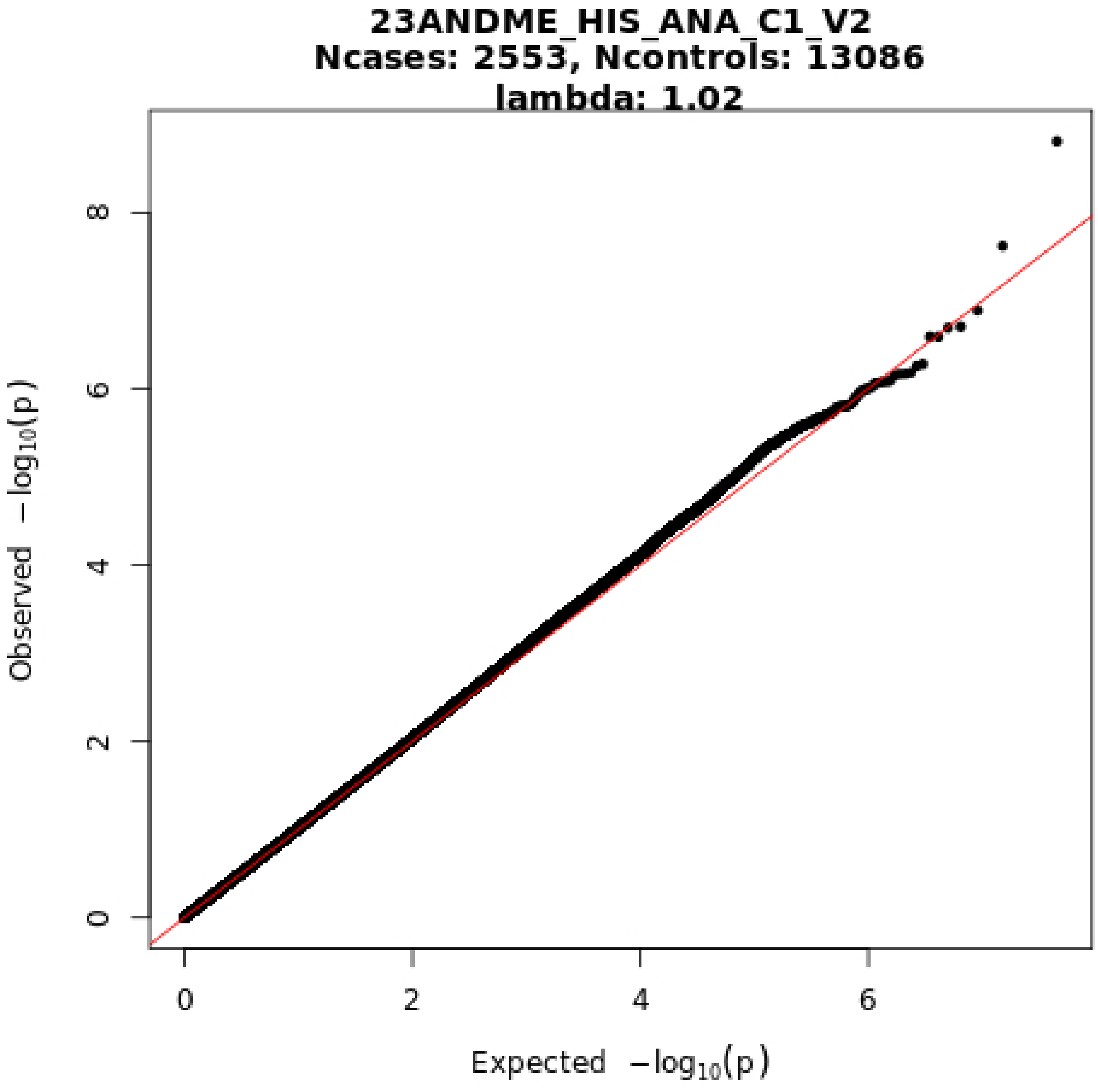
23ANDME_HIS_ANA_C1_V2
Ncases: 2553, Ncontrols: 13086
lambda: 1.02
Observed −log10(p)
Expected −log10(p)
0
2
4
6
8
0
2
4
6

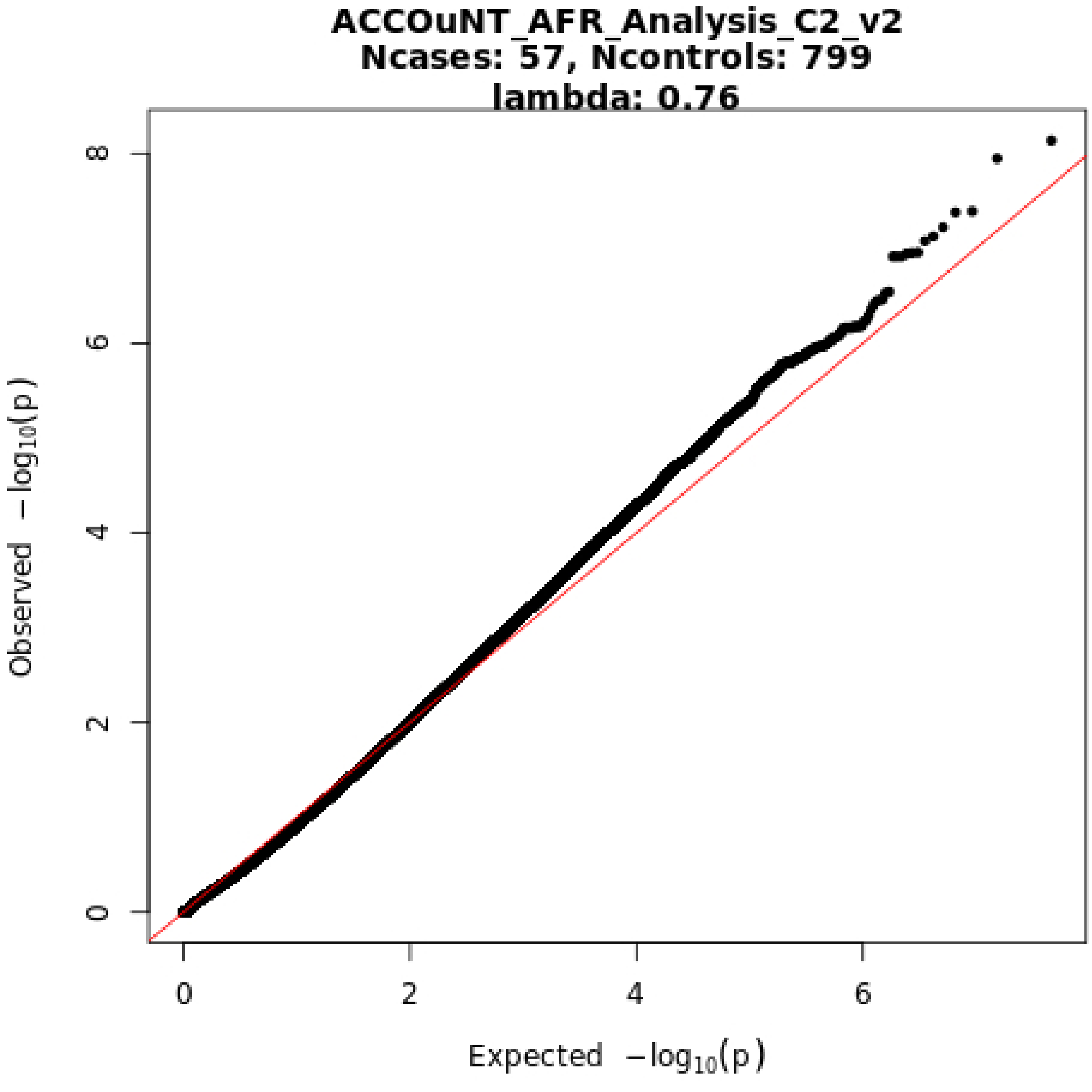
ACCOuNT_AFR_Analysis_C2_v2
Ncases: 57, Ncontrols: 799
lambda: 0.76
Observed −log10(p)
Expected −log10(p)
0
2
4
6
8
0
2
4
6

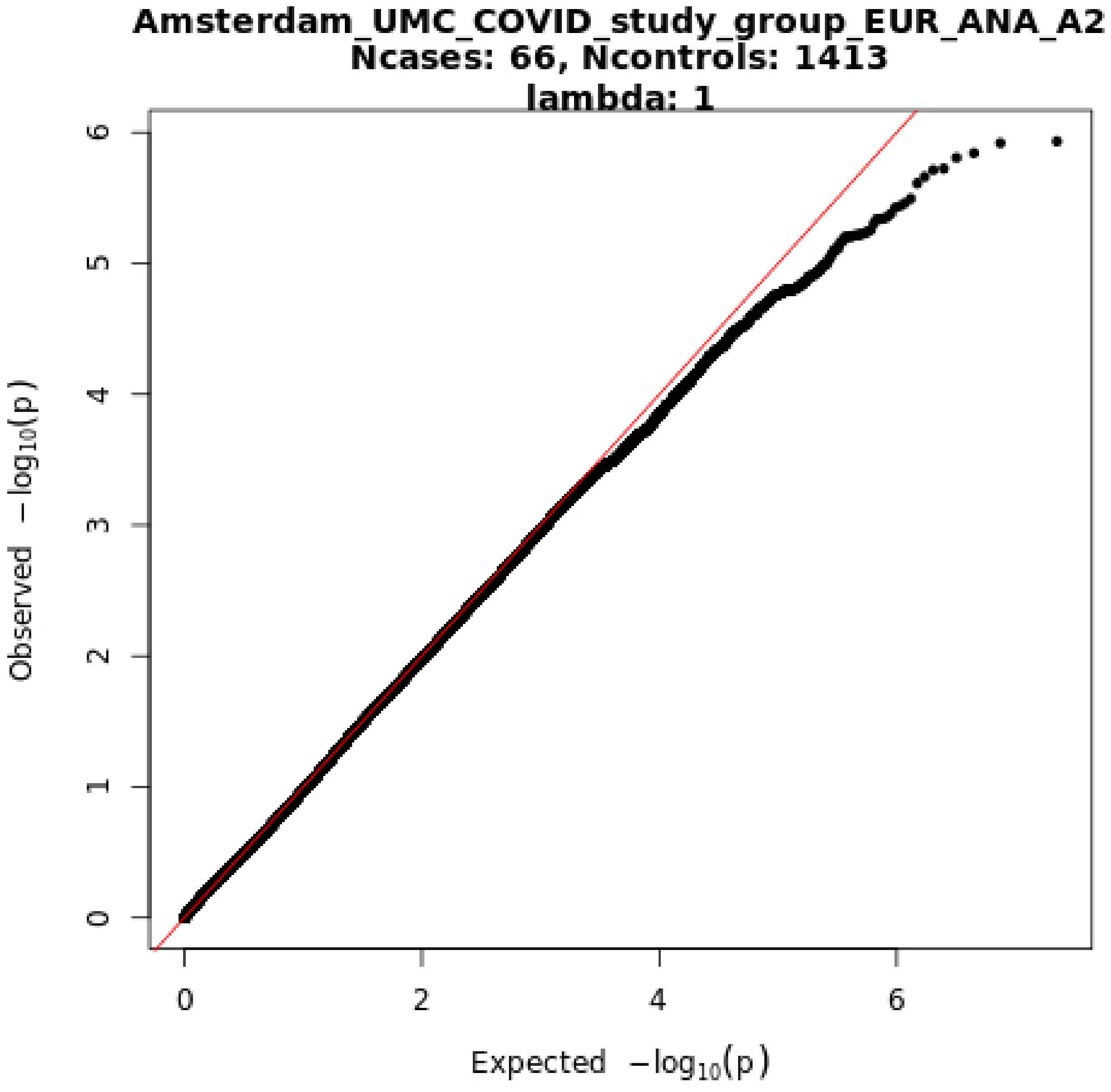
Amsterdam_UMC_COVID_study_group_EUR_ANA_A2
Ncases: 66, Ncontrols: 1413
lambda: 1
Observed $-\log_{10}(p)$
Expected $-\log_{10}(p)$
0
1
2
3
4
5
6
0
2
4
6

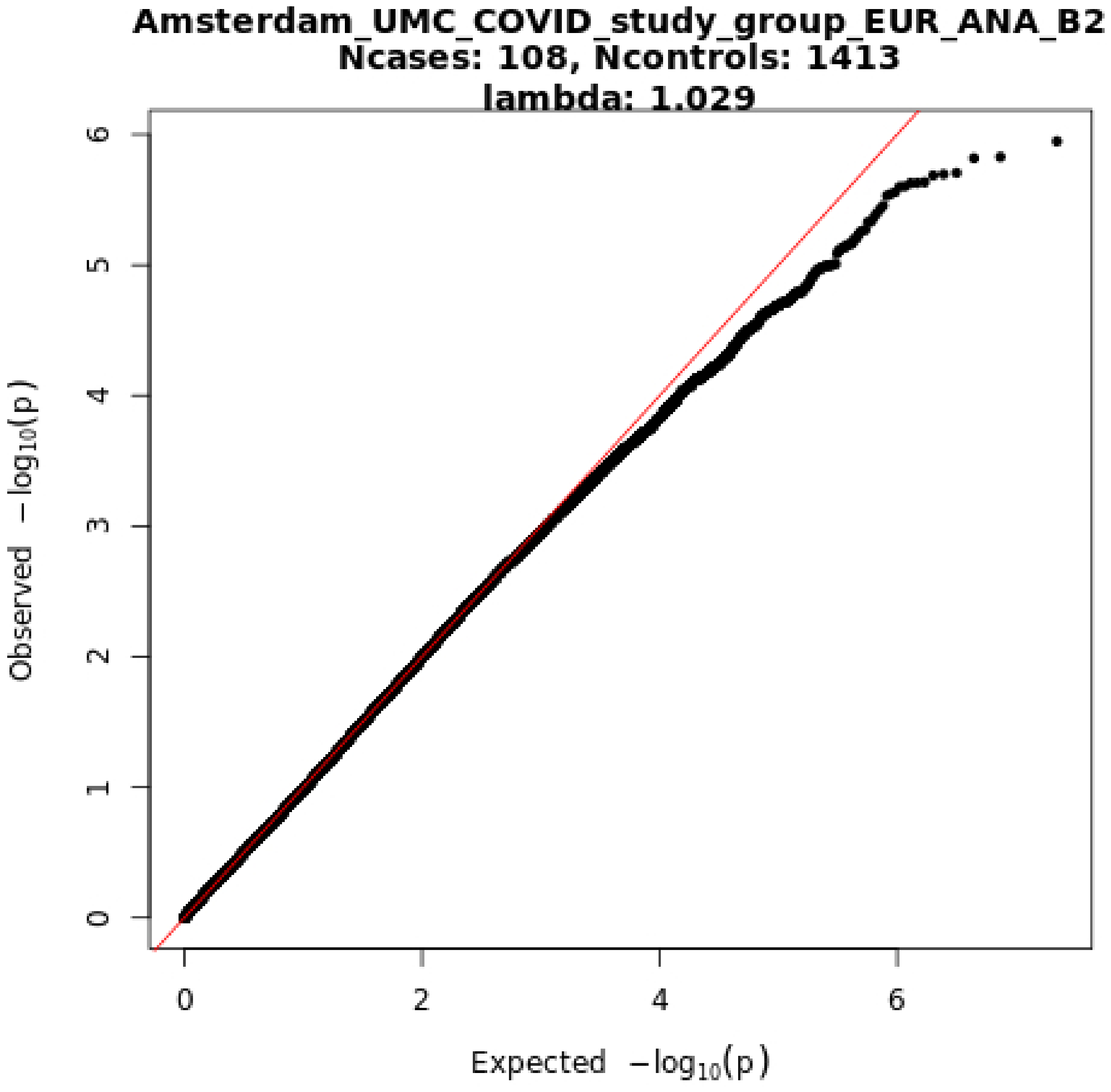
Amsterdam_UMC_COVID_study_group_EUR_ANA_B2
Ncases: 108, Ncontrols: 1413
lambda: 1.029
Observed −log10(p)
Expected −log10(p)
0
1
2
3
4
5
6
0
2
4
6

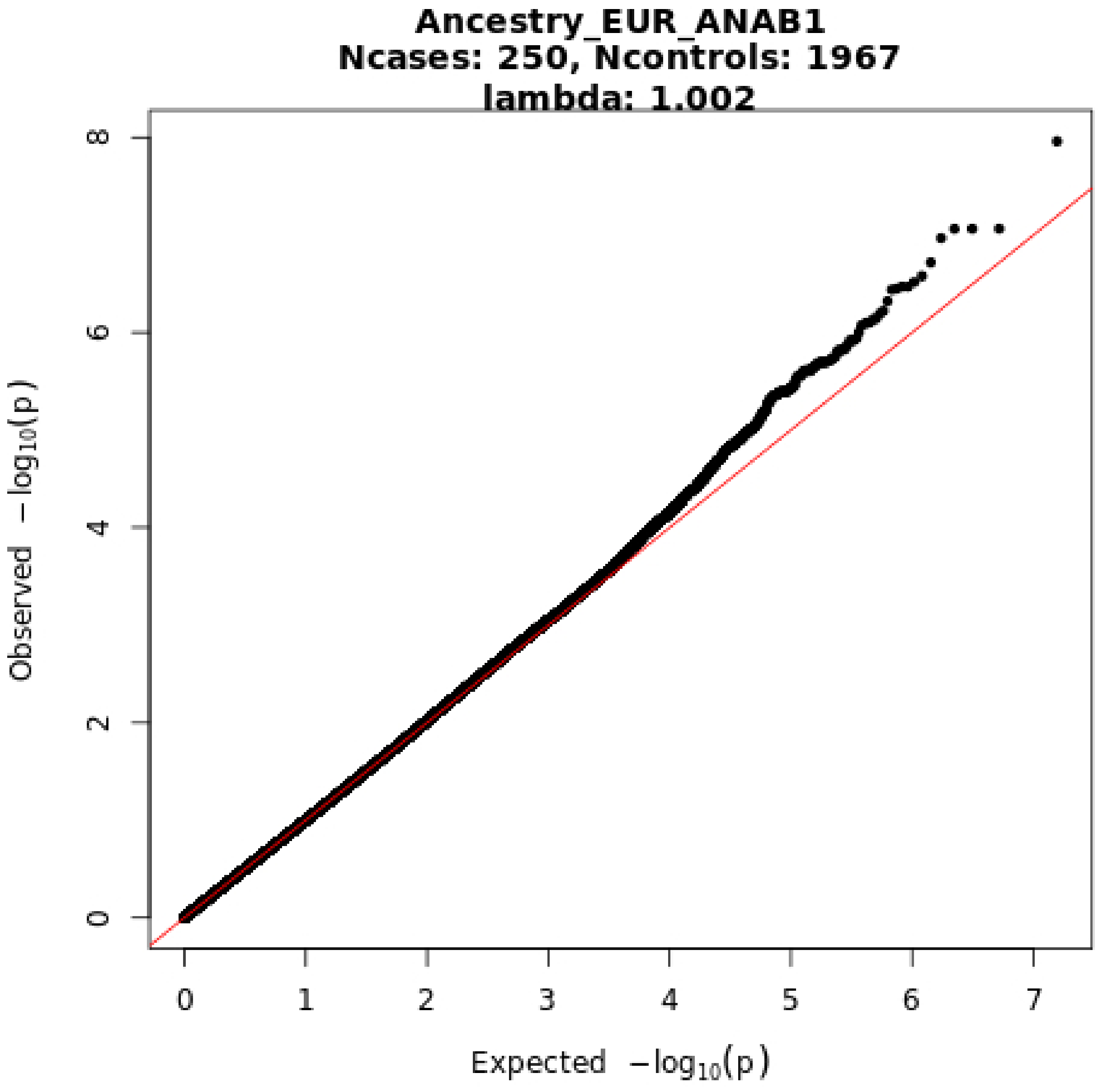
Ancestry_EUR_ANAB1
Ncases: 250, Ncontrols: 1967
lambda: 1.002
Observed −log10(p)
Expected −log10(p)
0
2
4
6
8
0
1
2
3
4
5
6
7

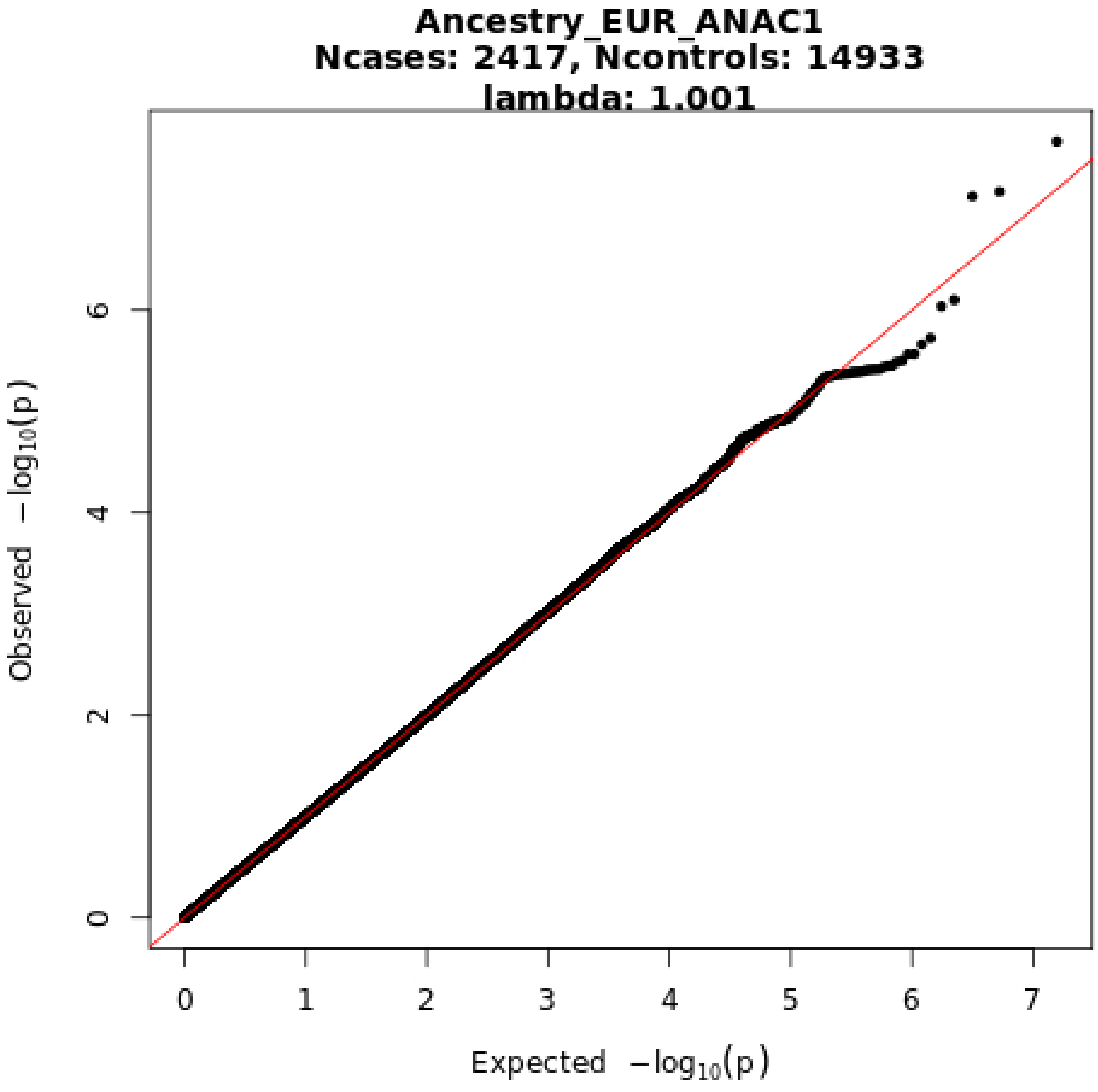
Ancestry_EUR_ANAC1
Ncases: 2417, Ncontrols: 14933
lambda: 1.001
0
2
4
6
0
1
2
3
4
5
6
7
Observed −log10(p)
Expected −log10(p)

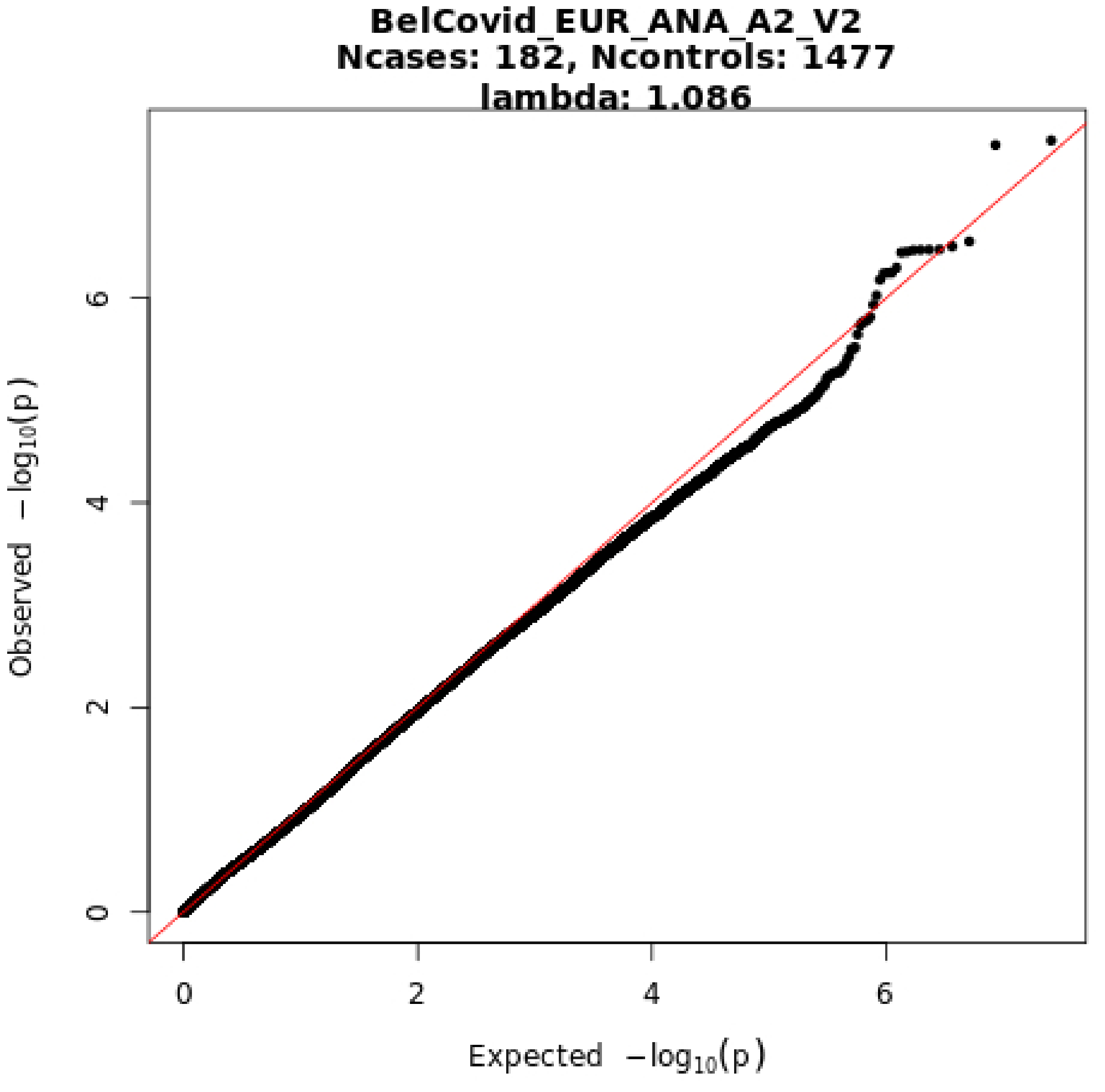
BelCovid_EUR_ANA_A2_V2
Ncases: 182, Ncontrols: 1477
lambda: 1.086
Observed −log10(p)
Expected −log10(p)
0
2
4
6
0
2
4
6

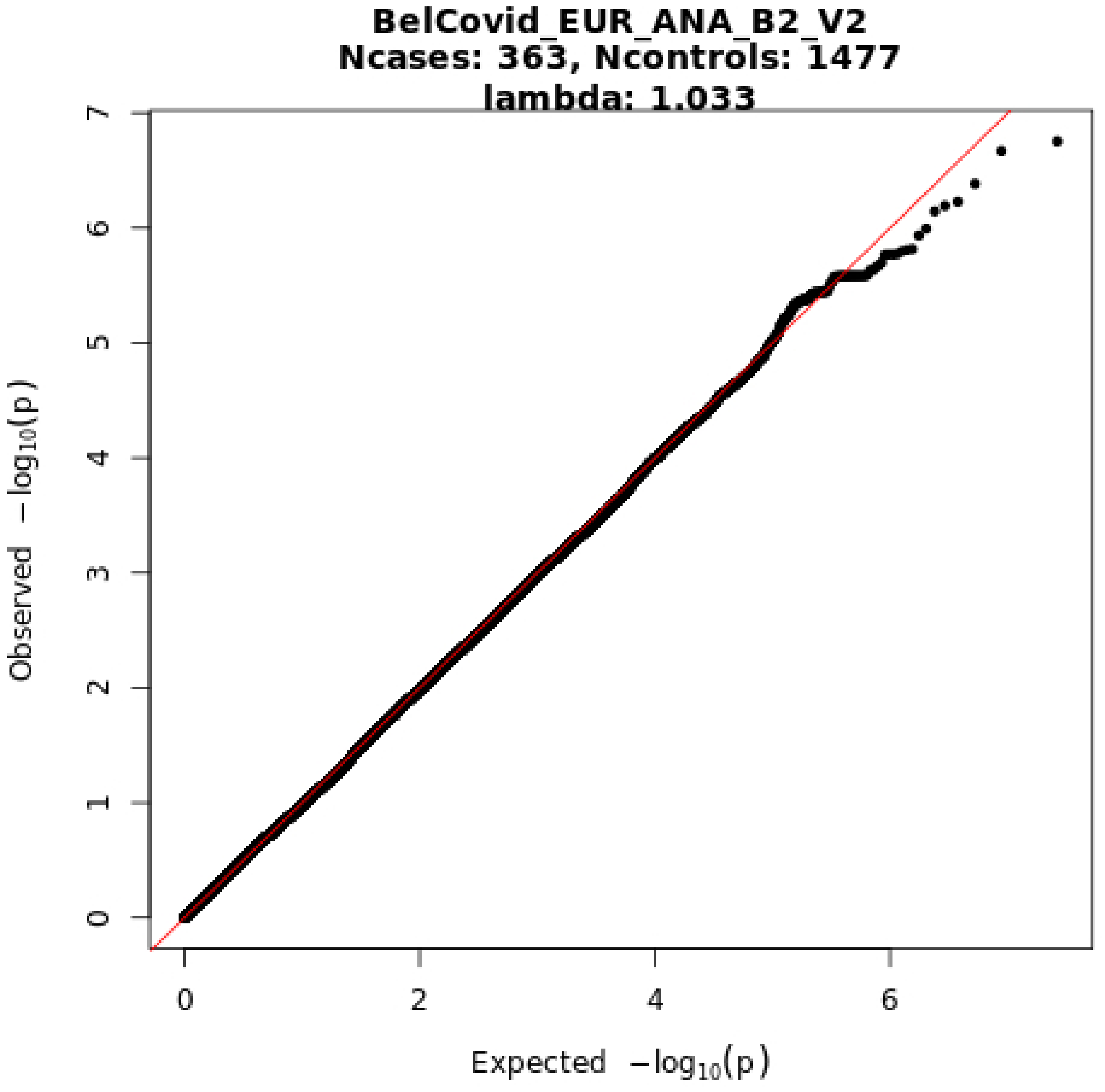
BelCovid_EUR_ANA_B2_V2
Ncases: 363, Ncontrols: 1477
lambda: 1.033
Observed −log10(p)
Expected −log10(p)
0
1
2
3
4
5
6
7
0
2
4
6

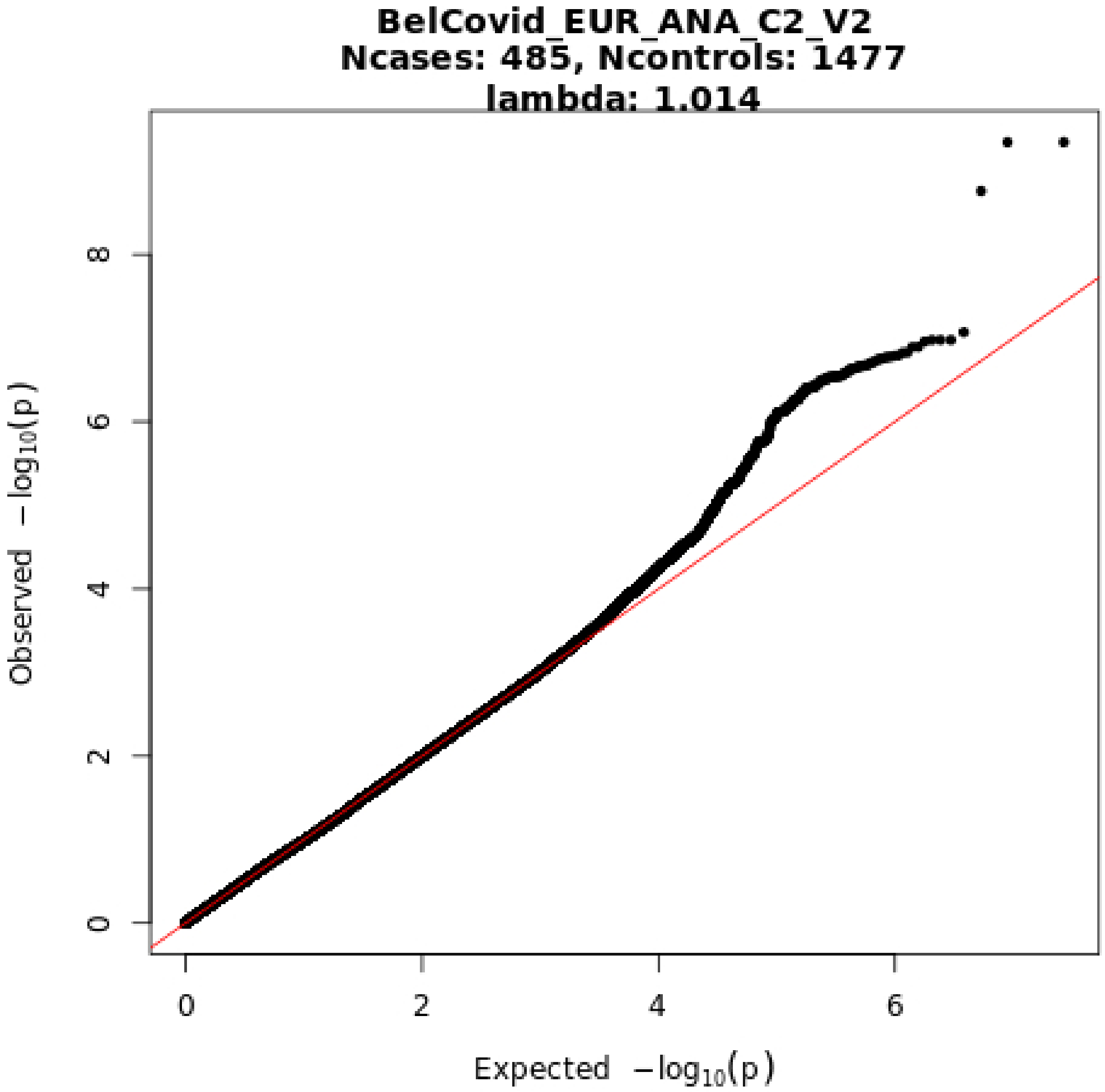

BelCovid_EUR_ANA_C2_V2
Ncases: 485, Ncontrols: 1477
lambda: 1.014
Observed −log10(p)
Expected −log10(p)
0
2
4
6
8
0
2
4
6

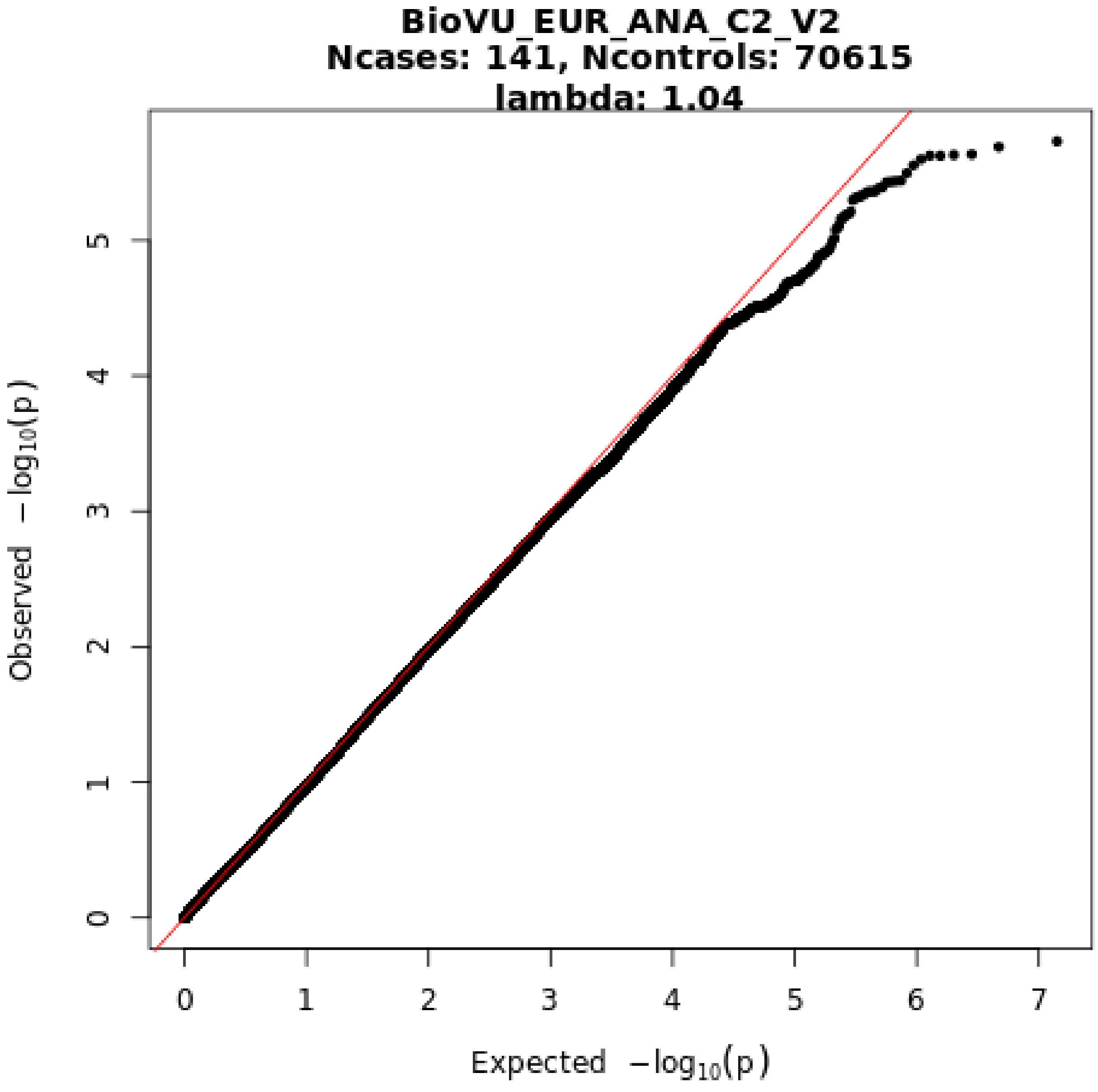
BioVU_EUR_ANA_C2_V2
Ncases: 141, Ncontrols: 70615
lambda: 1.04
Observed −log10(p)
Expected −log10(p)
0
1
2
3
4
5
6
7

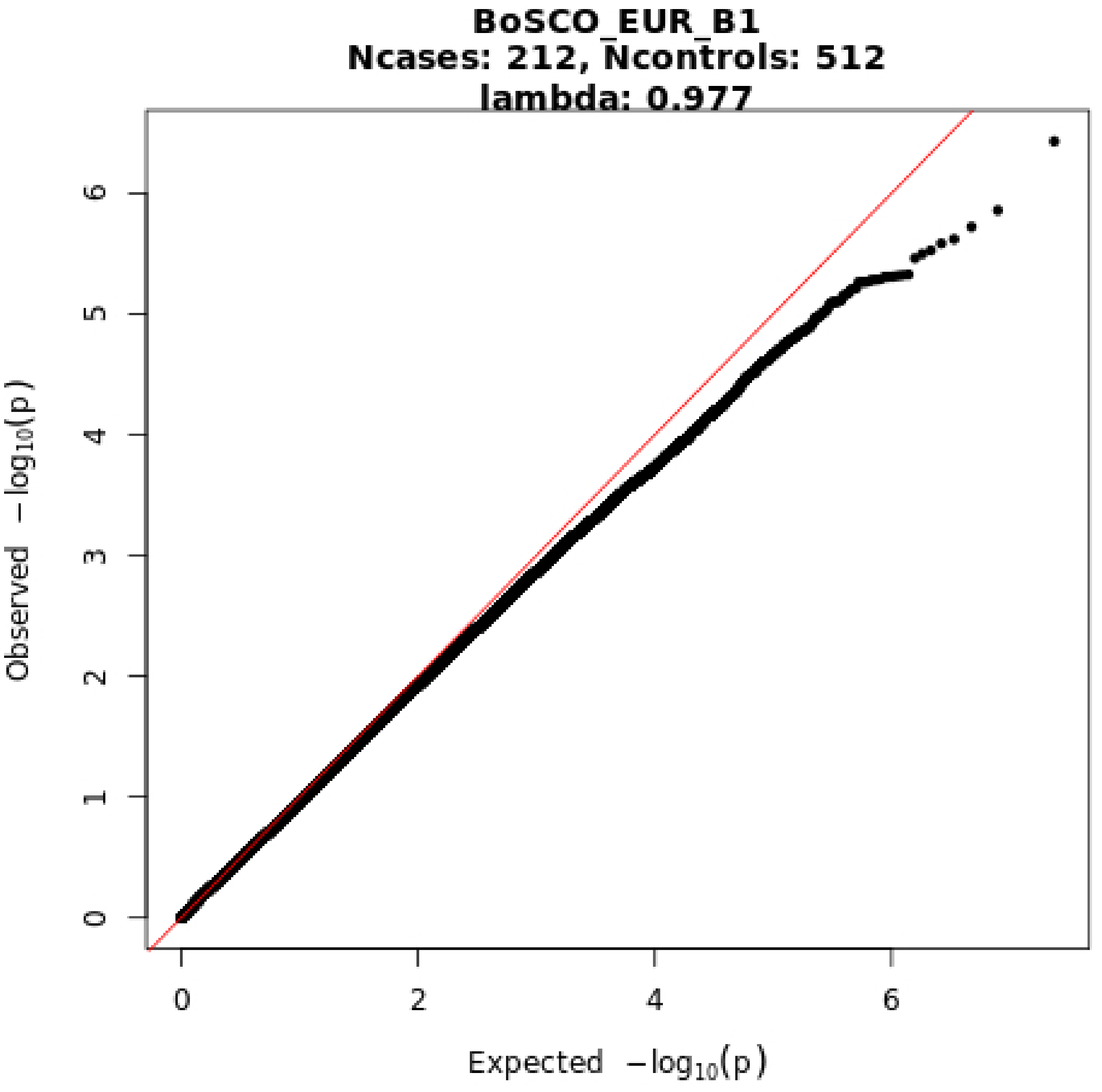

BoSCO_EUR_B1
Ncases: 212, Ncontrols: 512
lambda: 0.977
Observed −log10(p)
Expected −log10(p)
0
1
2
3
4
5
6
0
2
4
6

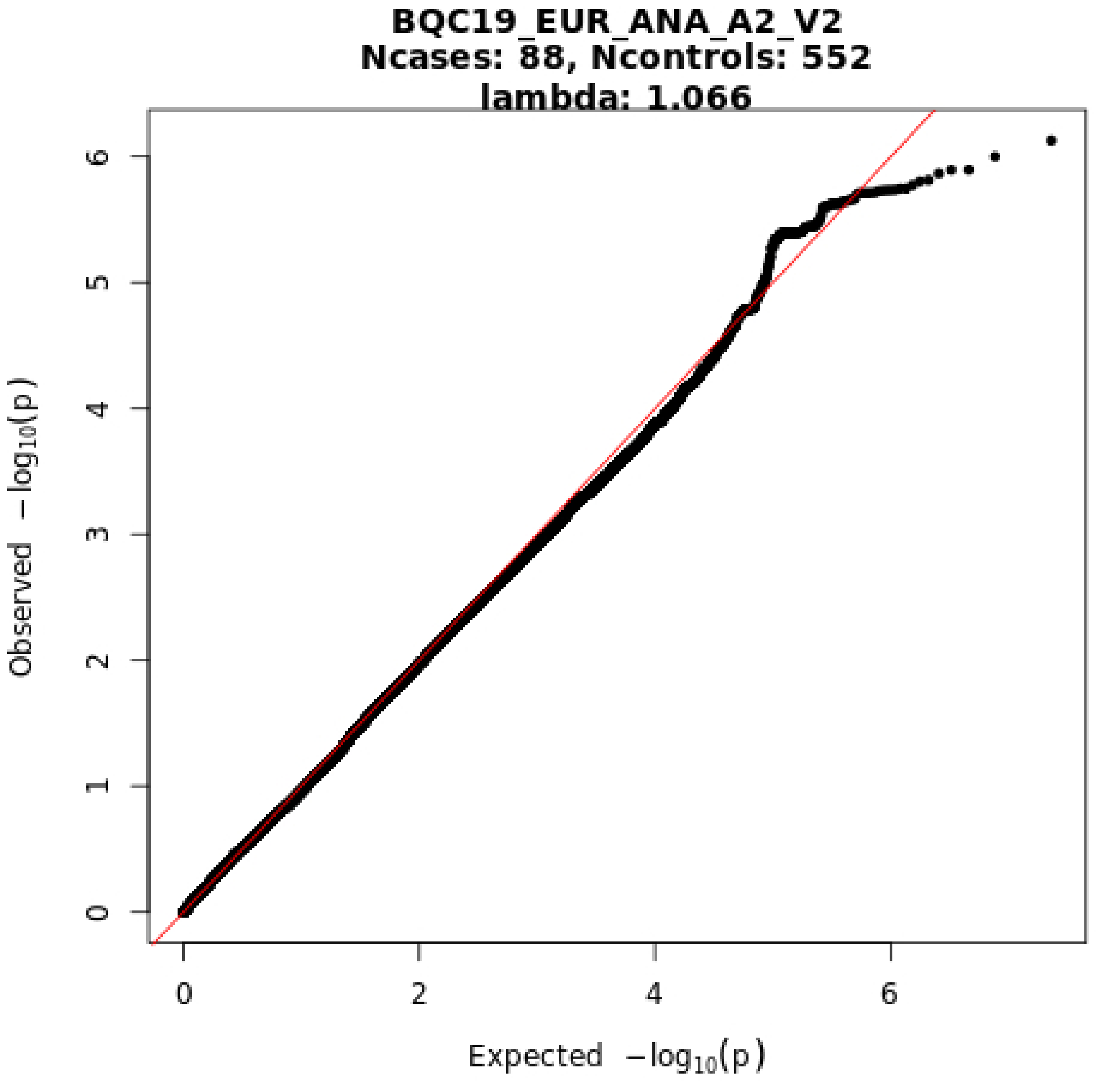
BQC19_EUR_ANA_A2_V2
Ncases: 88, Ncontrols: 552
lambda: 1.066
Observed −log10(p)
Expected −log10(p)
0
1
2
3
4
5
6
0
2
4
6

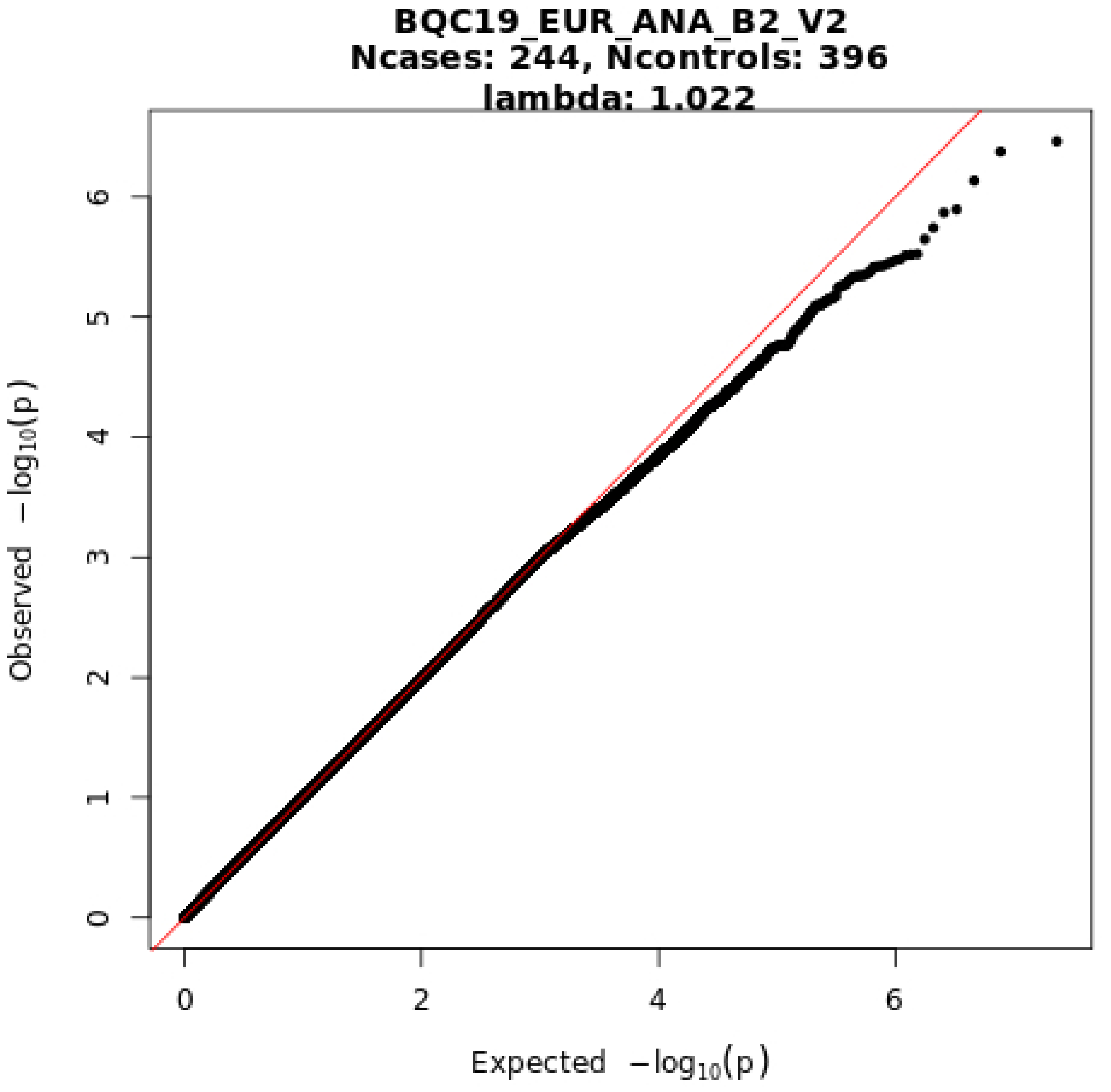

BQC19_EUR_ANA_B2_V2
Ncases: 244, Ncontrols: 396
lambda: 1.022
Observed −log10(p)
Expected −log10(p)
0
1
2
3
4
5
6
0
2
4
6

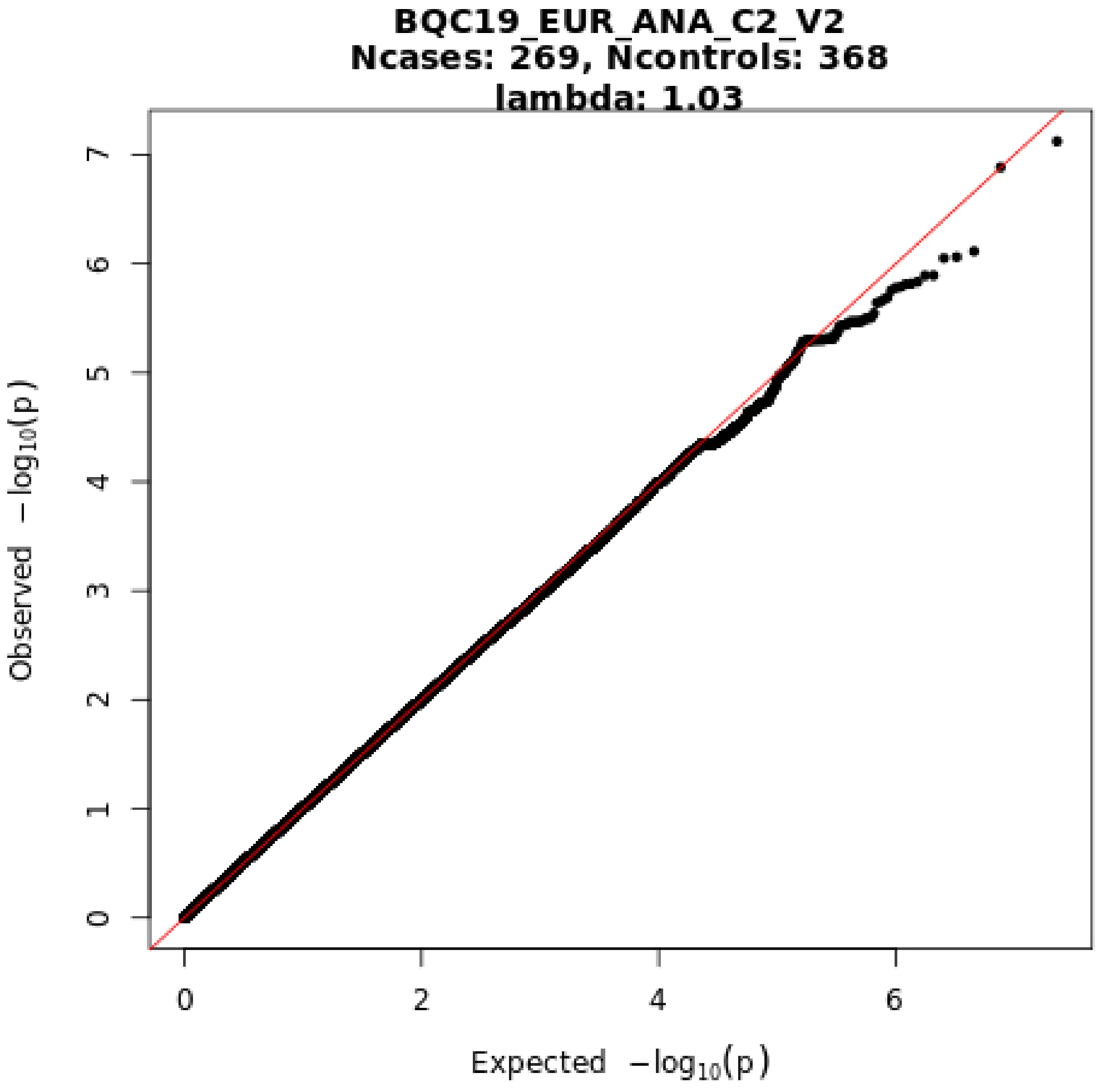
BQC19_EUR_ANA_C2_V2
Ncases: 269, Ncontrols: 368
lambda: 1.03
Observed −log10(p)
Expected −log10(p)
0
1
2
3
4
5
6
7
0
2
4
6

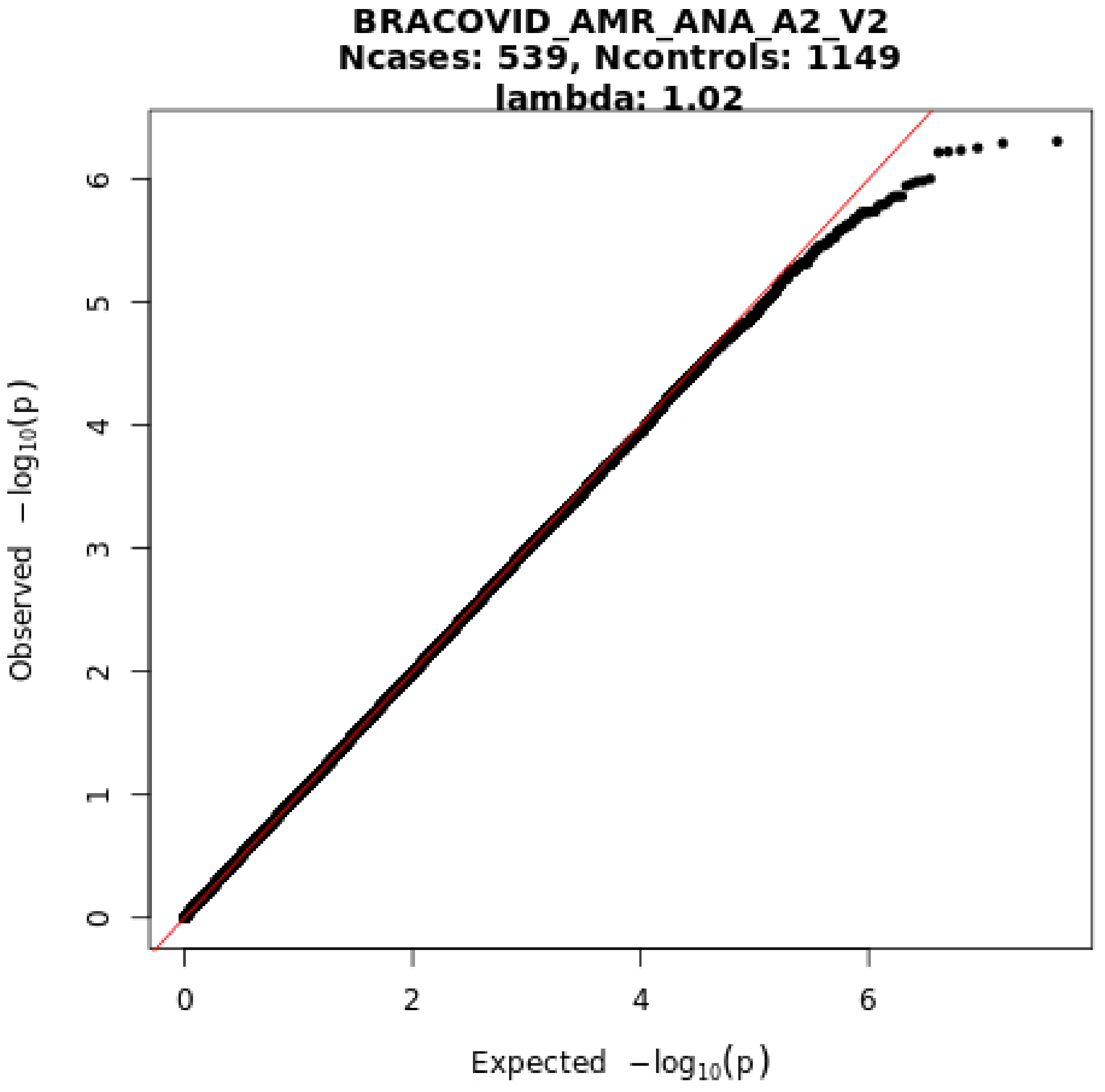
BRACOVID_AMR_ANA_A2_V2
Ncases: 539, Ncontrols: 1149
lambda: 1.02
Observed −log10(p)
Expected −log10(p)
0
1
2
3
4
5
6
0
2
4
6

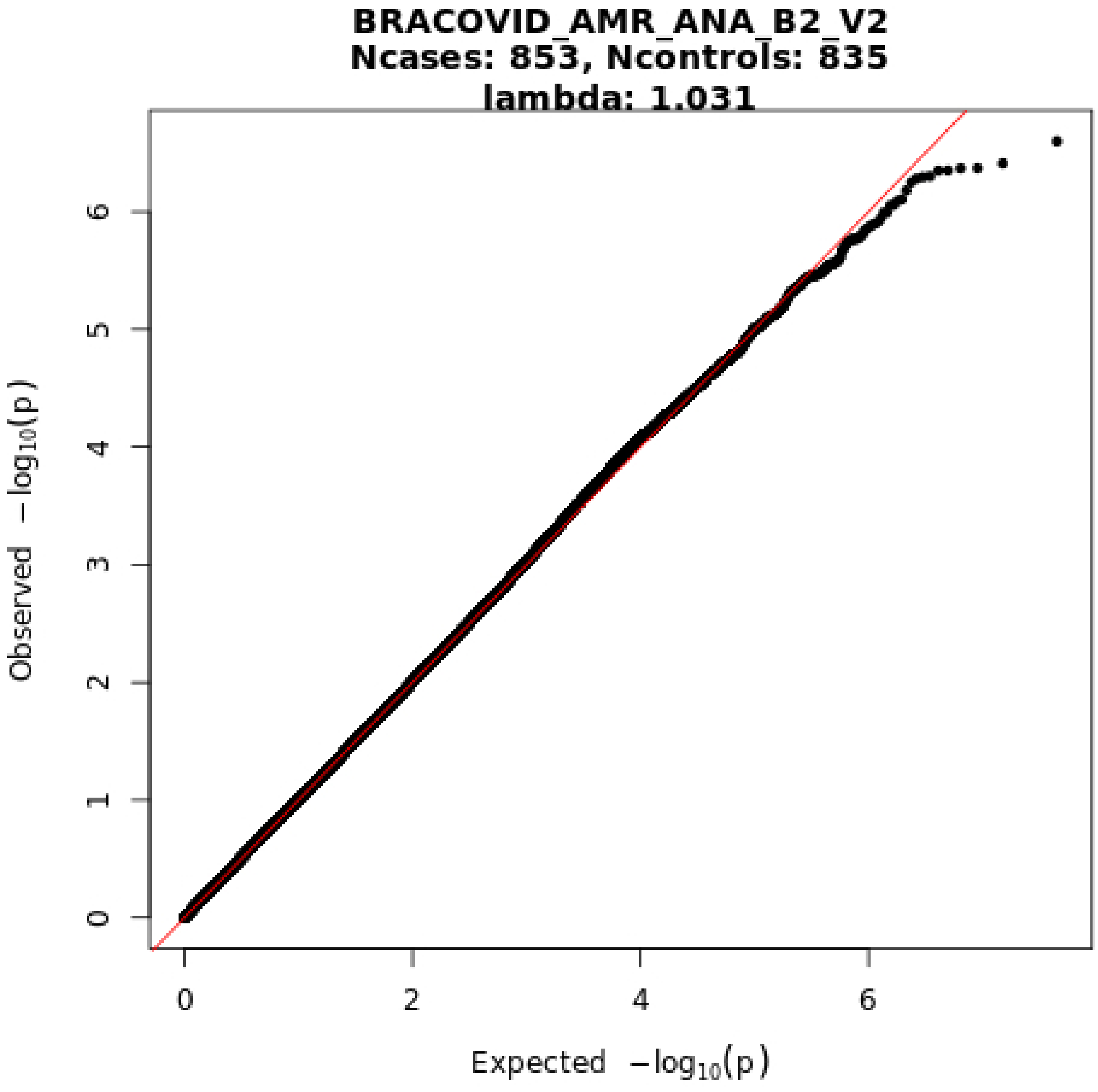
BRACOVID_AMR_ANA_B2_V2
Ncases: 853, Ncontrols: 835
lambda: 1.031
Observed −log10(p)
Expected −log10(p)
0
1
2
3
4
5
6
0
2
4
6

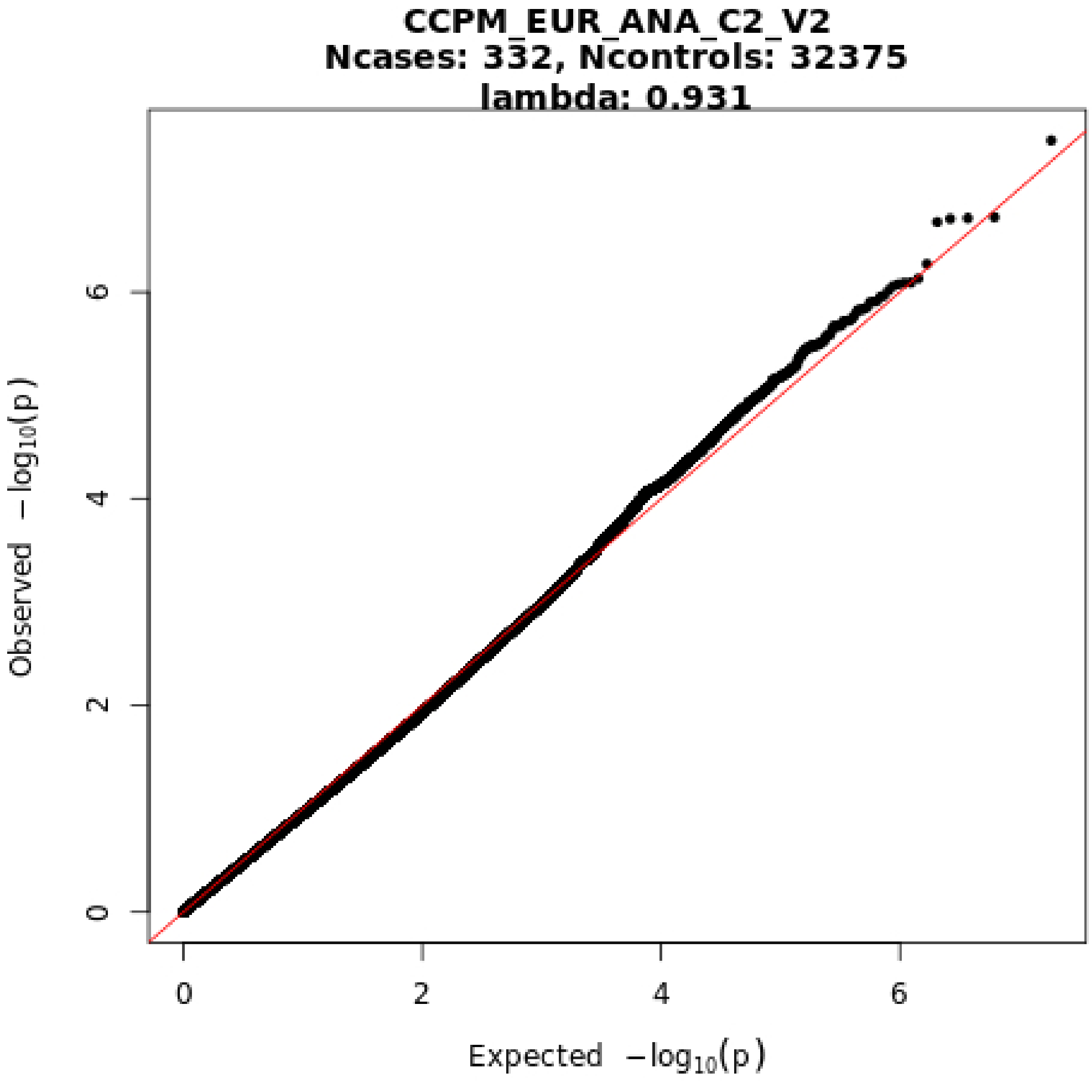
CCPM_EUR_ANA_C2_V2
Ncases: 332, Ncontrols: 32375
lambda: 0.931
Observed −log10(p)
Expected −log10(p)
0
2
4
6
0
2
4
6

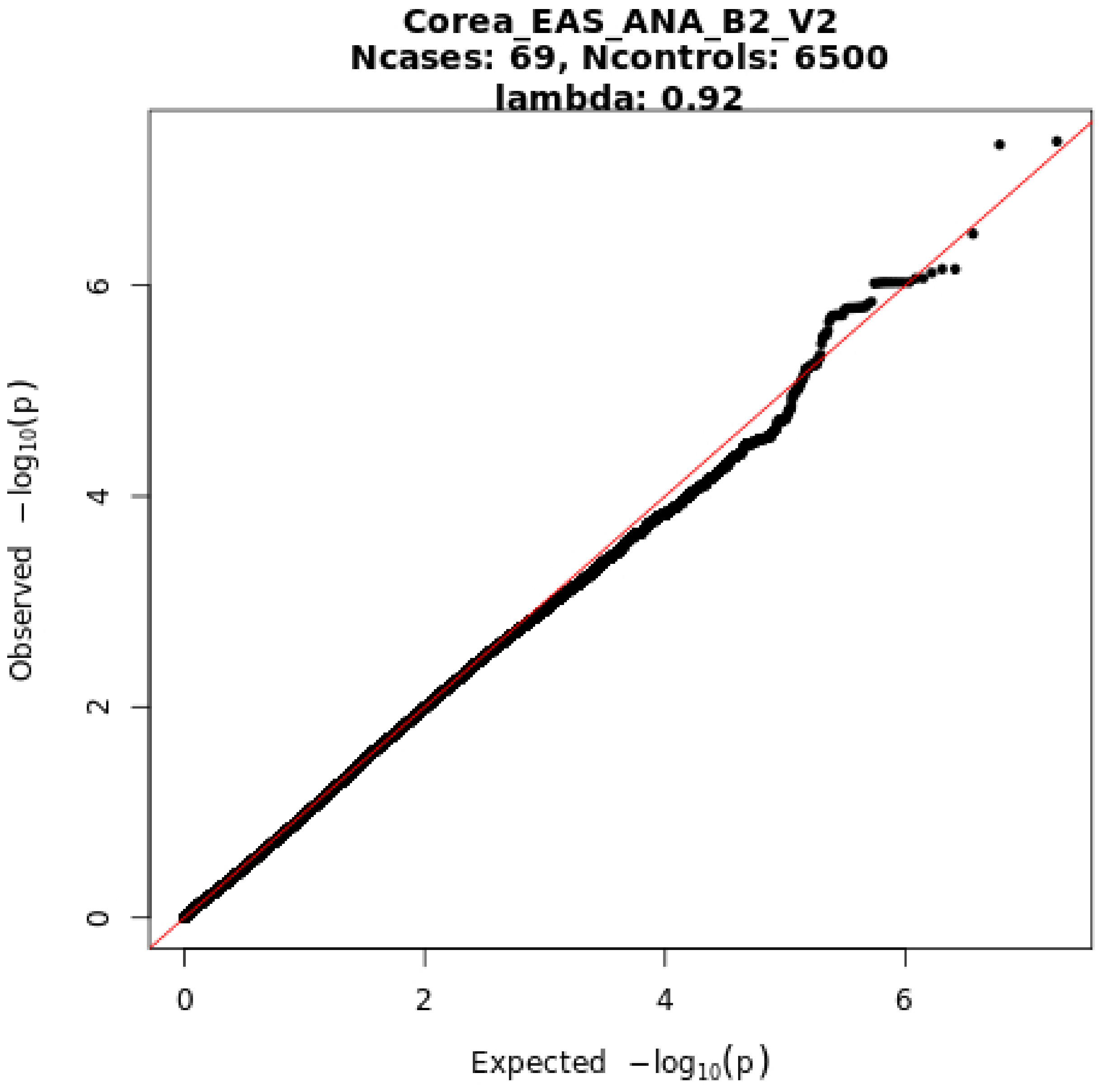
Corea_EAS_ANA_B2_V2
Ncases: 69, Ncontrols: 6500
lambda: 0.92
Observed −log10(p)
Expected −log10(p)
0
2
4
6
0
2
4
6

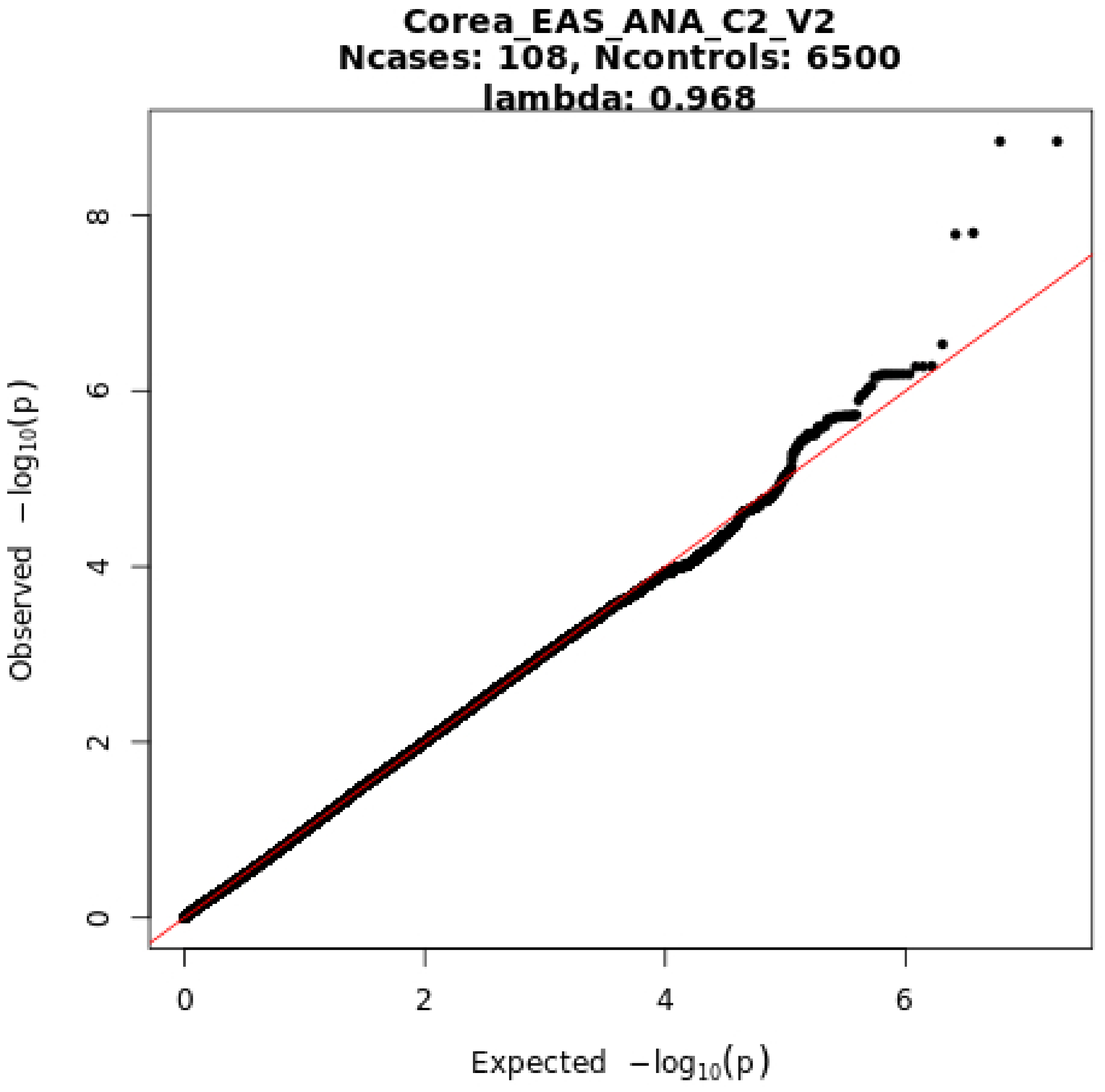
Corea_EAS_ANA_C2_V2
Ncases: 108, Ncontrols: 6500
lambda: 0.968
Observed −log10(p)
Expected −log10(p)
0
2
4
6
8

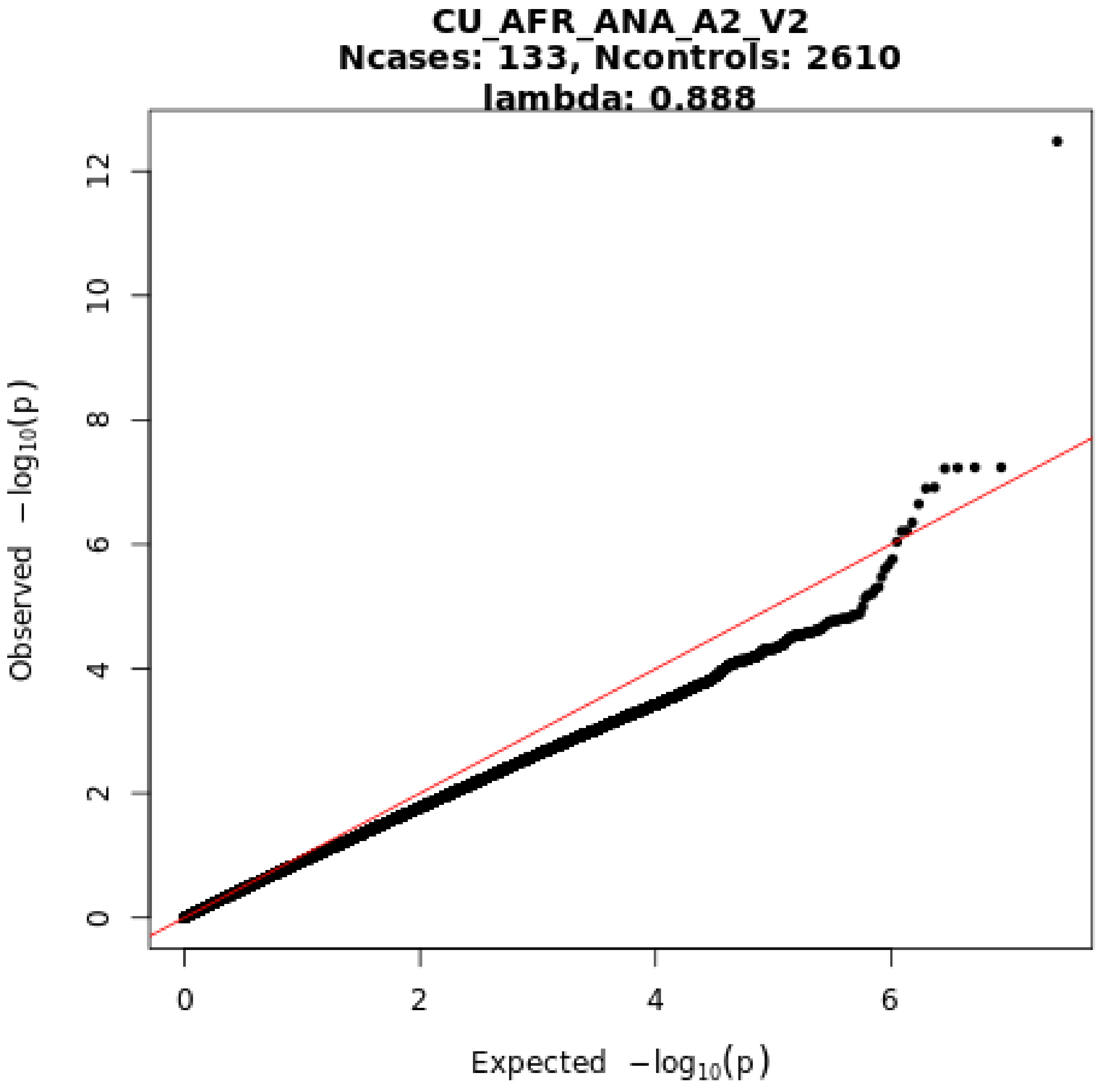
CU_AFR_ANA_A2_V2
Ncases: 133, Ncontrols: 2610
lambda: 0.888
Observed −log10(p)
Expected −log10(p)
0
2
4
6
8
10
12
0
2
4
6

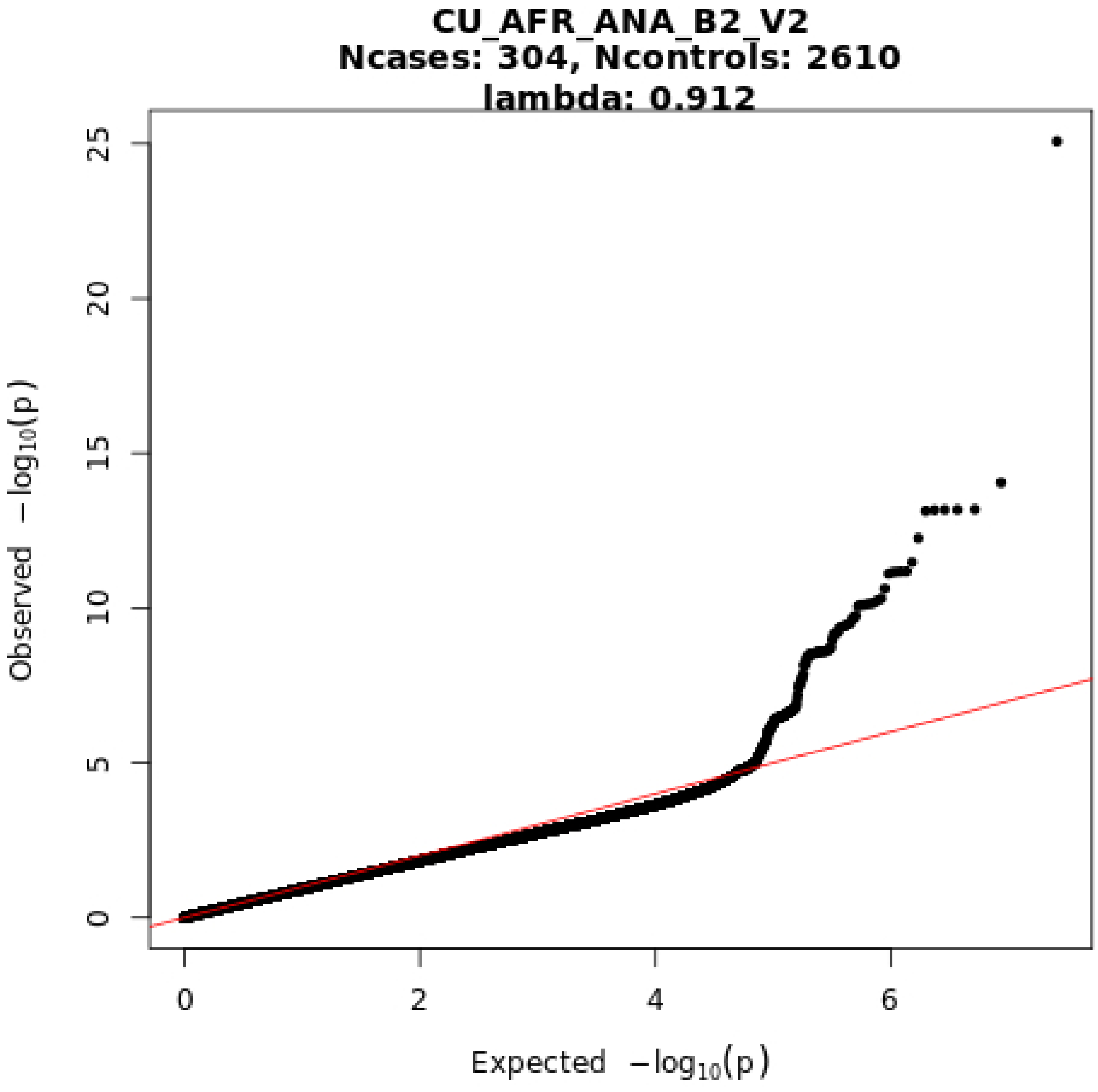
CU_AFR_ANA_B2_V2
Ncases: 304, Ncontrols: 2610
lambda: 0.912
Observed −log10(p)
Expected −log10(p)
0
5
10
15
20
25
0
2
4
6

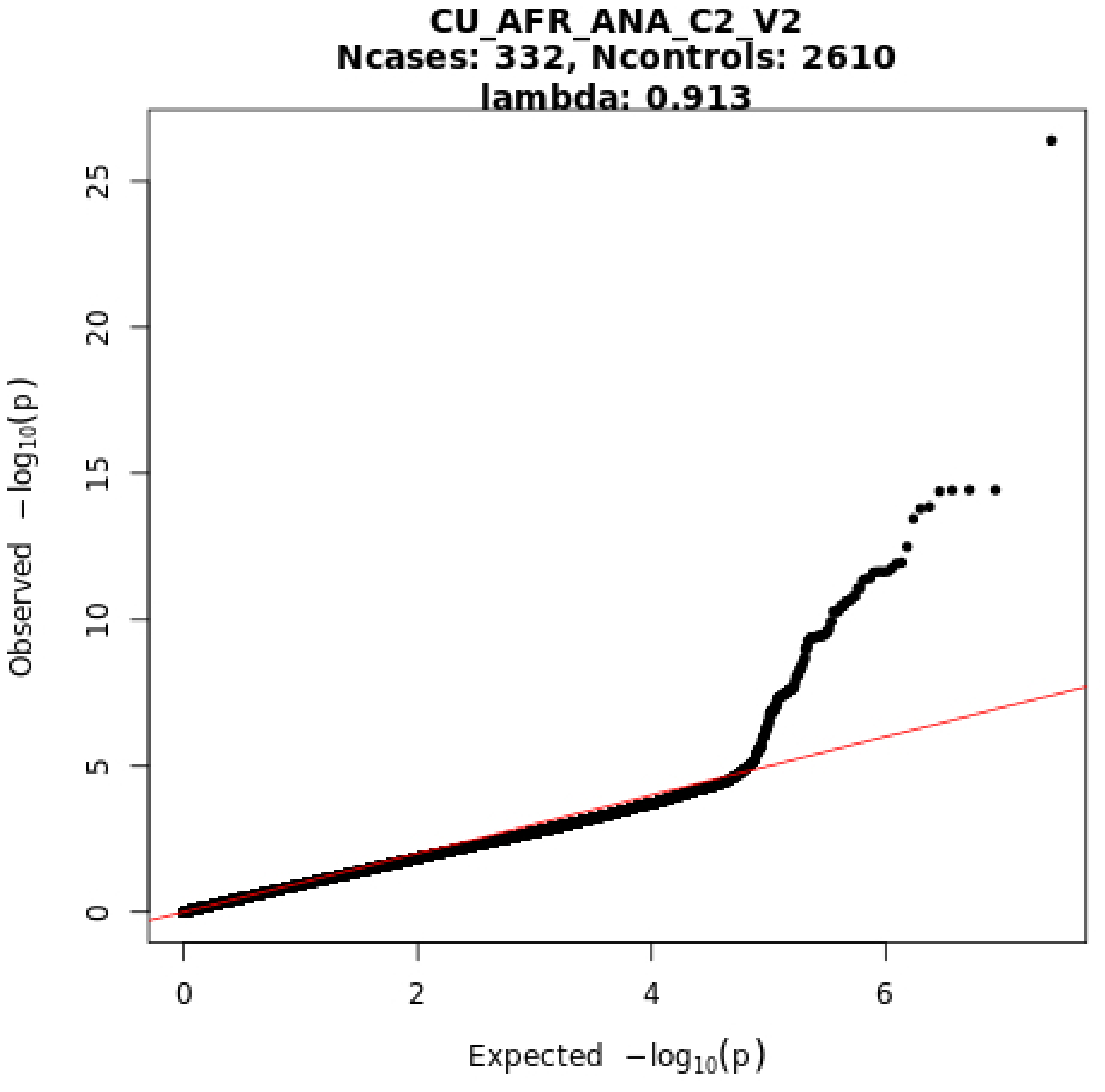

CU_AFR_ANA_C2_V2
Ncases: 332, Ncontrols: 2610
lambda: 0.913
Observed −log10(p)
Expected −log10(p)
0
5
10
15
20
25
0
2
4
6

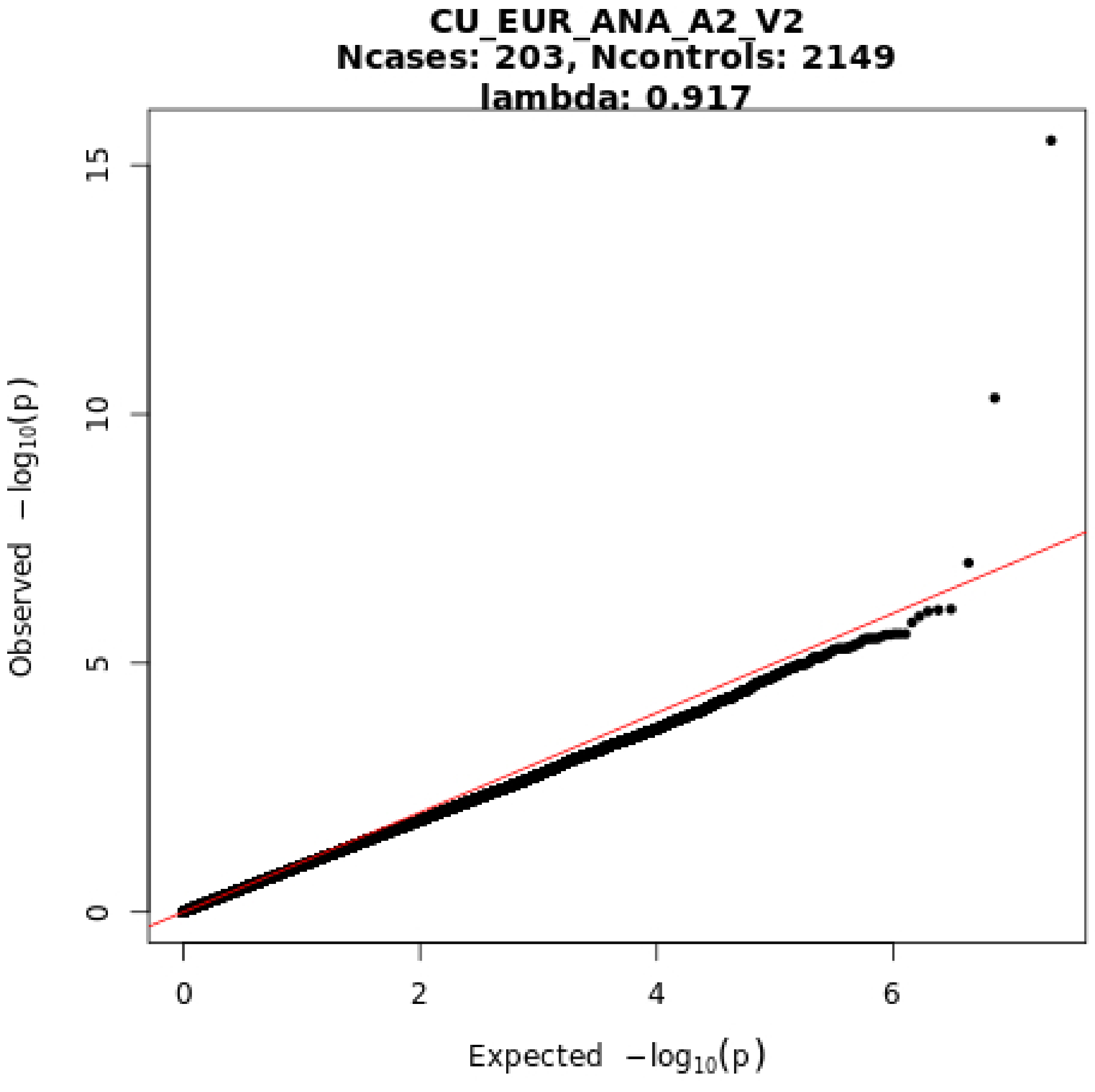
CU_EUR_ANA_A2_V2
Ncases: 203, Ncontrols: 2149
lambda: 0.917
Observed −log10(p)
Expected −log10(p)
0
5
10
15
0
2
4
6

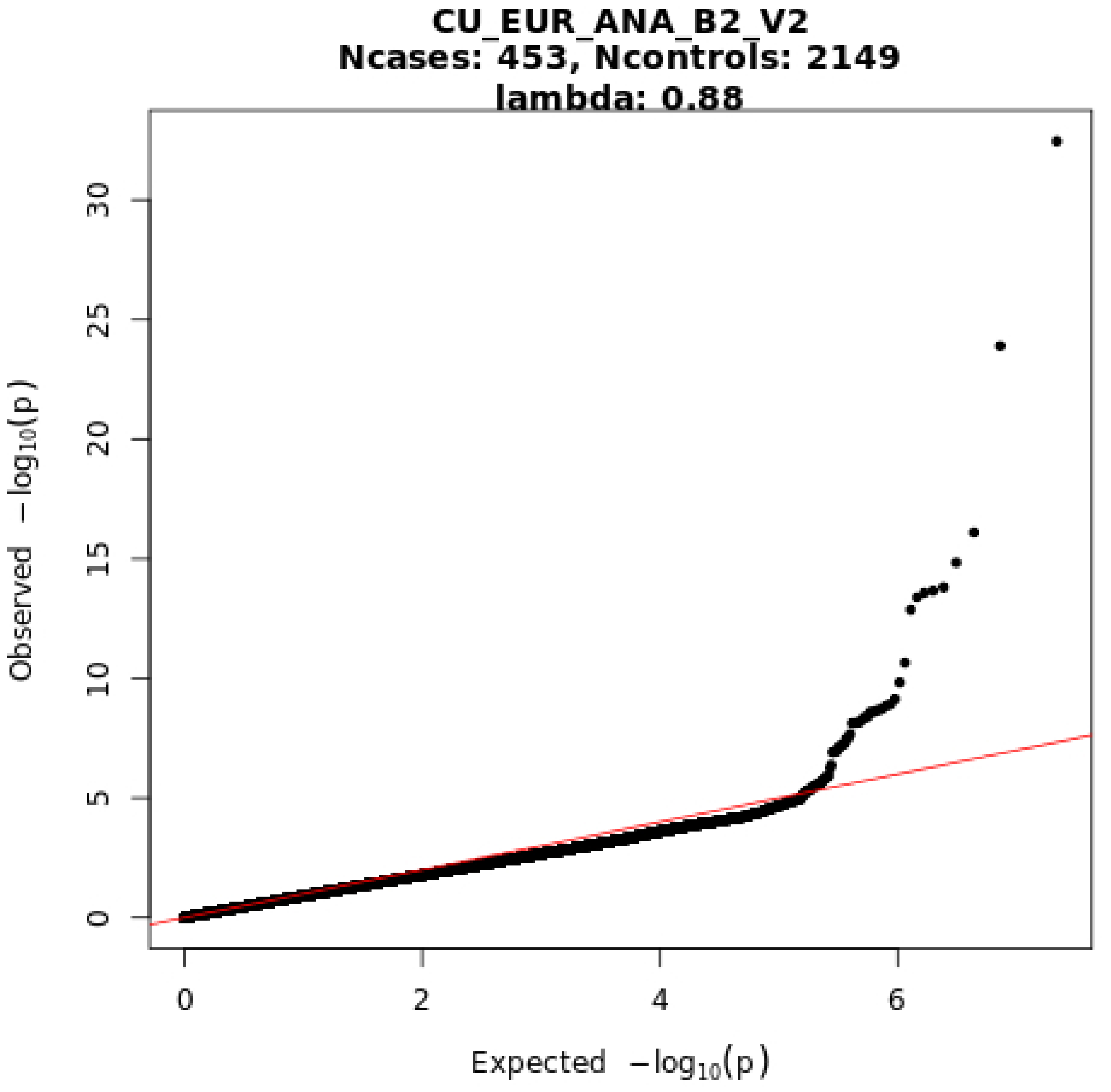
CU_EUR_ANA_B2_V2
Ncases: 453, Ncontrols: 2149
lambda: 0.88
Observed $-\log_{10}(p)$
Expected $-\log_{10}(p)$
0
5
10
15
20
25
30
0
2
4
6

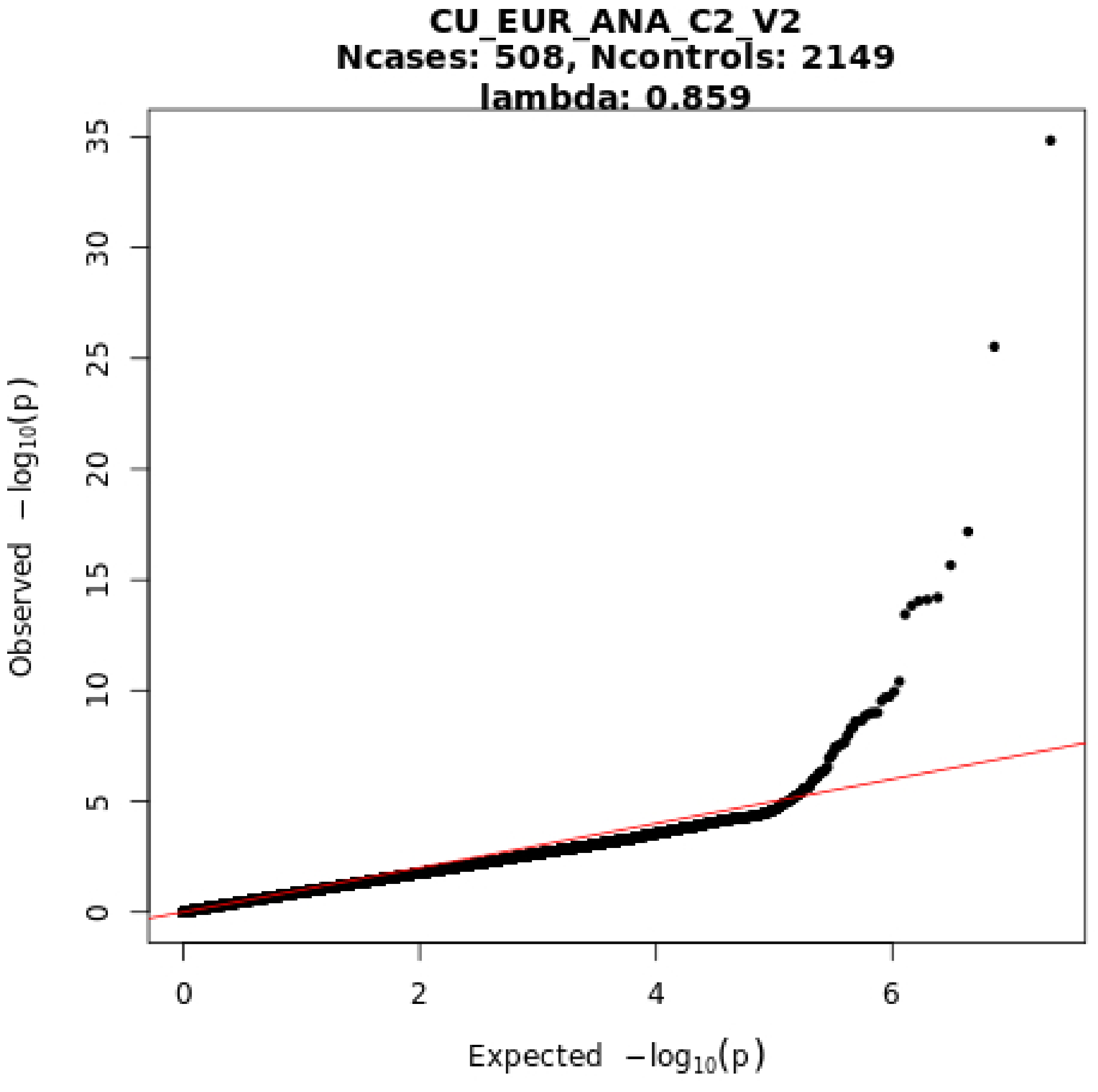
CU_EUR_ANA_C2_V2
Ncases: 508, Ncontrols: 2149
lambda: 0.859
Observed −log10(p)
Expected −log10(p)
0
5
10
15
20
25
30
35
0
2
4
6

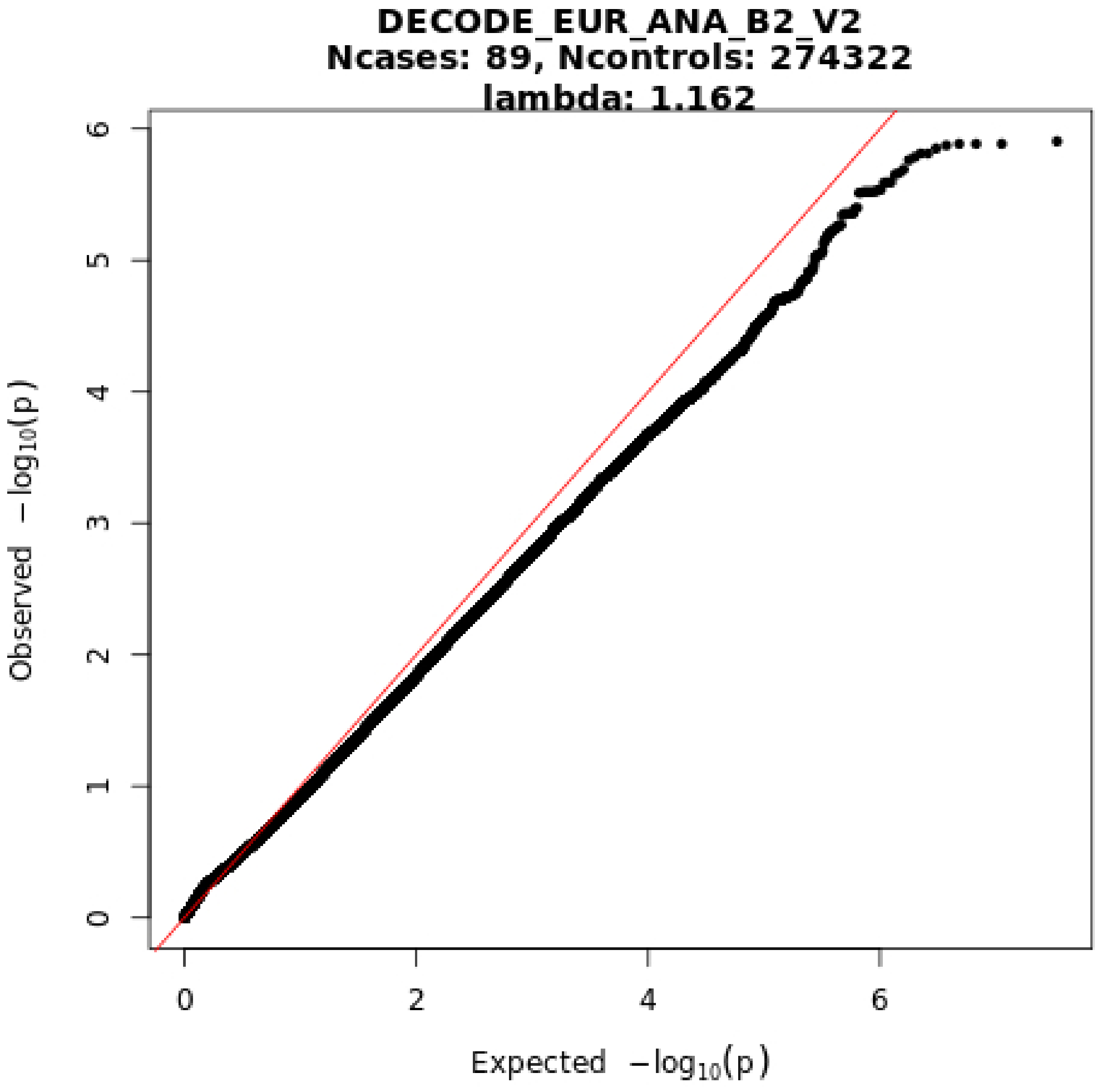
DECODE_EUR_ANA_B2_V2
Ncases: 89, Ncontrols: 274322
lambda: 1.162
Observed −log10(p)
Expected −log10(p)
0
1
2
3
4
5
6
0
2
4
6

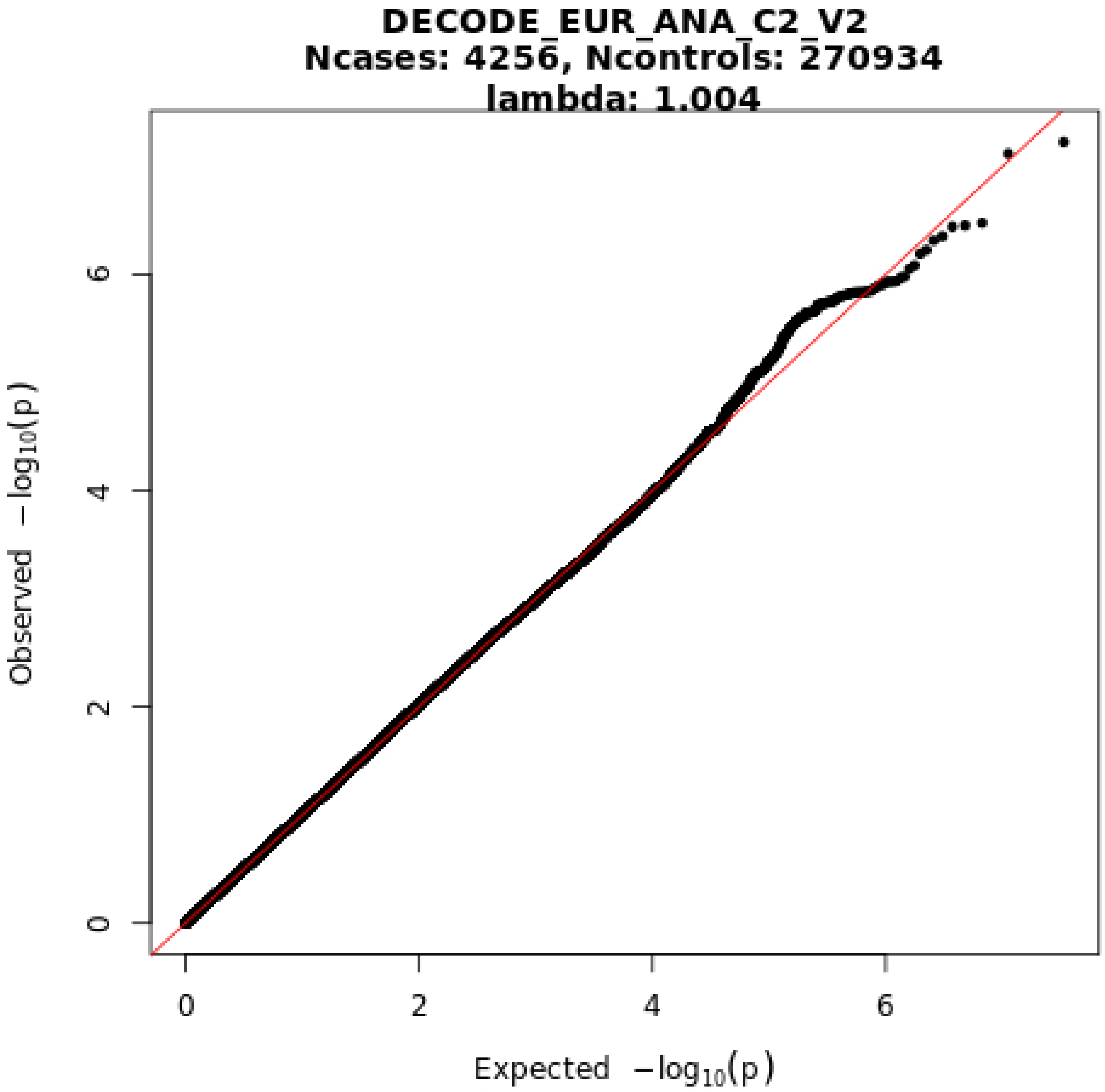
DECODE_EUR_ANA_C2_V2
Ncases: 4256, Ncontrols: 270934
lambda: 1.004
Observed −log10(p)
Expected −log10(p)
0
2
4
6

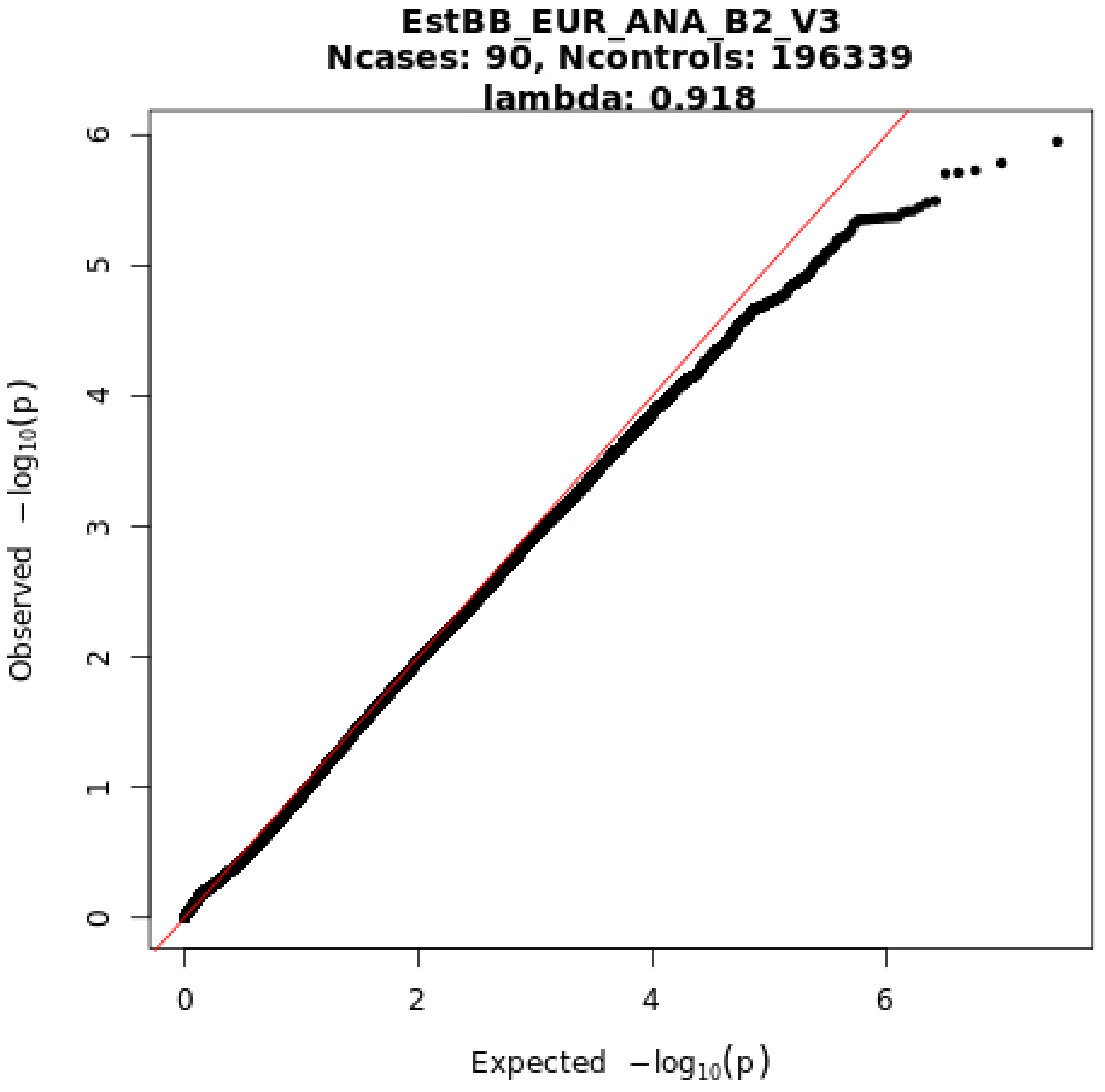
EstBB_EUR_ANA_B2_V3
Ncases: 90, Ncontrols: 196339
lambda: 0.918
Observed −log10(p)
Expected −log10(p)
0
1
2
3
4
5
6
0
2
4
6

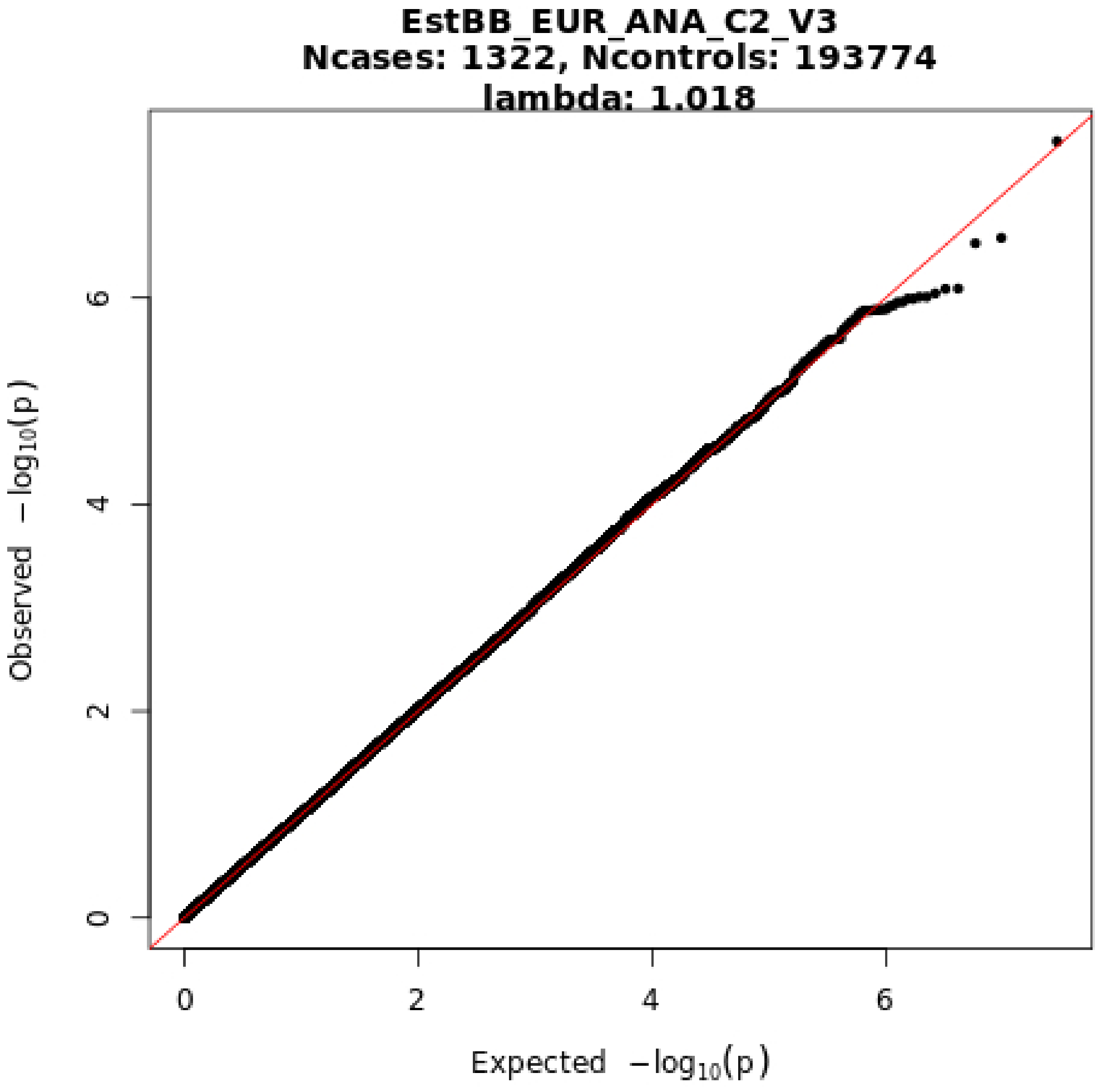
EstBB_EUR_ANA_C2_V3
Ncases: 1322, Ncontrols: 193774
lambda: 1.018
Observed −log10(p)
Expected −log10(p)
0
2
4
6
0
2
4
6

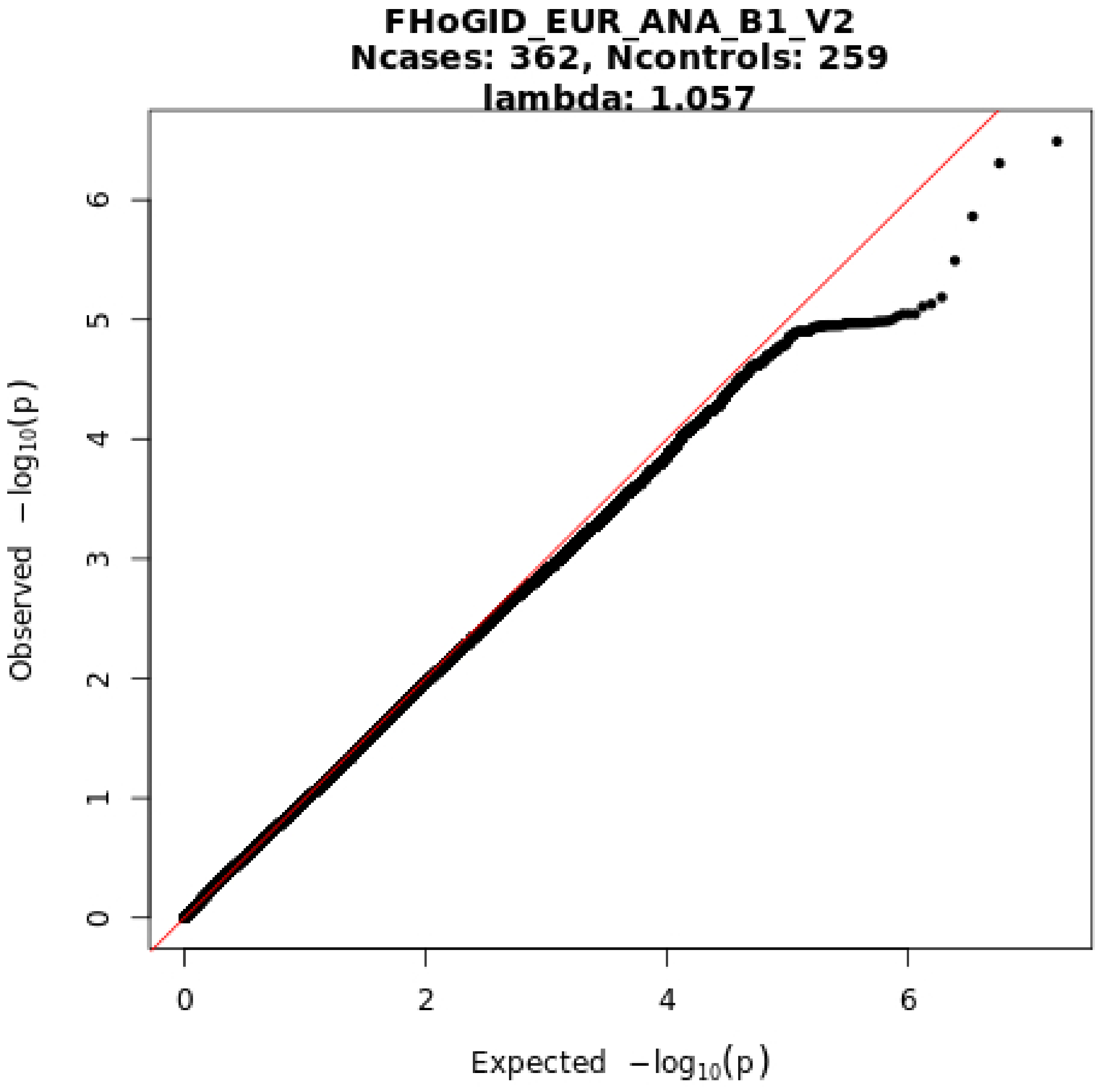
FHoGID_EUR_ANA_B1_V2
Ncases: 362, Ncontrols: 259
lambda: 1.057
Observed −log10(p)
Expected −log10(p)
0
1
2
3
4
5
6
0
2
4
6

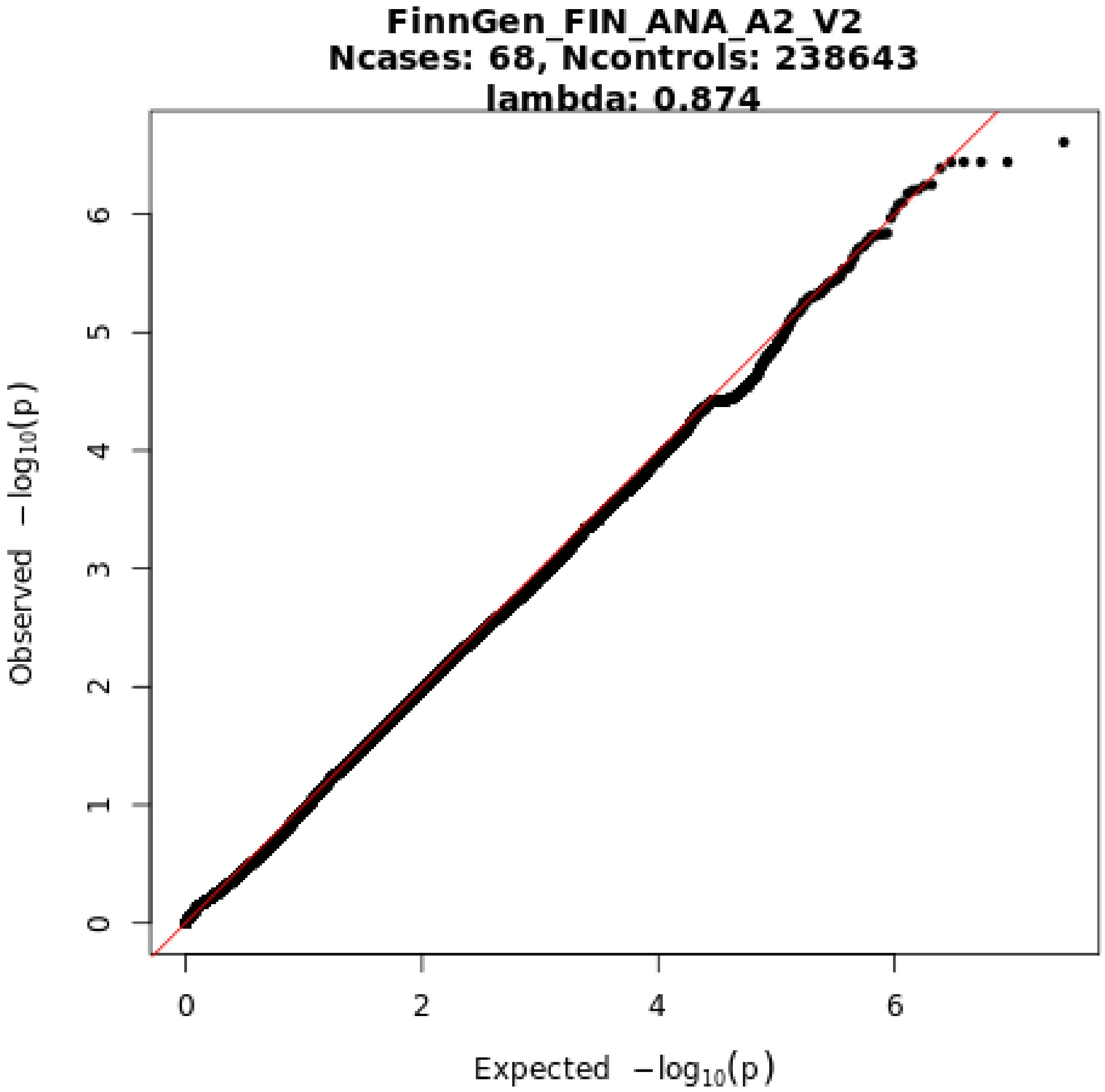
FinnGen_FIN_ANA_A2_V2
Ncases: 68, Ncontrols: 238643
lambda: 0.874
Observed −log10(p)
Expected −log10(p)
0
1
2
3
4
5
6
0
2
4
6

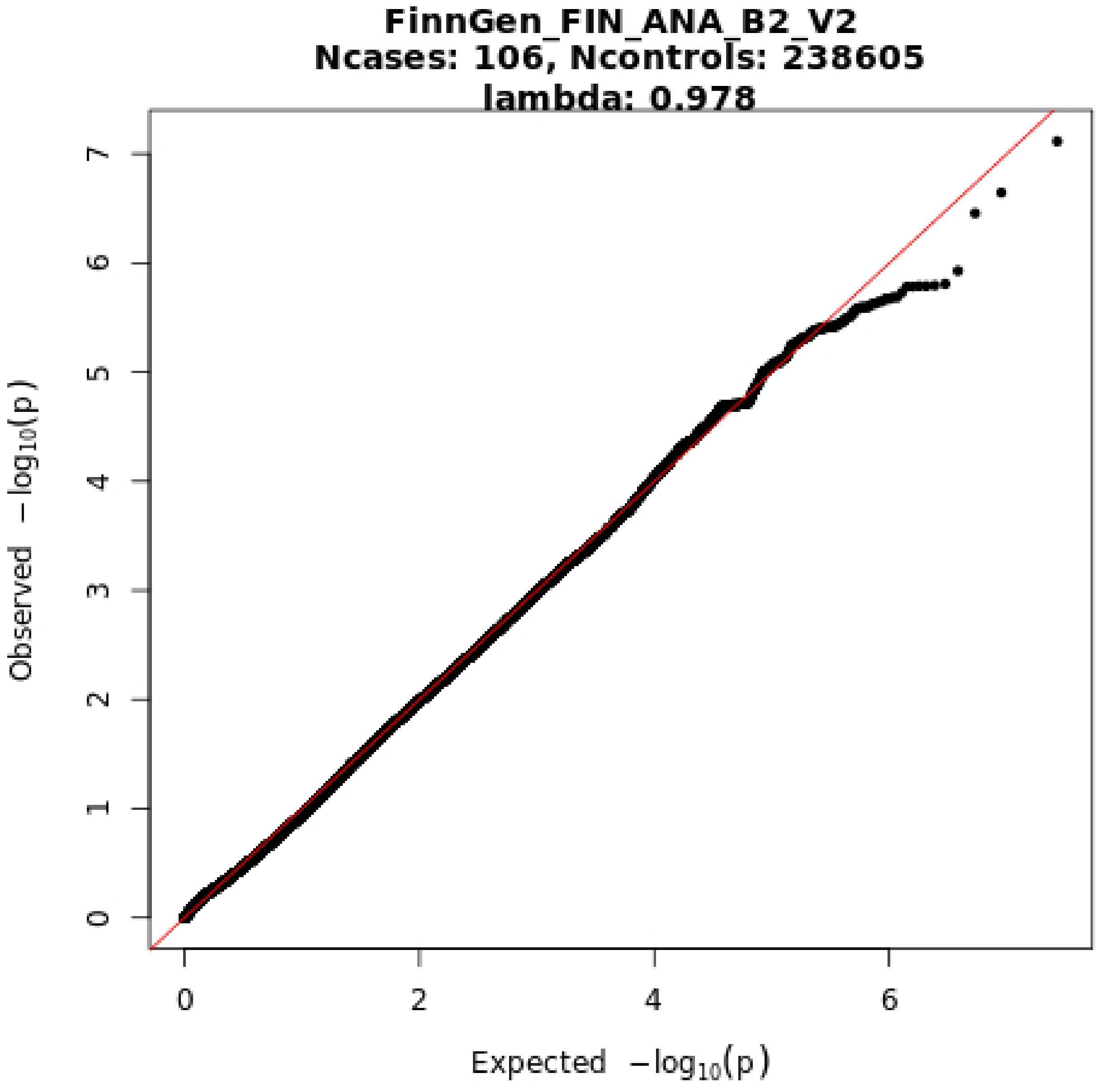
FinnGen_FIN_ANA_B2_V2
Ncases: 106, Ncontrols: 238605
lambda: 0.978
Observed −log10(p)
Expected −log10(p)
0
1
2
3
4
5
6
7

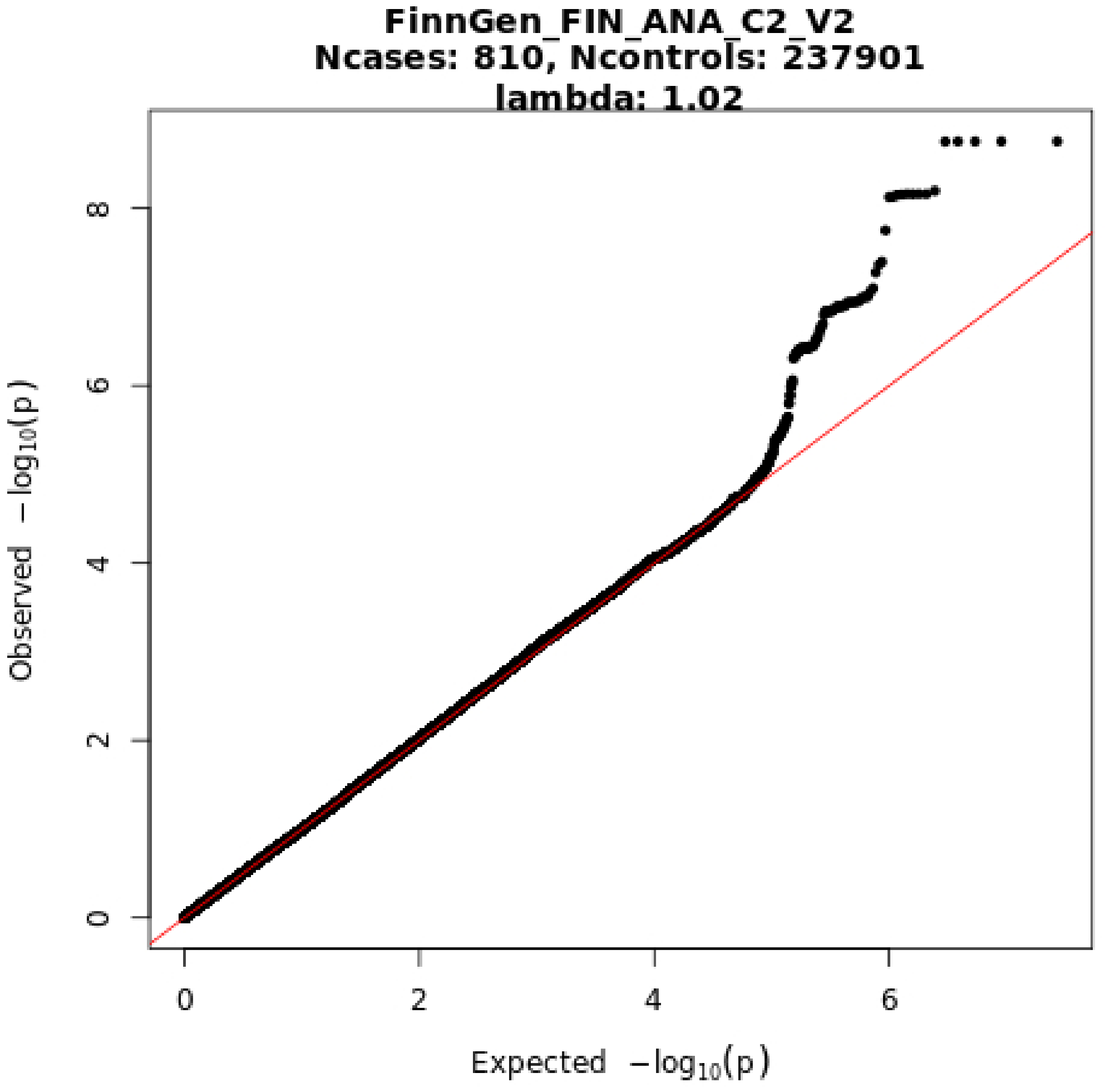
FinnGen_FIN_ANA_C2_V2
Ncases: 810, Ncontrols: 237901
lambda: 1.02
Observed −log10(p)
Expected −log10(p)
0
2
4
6
8
0
2
4
6

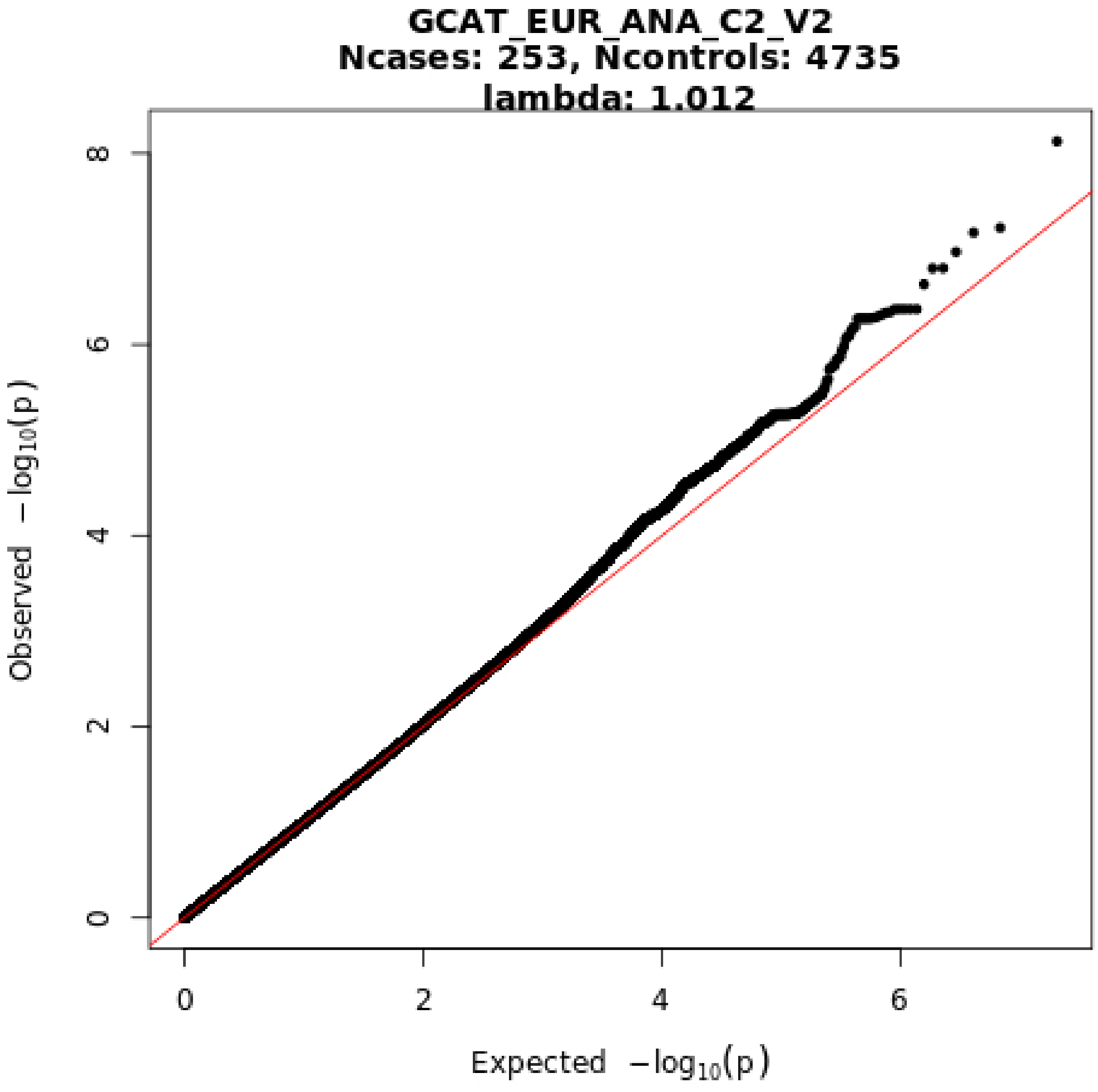
GCAT_EUR_ANA_C2_V2
Ncases: 253, Ncontrols: 4735
lambda: 1.012
Observed −log10(p)
Expected −log10(p)
0
2
4
6
8
0
2
4
6

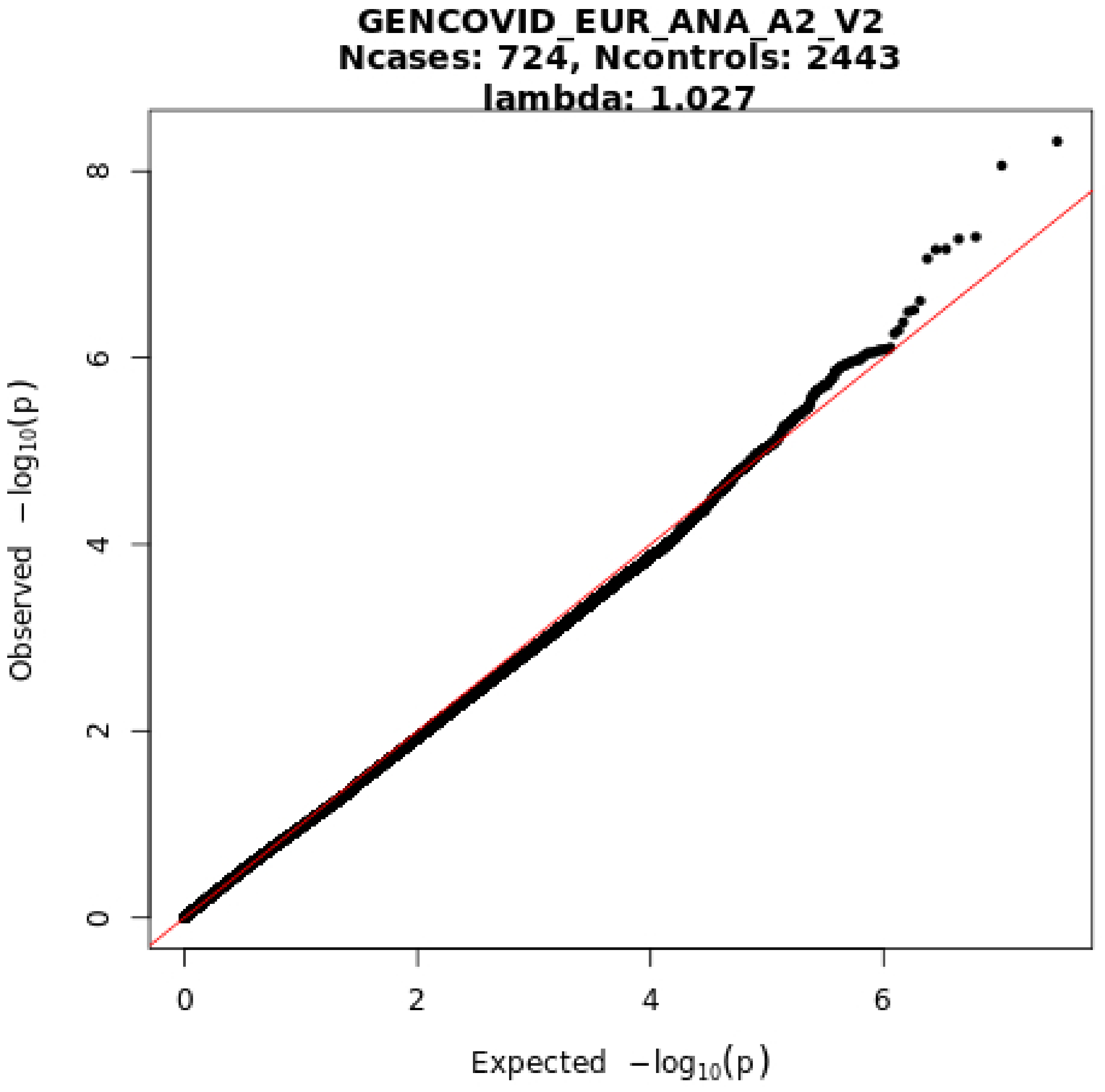
GENCOVID_EUR_ANA_A2_V2
Ncases: 724, Ncontrols: 2443
lambda: 1.027
Observed $-\log_{10}(p)$
Expected $-\log_{10}(p)$
0
2
4
6
8

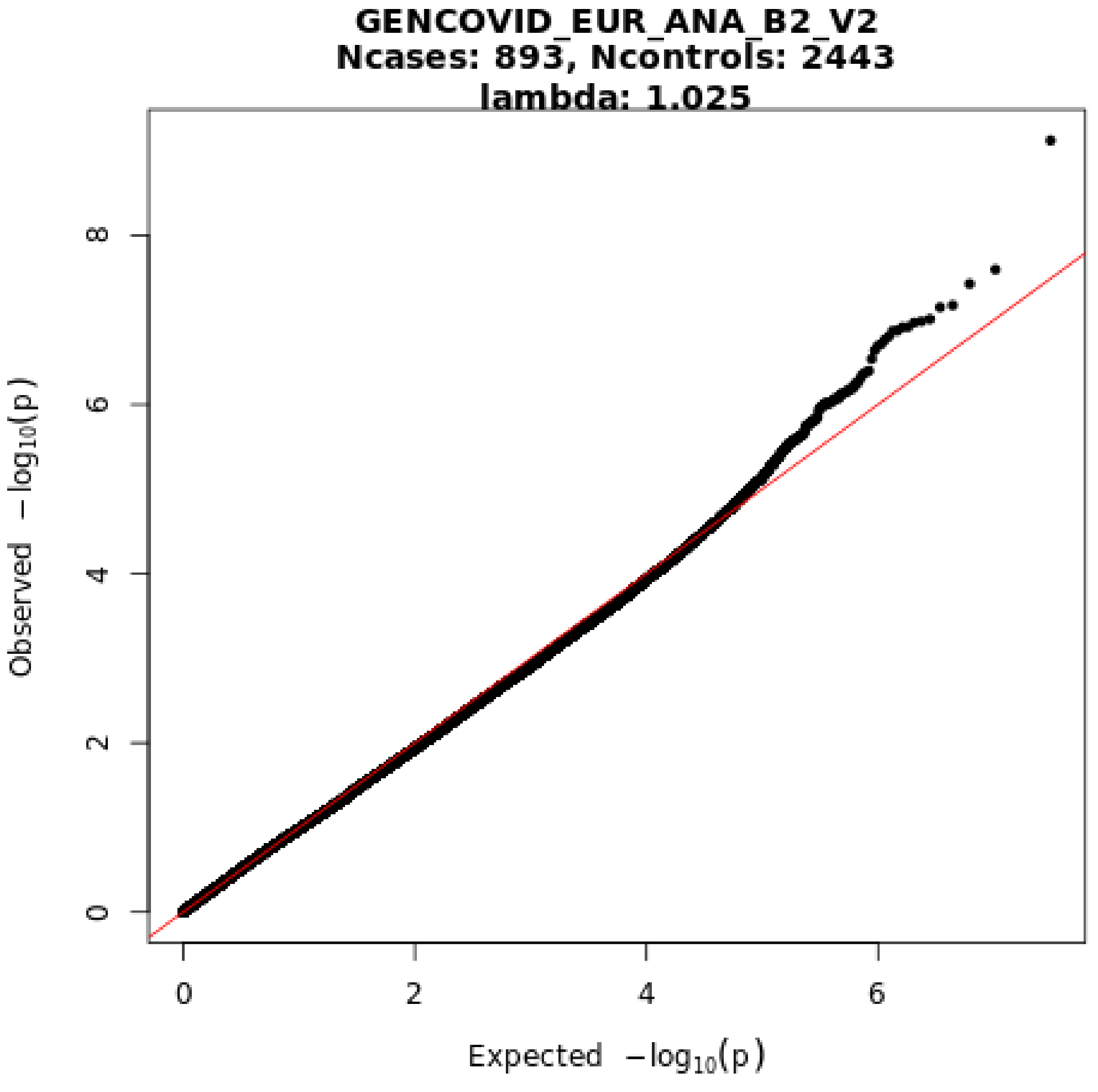
GENCOVID_EUR_ANA_B2_V2
Ncases: 893, Ncontrols: 2443
lambda: 1.025
Observed −log10(p)
Expected −log10(p)
0
2
4
6
8
0
2
4
6

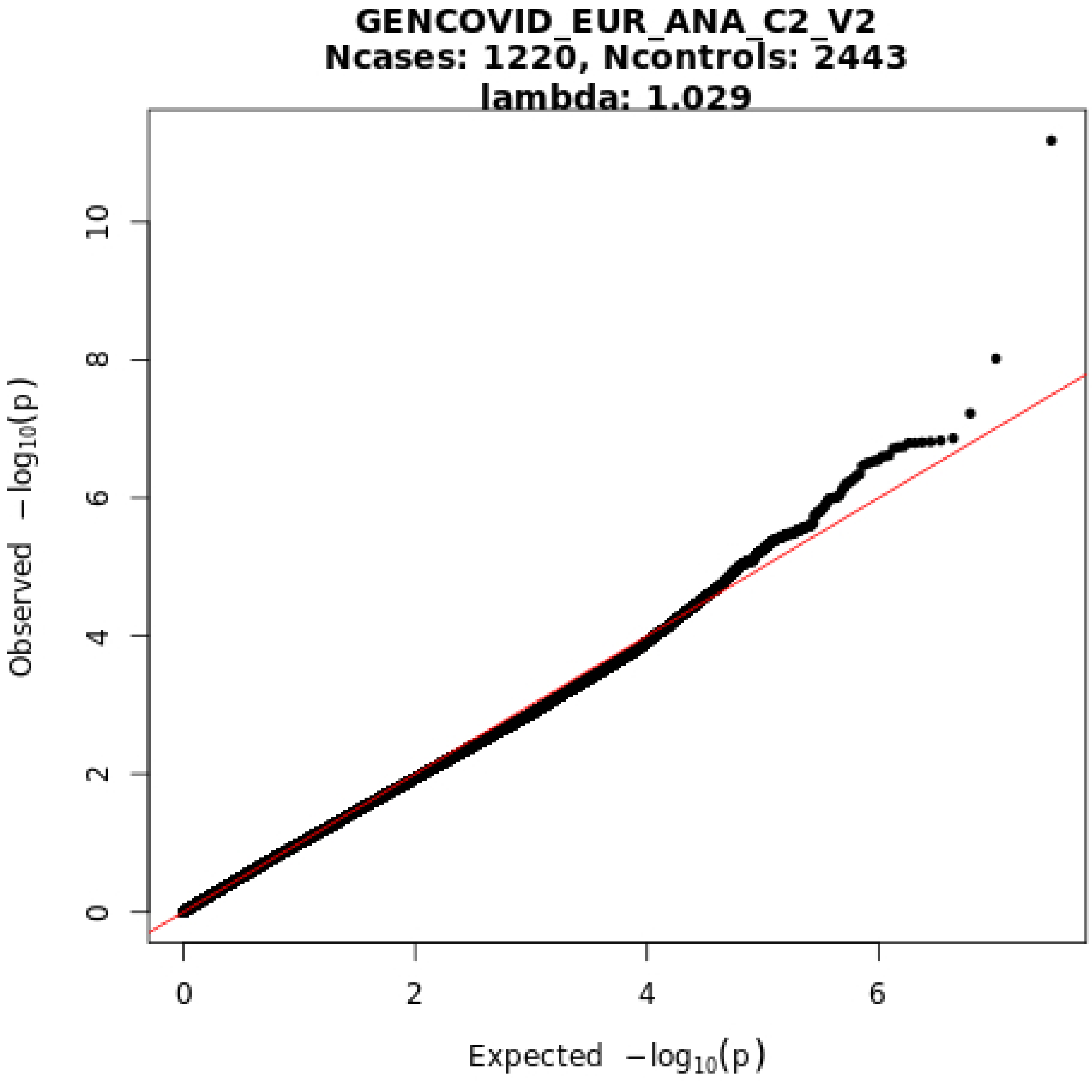
GENCOVID_EUR_ANA_C2_V2
Ncases: 1220, Ncontrols: 2443
lambda: 1.029
Observed −log10(p)
Expected −log10(p)
0
2
4
6
8
10
0
2
4
6

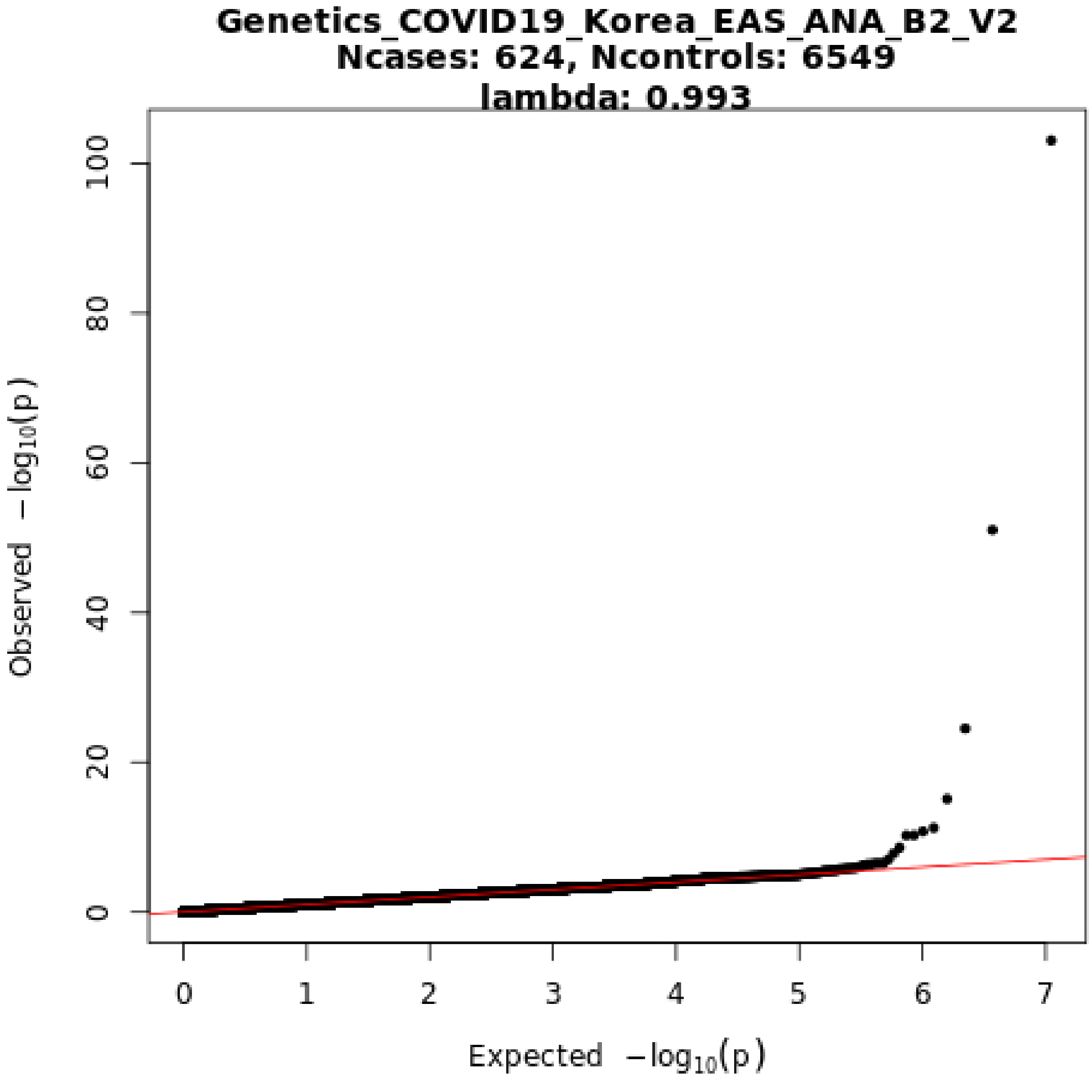
Genetics_COVID19_Korea_EAS_ANA_B2_V2
Ncases: 624, Ncontrols: 6549
lambda: 0.993
Observed −log10(p)
Expected −log10(p)
0
20
40
60
80
100
0
1
2
3
4
5
6
7

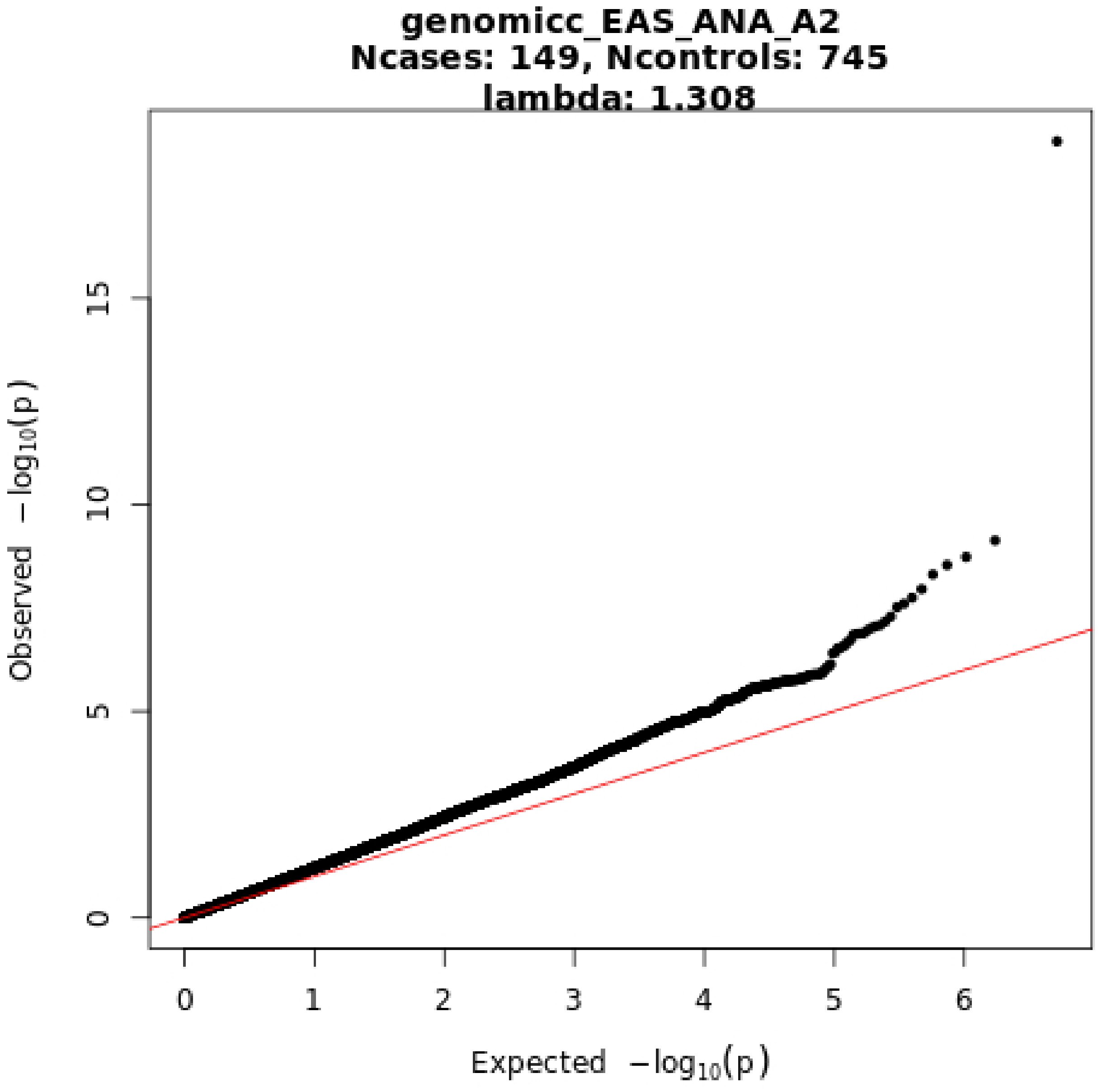
genomicc_EAS_ANA_A2
Ncases: 149, Ncontrols: 745
lambda: 1.308
Observed −log10(p)
Expected −log10(p)
0
5
10
15
0
1
2
3
4
5
6

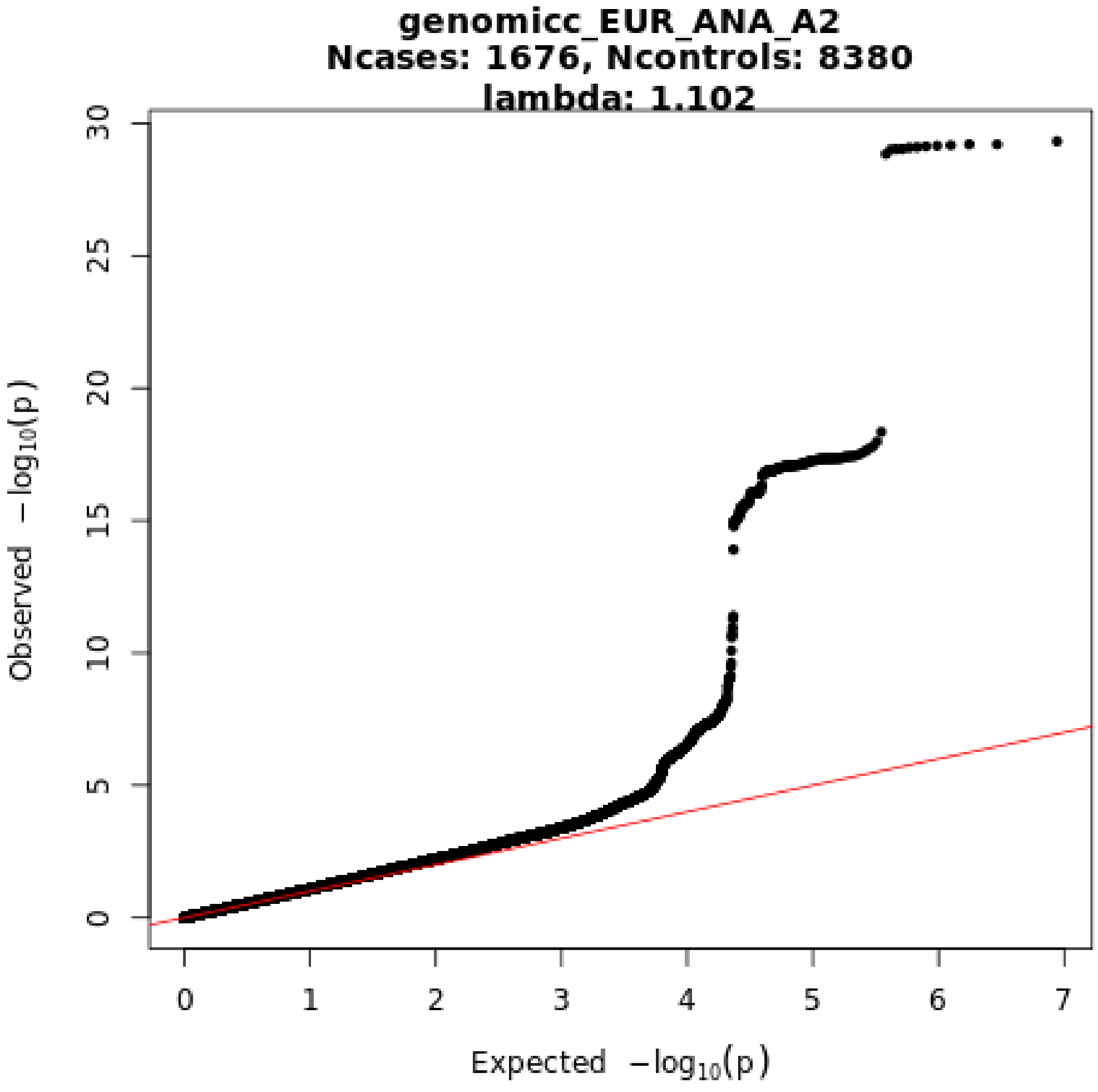

genomicc_EUR_ANA_A2
Ncases: 1676, Ncontrols: 8380
lambda: 1.102
Observed −log10(p)
Expected −log10(p)
0
5
10
15
20
25
30
0
1
2
3
4
5
6
7

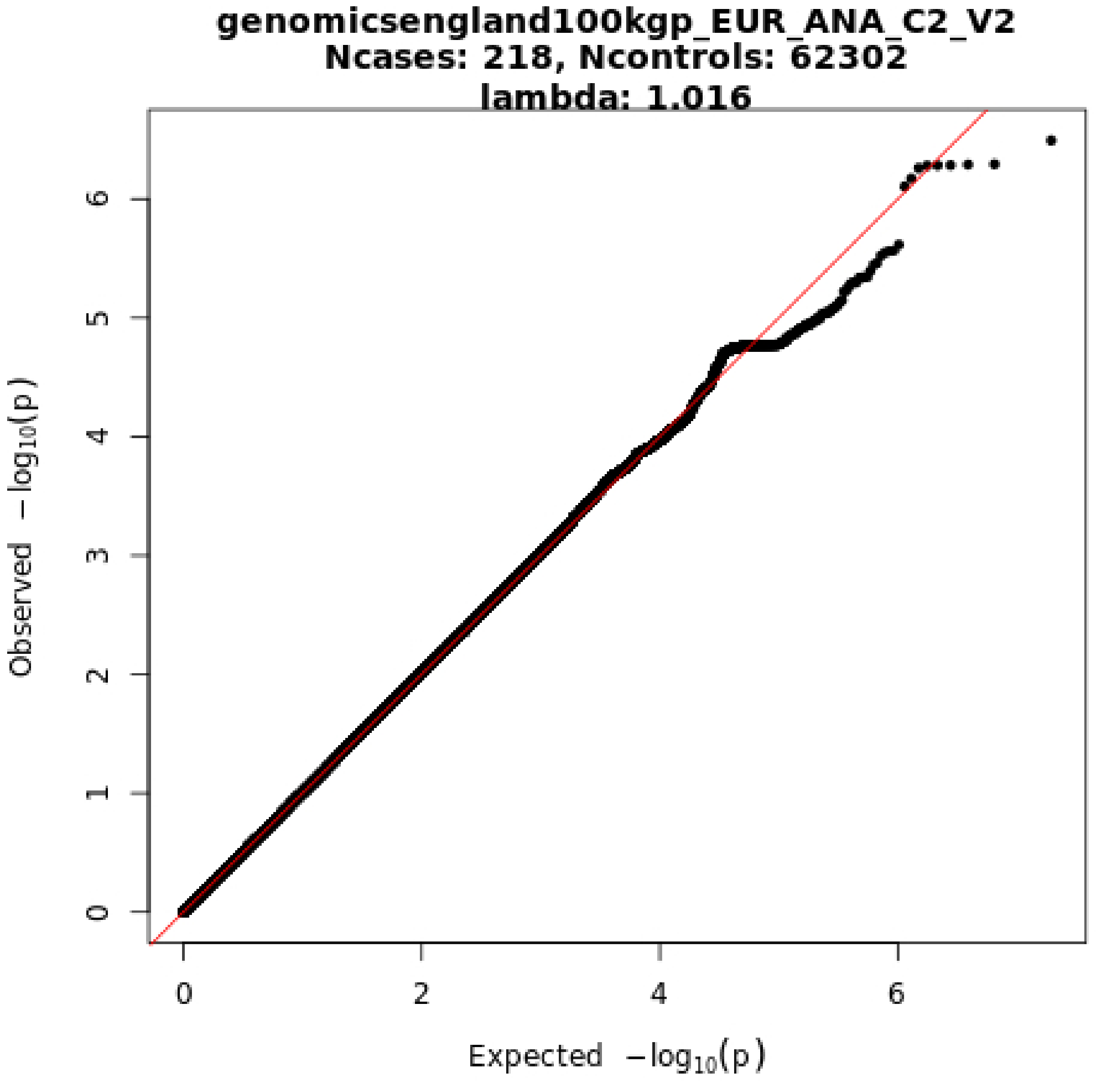
genomicsengland100kgp_EUR_ANA_C2_V2
Ncases: 218, Ncontrols: 62302
lambda: 1.016
Observed −log10(p)
Expected −log10(p)
0
1
2
3
4
5
6
0
2
4
6

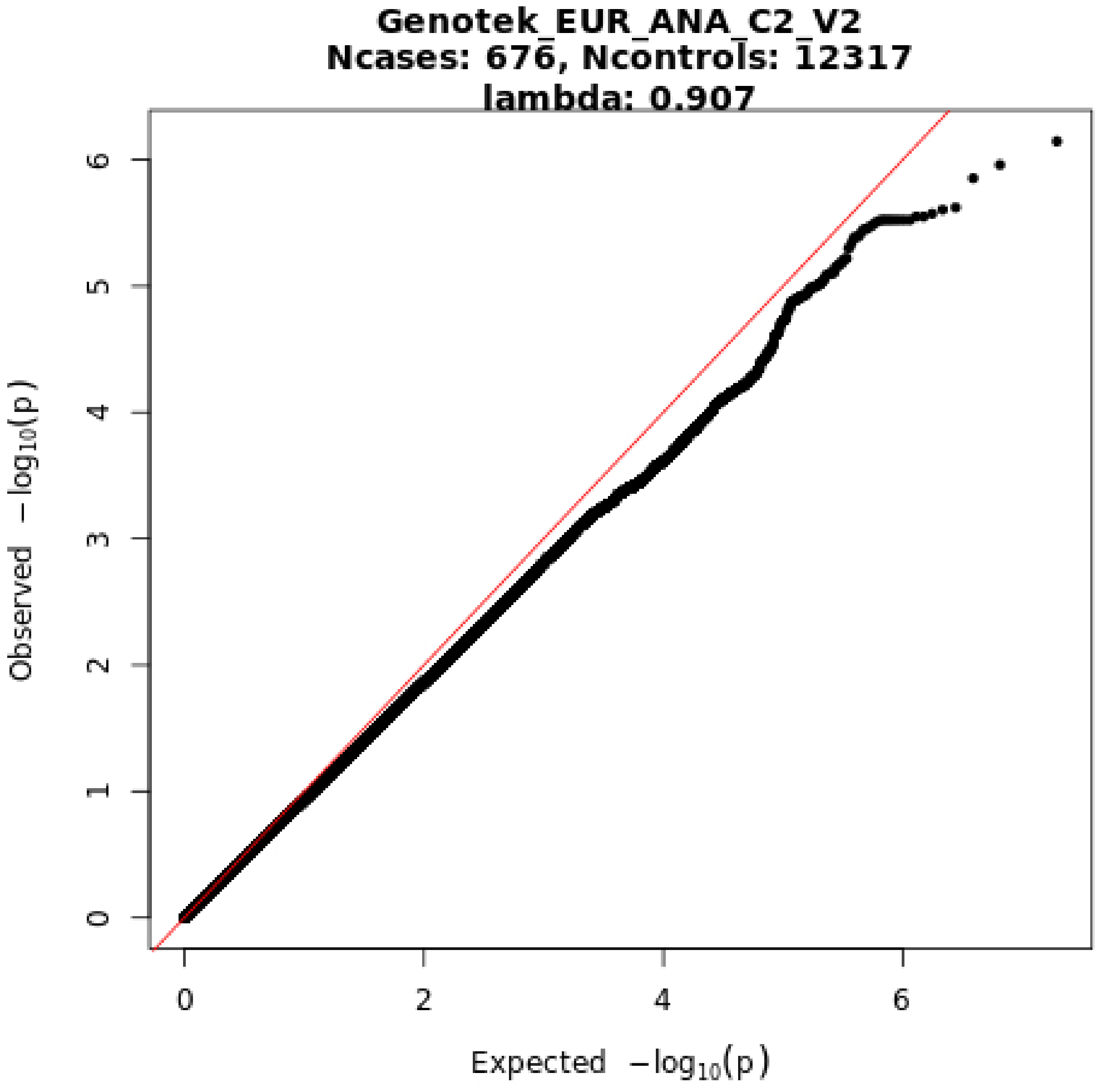
Genotek_EUR_ANA_C2_V2
Ncases: 676, Ncontrols: 12317
lambda: 0.907
Observed −log10(p)
Expected −log10(p)
0
1
2
3
4
5
6
0
2
4
6

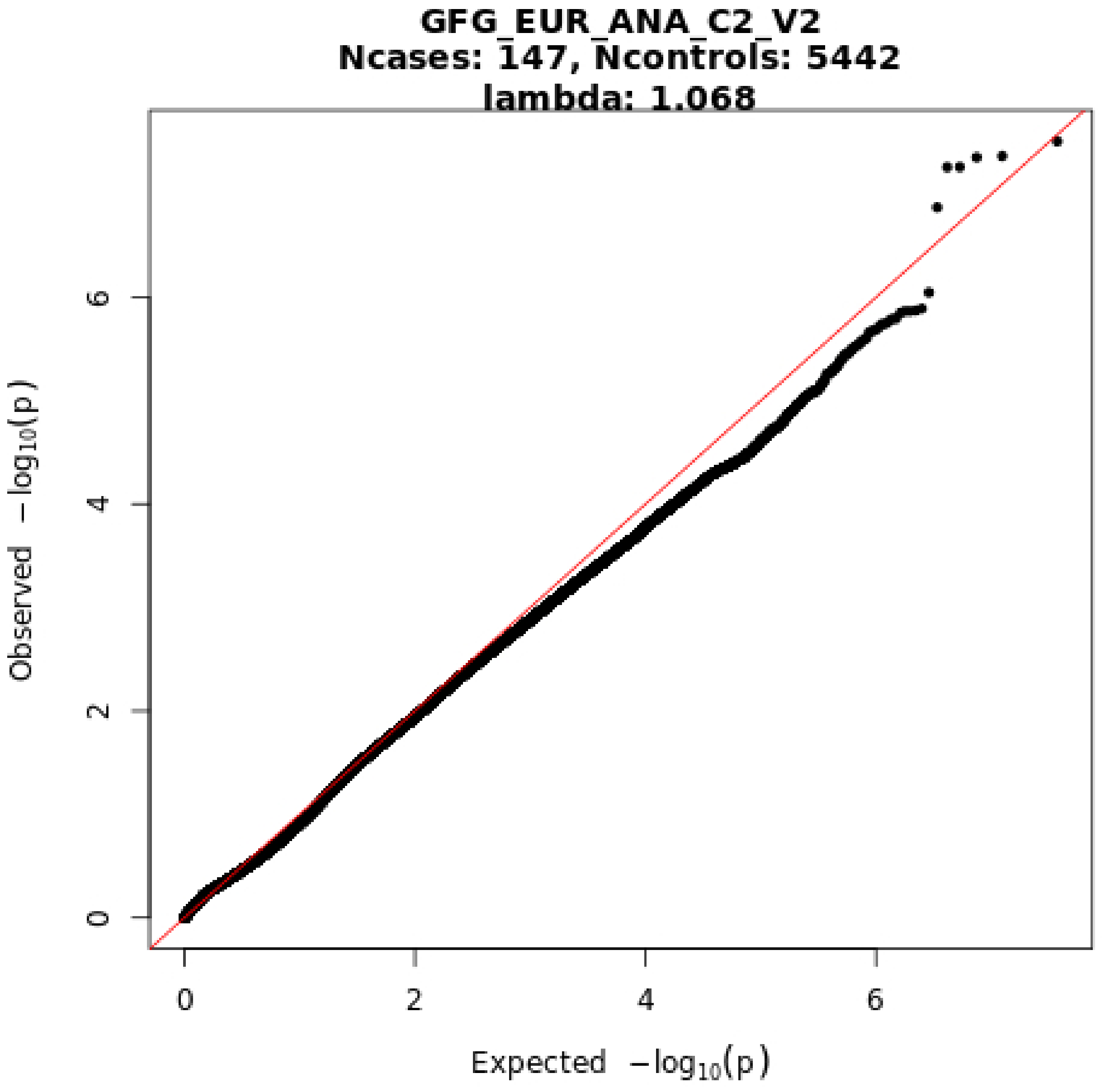
GFG_EUR_ANA_C2_V2
Ncases: 147, Ncontrols: 5442
lambda: 1.068
Observed −log10(p)
Expected −log10(p)
0
2
4
6

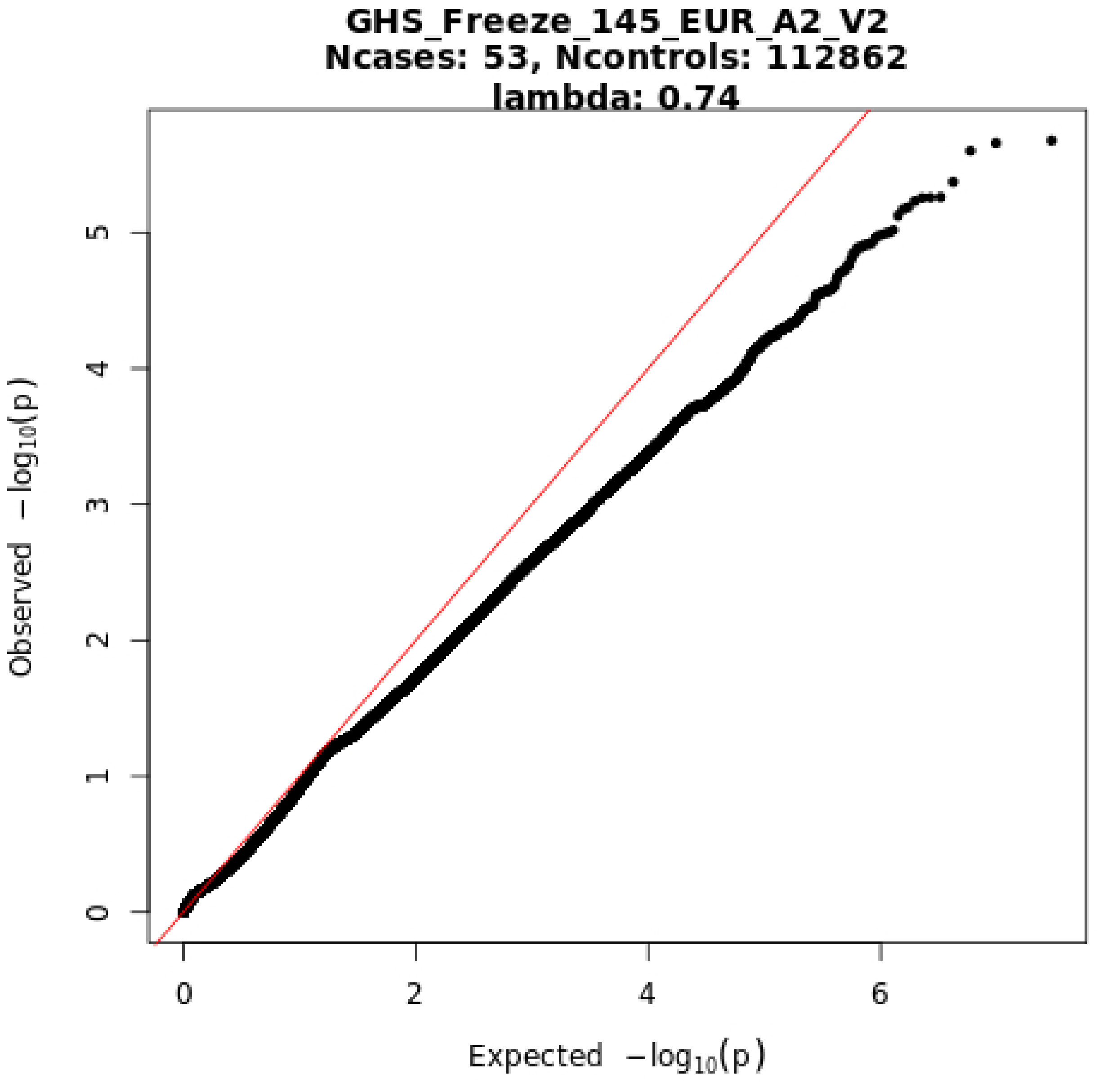
GHS_Freeze_145_EUR_A2_V2
Ncases: 53, Ncontrols: 112862
lambda: 0.74
Observed −log10(p)
Expected −log10(p)
0
1
2
3
4
5
0
2
4
6

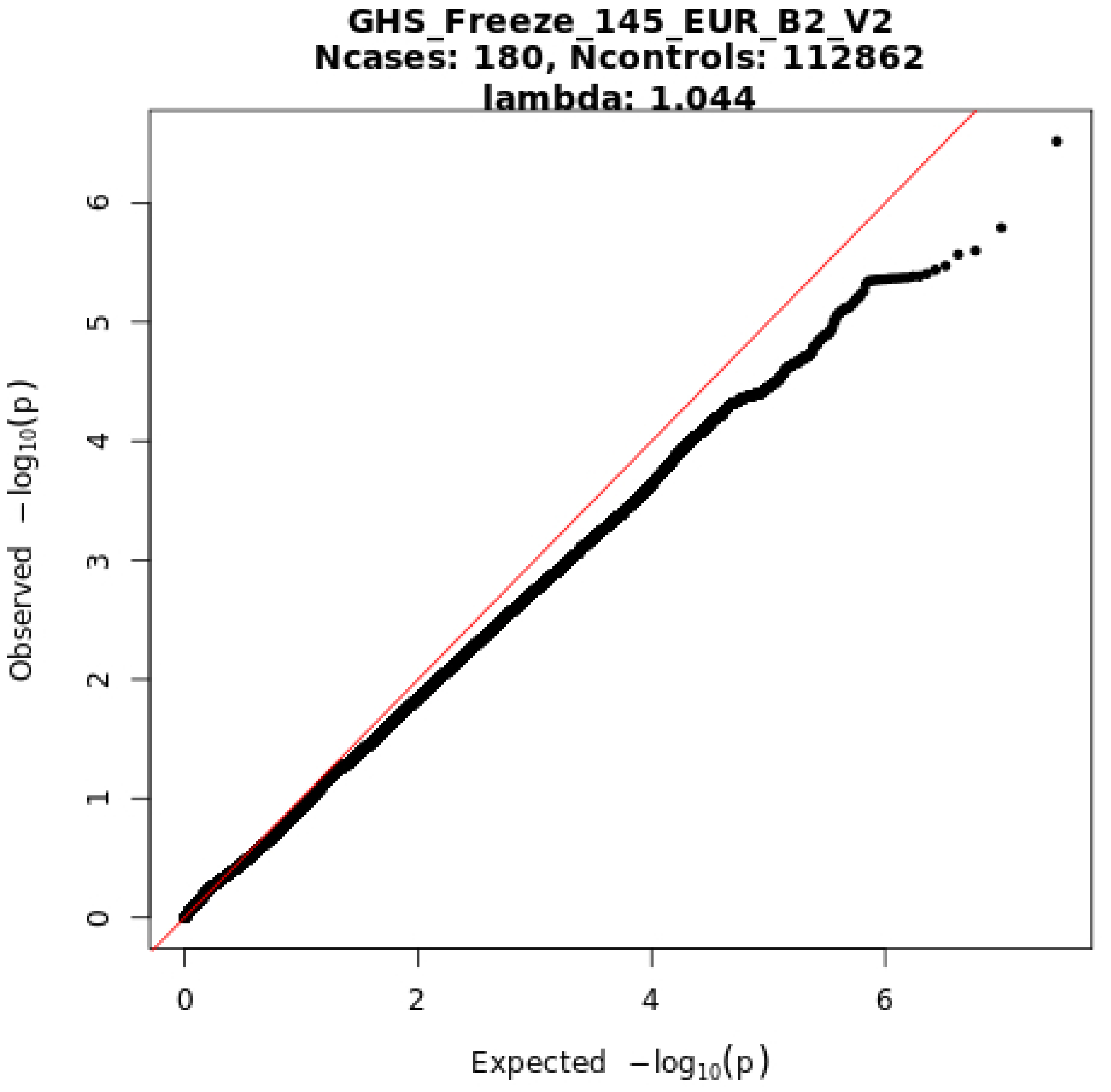
GHS_Freeze_145_EUR_B2_V2
Ncases: 180, Ncontrols: 112862
lambda: 1.044
Observed −log10(p)
Expected −log10(p)
0
1
2
3
4
5
6
0
2
4
6

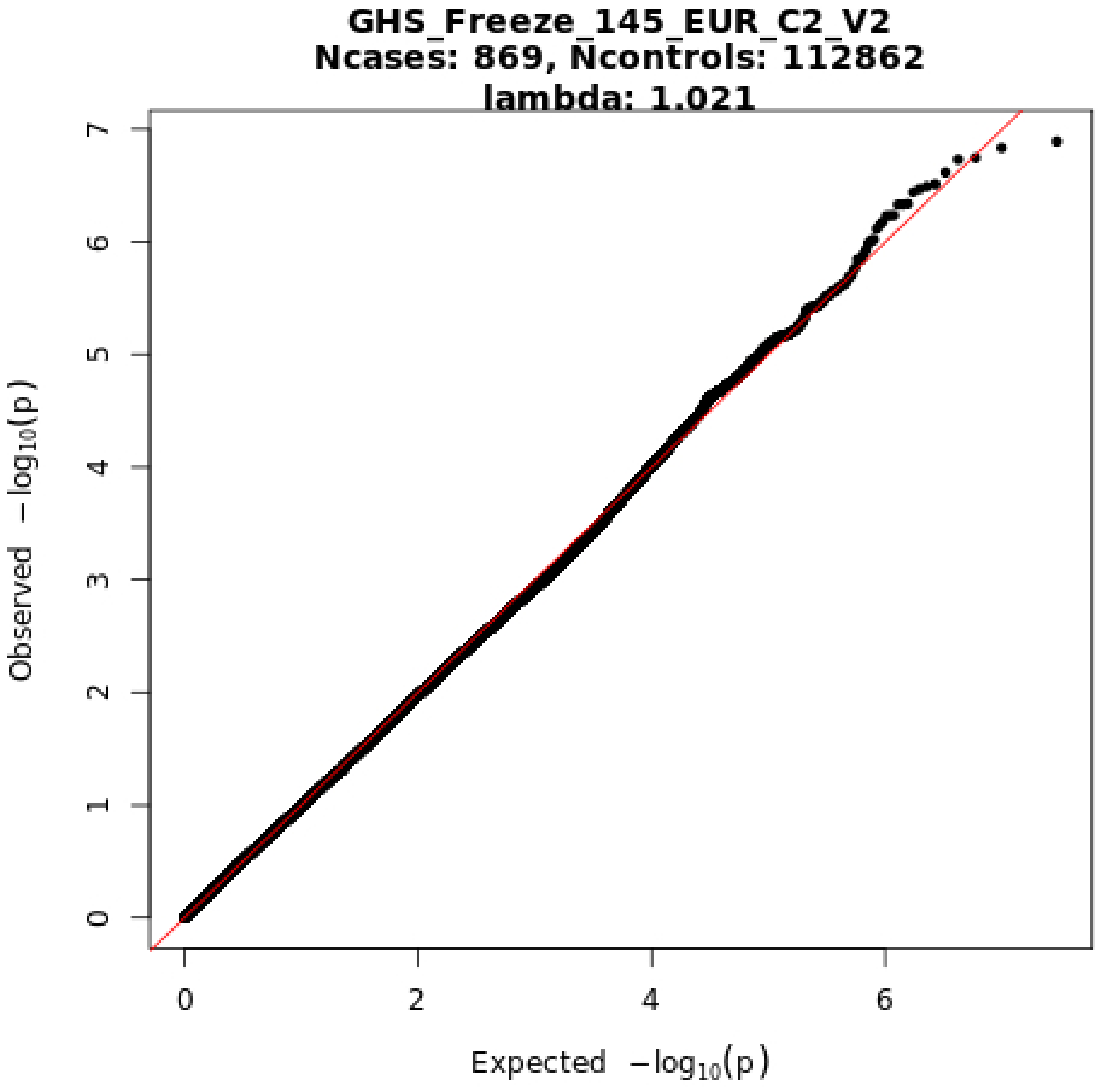
GHS_Freeze_145_EUR_C2_V2
Ncases: 869, Ncontrols: 112862
lambda: 1.021
Observed −log10(p)
Expected −log10(p)
0
1
2
3
4
5
6
7
0
2
4
6

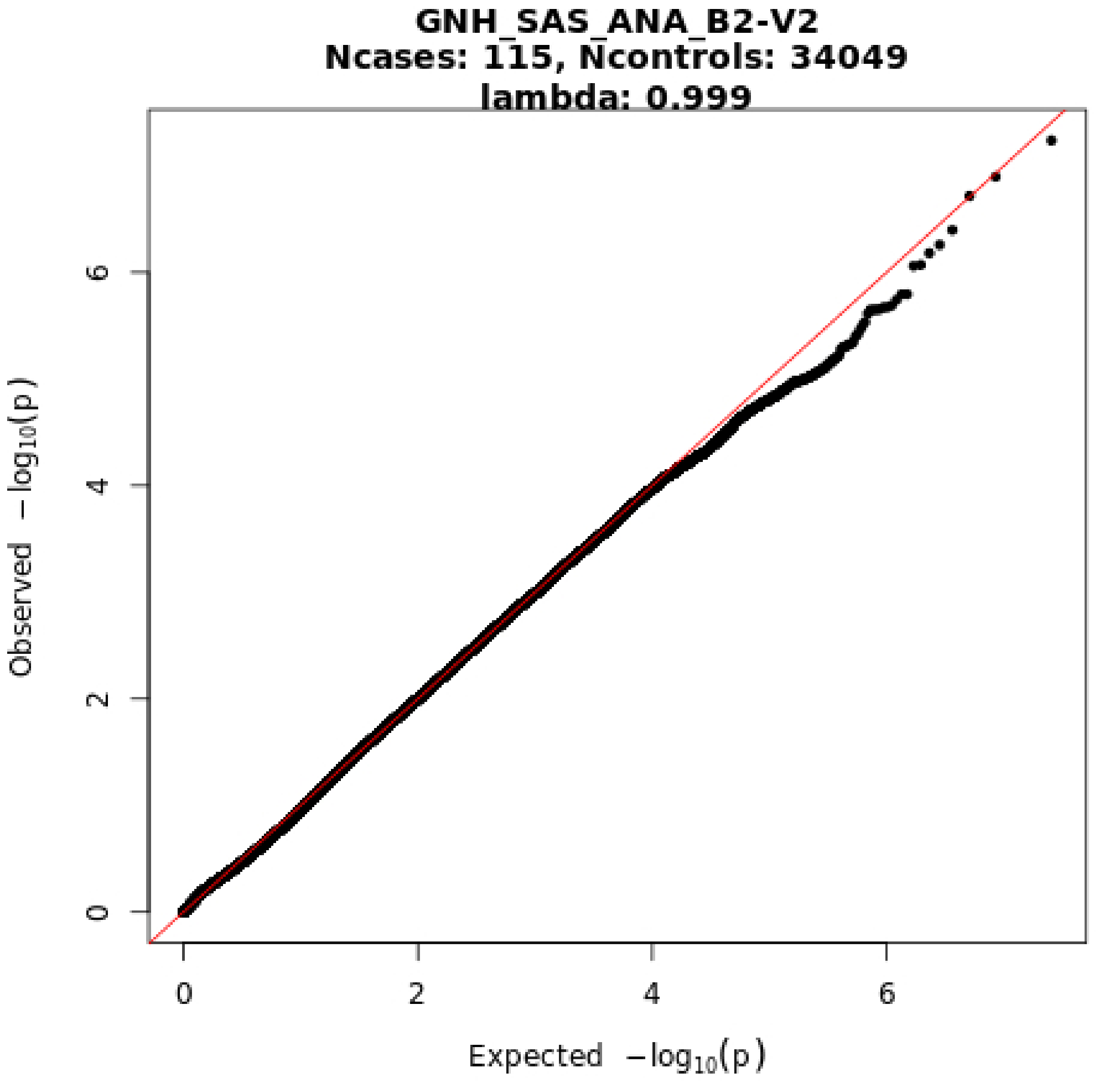
GNH_SAS_ANA_B2-V2
Ncases: 115, Ncontrols: 34049
lambda: 0.999
Observed −log10(p)
Expected −log10(p)
0
2
4
6
0
2
4
6

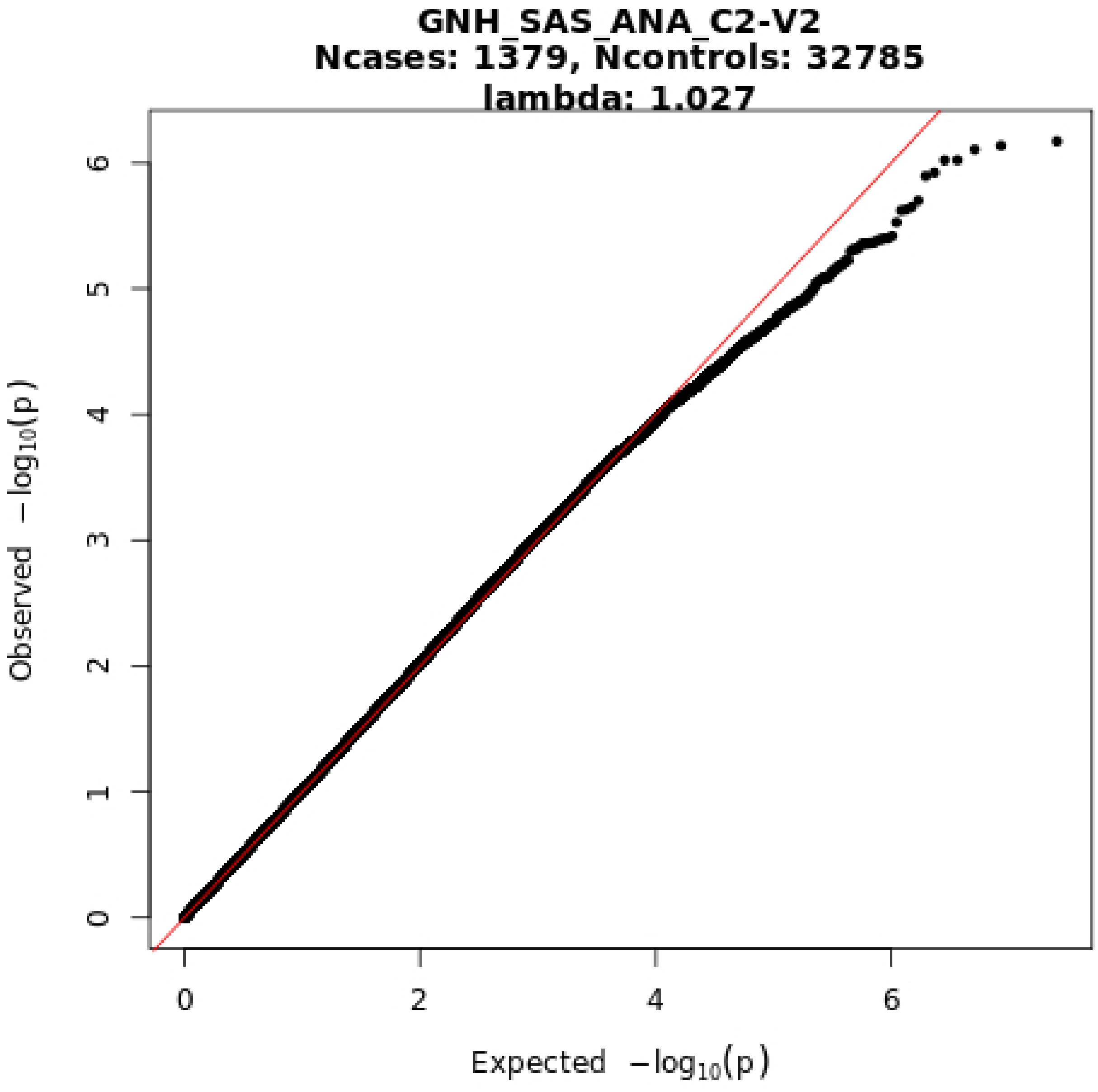
GNH_SAS_ANA_C2-V2
Ncases: 1379, Ncontrols: 32785
lambda: 1.027
Observed −log10(p)
Expected −log10(p)
0
1
2
3
4
5
6
0
2
4
6

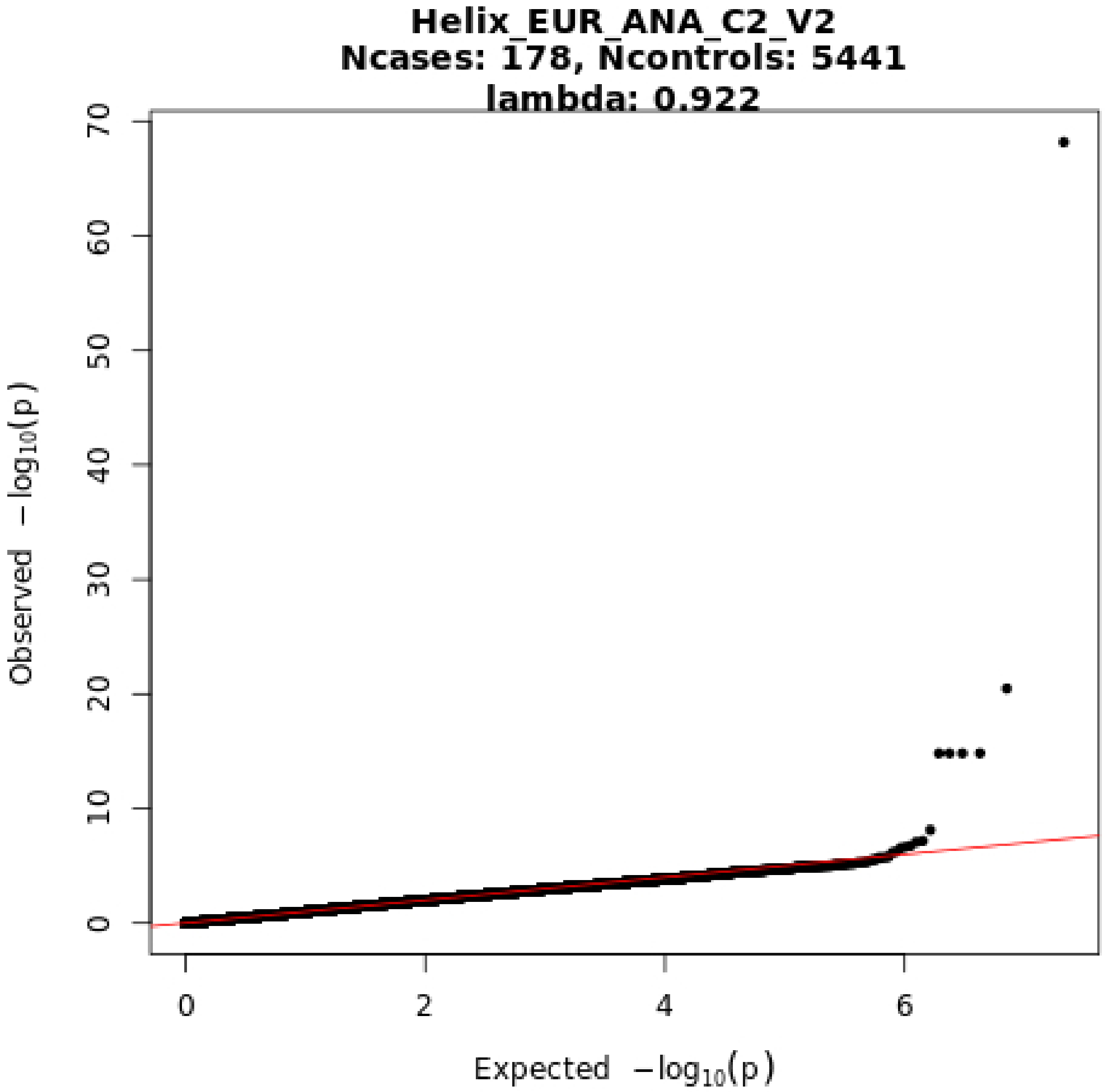
Helix_EUR_ANA_C2_V2
Ncases: 178, Ncontrols: 5441
lambda: 0.922
0
10
20
30
40
50
60
70
Observed $-\log_{10}(p)$
0
2
4
6
Expected $-\log_{10}(p)$
