## Extended data and supplement for "Mapping the human genetic architecture of COVID-19 by worldwide meta-analysis": Extended_Data_Figure_8.pdf

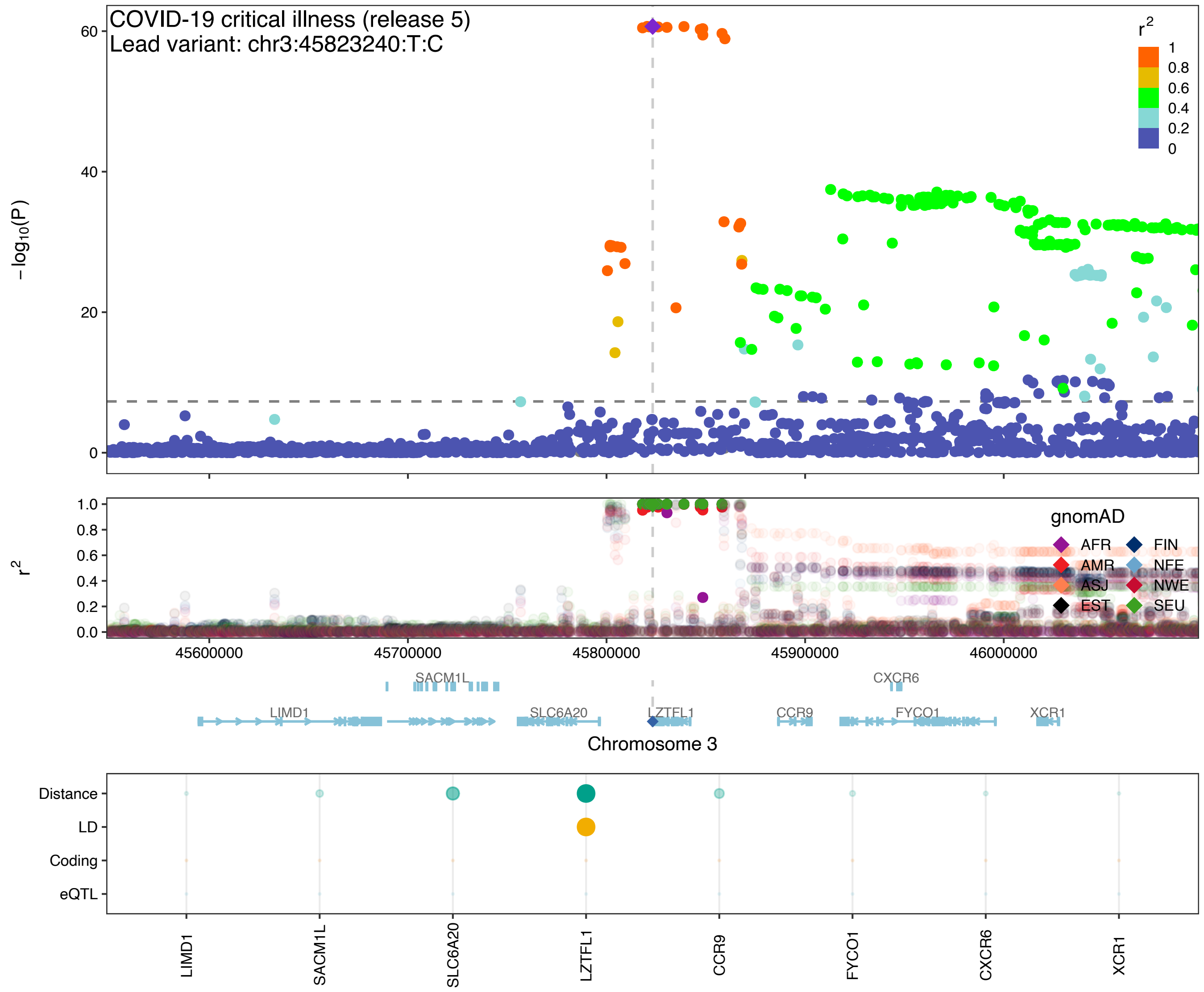

COVID-19 critical illness (release 5)
Lead variant: chr3:45823240:T:C
−log10(P)
r2
1
0.8
0.6
0.4
0.2
0
gnomAD
AFR
FIN
AMR
NFE
ASJ
NWE
EST
SEU
45600000
45700000
45800000
45900000
46000000
SACM1L
CXCR6
LIMD1
SLC6A20
LZTFL1
CCR9
FYCO1
XCR1
Chromosome 3
Distance
LD
Coding
eQTL
LIMD1
SACM1L
SLC6A20
LZTFL1
CCR9
FYCO1
CXCR6
XCR1

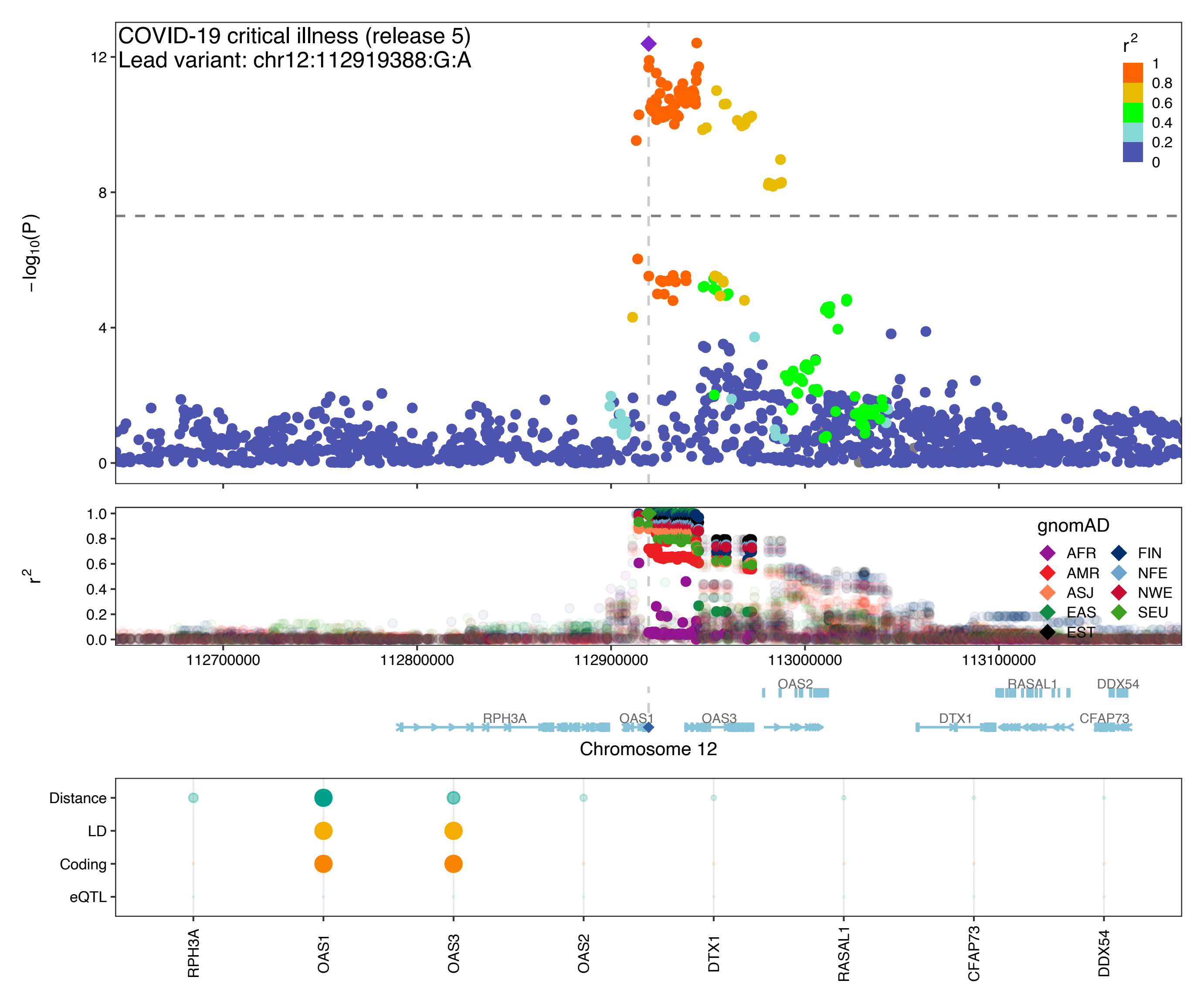

COVID-19 critical illness (release 5)
Lead variant: chr12:112919388:G:A
−log10(P)
r2
1
0.8
0.6
0.4
0.2
0
gnomAD
AFR
AMR
ASJ
EAS
EST
FIN
NFE
NWE
SEU
112700000
112800000
112900000
113000000
113100000
OAS2
RASAL1
DDX54
RPH3A
OAS1
OAS3
DTX1
CFAP73
Chromosome 12
Distance
LD
Coding
eQTL
RPH3A
OAS1
OAS3
OAS2
DTX1
RASAL1
CFAP73
DDX54

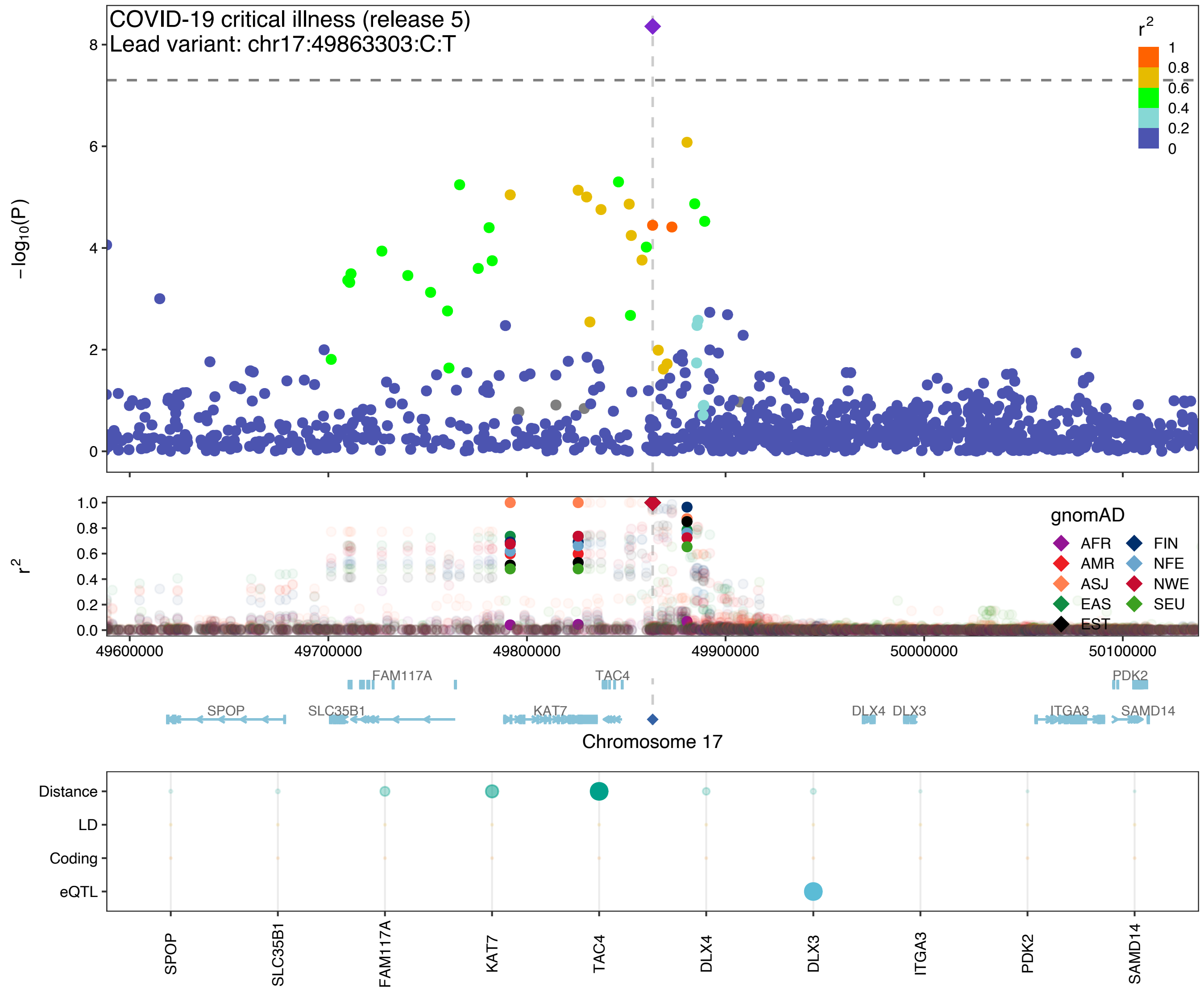
COVID-19 critical illness (release 5)
Lead variant: chr17:49863303:C:T
−log10(P)
r2
1
0.8
0.6
0.4
0.2
0
gnomAD
AFR
FIN
AMR
NFE
ASJ
NWE
EAS
SEU
EST
49600000
49700000
49800000
49900000
50000000
50100000
FAM117A
TAC4
PDK2
SPOP
SLC35B1
KAT7
DLX4
DLX3
ITGA3
SAMD14
Chromosome 17
Distance
LD
Coding
eQTL
SPOP
SLC35B1
FAM117A
KAT7
TAC4
DLX4
DLX3
ITGA3
PDK2
SAMD14

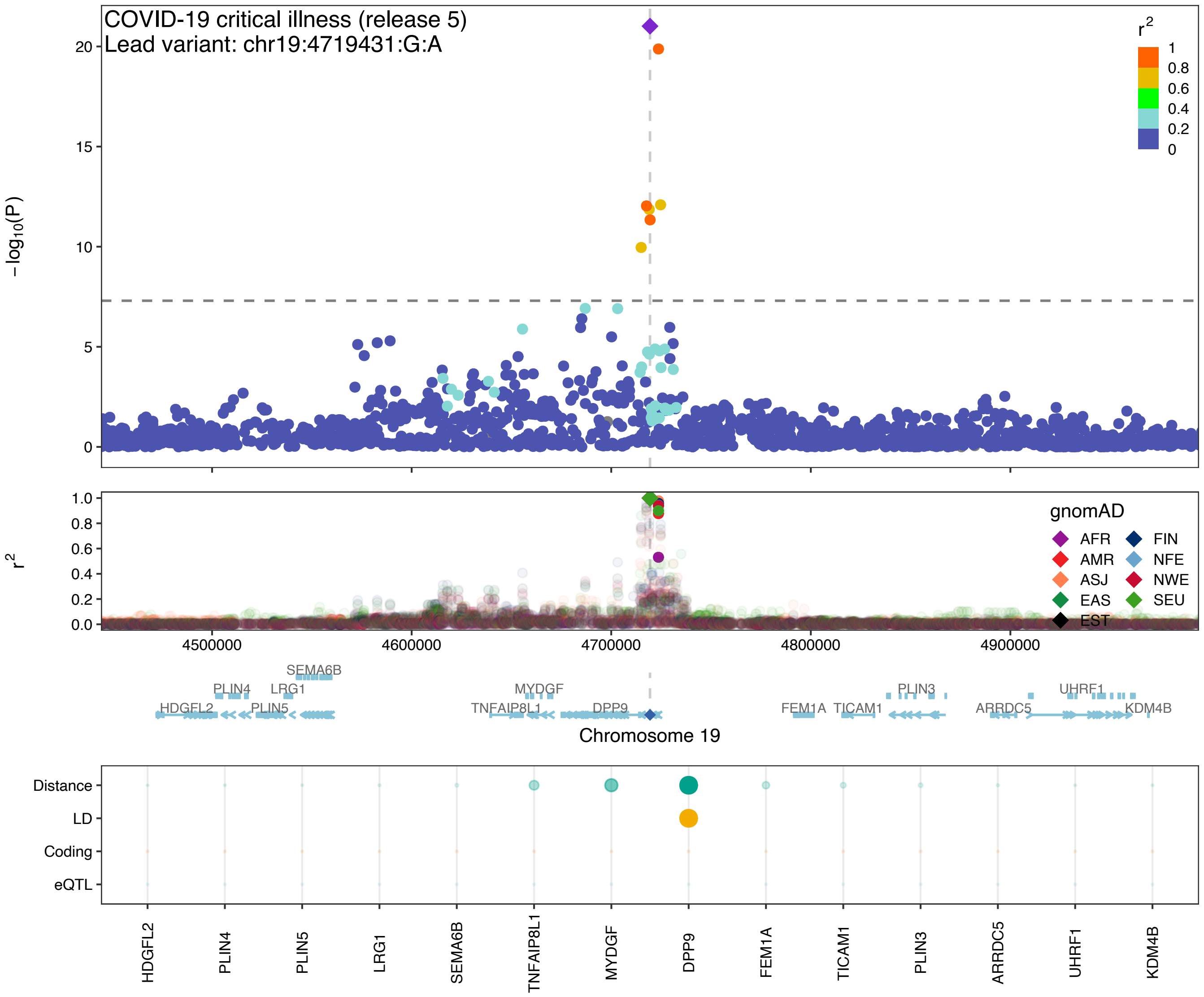

COVID-19 critical illness (release 5)
Lead variant: chr19:4719431:G:A
−log10(P)
r2
gnomAD
AFR
AMR
ASJ
EAS
EST
FIN
NFE
NWE
SEU
4500000
4600000
4700000
4800000
4900000
SEMA6B
PLIN4
LRG1
HDGFL2
PLIN5
MYDGF
TNFAIP8L1
DPP9
FEM1A
TICAM1
PLIN3
ARRDC5
UHRF1
KDM4B
Chromosome 19
Distance
LD
Coding
eQTL

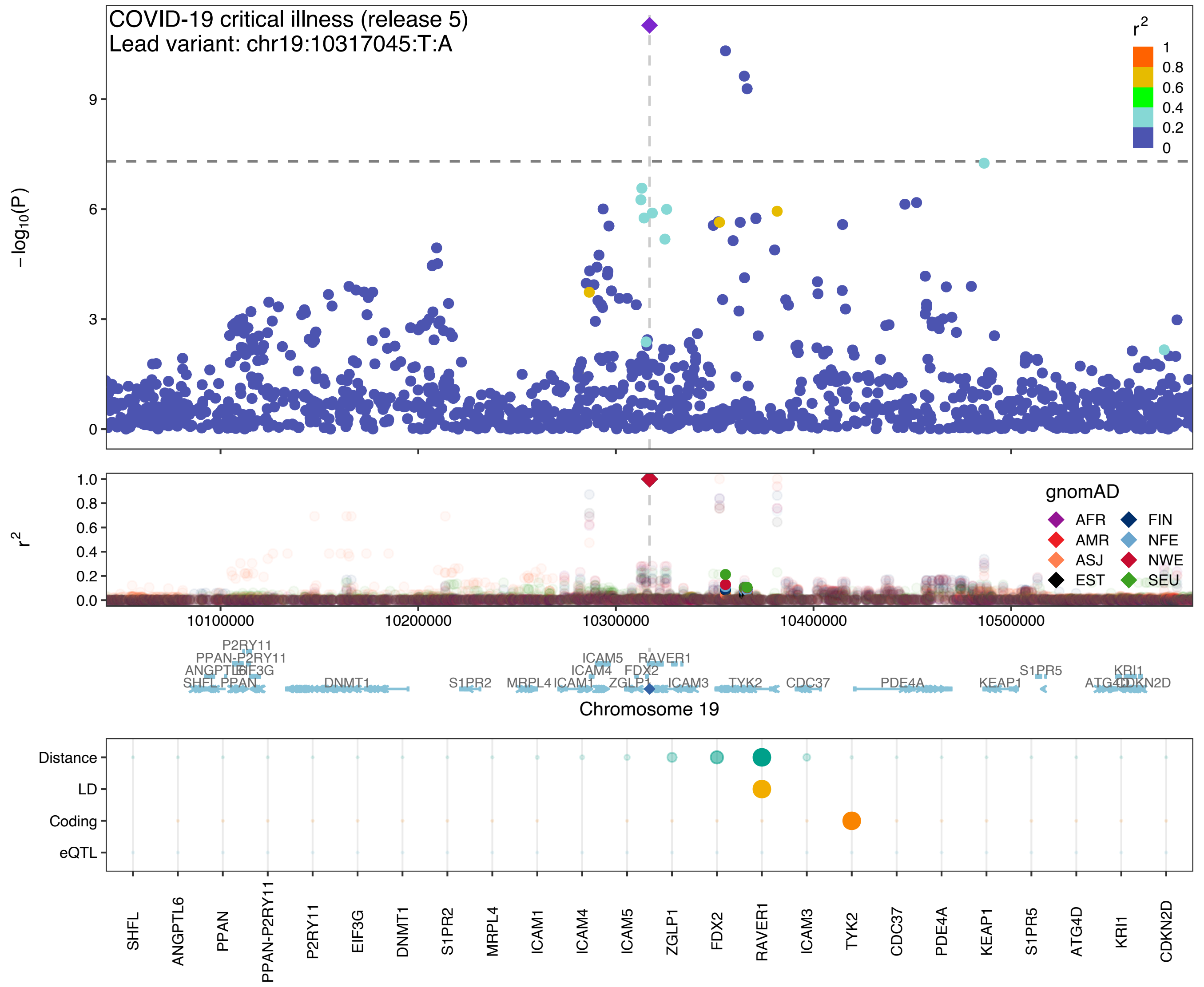

COVID-19 critical illness (release 5)
Lead variant: chr19:10317045:T:A
$-\log_{10}(P)$
$r^2$
1
0.8
0.6
0.4
0.2
0
gnomAD
AFR
AMR
ASJ
EST
FIN
NFE
NWE
SEU
10100000
10200000
10300000
10400000
10500000
Chromosome 19
P2RY11
PPAN-P2RY11
ANGPTL6
EIF3G
SHFL
PPAN
DNMT1
S1PR2
MRPL4
ICAM1
ICAM5
ICAM4
FDX2
ZGLP1
RAVER1
ICAM3
TYK2
CDC37
PDE4A
KEAP1
S1PR5
KRI1
ATG4D
CDKN2D
Distance
LD
Coding
eQTL
SHFL
ANGPTL6
PPAN
PPAN-P2RY11
P2RY11
EIF3G
DNMT1
S1PR2
MRPL4
ICAM1
ICAM4
ICAM5
ZGLP1
FDX2
RAVER1
ICAM3
TYK2
CDC37
PDE4A
KEAP1
S1PR5
ATG4D
KRI1
CDKN2D

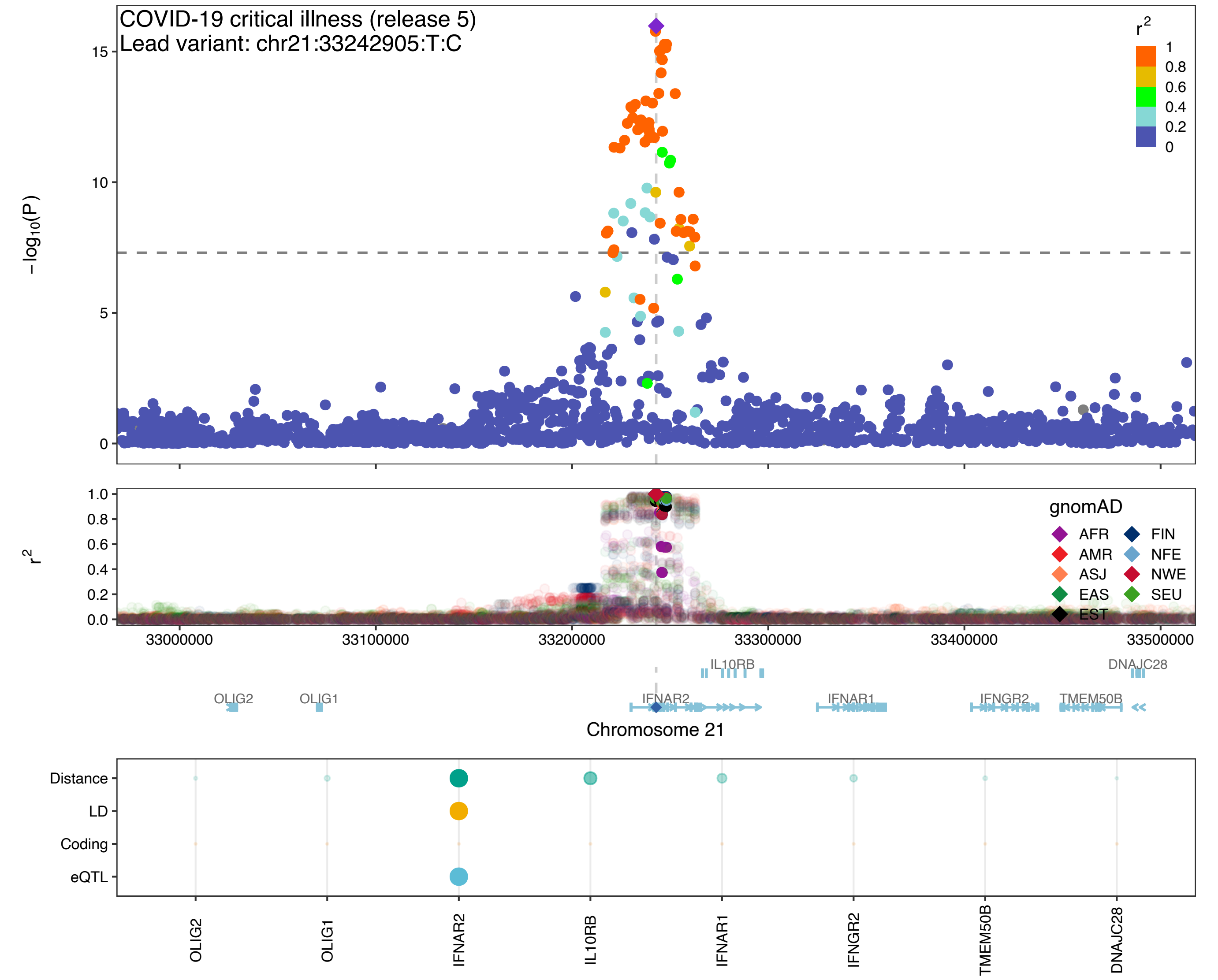
COVID-19 critical illness (release 5)
Lead variant: chr21:33242905:T:C
r²
1
0.8
0.6
0.4
0.2
0
−log10(P)
15
10
5
0
r²
1.0
0.8
0.6
0.4
0.2
0.0
gnomAD
AFR
AMR
ASJ
EAS
EST
FIN
NFE
NWE
SEU
33000000
33100000
33200000
33300000
33400000
33500000
IL10RB
DNAJC28
OLIG2
OLIG1
IFNAR2
IFNAR1
IFNGR2
TMEM50B
Chromosome 21
Distance
LD
Coding
eQTL
OLIG2
OLIG1
IFNAR2
IL10RB
IFNAR1
IFNGR2
TMEM50B
DNAJC28

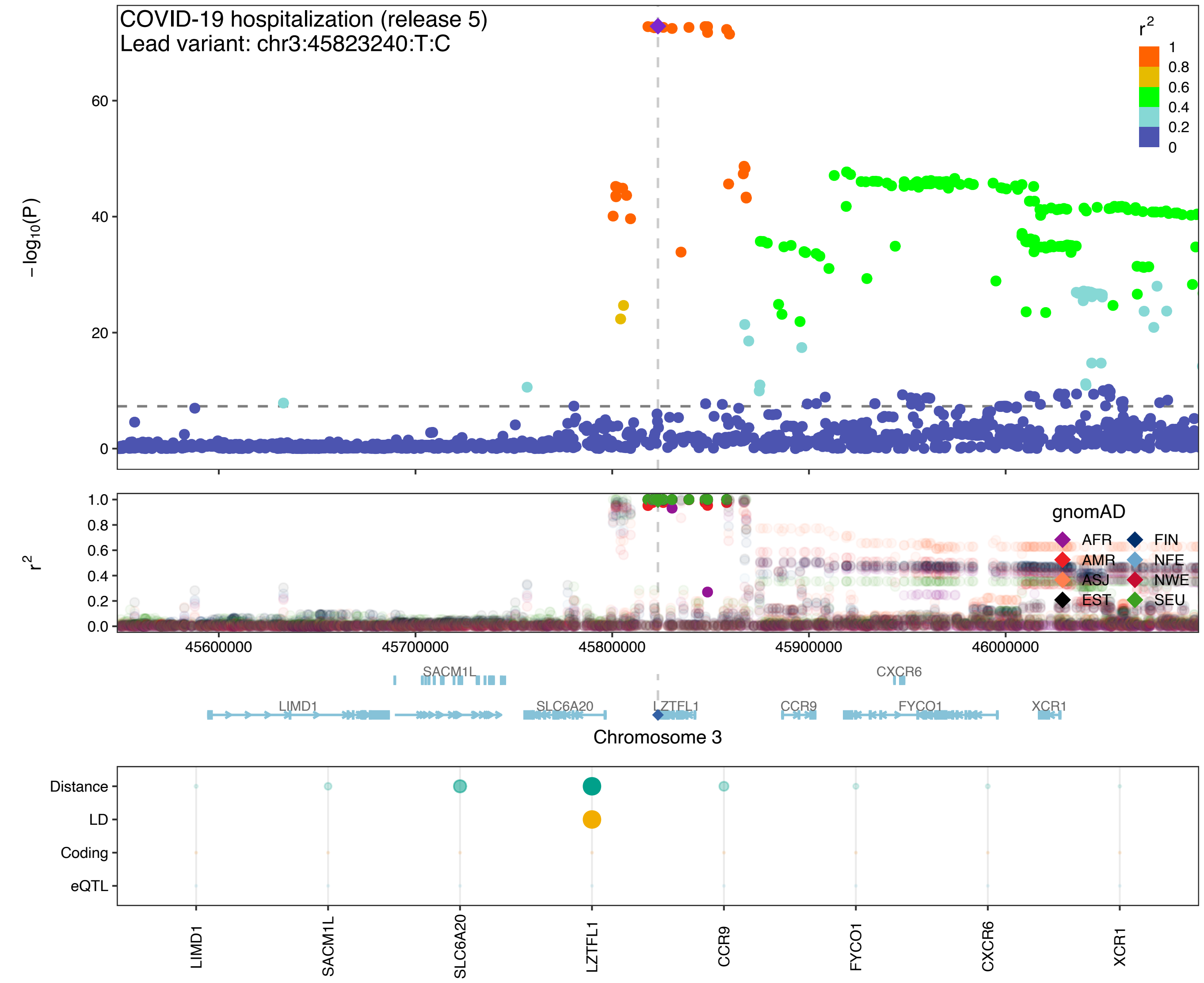
COVID-19 hospitalization (release 5)
Lead variant: chr3:45823240:T:C
r2
1
0.8
0.6
0.4
0.2
0
60
40
20
0
−log10(P)
1.0
0.8
0.6
0.4
0.2
0.0
r2
gnomAD
AFR
AMR
ASJ
EST
FIN
NFE
NWE
SEU
45600000
45700000
45800000
45900000
46000000
SACM1L
CXCR6
LIMD1
SLC6A20
LZTFL1
CCR9
FYCO1
XCR1
Chromosome 3
Distance
LD
Coding
eQTL
LIMD1
SACM1L
SLC6A20
LZTFL1
CCR9
FYCO1
CXCR6
XCR1

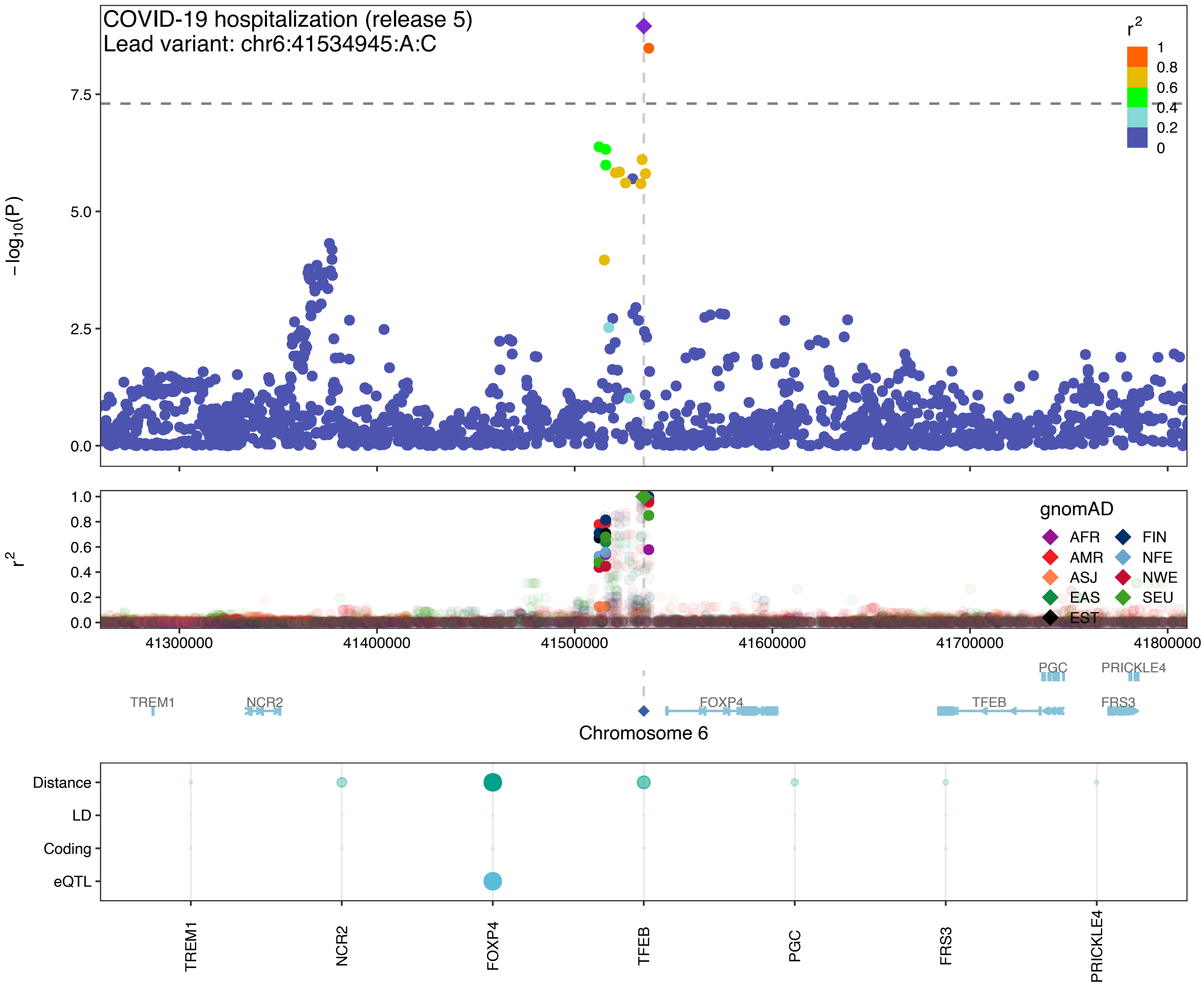

COVID-19 hospitalization (release 5)
Lead variant: chr6:41534945:A:C
$-\log_{10}(P)$
0.0
2.5
5.0
7.5
$r^2$
1
0.8
0.6
0.4
0.2
0
1.0
gnomAD
AFR
AMR
ASJ
EAS
EST
FIN
NFE
NWE
SEU
41300000
41400000
41500000
41600000
41700000
41800000
TREM1
NCR2
FOXP4
TFEB
PGC
PRICKLE4
FRS3
Chromosome 6
Distance
LD
Coding
eQTL

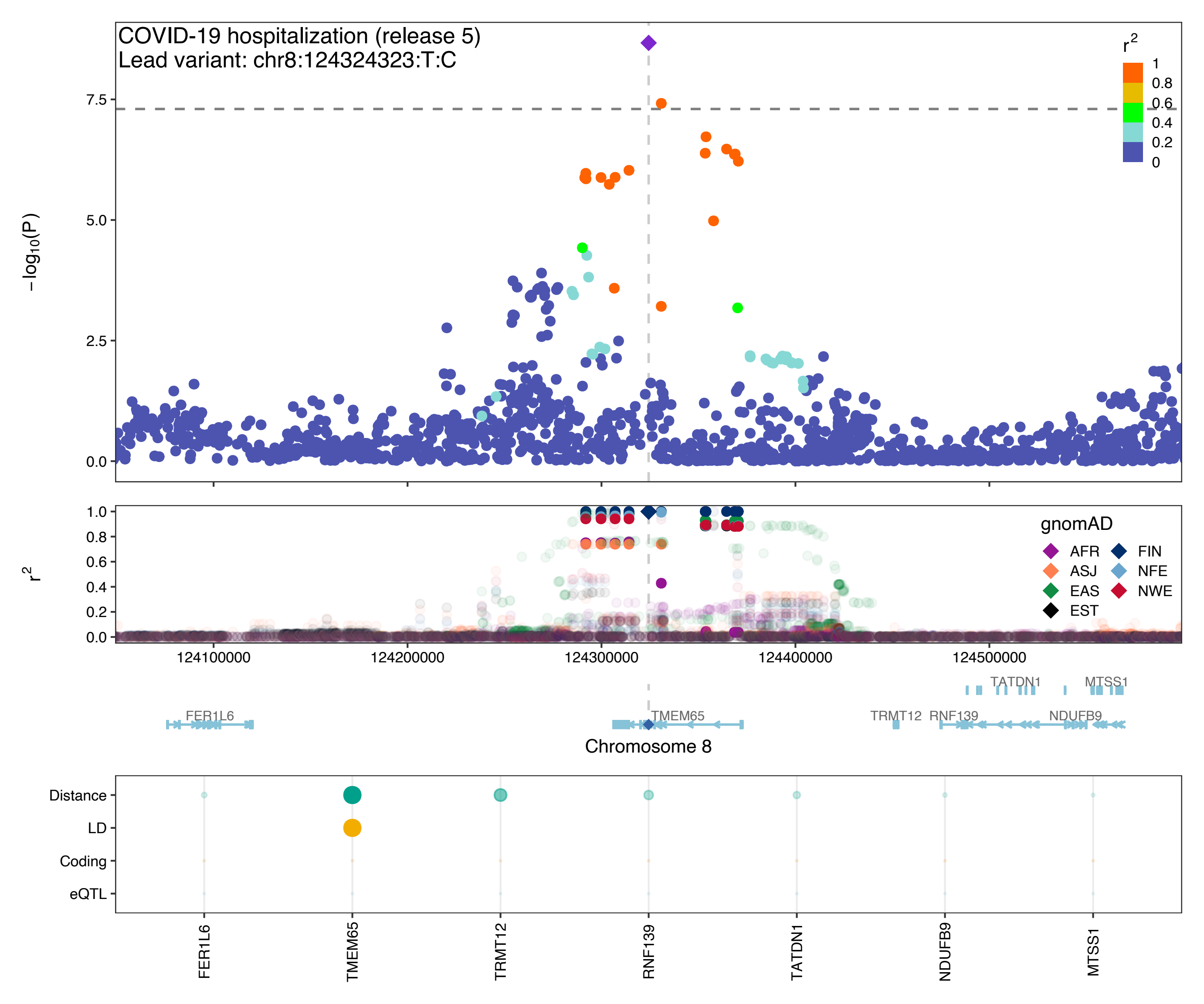

COVID-19 hospitalization (release 5)
Lead variant: chr8:124324323:T:C
r2
1
0.8
0.6
0.4
0.2
0
−log10(P)
7.5
5.0
2.5
0.0
r2
1.0
0.8
0.6
0.4
0.2
0.0
gnomAD
AFR
FIN
ASJ
NFE
EAS
NWE
EST
124100000
124200000
124300000
124400000
124500000
TATDN1
MTSS1
FER1L6
TMEM65
TRMT12
RNF139
NDUFB9
Chromosome 8
Distance
LD
Coding
eQTL
FER1L6
TMEM65
TRMT12
RNF139
TATDN1
NDUFB9
MTSS1

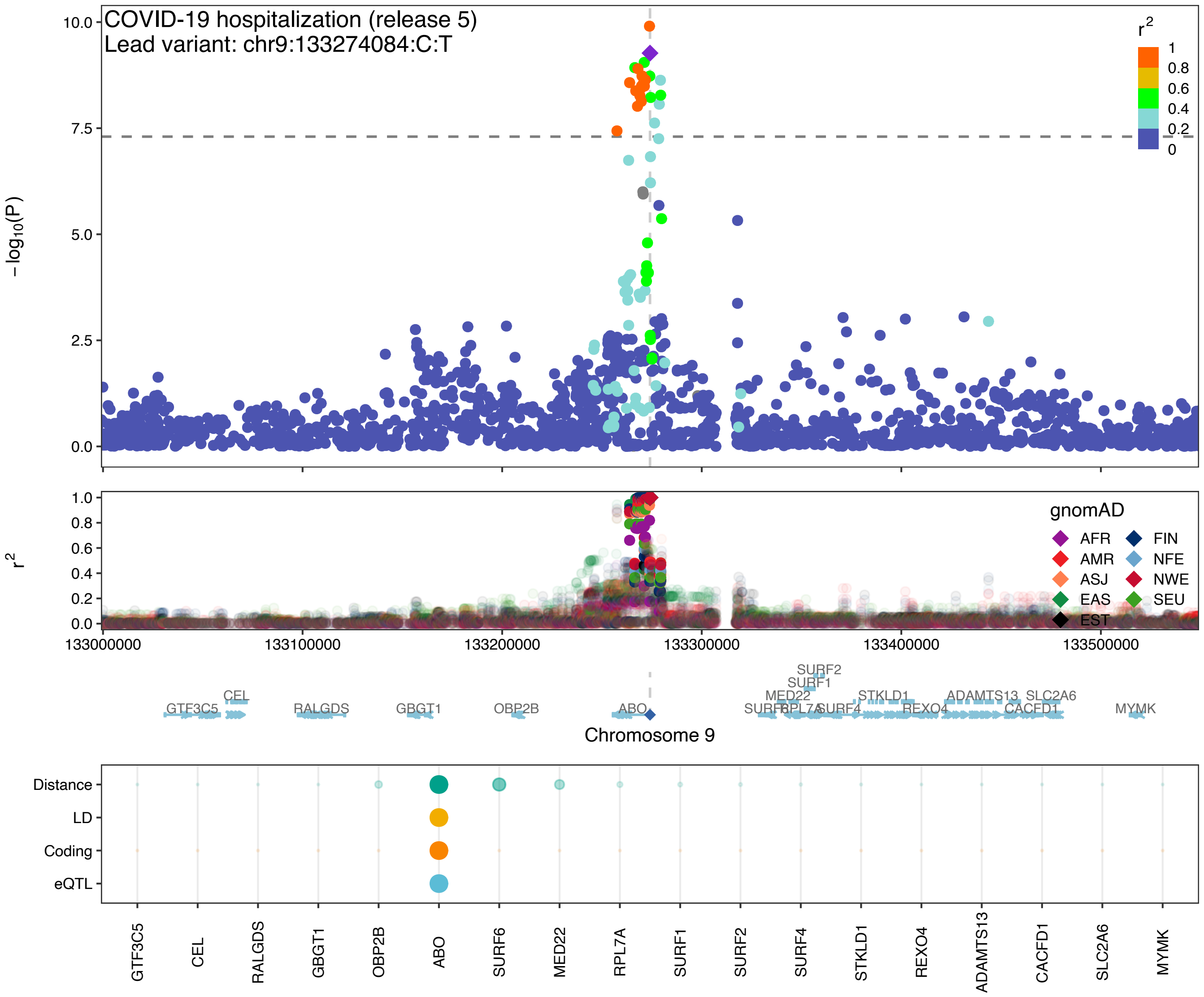
COVID-19 hospitalization (release 5)
Lead variant: chr9:133274084:C:T
−log10(P)
r2
gnomAD
AFR
AMR
ASJ
EAS
EST
FIN
NFE
NWE
SEU
133000000
133100000
133200000
133300000
133400000
133500000
Chromosome 9
Distance
LD
Coding
eQTL
GTF3C5
CEL
RALGDS
GBGT1
OBP2B
ABO
SURF6
MED22
RPL7A
SURF1
SURF2
SURF4
STKLD1
REXO4
ADAMTS13
CACFD1
SLC2A6
MYMK

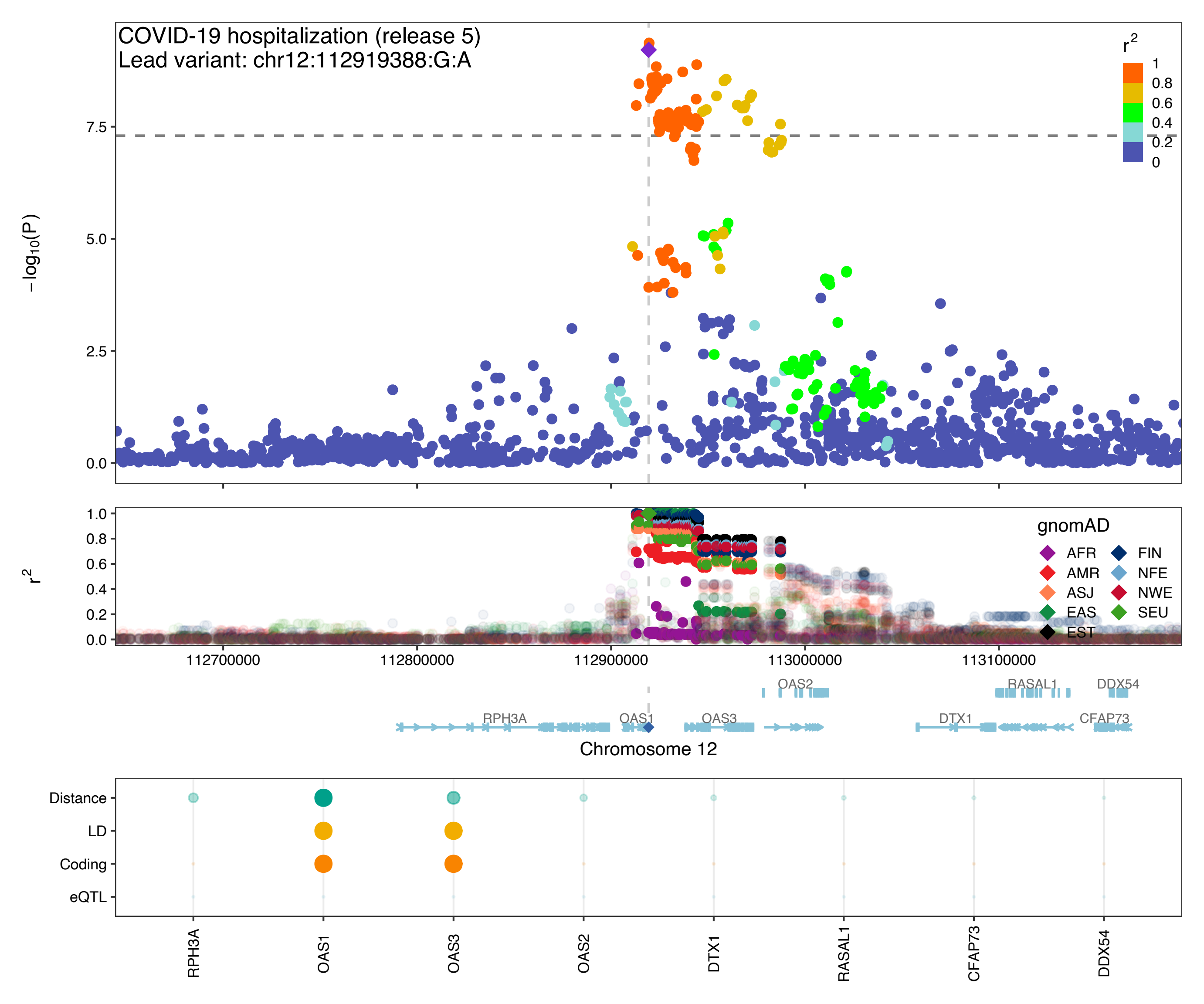
COVID-19 hospitalization (release 5)
Lead variant: chr12:112919388:G:A
$-\log_{10}(P)$
$r^2$
gnomAD
AFR
AMR
ASJ
EAS
EST
FIN
NFE
NWE
SEU
112700000
112800000
112900000
113000000
113100000
OAS2
RASAL1
DDX54
RPH3A
OAS1
OAS3
DTX1
CFAP73
Chromosome 12
Distance
LD
Coding
eQTL
RPH3A
OAS1
OAS3
OAS2
DTX1
RASAL1
CFAP73
DDX54

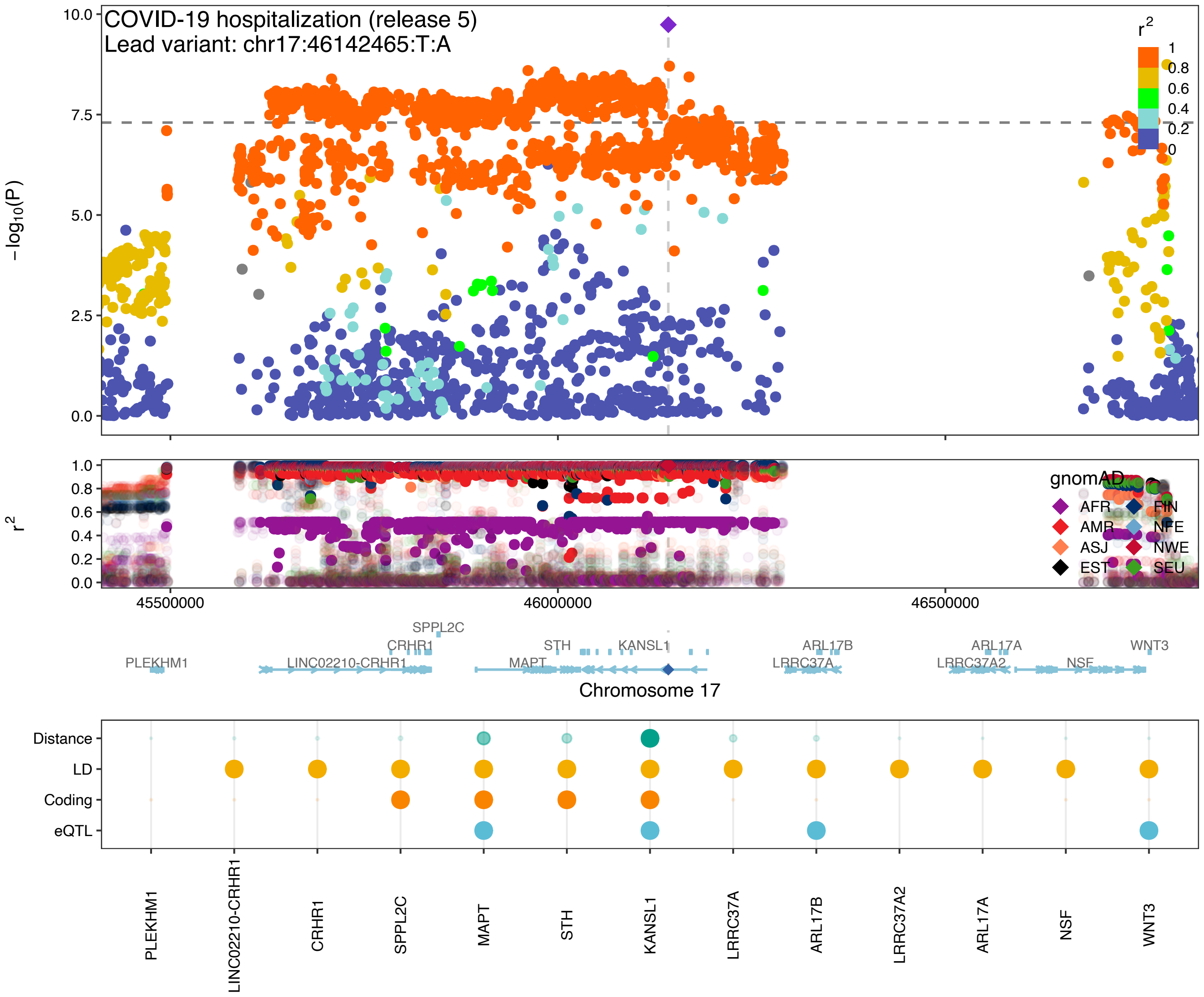

COVID-19 hospitalization (release 5)
Lead variant: chr17:46142465:T:A
−log10(P)
r2
gnomAD
AFR
AMR
ASJ
EST
FIN
NFE
NWE
SEU
45500000
46000000
46500000
SPPL2C
CRHR1
STH
KANSL1
ARL17B
ARL17A
WNT3
PLEKHM1
LINC02210-CRHR1
MAPT
LRRC37A
LRRC37A2
NSF
Chromosome 17
Distance
LD
Coding
eQTL
PLEKHM1
LINC02210-CRHR1
CRHR1
SPPL2C
MAPT
STH
KANSL1
LRRC37A
ARL17B
LRRC37A2
ARL17A
NSF
WNT3

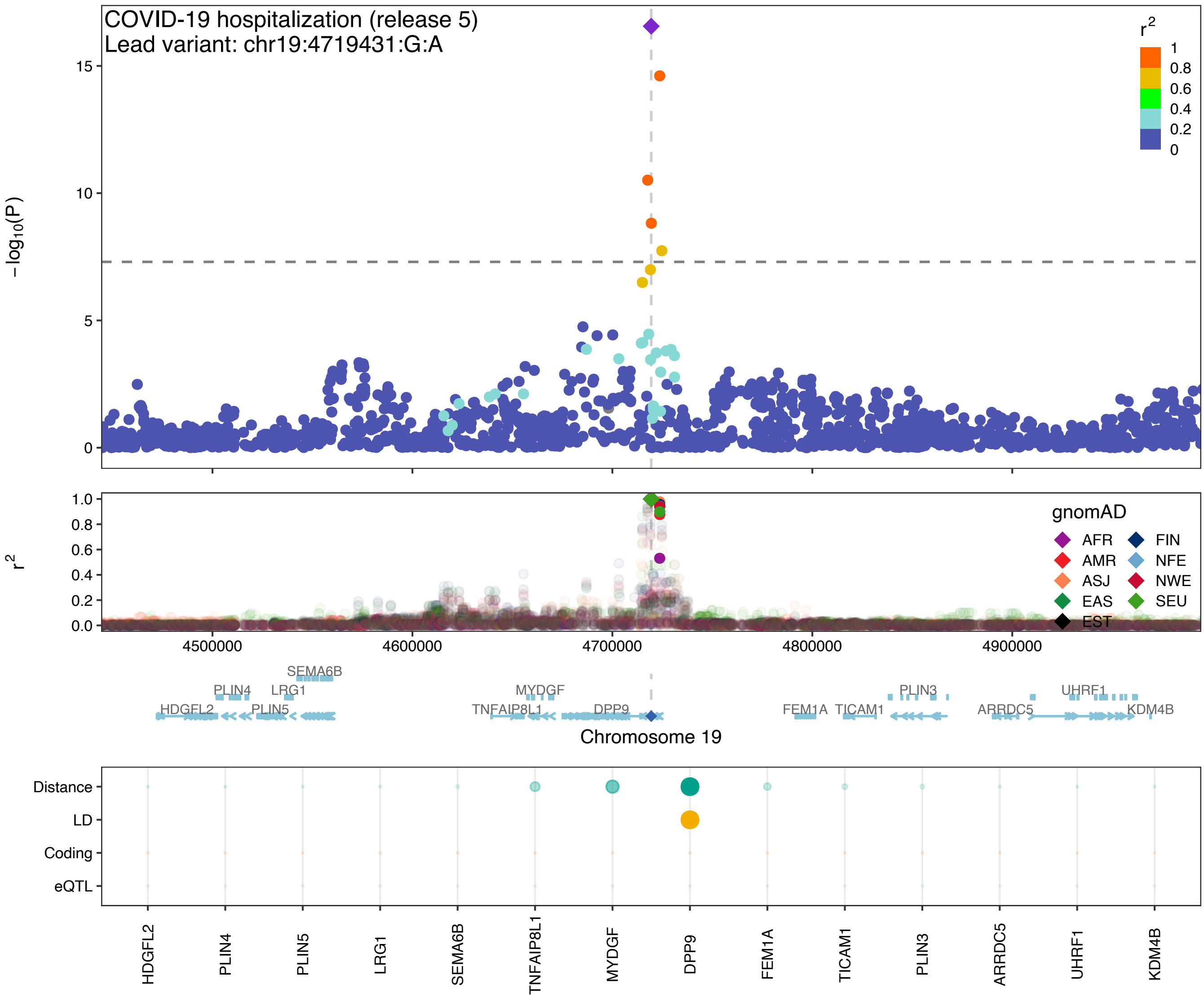

COVID-19 hospitalization (release 5)
Lead variant: chr19:4719431:G:A
−log10(P)
r2
gnomAD
AFR
AMR
ASJ
EAS
EST
FIN
NFE
NWE
SEU
4500000
4600000
4700000
4800000
4900000
SEMA6B
PLIN4
LRG1
HDGFL2
PLIN5
MYDGF
TNFAIP8L1
DPP9
FEM1A
TICAM1
PLIN3
ARRDC5
UHRF1
KDM4B
Chromosome 19
Distance
LD
Coding
eQTL
HDGFL2
PLIN4
PLIN5
LRG1
SEMA6B
TNFAIP8L1
MYDGF
DPP9
FEM1A
TICAM1
PLIN3
ARRDC5
UHRF1
KDM4B

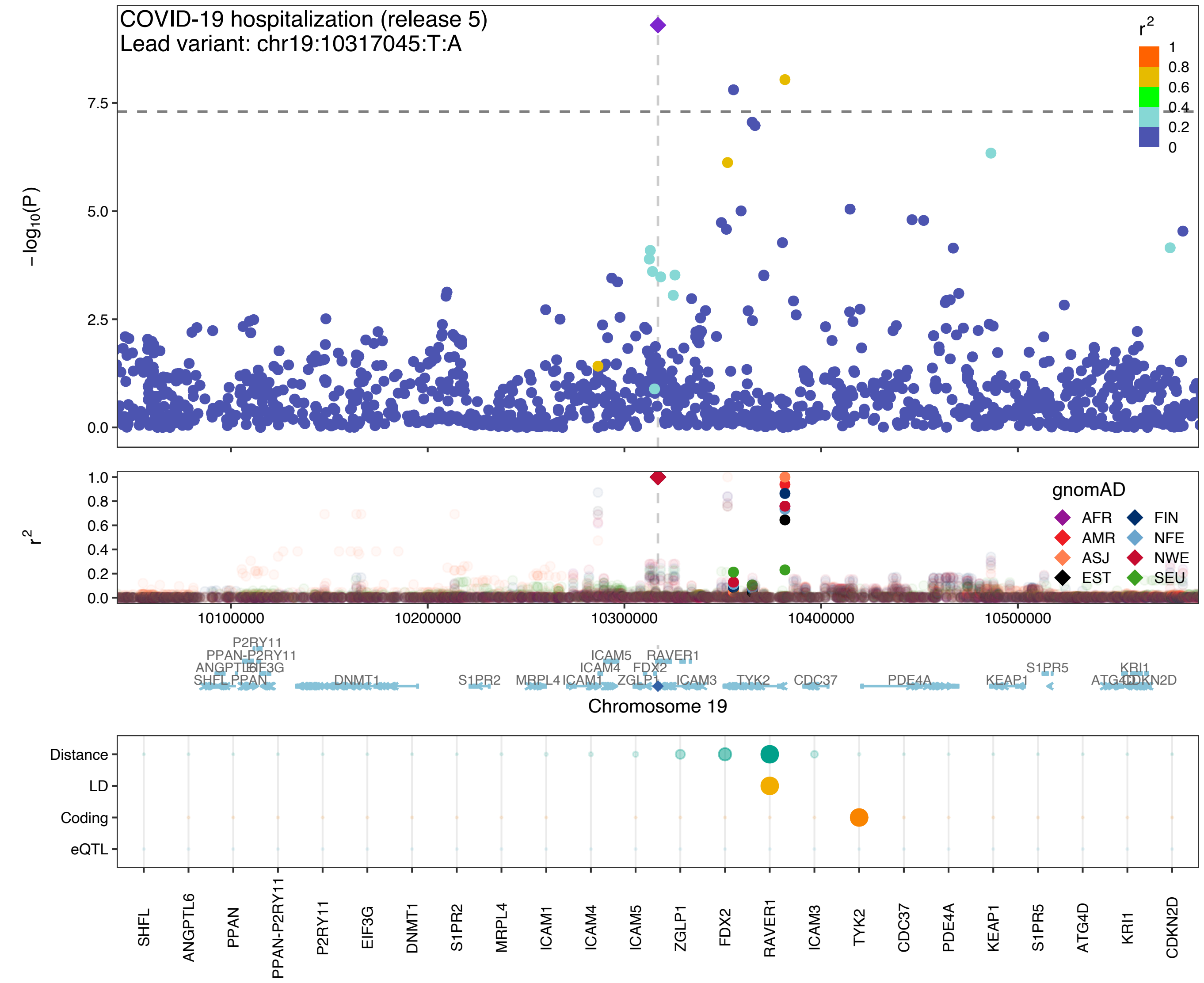
COVID-19 hospitalization (release 5)
Lead variant: chr19:10317045:T:A
$-\log_{10}(P)$
$r^2$
1
0.8
0.6
0.4
0.2
0
gnomAD
AFR
AMR
ASJ
EST
FIN
NFE
NWE
SEU
10100000
10200000
10300000
10400000
10500000
Chromosome 19
Distance
LD
Coding
eQTL
SHFL
ANGPTL6
PPAN
PPAN-P2RY11
P2RY11
EIF3G
DNMT1
S1PR2
MRPL4
ICAM1
ICAM4
ICAM5
ZGLP1
FDX2
RAVER1
ICAM3
TYK2
CDC37
PDE4A
KEAP1
S1PR5
ATG4D
KRI1
CDKN2D

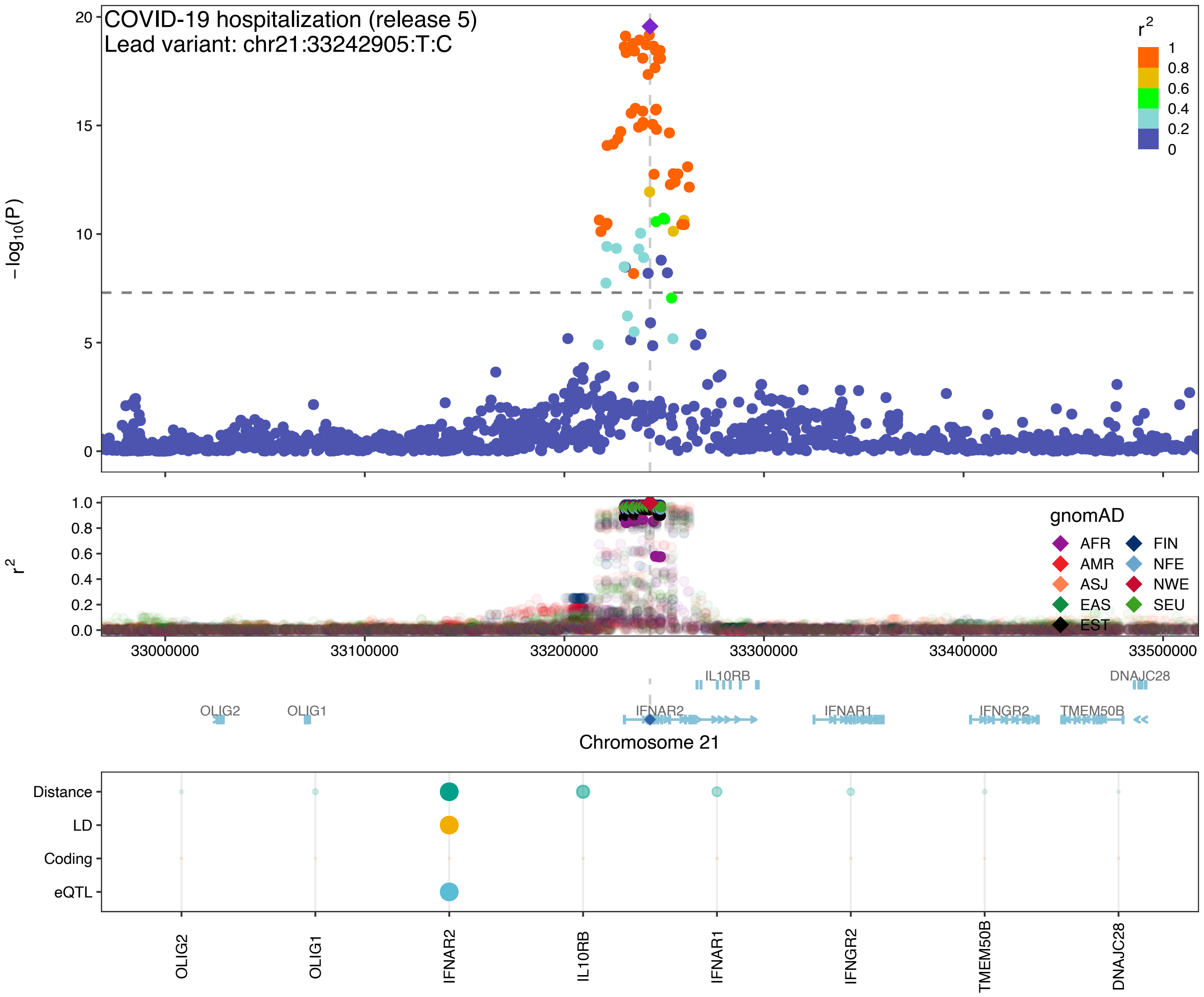

COVID-19 hospitalization (release 5)
Lead variant: chr21:33242905:T:C
r2
1
0.8
0.6
0.4
0.2
0
−log10(P)
20
15
10
5
0
r2
1.0
0.8
0.6
0.4
0.2
0.0
gnomAD
AFR
FIN
AMR
NFE
ASJ
NWE
EAS
SEU
EST
33000000
33100000
33200000
33300000
33400000
33500000
IL10RB
DNAJC28
OLIG2
OLIG1
IFNAR2
IFNAR1
IFNGR2
TMEM50B
Chromosome 21
Distance
LD
Coding
eQTL
OLIG2
OLIG1
IFNAR2
IL10RB
IFNAR1
IFNGR2
TMEM50B
DNAJC28

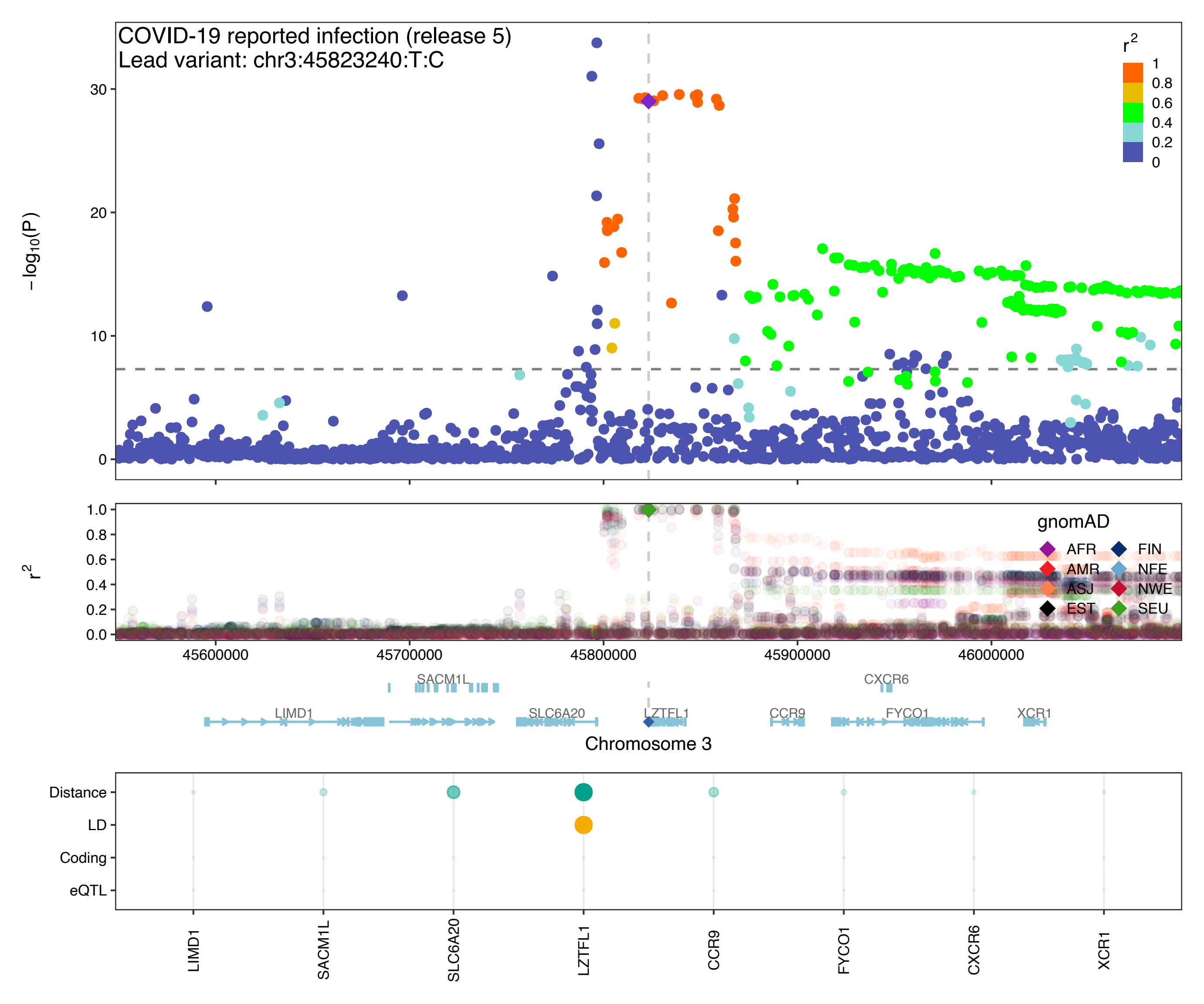
COVID-19 reported infection (release 5)
Lead variant: chr3:45823240:T:C
$r^2$
1
0.8
0.6
0.4
0.2
0
30
20
10
0
$-\log_{10}(P)$
$r^2$
1.0
0.8
0.6
0.4
0.2
0.0
gnomAD
AFR
FIN
AMR
NFE
ASJ
NWE
EST
SEU
45600000
45700000
45800000
45900000
46000000
SACM1L
CXCR6
LIMD1
SLC6A20
LZTFL1
CCR9
FYCO1
XCR1
Chromosome 3
Distance
LD
Coding
eQTL
LIMD1
SACM1L
SLC6A20
LZTFL1
CCR9
FYCO1
CXCR6
XCR1

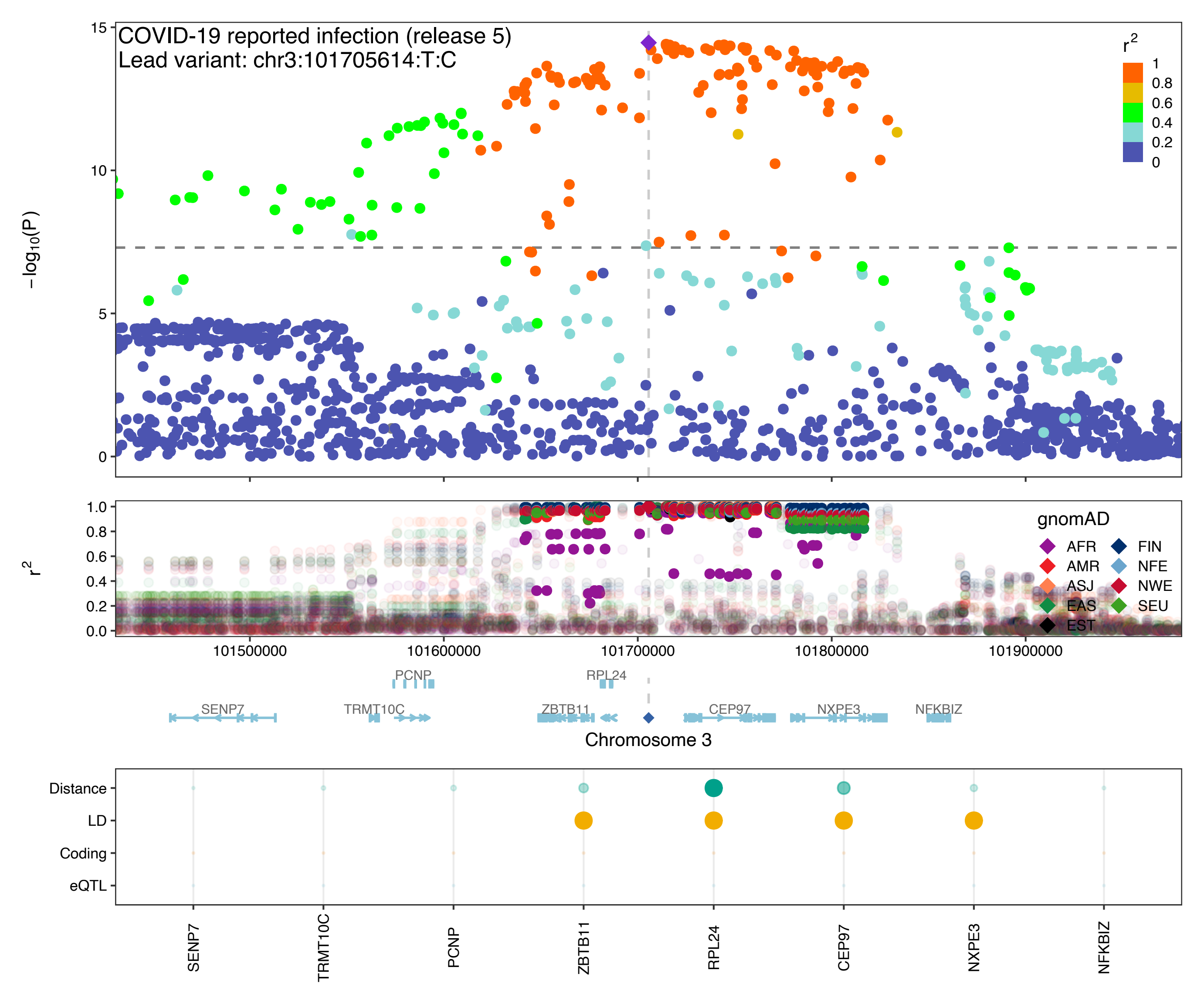

COVID-19 reported infection (release 5)
Lead variant: chr3:101705614:T:C
−log10(P)
15
10
5
0
r2
1
0.8
0.6
0.4
0.2
0
1.0
0.8
0.6
0.4
0.2
0.0
gnomAD
AFR
AMR
ASJ
EAS
EST
FIN
NFE
NWE
SEU
101500000
101600000
101700000
101800000
101900000
PCNP
RPL24
SENP7
TRMT10C
ZBTB11
CEP97
NXPE3
NFKBIZ
Chromosome 3
Distance
LD
Coding
eQTL
SENP7
TRMT10C
PCNP
ZBTB11
RPL24
CEP97
NXPE3
NFKBIZ

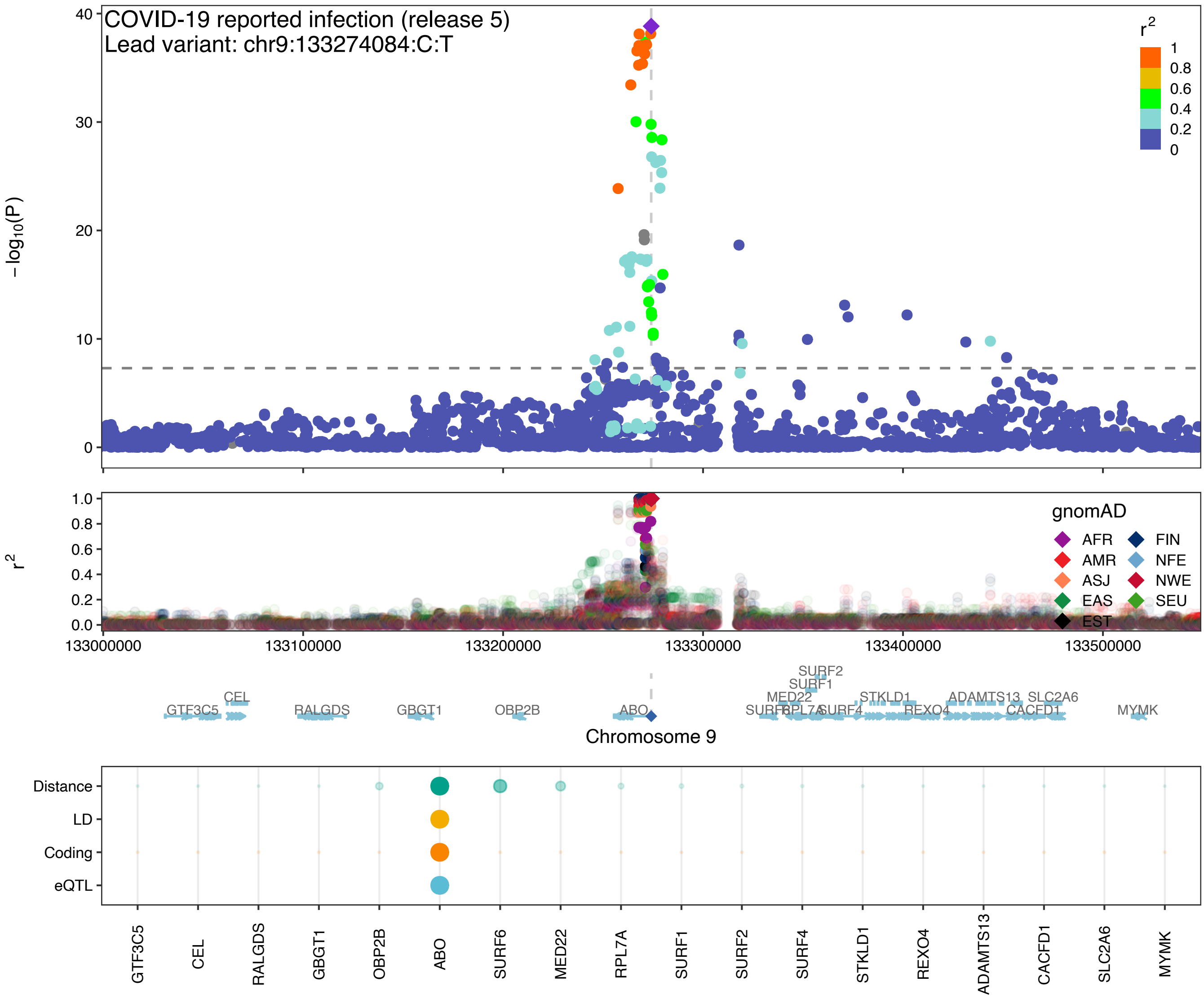
COVID-19 reported infection (release 5)
Lead variant: chr9:133274084:C:T
−log10(P)
r2
gnomAD
AFR
AMR
ASJ
EAS
EST
FIN
NFE
NWE
SEU
Chromosome 9
Distance
LD
Coding
eQTL
GTF3C5
CEL
RALGDS
GBGT1
OBP2B
ABO
SURF6
MED22
RPL7A
SURF1
SURF2
SURF4
STKLD1
REXO4
ADAMTS13
CACFD1
SLC2A6
MYMK

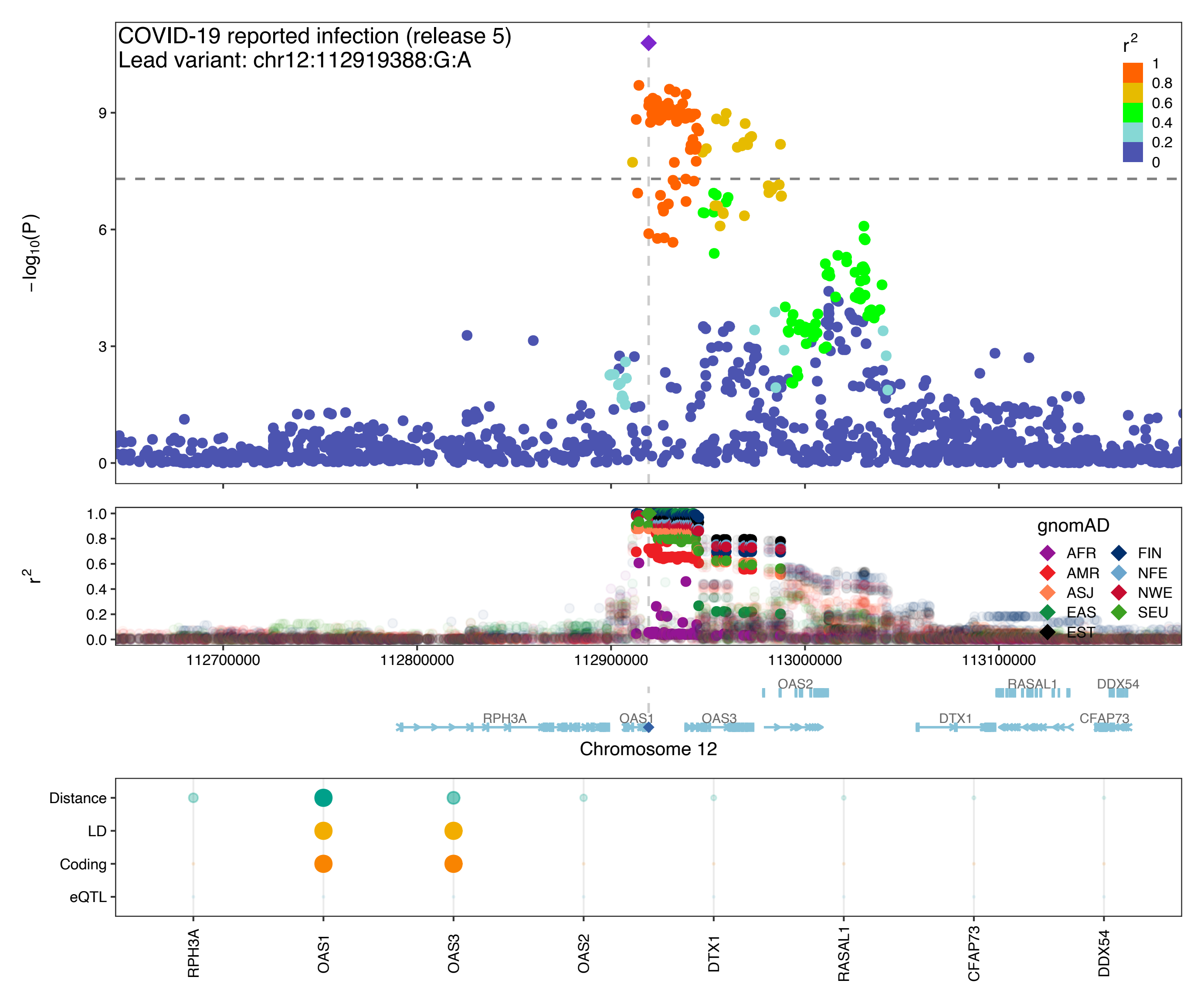

COVID-19 reported infection (release 5)
Lead variant: chr12:112919388:G:A
−log10(P)
r2
1
0.8
0.6
0.4
0.2
0
gnomAD
AFR
AMR
ASJ
EAS
EST
FIN
NFE
NWE
SEU
112700000
112800000
112900000
113000000
113100000
OAS2
RASAL1
DDX54
RPH3A
OAS1
OAS3
DTX1
CFAP73
Chromosome 12
Distance
LD
Coding
eQTL
RPH3A
OAS1
OAS3
OAS2
DTX1
RASAL1
CFAP73
DDX54

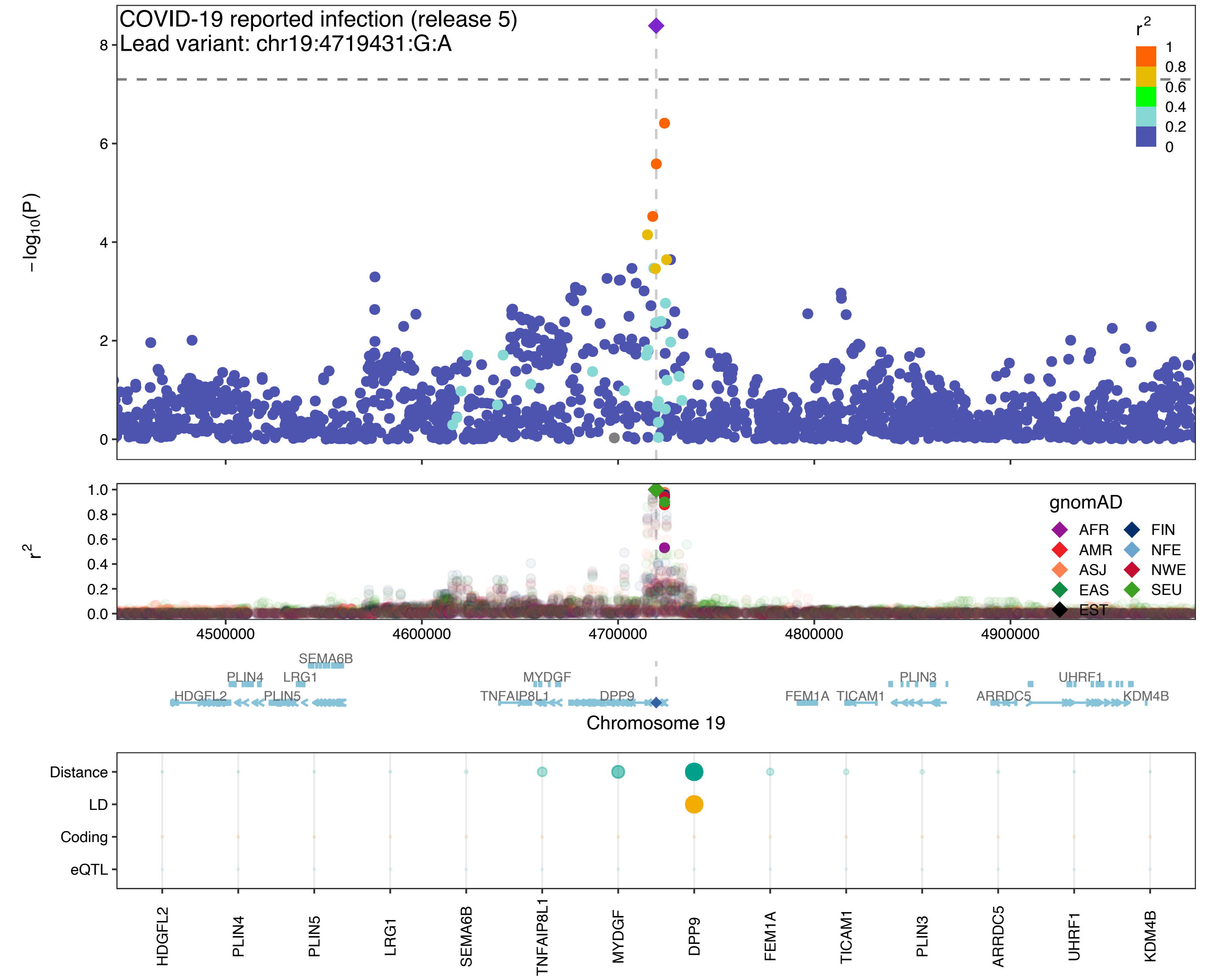
COVID-19 reported infection (release 5)
Lead variant: chr19:4719431:G:A
$-log_{10}(P)$
$r^2$
1
0.8
0.6
0.4
0.2
0
gnomAD
AFR
AMR
ASJ
EAS
EST
FIN
NFE
NWE
SEU
4500000
4600000
4700000
4800000
4900000
SEMA6B
PLIN4
LRG1
HDGFL2
PLIN5
MYDGF
TNFAIP8L1
DPP9
FEM1A
TICAM1
PLIN3
ARRDC5
UHRF1
KDM4B
Chromosome 19
Distance
LD
Coding
eQTL
HDGFL2
PLIN4
PLIN5
LRG1
SEMA6B
TNFAIP8L1
MYDGF
DPP9
FEM1A
TICAM1
PLIN3
ARRDC5
UHRF1
KDM4B

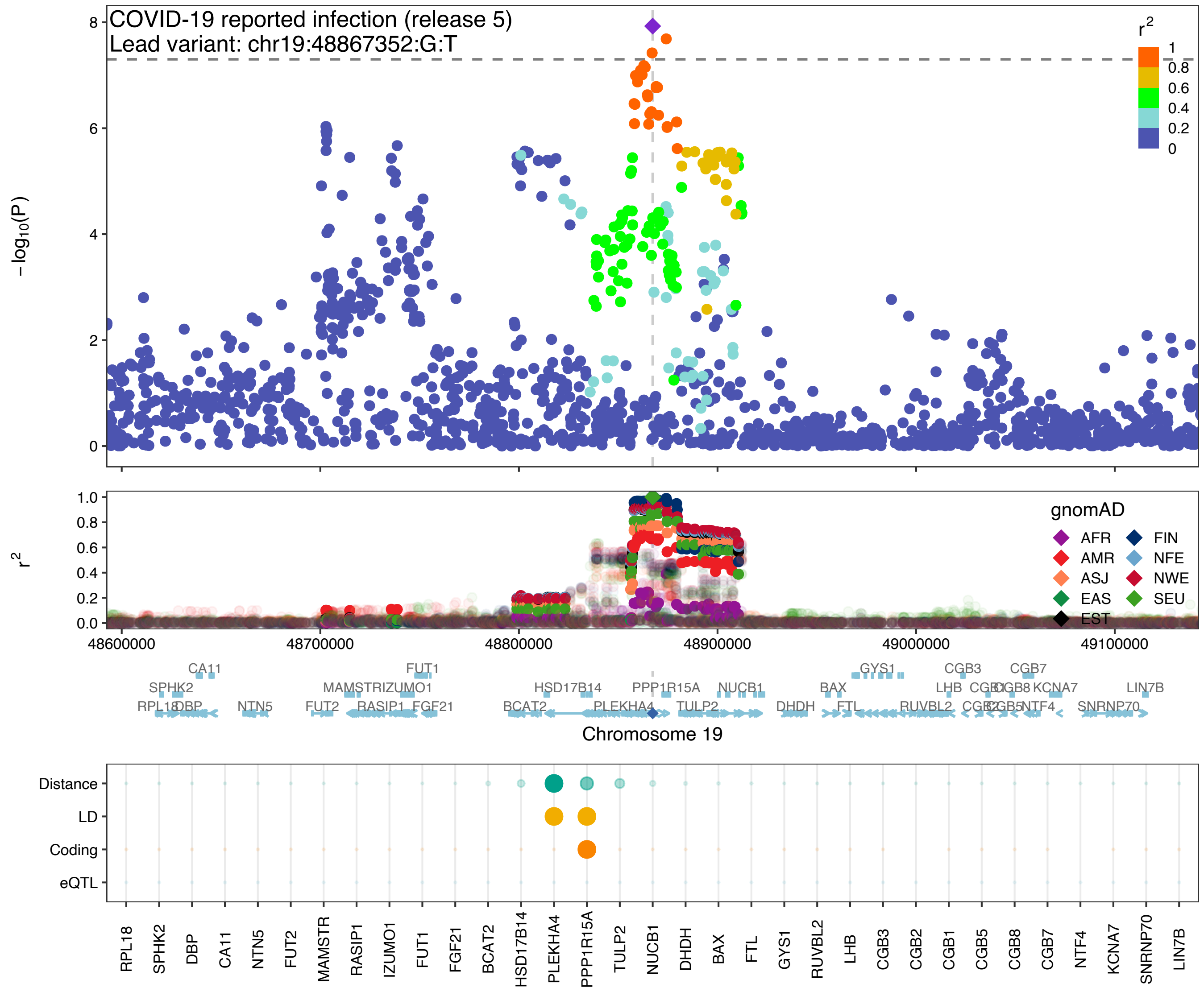

COVID-19 reported infection (release 5)
Lead variant: chr19:48867352:G:T
−log10(P)
r2
1
0.8
0.6
0.4
0.2
0
gnomAD
AFR
AMR
ASJ
EAS
EST
FIN
NFE
NWE
SEU
48600000
48700000
48800000
48900000
49000000
49100000
Chromosome 19
Distance
LD
Coding
eQTL
RPL18
SPHK2
DBP
CA11
NTN5
FUT2
MAMSTR
RASIP1
IZUMO1
FUT1
FGF21
BCAT2
HSD17B14
PLEKHA4
PPP1R15A
TULP2
NUCB1
DHDH
BAX
FTL
GYS1
RUVBL2
LHB
CGB3
CGB2
CGB1
CGB5
CGB8
CGB7
NTF4
KCNA7
SNRNP70
LIN7B
