## Extended data and supplement for "Mapping the human genetic architecture of COVID-19 by worldwide meta-analysis": Extended_Data_Figure_10.pdf

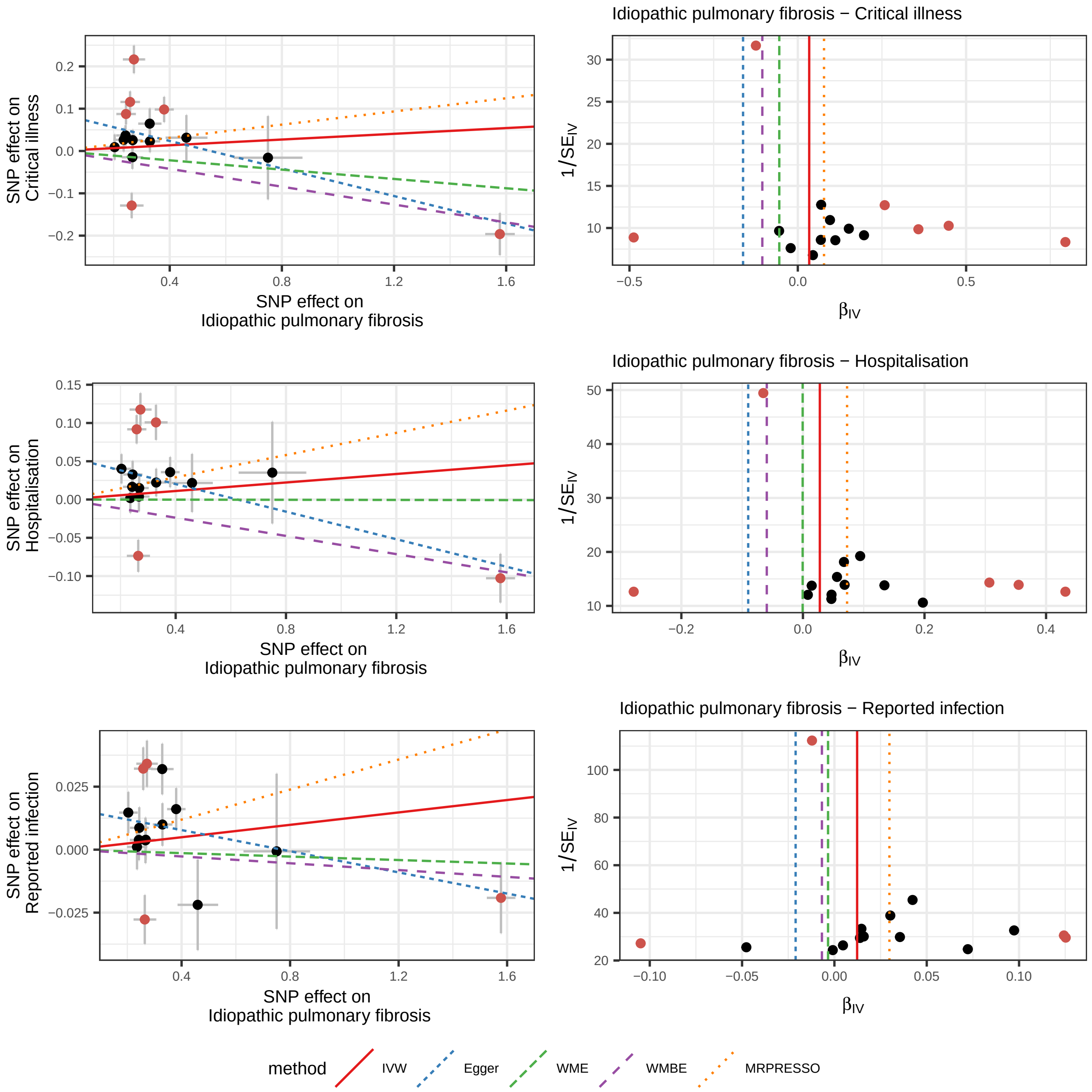

Idiopathic pulmonary fibrosis − Critical illness
Idiopathic pulmonary fibrosis − Hospitalisation
Idiopathic pulmonary fibrosis − Reported infection
SNP effect on Critical illness
SNP effect on Hospitalisation
SNP effect on Reported infection
SNP effect on Idiopathic pulmonary fibrosis
1/SE_IV
β_IV
method
IVW
Egger
WME
WMBE
MRPRESSO

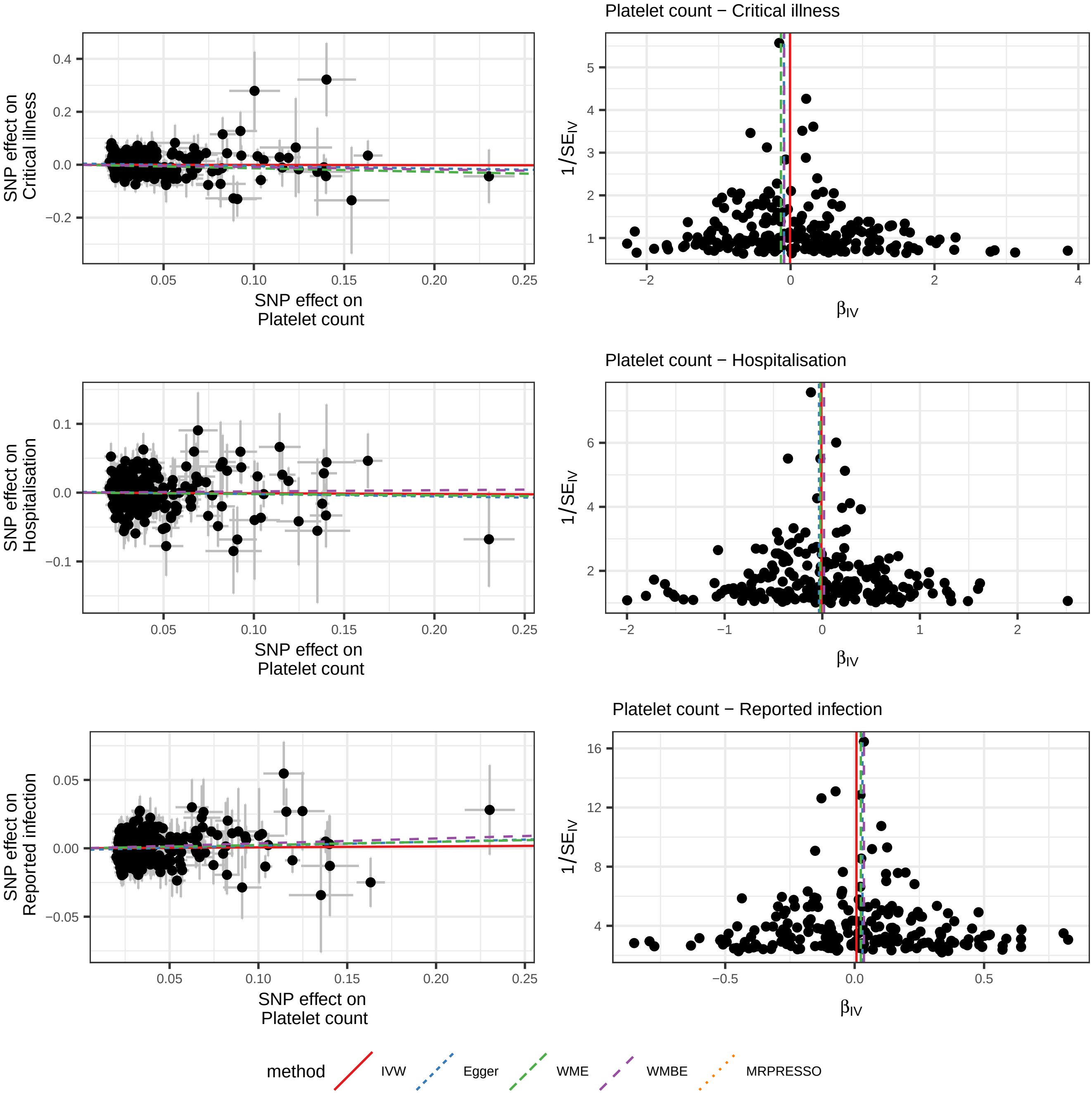

Platelet count − Critical illness
Platelet count − Hospitalisation
Platelet count − Reported infection
SNP effect on Critical illness
SNP effect on Hospitalisation
SNP effect on Reported infection
SNP effect on Platelet count
1/SE_IV
β_IV
method
IVW
Egger
WME
WMBE
MRPRESSO

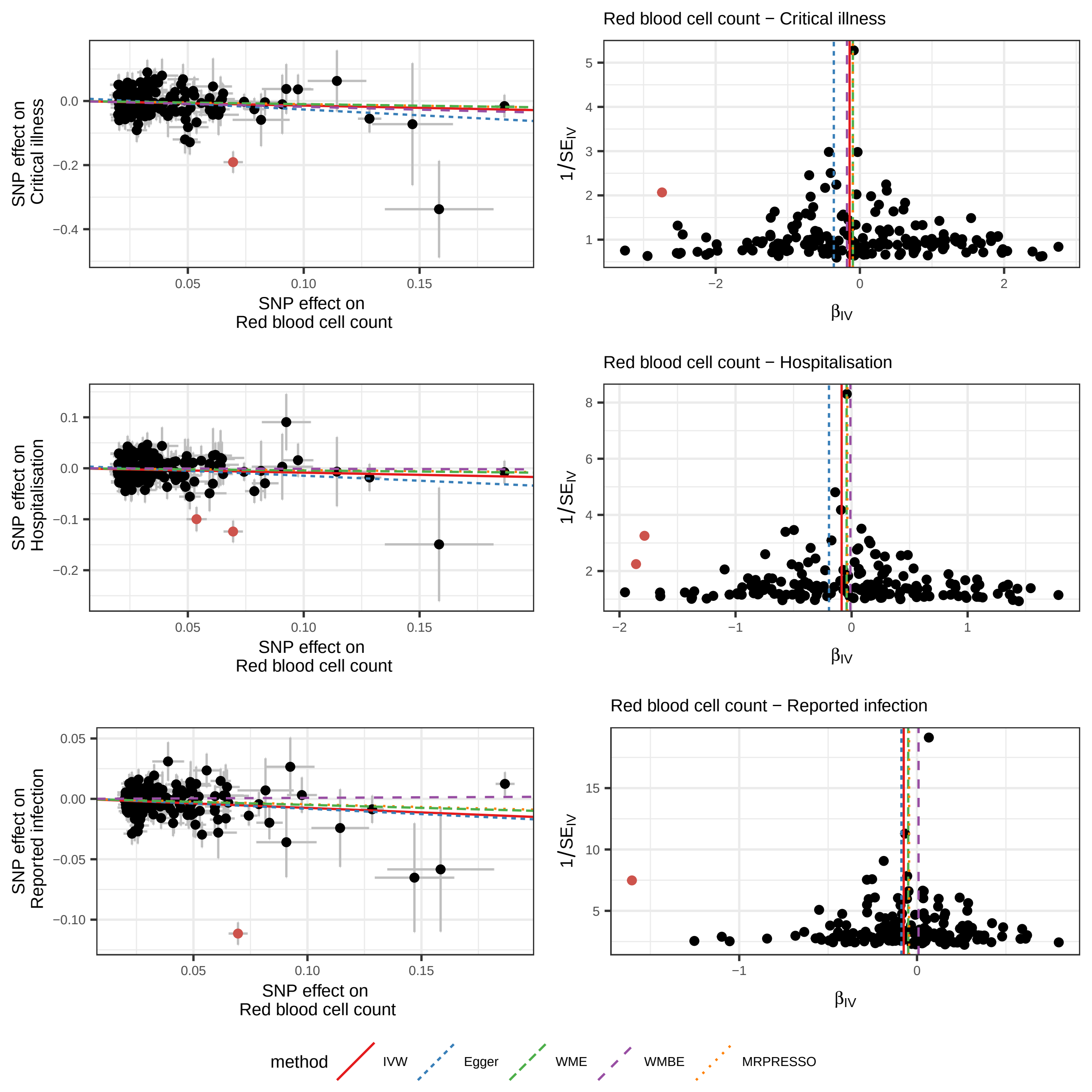

Red blood cell count − Critical illness
SNP effect on Critical illness
SNP effect on Red blood cell count
1/SE_IV
β_IV
Red blood cell count − Hospitalisation
SNP effect on Hospitalisation
SNP effect on Red blood cell count
1/SE_IV
β_IV
Red blood cell count − Reported infection
SNP effect on Reported infection
SNP effect on Red blood cell count
1/SE_IV
β_IV
method
IVW
Egger
WME
WMBE
MRPRESSO

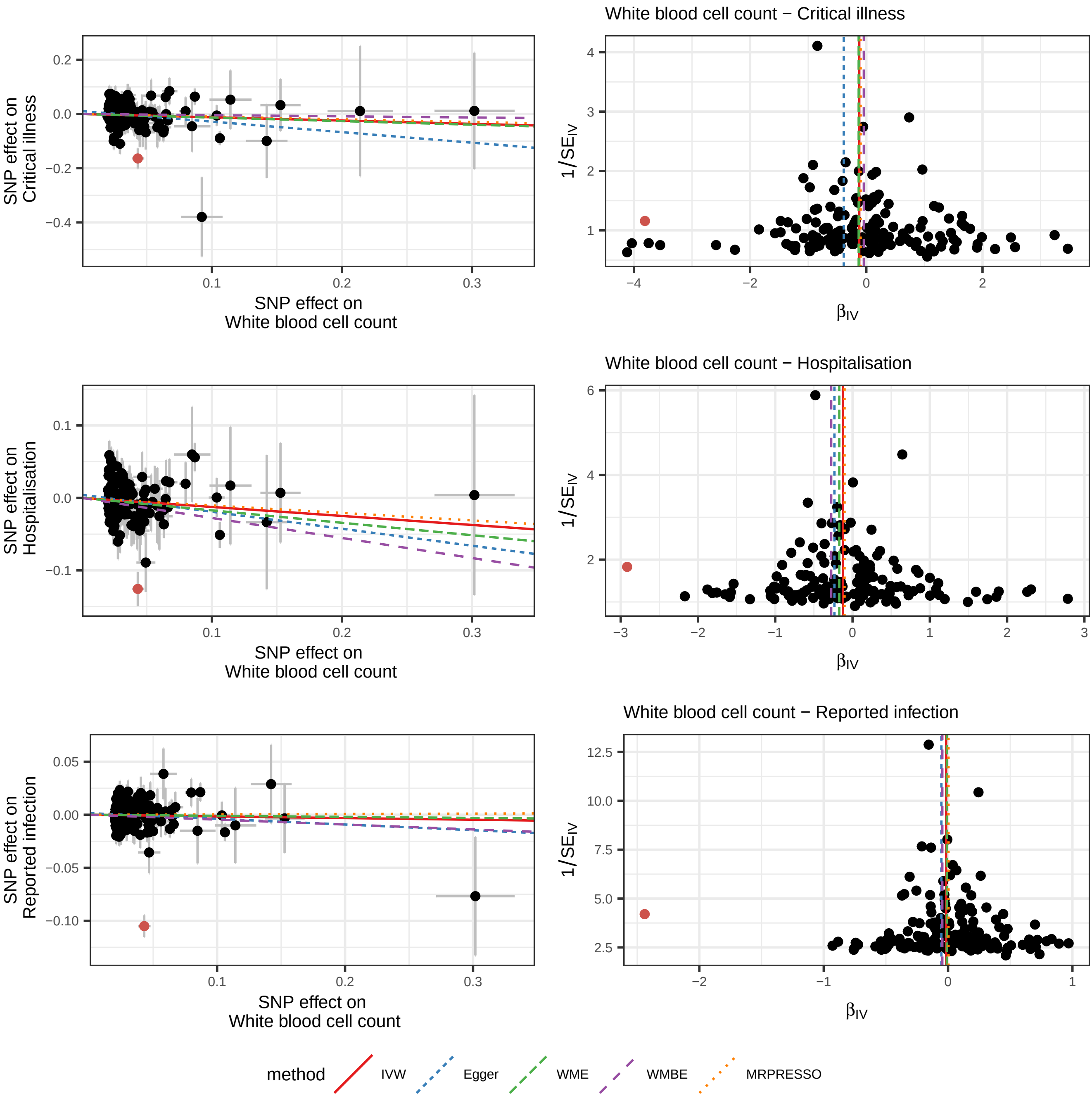

White blood cell count − Critical illness
SNP effect on Critical illness
SNP effect on White blood cell count
1/SE_IV
β_IV
White blood cell count − Hospitalisation
SNP effect on Hospitalisation
SNP effect on White blood cell count
1/SE_IV
β_IV
White blood cell count − Reported infection
SNP effect on Reported infection
SNP effect on White blood cell count
1/SE_IV
β_IV
method
IVW
Egger
WME
WMBE
MRPRESSO

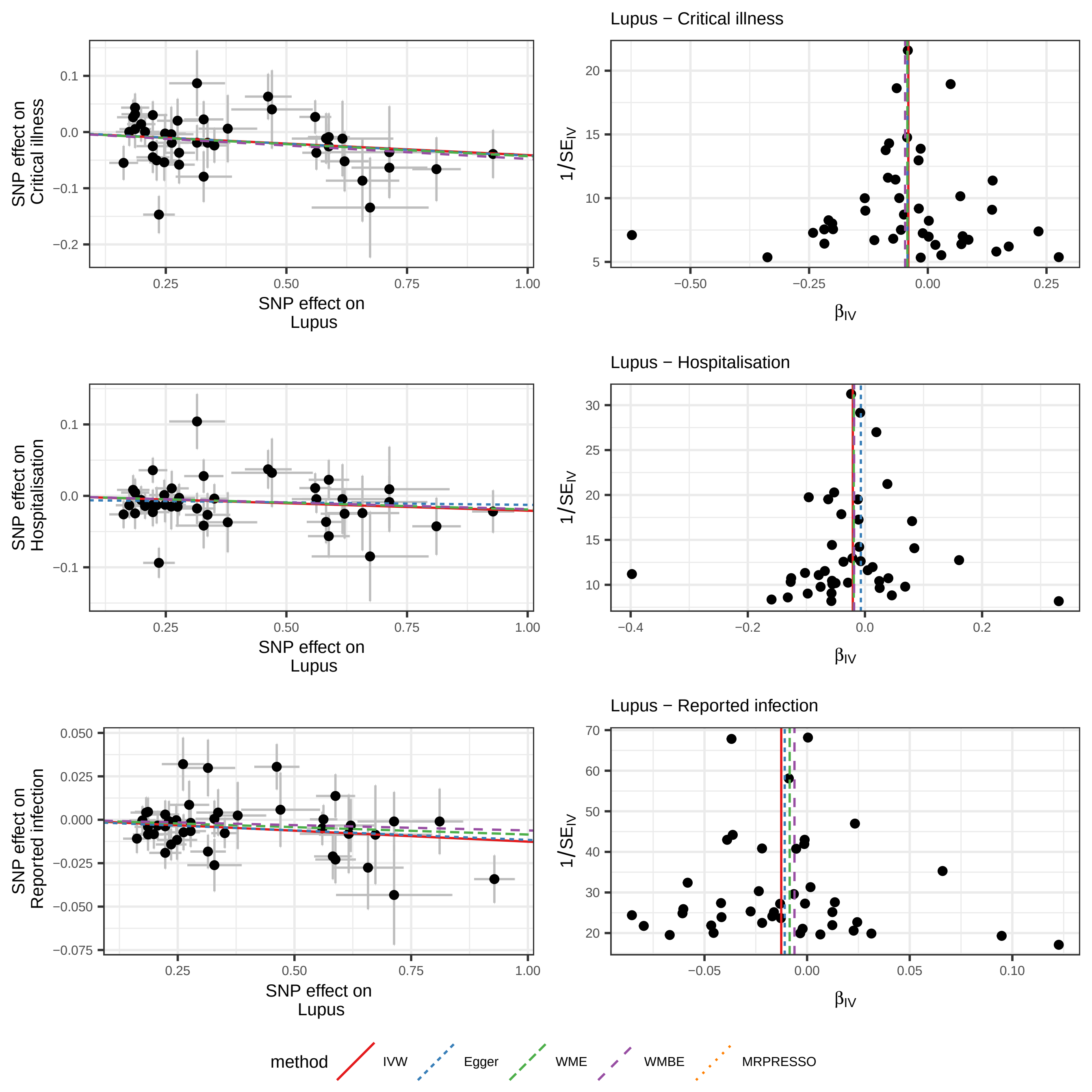

Lupus − Critical illness
SNP effect on Critical illness
SNP effect on Lupus
$1/SE_{IV}$
$\beta_{IV}$
Lupus − Hospitalisation
SNP effect on Hospitalisation
SNP effect on Lupus
$1/SE_{IV}$
$\beta_{IV}$
Lupus − Reported infection
SNP effect on Reported infection
SNP effect on Lupus
$1/SE_{IV}$
$\beta_{IV}$
method
IVW
Egger
WME
WMBE
MRPRESSO

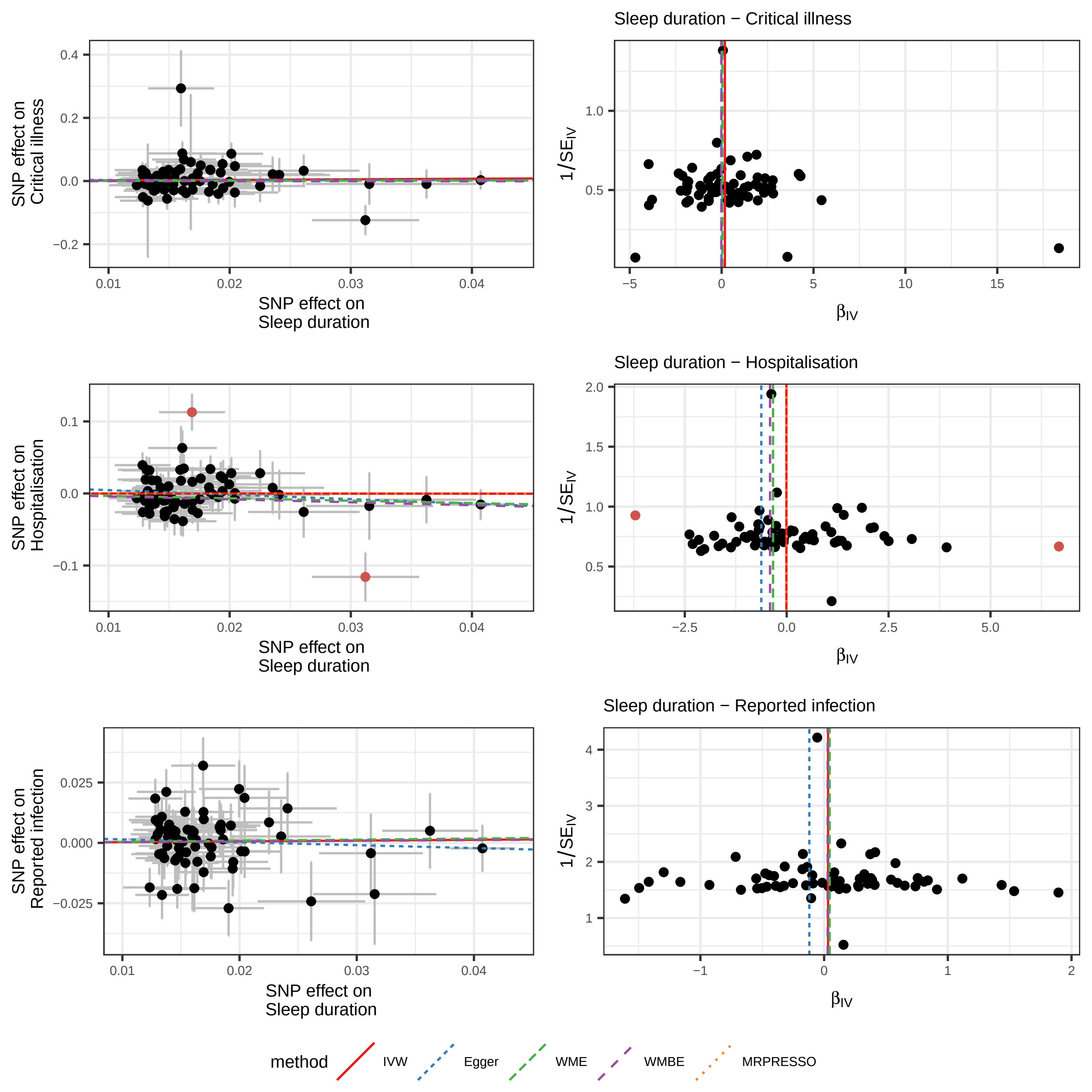

Sleep duration − Critical illness
Sleep duration − Hospitalisation
Sleep duration − Reported infection
SNP effect on Critical illness
SNP effect on Hospitalisation
SNP effect on Reported infection
SNP effect on Sleep duration
$1/SE_{IV}$
$\beta_{IV}$
method
IVW
Egger
WME
WMBE
MRPRESSO

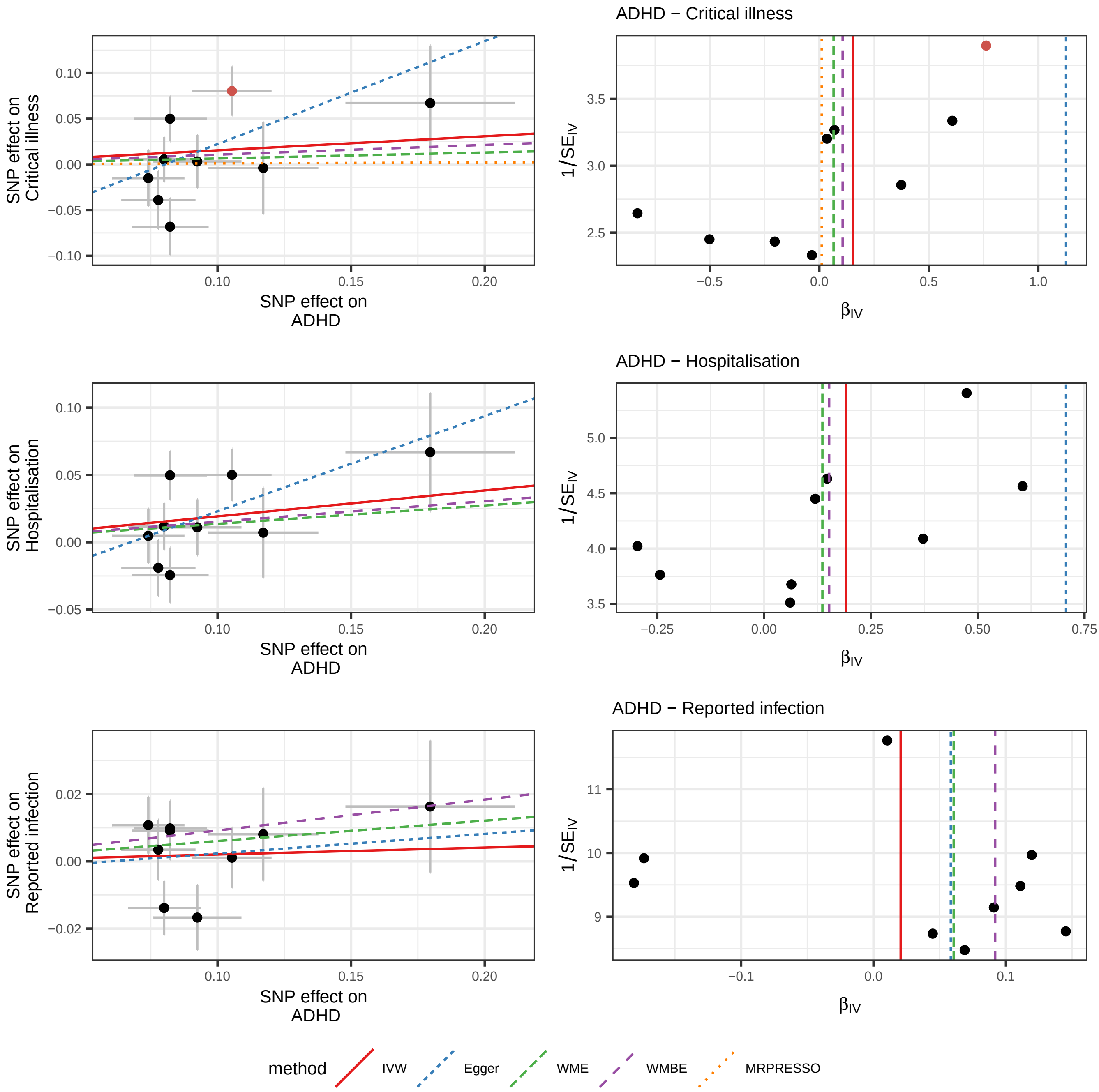

ADHD − Critical illness
ADHD − Hospitalisation
ADHD − Reported infection
SNP effect on Critical illness
SNP effect on Hospitalisation
SNP effect on Reported infection
SNP effect on ADHD
1/SE$_{IV}$
β$_{IV}$
method
IVW
Egger
WME
WMBE
MRPRESSO

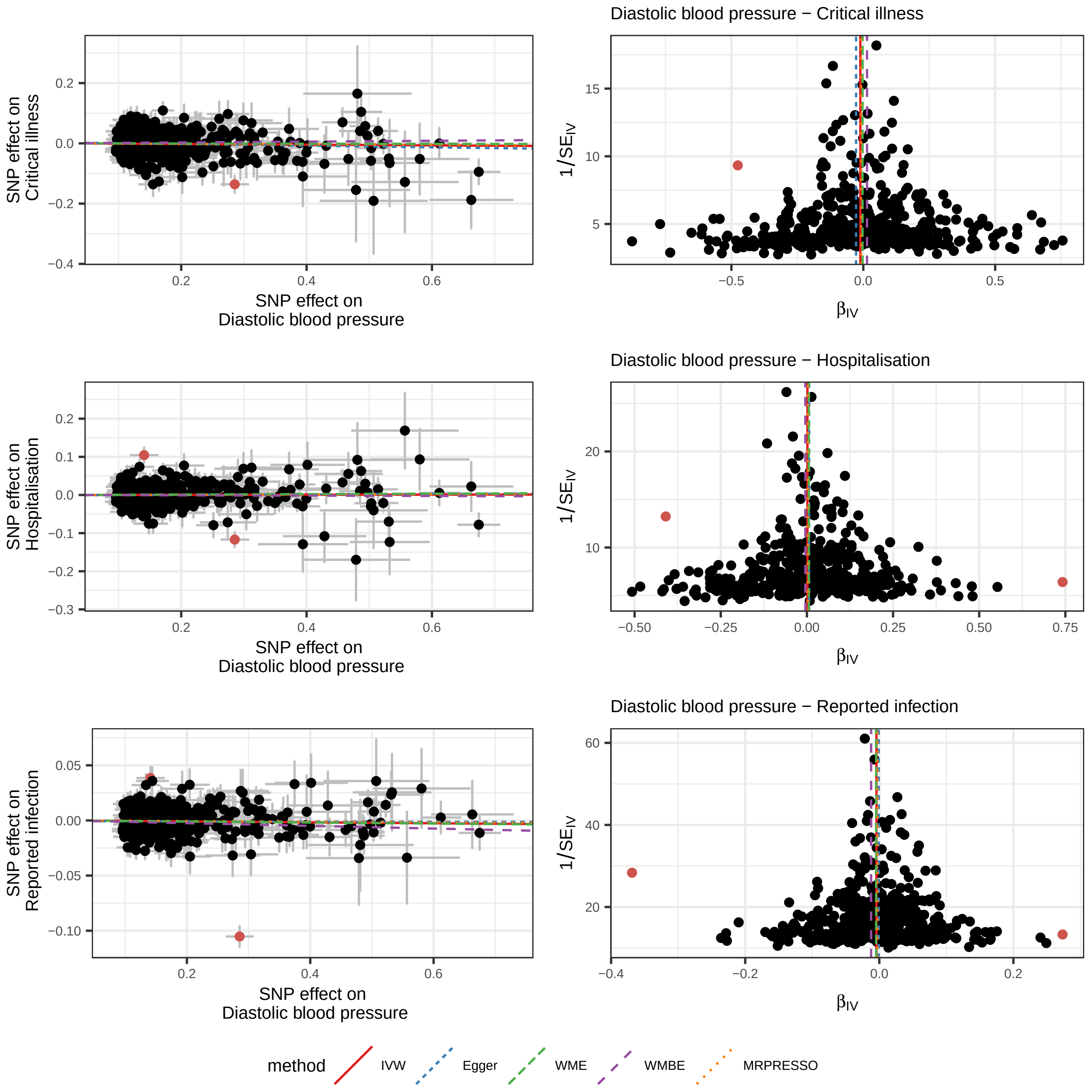

Diastolic blood pressure − Critical illness
Diastolic blood pressure − Hospitalisation
Diastolic blood pressure − Reported infection
SNP effect on Critical illness
SNP effect on Hospitalisation
SNP effect on Reported infection
SNP effect on Diastolic blood pressure
$1/SE_{IV}$
$\beta_{IV}$
method
IVW
Egger
WME
WMBE
MRPRESSO

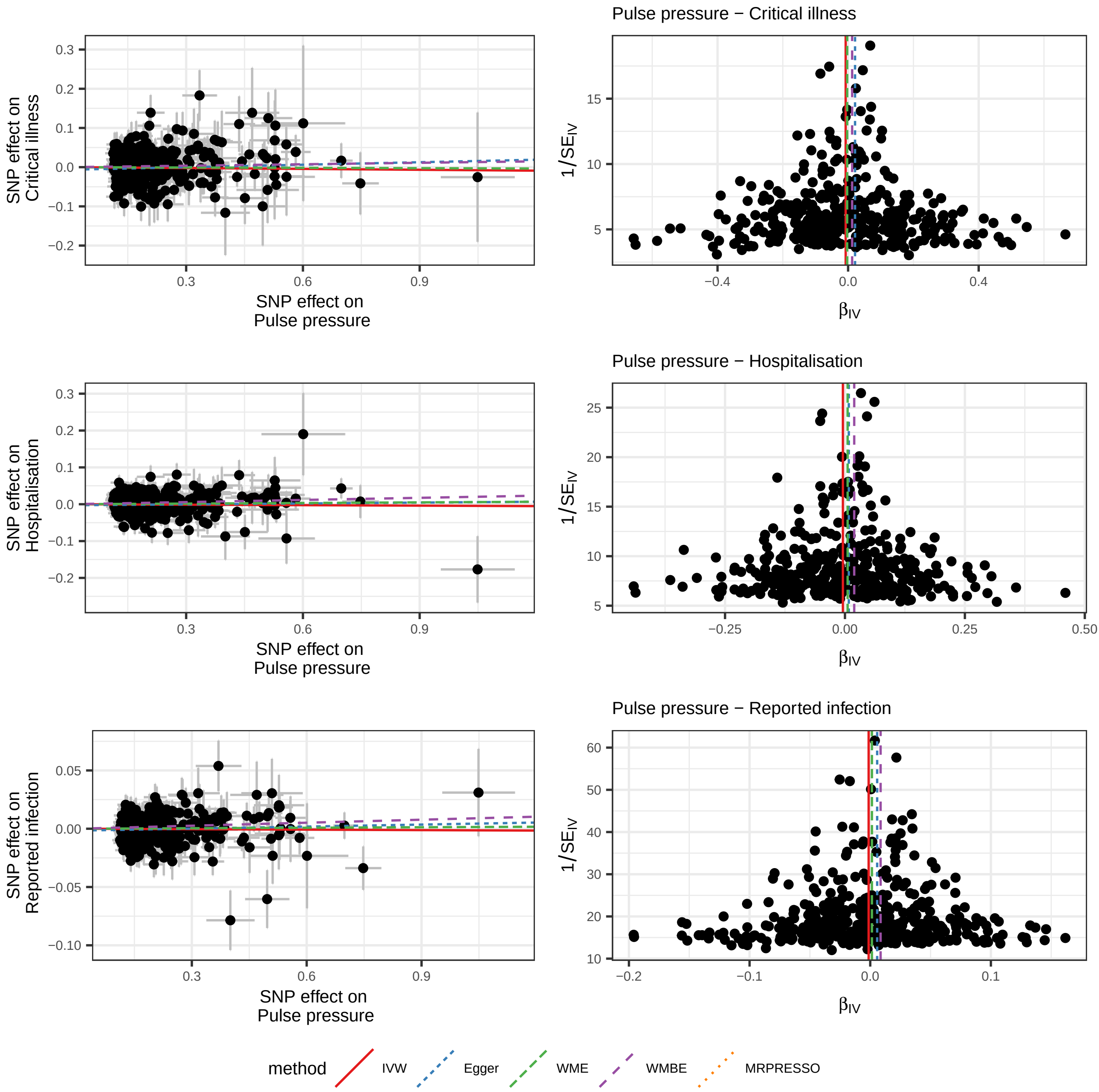

Pulse pressure − Critical illness
Pulse pressure − Hospitalisation
Pulse pressure − Reported infection
SNP effect on Critical illness
SNP effect on Hospitalisation
SNP effect on Reported infection
SNP effect on Pulse pressure
1/SE_IV
β_IV
method
IVW
Egger
WME
WMBE
MRPRESSO

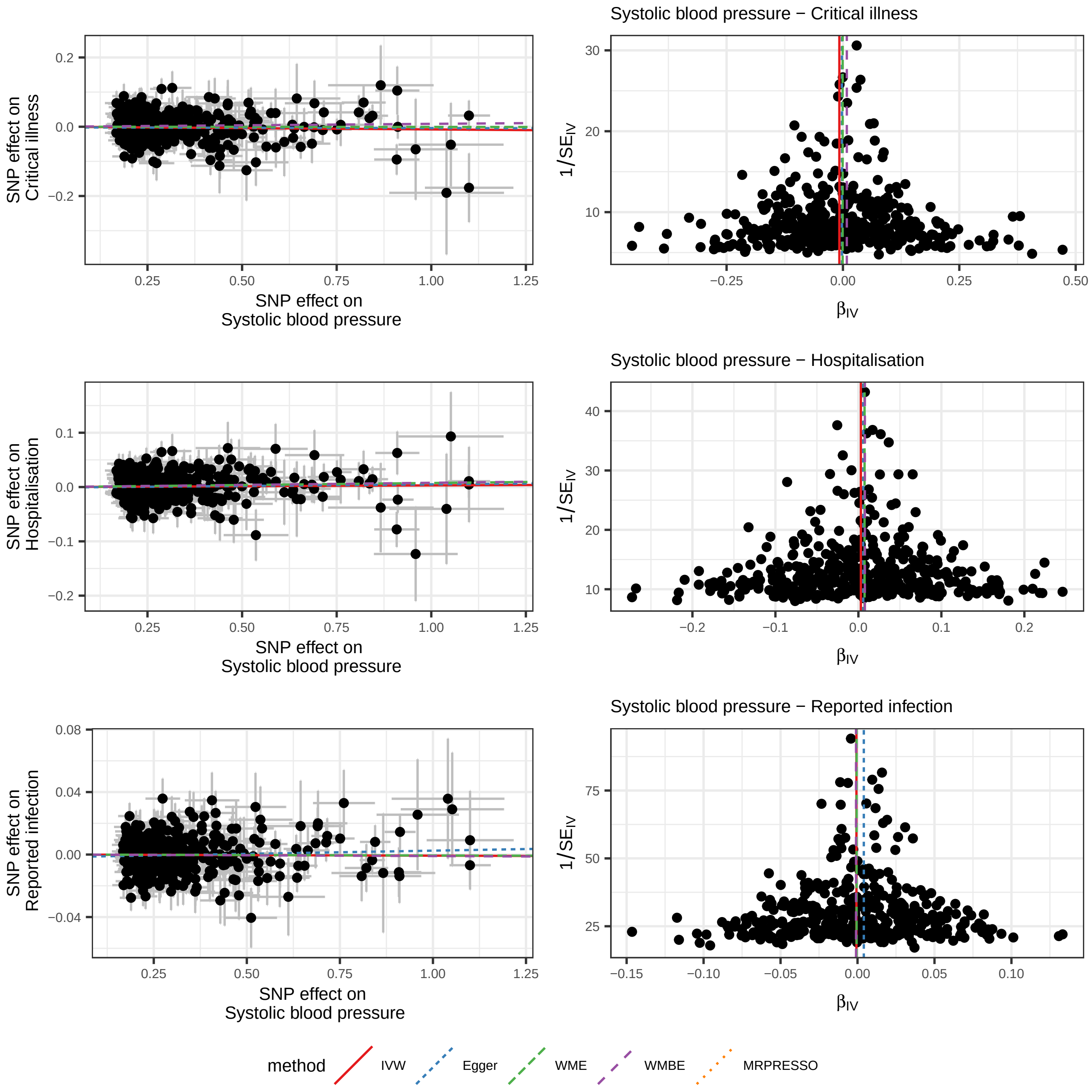

Systolic blood pressure − Critical illness
SNP effect on Critical illness
SNP effect on Systolic blood pressure
$1/SE_{IV}$
$\beta_{IV}$
Systolic blood pressure − Hospitalisation
SNP effect on Hospitalisation
SNP effect on Systolic blood pressure
$1/SE_{IV}$
$\beta_{IV}$
Systolic blood pressure − Reported infection
SNP effect on Reported infection
SNP effect on Systolic blood pressure
$1/SE_{IV}$
$\beta_{IV}$
method
IVW
Egger
WME
WMBE
MRPRESSO

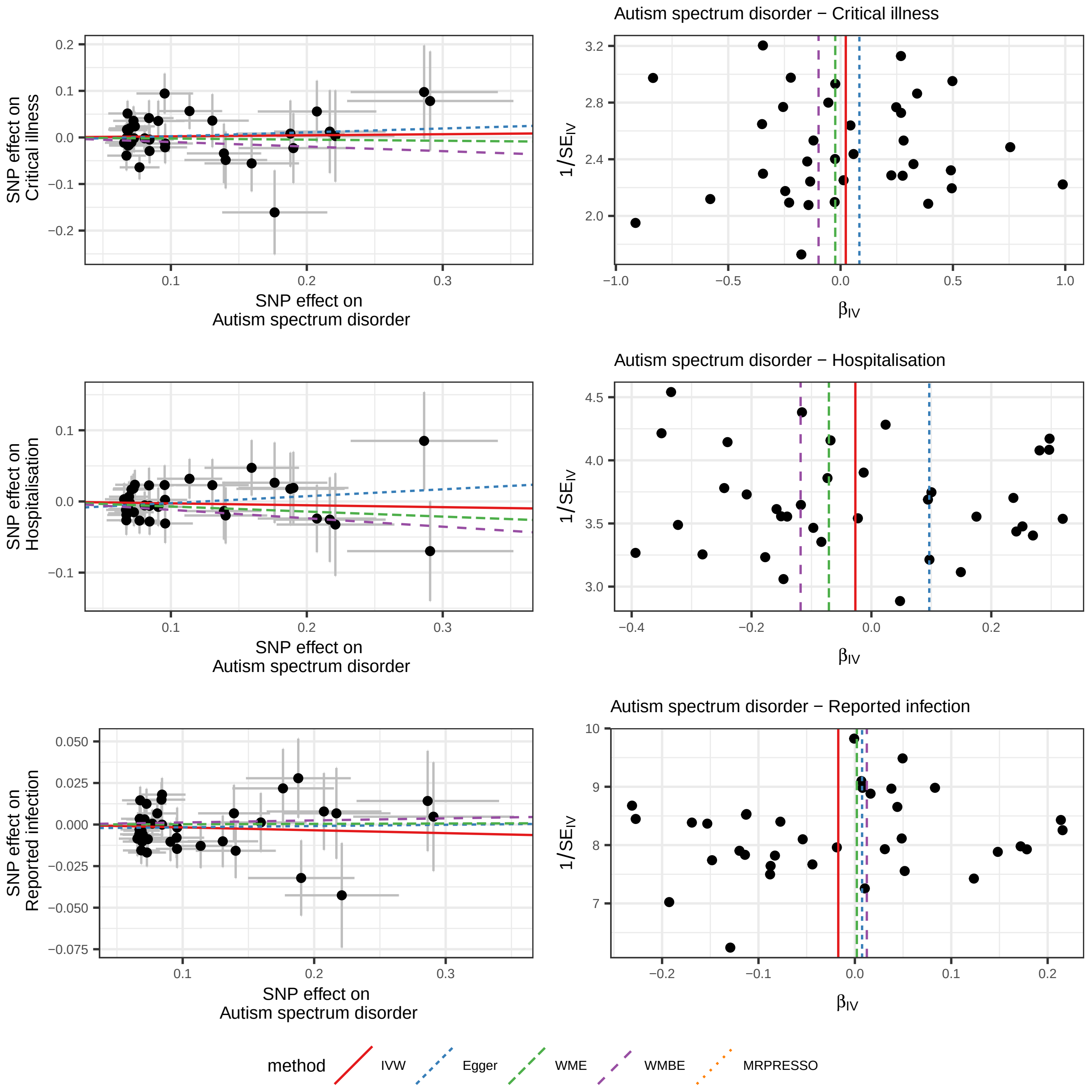

Autism spectrum disorder − Critical illness
SNP effect on Critical illness
SNP effect on Autism spectrum disorder
1/SE_IV
β_IV
Autism spectrum disorder − Hospitalisation
SNP effect on Hospitalisation
SNP effect on Autism spectrum disorder
1/SE_IV
β_IV
Autism spectrum disorder − Reported infection
SNP effect on Reported infection
SNP effect on Autism spectrum disorder
1/SE_IV
β_IV
method
IVW
Egger
WME
WMBE
MRPRESSO

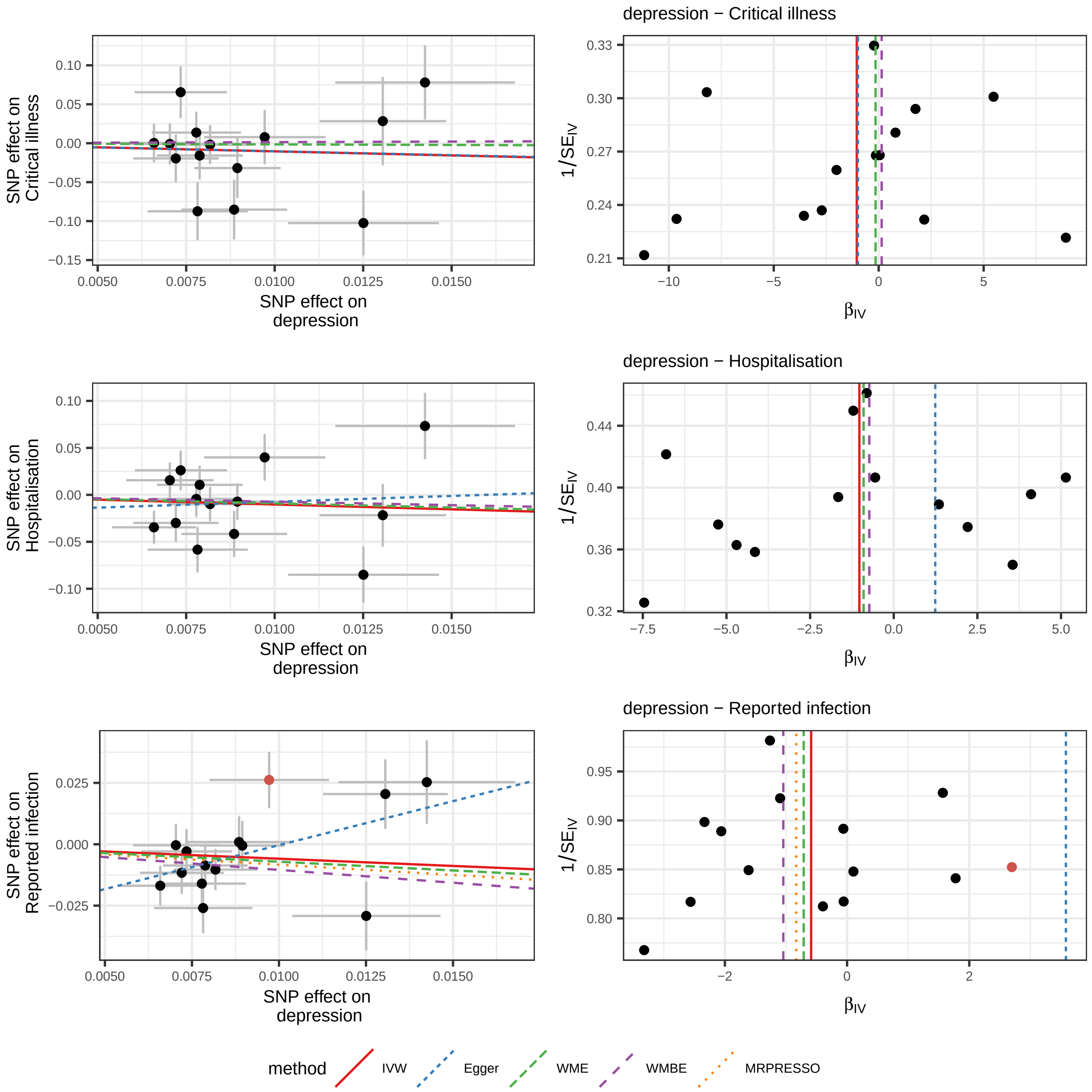

depression − Critical illness
depression − Hospitalisation
depression − Reported infection
SNP effect on Critical illness
SNP effect on Hospitalisation
SNP effect on Reported infection
SNP effect on depression
1/SE_IV
β_IV
method
IVW
Egger
WME
WMBE
MRPRESSO

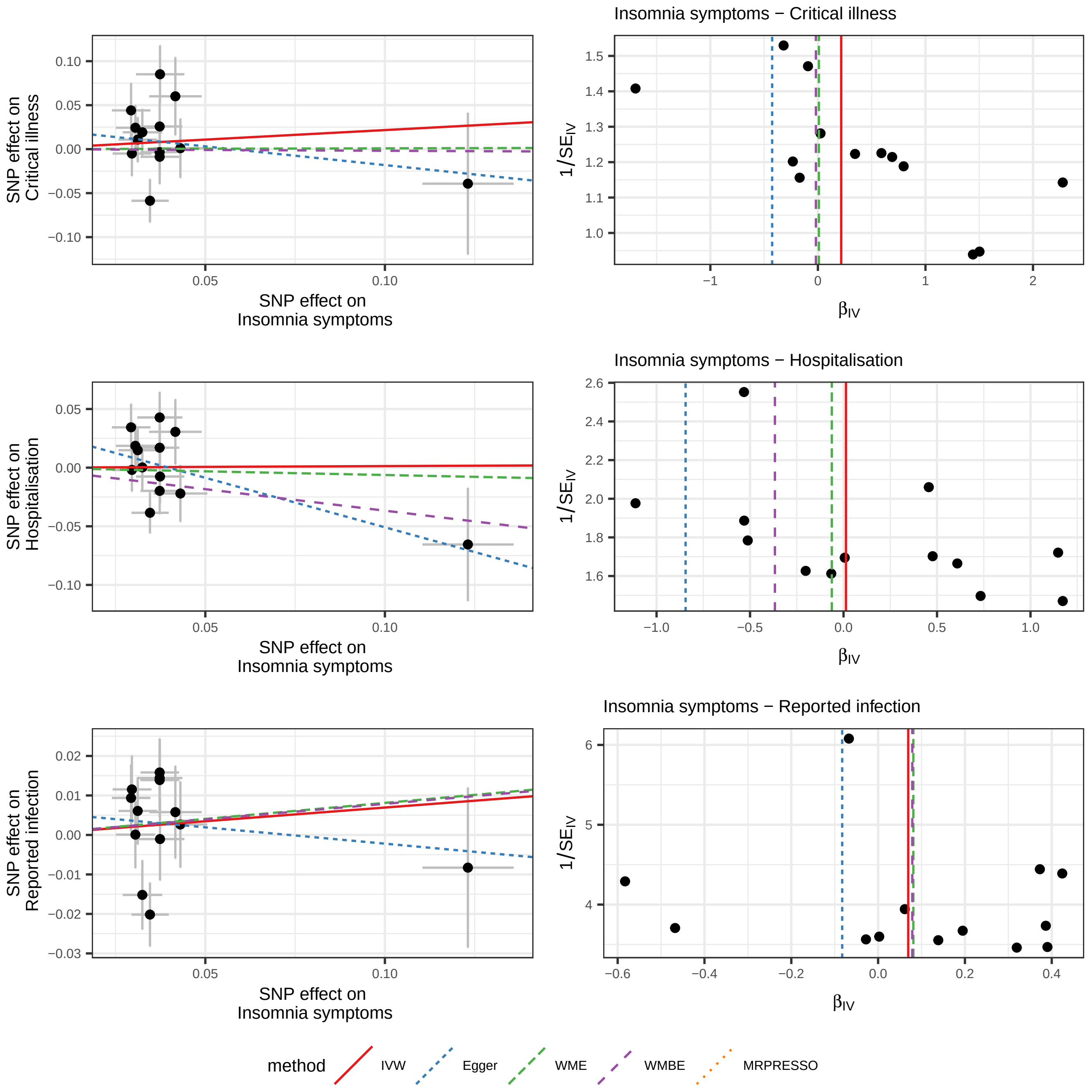

Insomnia symptoms − Critical illness
SNP effect on Critical illness
SNP effect on Insomnia symptoms
Insomnia symptoms − Hospitalisation
SNP effect on Hospitalisation
SNP effect on Insomnia symptoms
Insomnia symptoms − Reported infection
SNP effect on Reported infection
SNP effect on Insomnia symptoms
$1/SE_{IV}$
$\beta_{IV}$
method
IVW
Egger
WME
WMBE
MRPRESSO

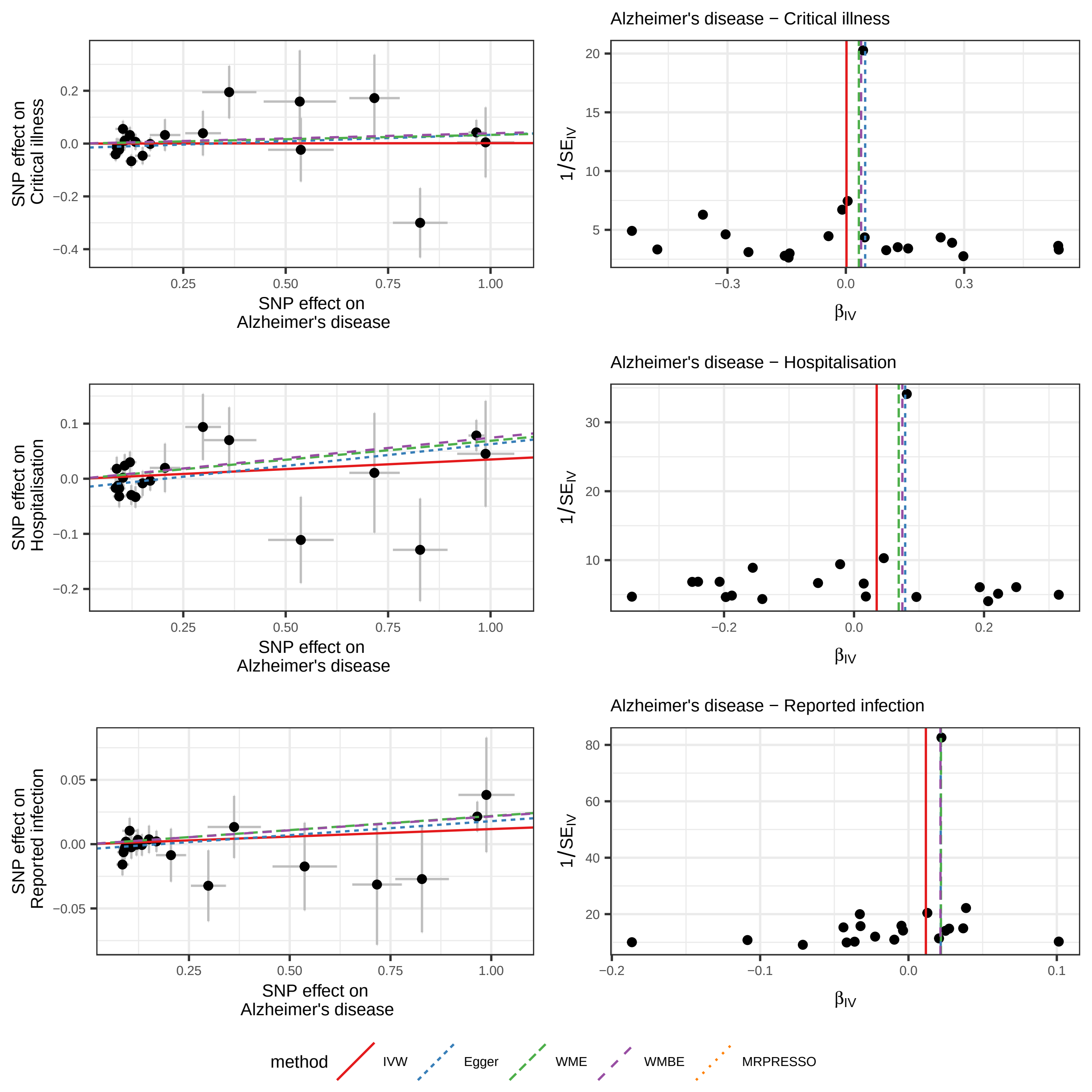

Alzheimer's disease − Critical illness
Alzheimer's disease − Hospitalisation
Alzheimer's disease − Reported infection
SNP effect on Critical illness
SNP effect on Hospitalisation
SNP effect on Reported infection
SNP effect on Alzheimer's disease
1/SE_IV
β_IV
method
IVW
Egger
WME
WMBE
MRPRESSO

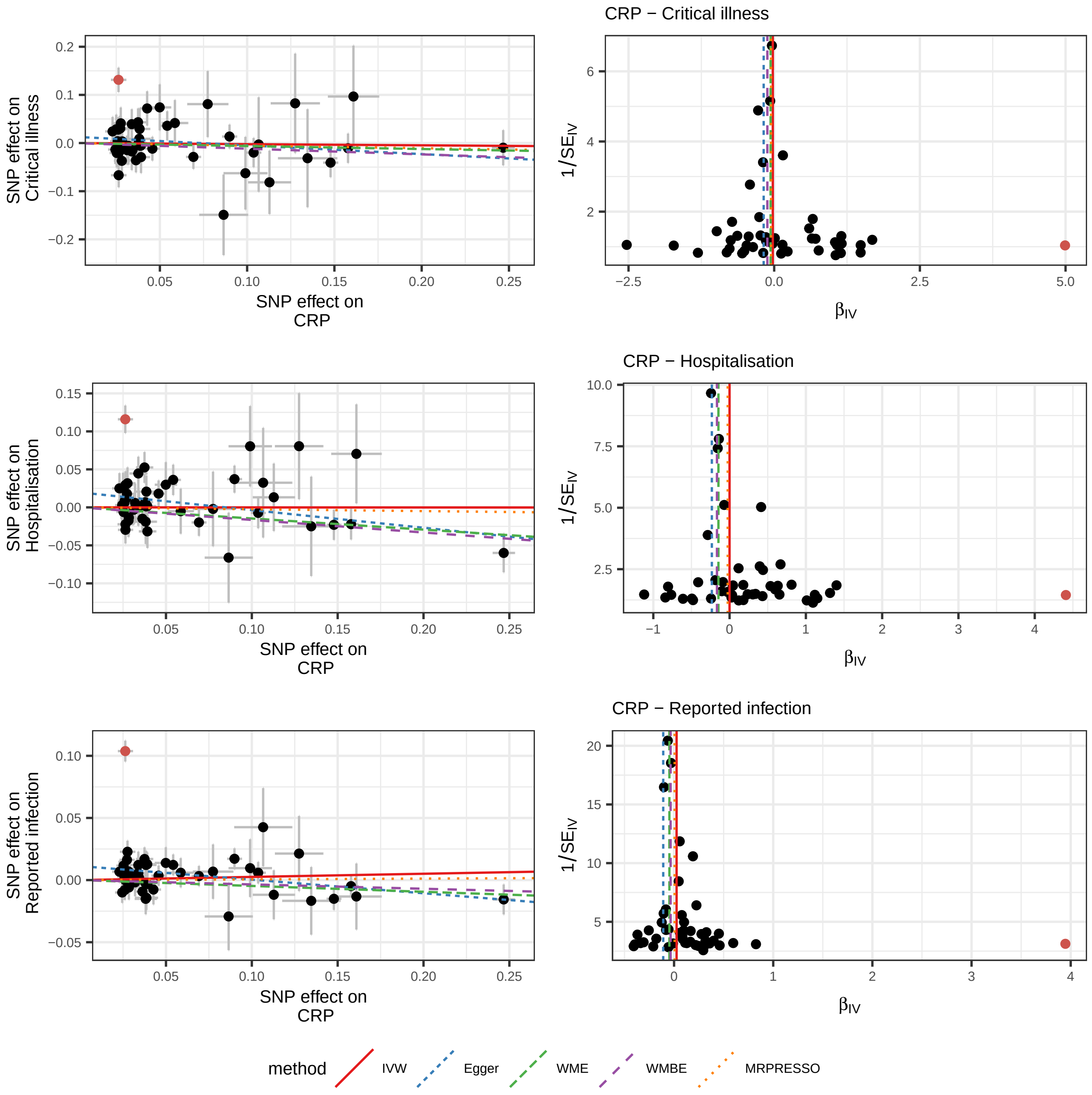

CRP − Critical illness
CRP − Hospitalisation
CRP − Reported infection
SNP effect on Critical illness
SNP effect on Hospitalisation
SNP effect on Reported infection
SNP effect on CRP
1/SE_IV
β_IV
method
IVW
Egger
WME
WMBE
MRPRESSO

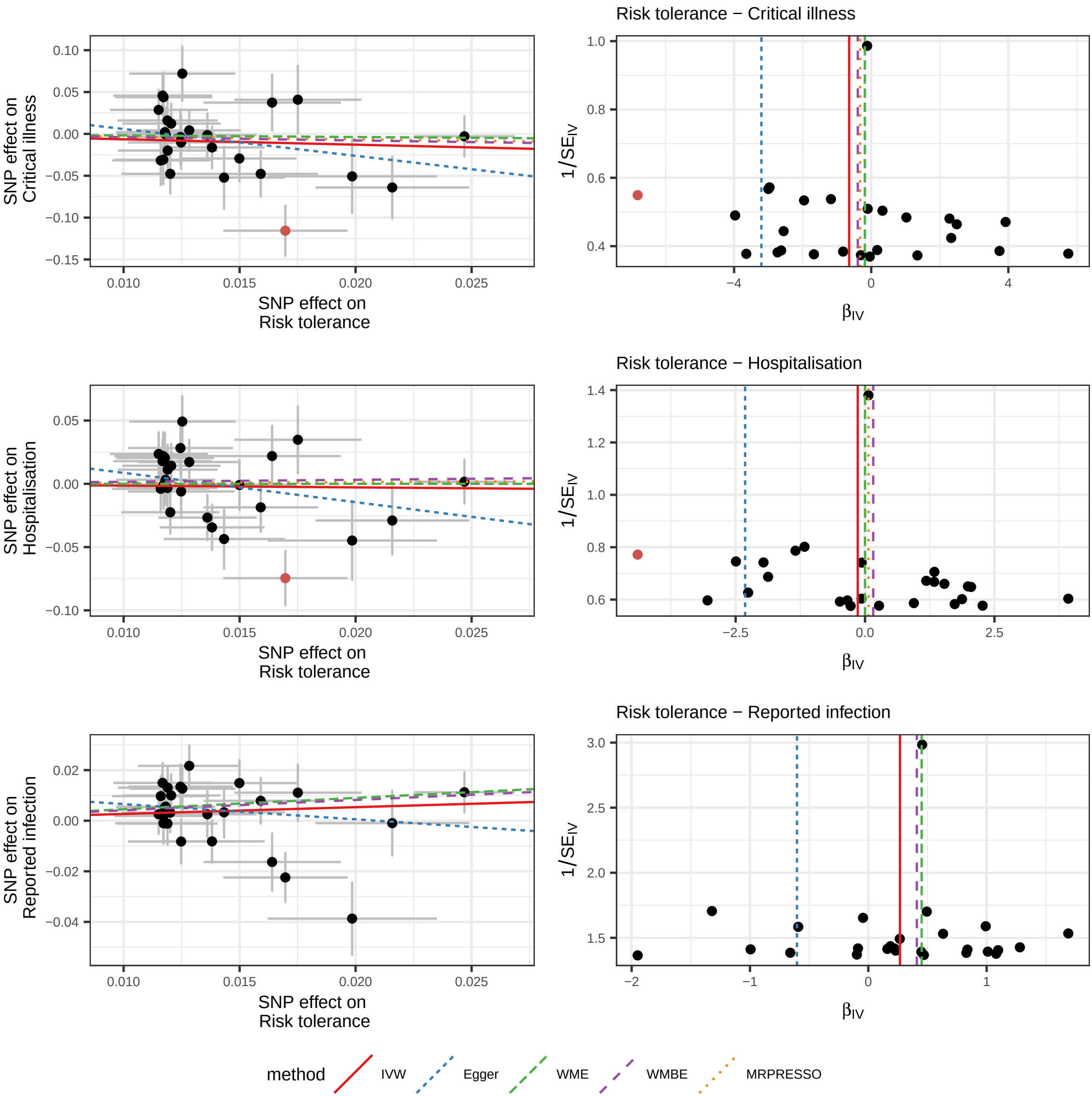

Risk tolerance − Critical illness
Risk tolerance − Hospitalisation
Risk tolerance − Reported infection
SNP effect on Critical illness
SNP effect on Hospitalisation
SNP effect on Reported infection
SNP effect on Risk tolerance
1/SE_IV
β_IV
method
IVW
Egger
WME
WMBE
MRPRESSO

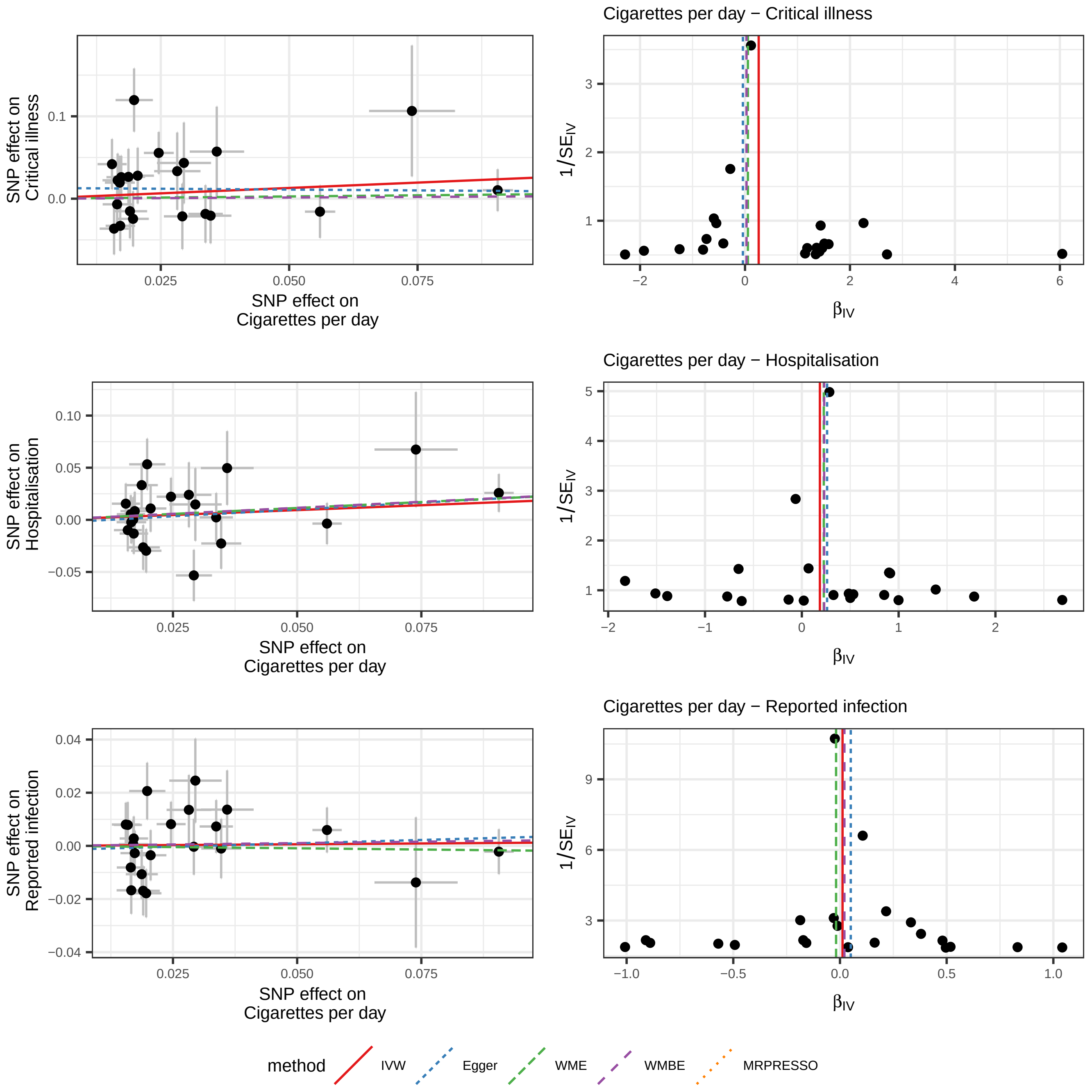

Cigarettes per day − Critical illness
SNP effect on Critical illness
0.1
0.0
0.025
0.050
0.075
SNP effect on Cigarettes per day
1/SE_IV
3
2
1
−2
0
2
4
6
β_IV
Cigarettes per day − Hospitalisation
SNP effect on Hospitalisation
0.10
0.05
0.00
−0.05
0.025
0.050
0.075
SNP effect on Cigarettes per day
1/SE_IV
5
4
3
2
1
−2
−1
0
1
2
β_IV
Cigarettes per day − Reported infection
SNP effect on Reported infection
0.04
0.02
0.00
−0.02
−0.04
0.025
0.050
0.075
SNP effect on Cigarettes per day
1/SE_IV
9
6
3
−1.0
−0.5
0.0
0.5
1.0
β_IV
method
IVW
Egger
WME
WMBE
MRPRESSO

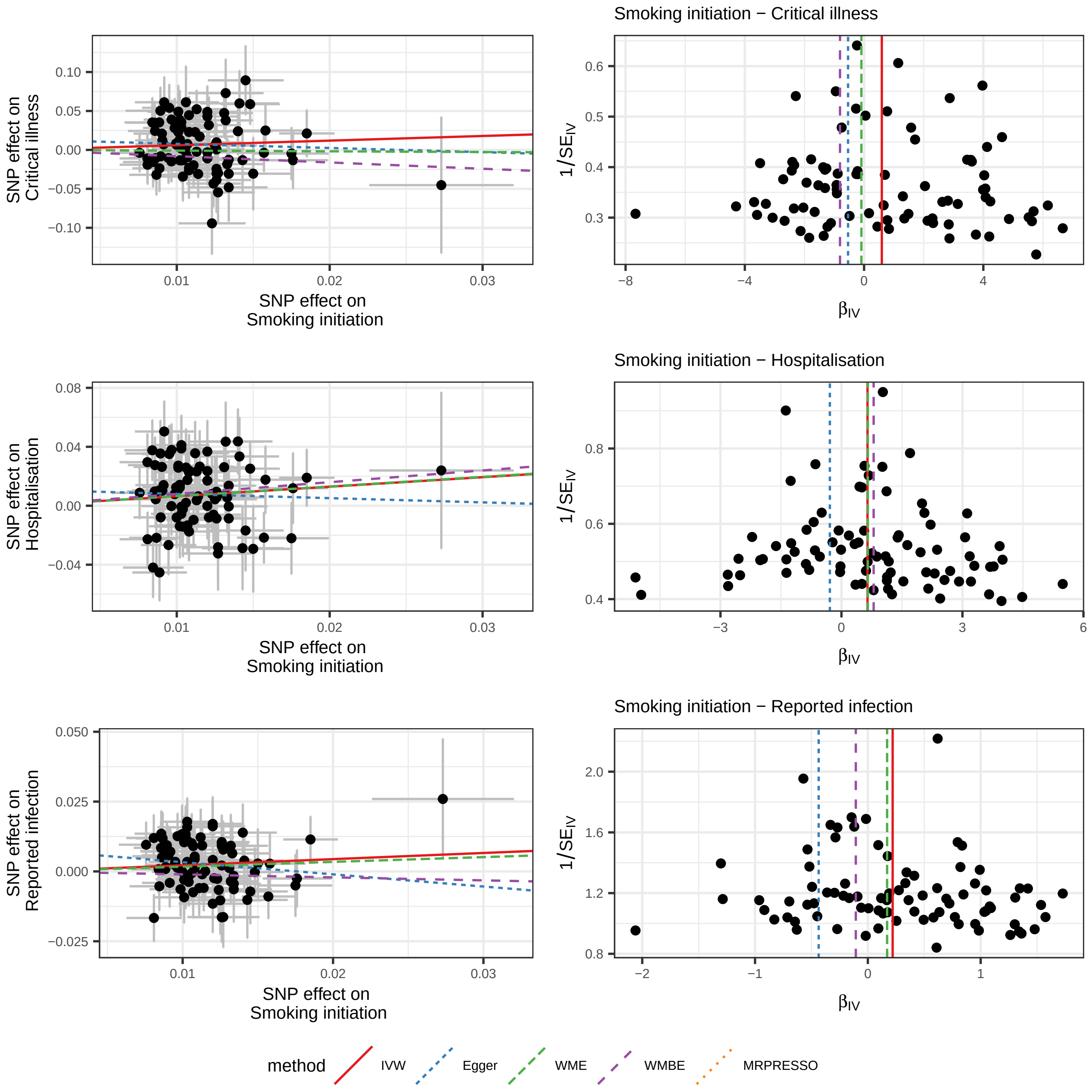
SNP effect on Critical illness
SNP effect on Smoking initiation
Smoking initiation − Critical illness
1/SE_IV
β_IV
SNP effect on Hospitalisation
SNP effect on Smoking initiation
Smoking initiation − Hospitalisation
1/SE_IV
β_IV
SNP effect on Reported infection
SNP effect on Smoking initiation
Smoking initiation − Reported infection
1/SE_IV
β_IV
method
IVW
Egger
WME
WMBE
MRPRESSO

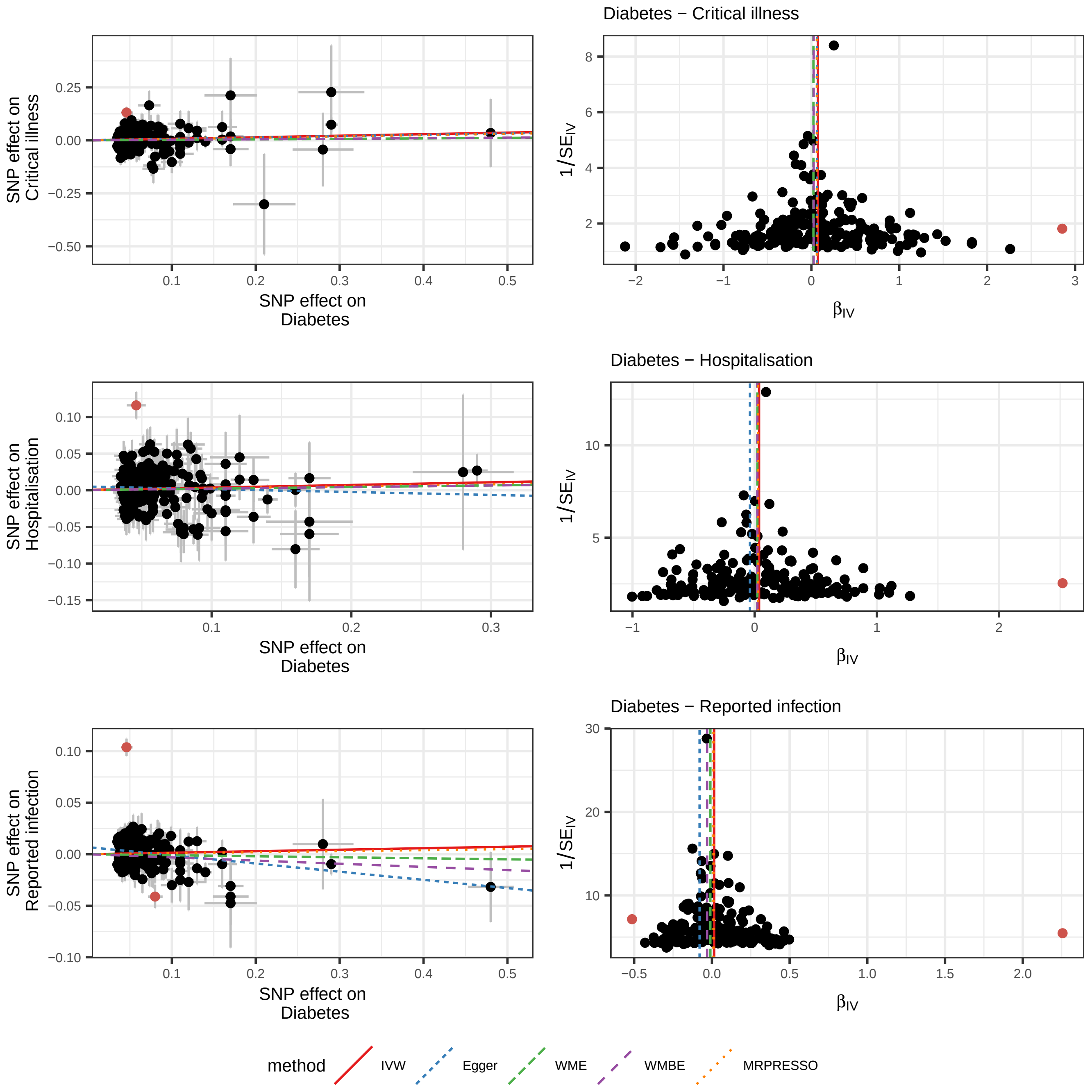
Diabetes − Critical illness
Diabetes − Hospitalisation
Diabetes − Reported infection
SNP effect on Critical illness
SNP effect on Hospitalisation
SNP effect on Reported infection
SNP effect on Diabetes
1/SE$_{IV}$
β$_{IV}$
method
IVW
Egger
WME
WMBE
MRPRESSO

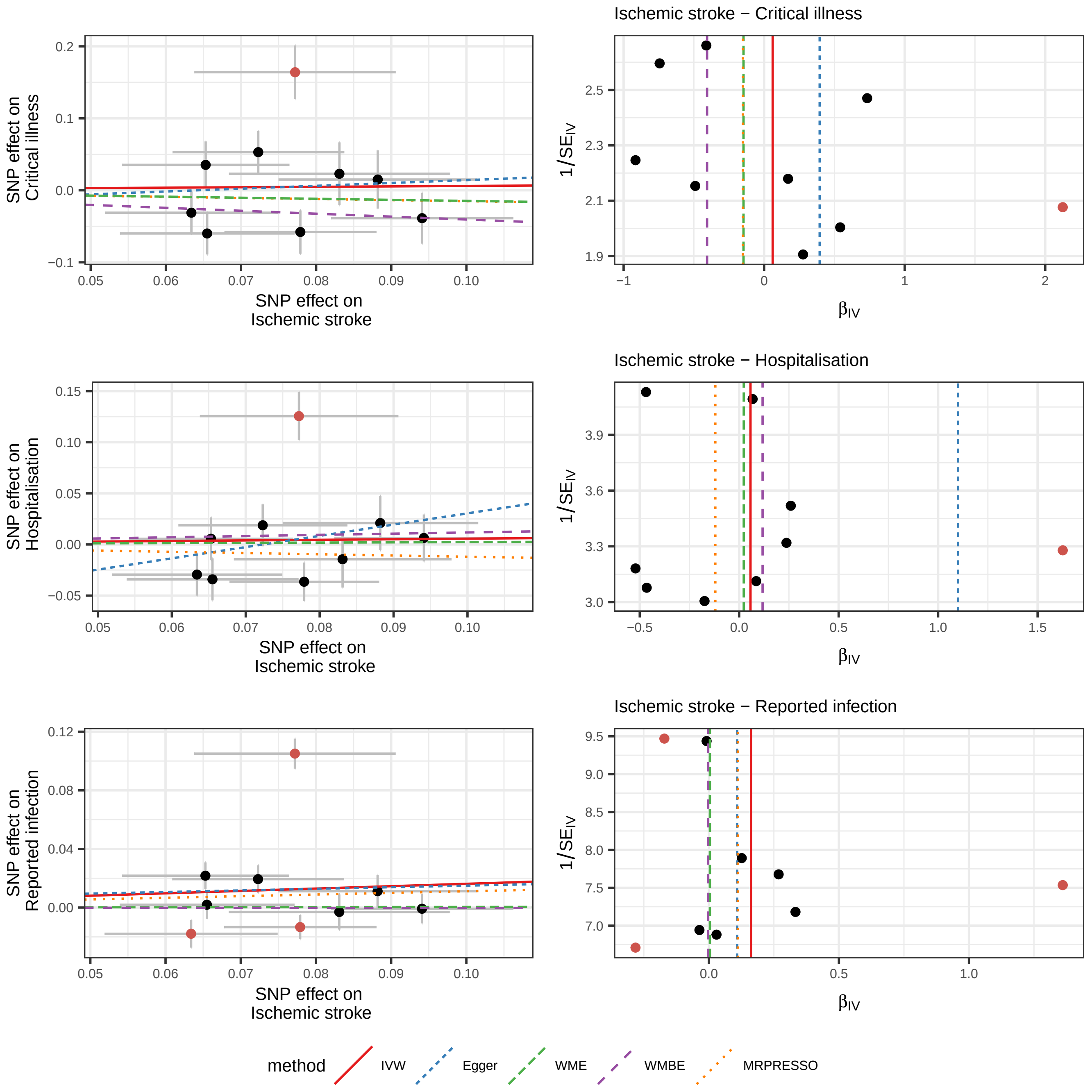

Ischemic stroke − Critical illness
Ischemic stroke − Hospitalisation
Ischemic stroke − Reported infection
SNP effect on Critical illness
SNP effect on Hospitalisation
SNP effect on Reported infection
SNP effect on Ischemic stroke
1/SE_IV
β_IV
method
IVW
Egger
WME
WMBE
MRPRESSO

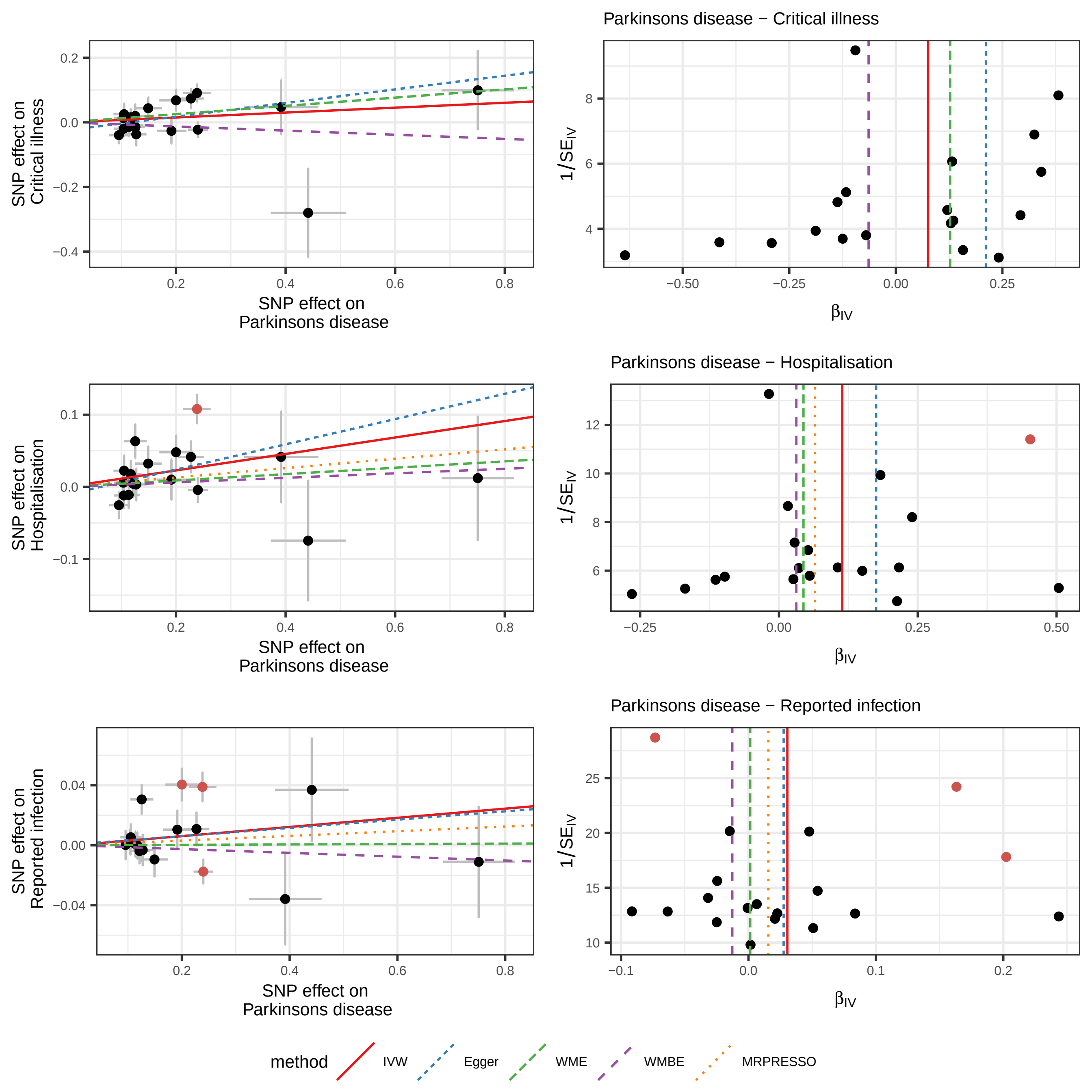

Parkinsons disease − Critical illness
SNP effect on Critical illness
SNP effect on Parkinsons disease
1/SE_IV
β_IV
Parkinsons disease − Hospitalisation
SNP effect on Hospitalisation
SNP effect on Parkinsons disease
1/SE_IV
β_IV
Parkinsons disease − Reported infection
SNP effect on Reported infection
SNP effect on Parkinsons disease
1/SE_IV
β_IV
method
IVW
Egger
WME
WMBE
MRPRESSO

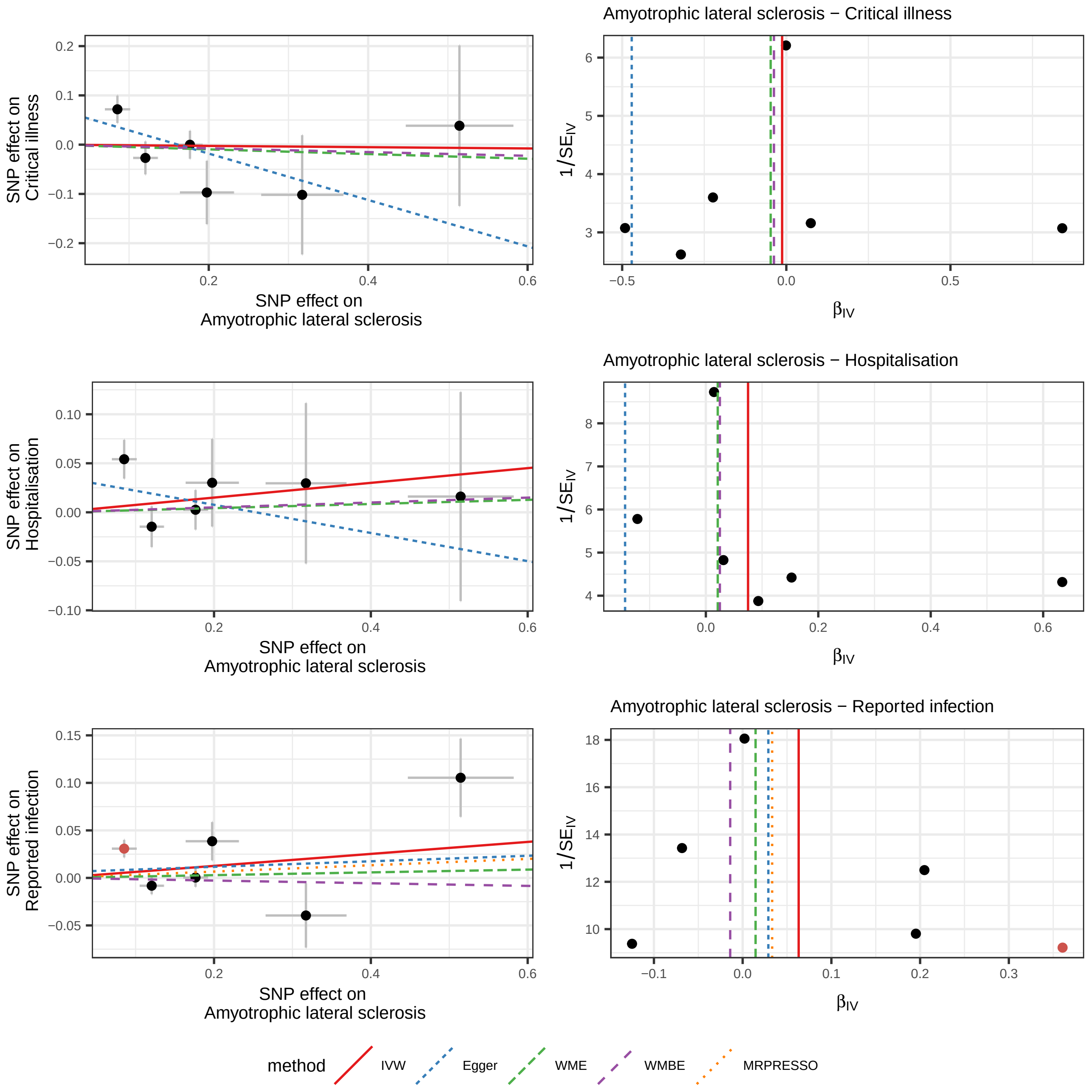
Amyotrophic lateral sclerosis − Critical illness
SNP effect on
Critical illness
SNP effect on
Amyotrophic lateral sclerosis
1/SE_IV
β_IV
0.2
0.1
0.0
−0.1
−0.2
0.4
0.6
6
5
4
3
−0.5
0.5
Amyotrophic lateral sclerosis − Hospitalisation
SNP effect on
Hospitalisation
SNP effect on
Amyotrophic lateral sclerosis
1/SE_IV
β_IV
0.10
0.05
0.00
−0.05
−0.10
8
7
Amyotrophic lateral sclerosis − Reported infection
SNP effect on
Reported infection
SNP effect on
Amyotrophic lateral sclerosis
1/SE_IV
β_IV
0.15
18
16
14
12
10
−0.1
0.3
method
IVW
Egger
WME
WMBE
MRPRESSO

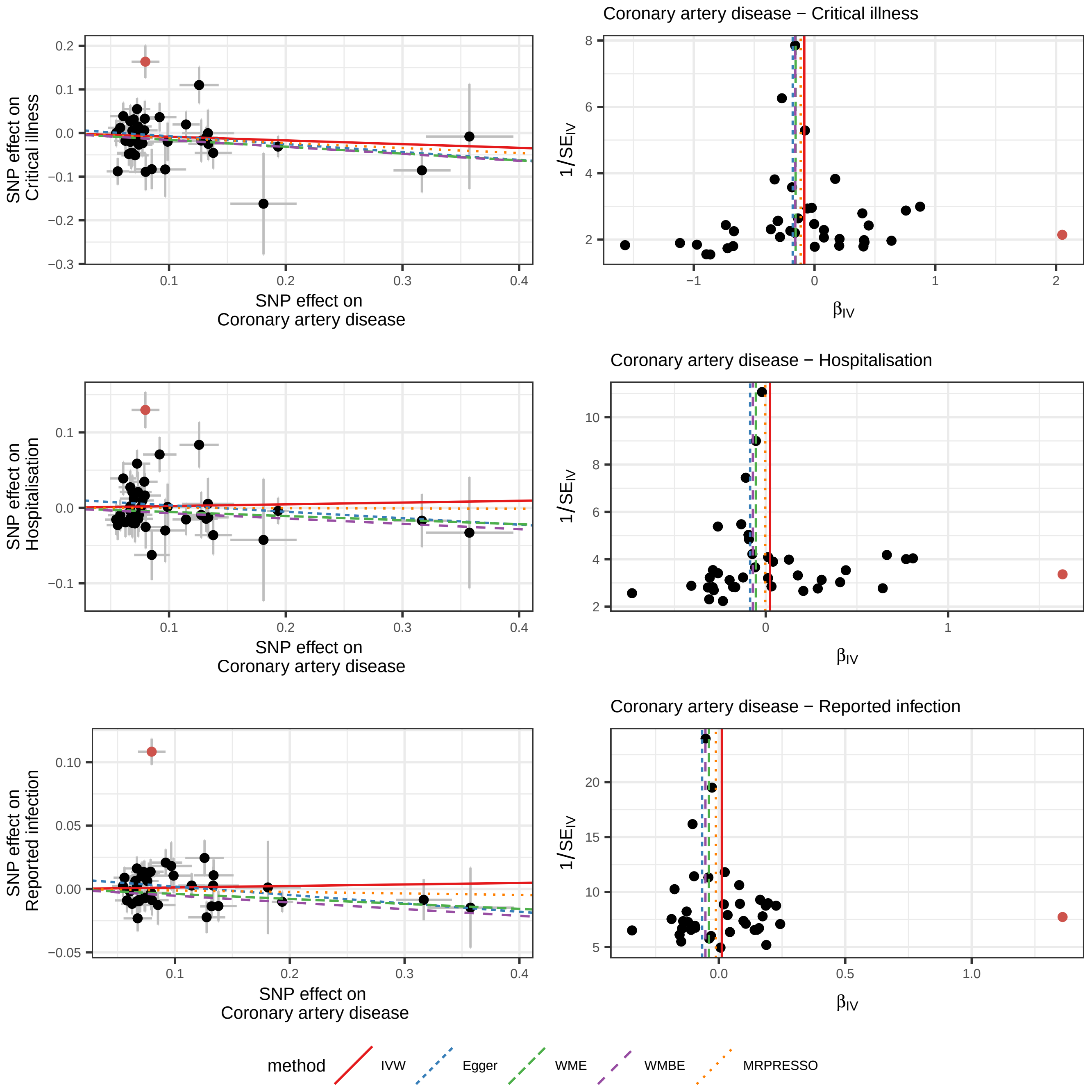

Coronary artery disease − Critical illness
SNP effect on Critical illness
SNP effect on Coronary artery disease
1/SE_IV
β_IV
Coronary artery disease − Hospitalisation
SNP effect on Hospitalisation
SNP effect on Coronary artery disease
1/SE_IV
β_IV
Coronary artery disease − Reported infection
SNP effect on Reported infection
SNP effect on Coronary artery disease
1/SE_IV
β_IV
method
IVW
Egger
WME
WMBE
MRPRESSO

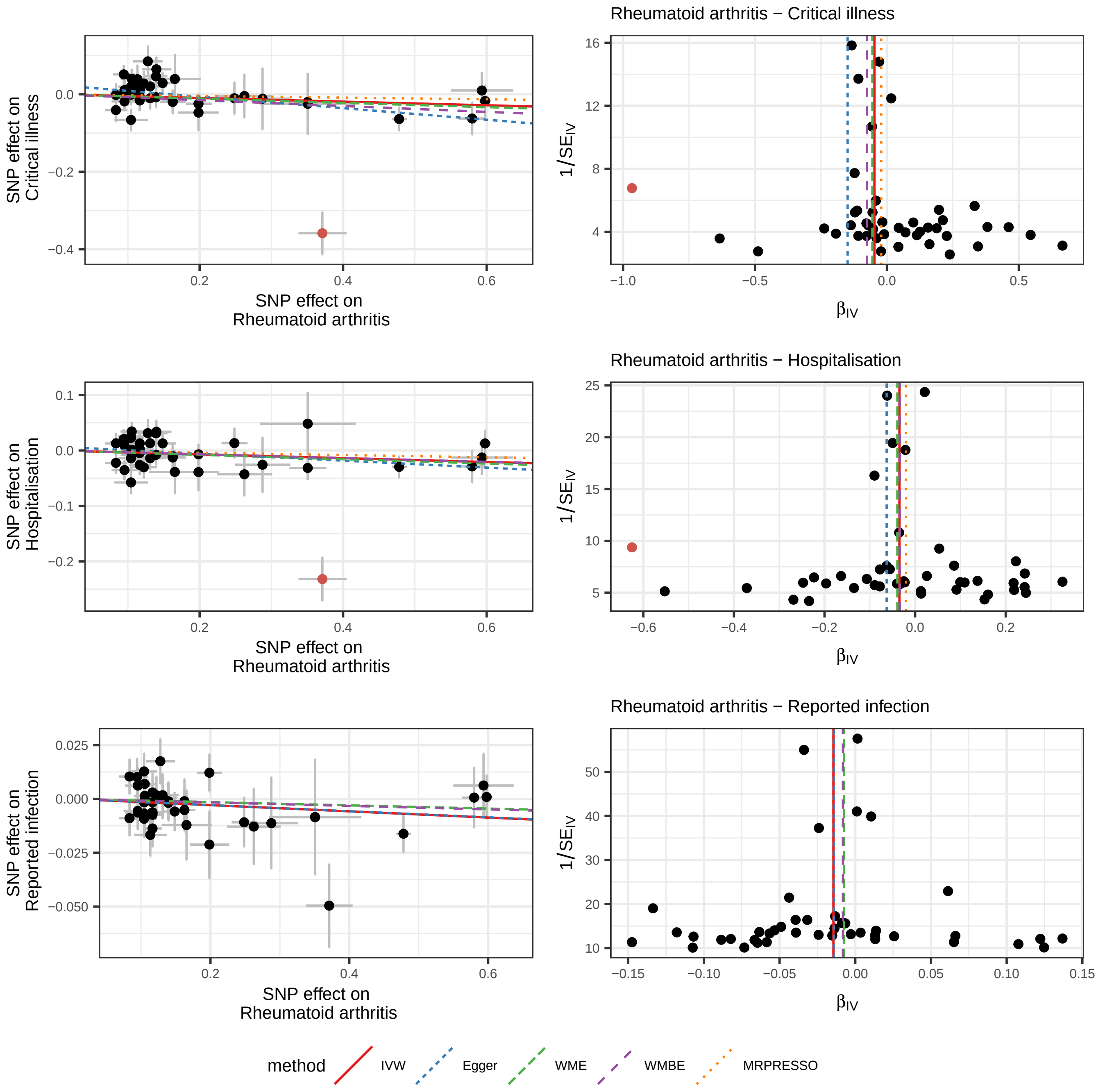
Rheumatoid arthritis − Critical illness
Rheumatoid arthritis − Hospitalisation
Rheumatoid arthritis − Reported infection
SNP effect on Critical illness
SNP effect on Hospitalisation
SNP effect on Reported infection
SNP effect on Rheumatoid arthritis
1/SE_IV
β_IV
method
IVW
Egger
WME
WMBE
MRPRESSO

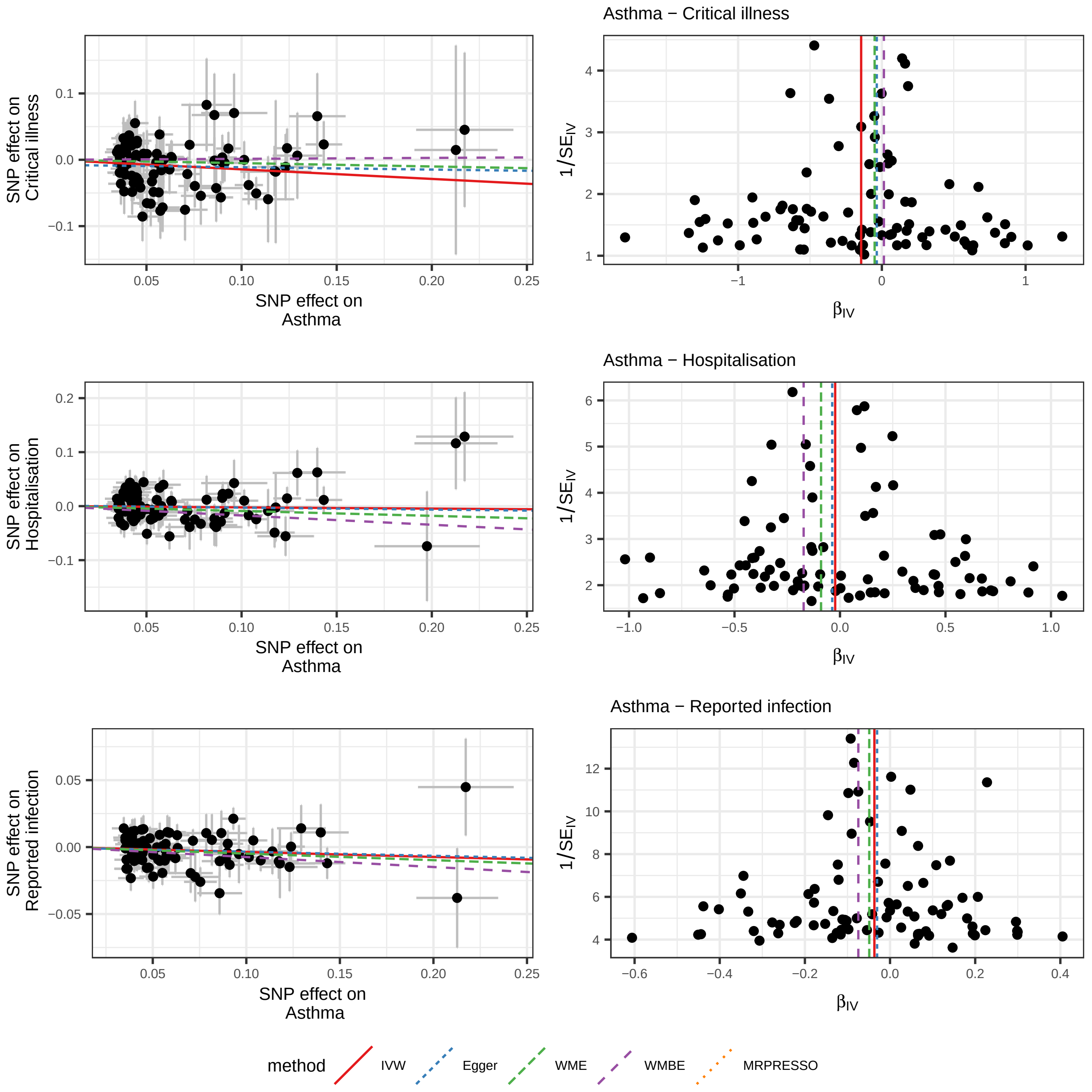

Asthma − Critical illness
SNP effect on Critical illness
SNP effect on Asthma
$1/SE_{IV}$
$\beta_{IV}$
Asthma − Hospitalisation
SNP effect on Hospitalisation
SNP effect on Asthma
$1/SE_{IV}$
$\beta_{IV}$
Asthma − Reported infection
SNP effect on Reported infection
SNP effect on Asthma
$1/SE_{IV}$
$\beta_{IV}$
method
IVW
Egger
WME
WMBE
MRPRESSO

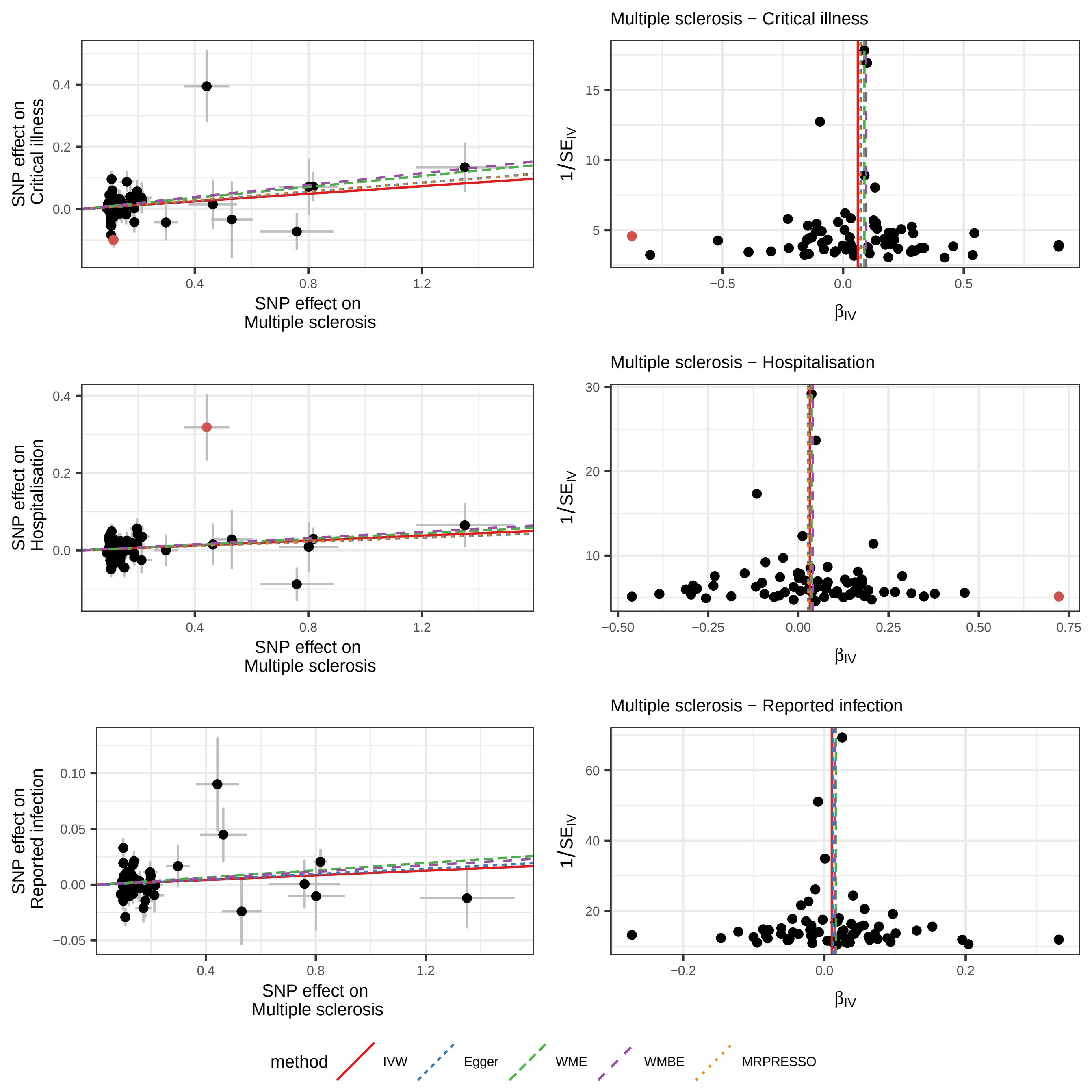

Multiple sclerosis − Critical illness
Multiple sclerosis − Hospitalisation
Multiple sclerosis − Reported infection
SNP effect on Critical illness
SNP effect on Hospitalisation
SNP effect on Reported infection
SNP effect on Multiple sclerosis
$1/SE_{IV}$
$\beta_{IV}$
method
IVW
Egger
WME
WMBE
MRPRESSO

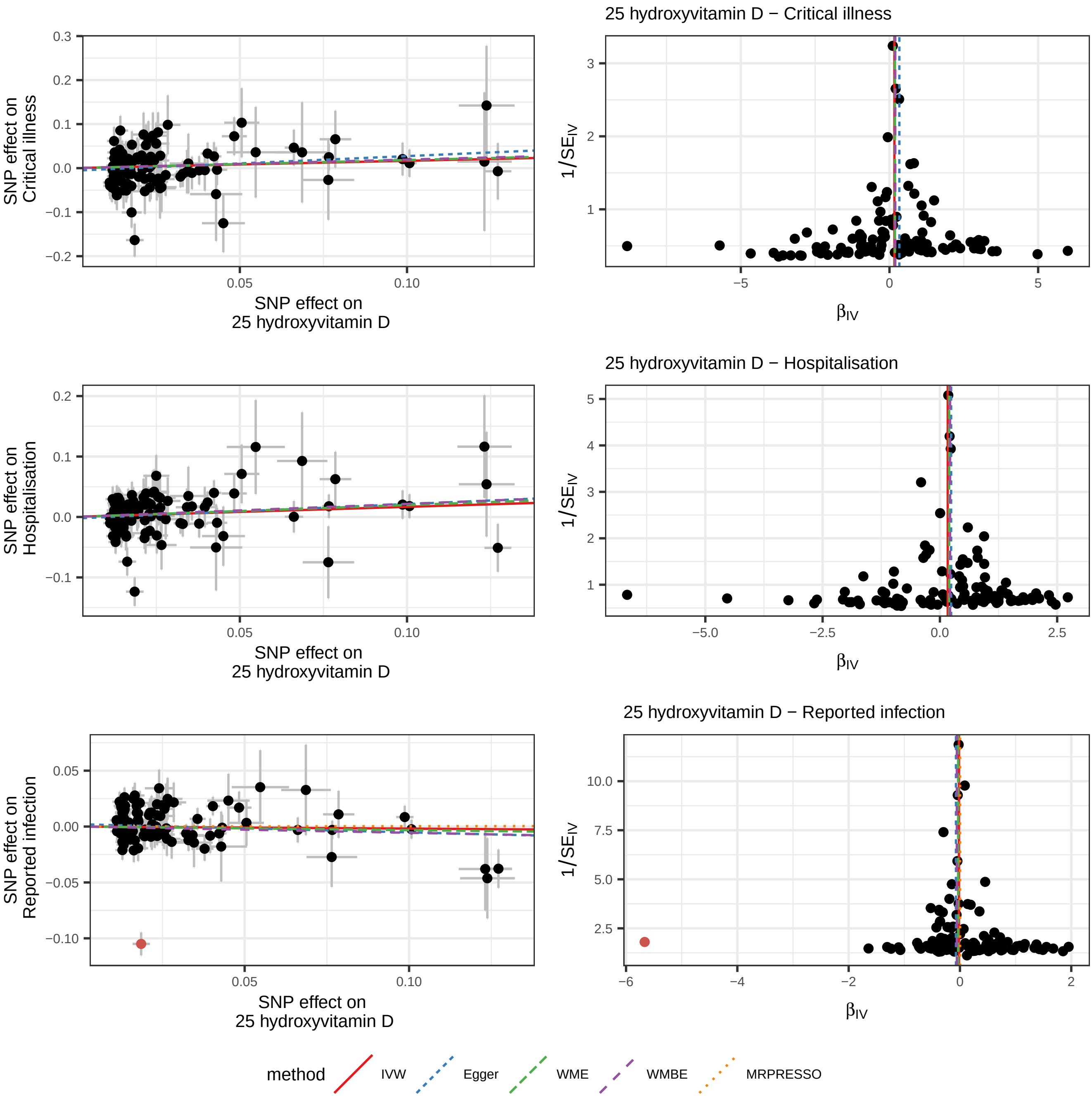
25 hydroxyvitamin D − Critical illness
SNP effect on Critical illness
SNP effect on 25 hydroxyvitamin D
1/SE_IV
β_IV
25 hydroxyvitamin D − Hospitalisation
SNP effect on Hospitalisation
SNP effect on 25 hydroxyvitamin D
1/SE_IV
β_IV
25 hydroxyvitamin D − Reported infection
SNP effect on Reported infection
SNP effect on 25 hydroxyvitamin D
1/SE_IV
β_IV
method
IVW
Egger
WME
WMBE
MRPRESSO

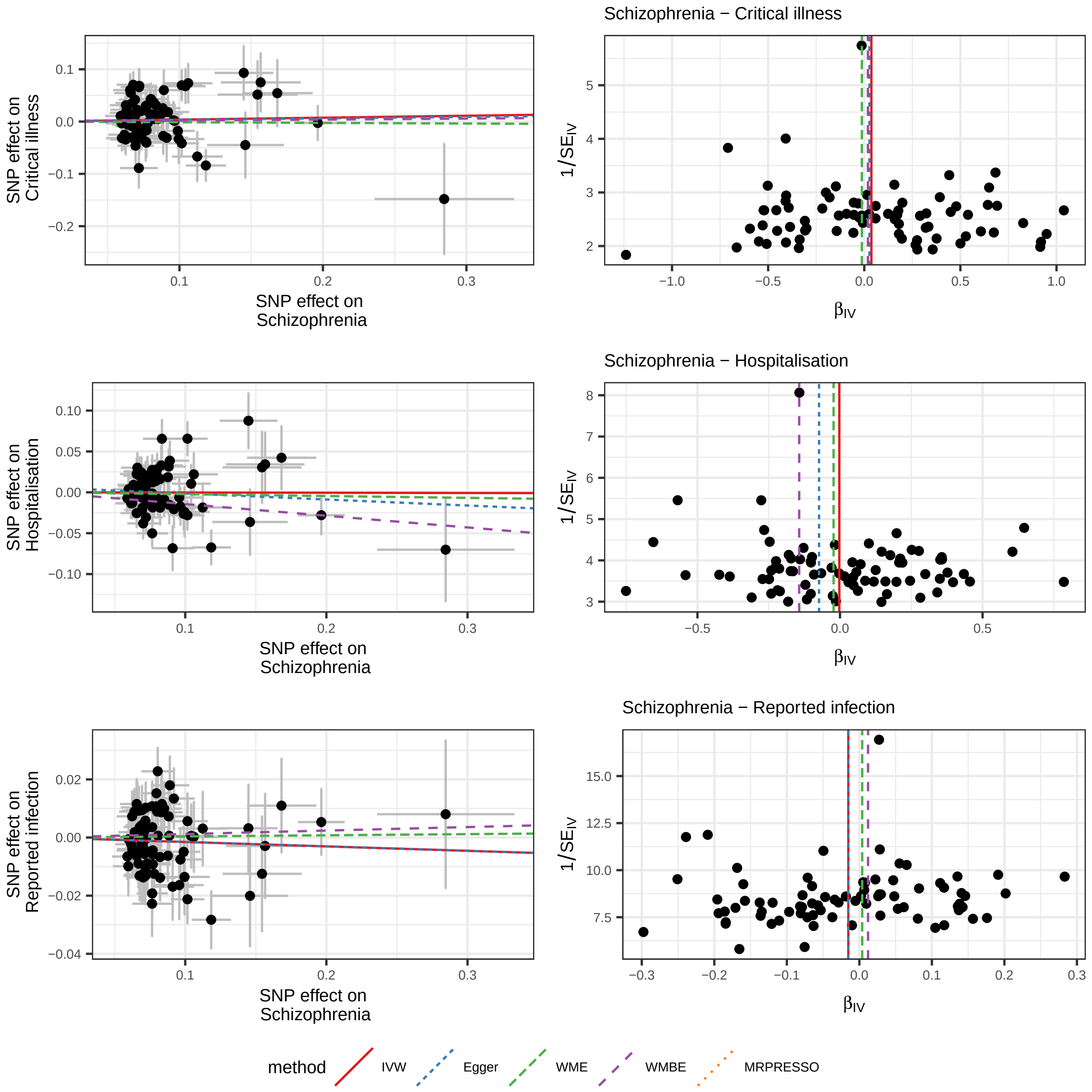

Schizophrenia − Critical illness
SNP effect on Critical illness
SNP effect on Schizophrenia
1/SE_IV
β_IV
Schizophrenia − Hospitalisation
SNP effect on Hospitalisation
SNP effect on Schizophrenia
1/SE_IV
β_IV
Schizophrenia − Reported infection
SNP effect on Reported infection
SNP effect on Schizophrenia
1/SE_IV
β_IV
method
IVW
Egger
WME
WMBE
MRPRESSO

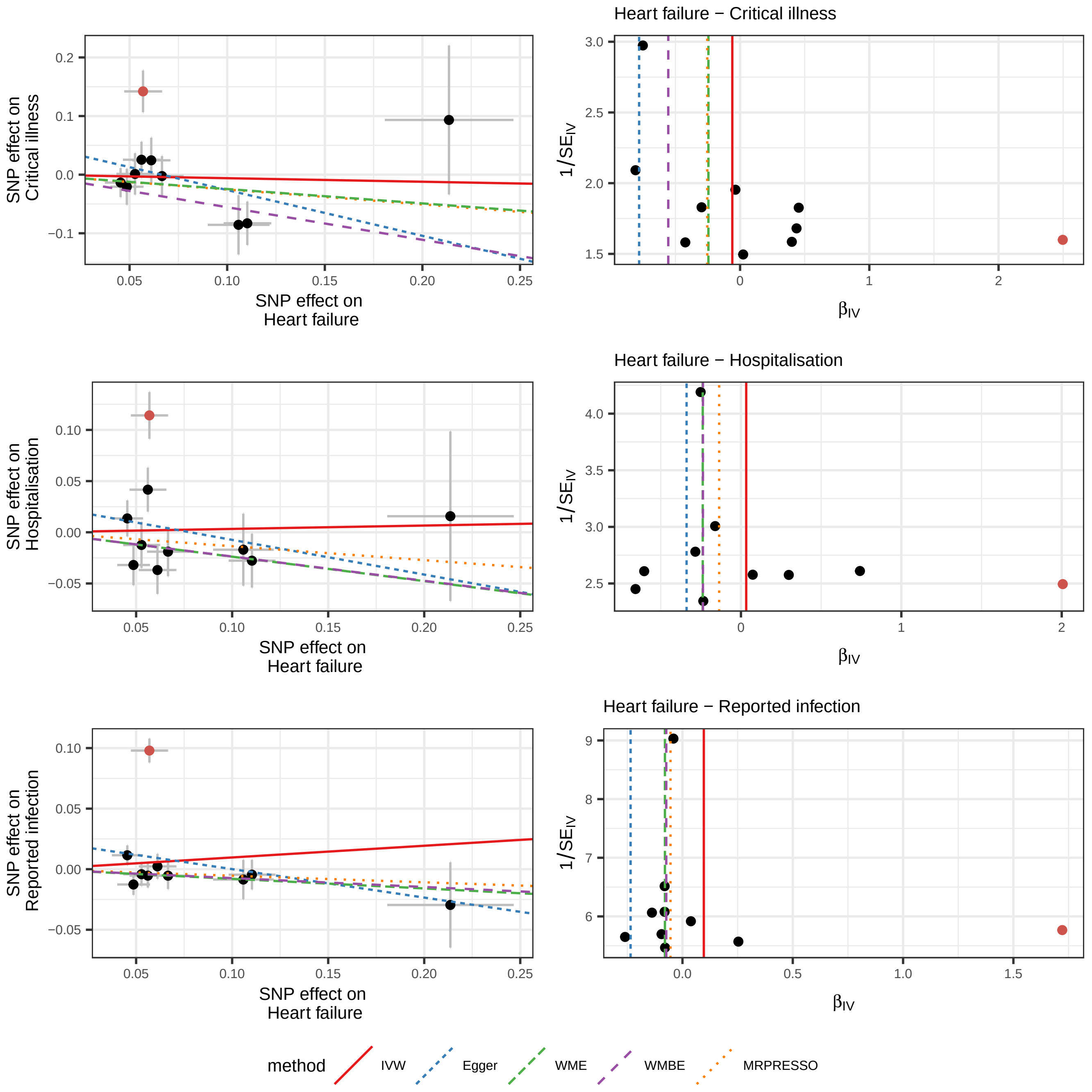

Heart failure − Critical illness
Heart failure − Hospitalisation
Heart failure − Reported infection
SNP effect on Critical illness
SNP effect on Hospitalisation
SNP effect on Reported infection
SNP effect on Heart failure
1/SE_IV
β_IV
method
IVW
Egger
WME
WMBE
MRPRESSO

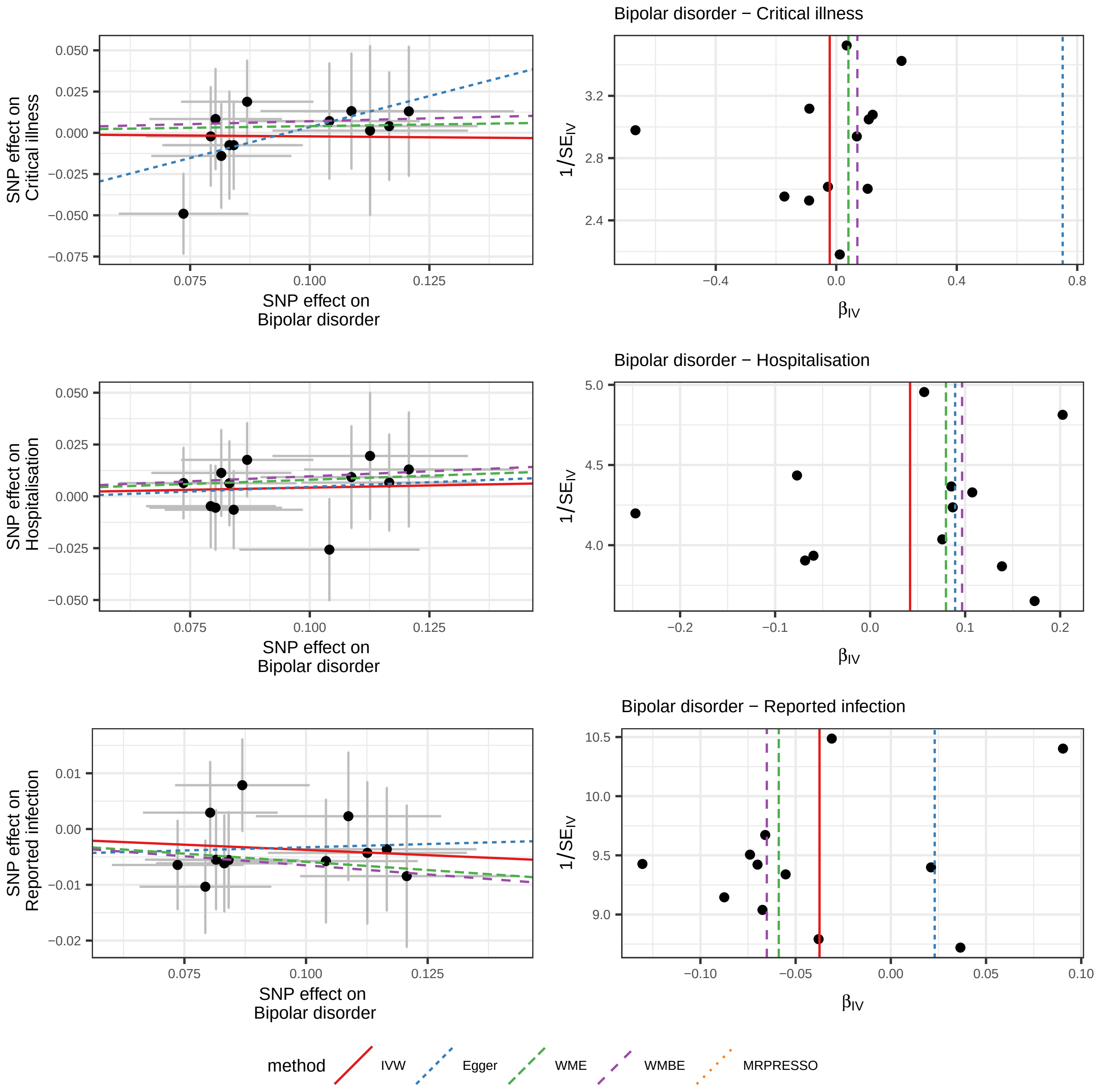

SNP effect on Critical illness
SNP effect on Bipolar disorder
Bipolar disorder − Critical illness
1/SE_IV
β_IV
SNP effect on Hospitalisation
SNP effect on Bipolar disorder
Bipolar disorder − Hospitalisation
1/SE_IV
β_IV
SNP effect on Reported infection
SNP effect on Bipolar disorder
Bipolar disorder − Reported infection
1/SE_IV
β_IV
method
IVW
Egger
WME
WMBE
MRPRESSO

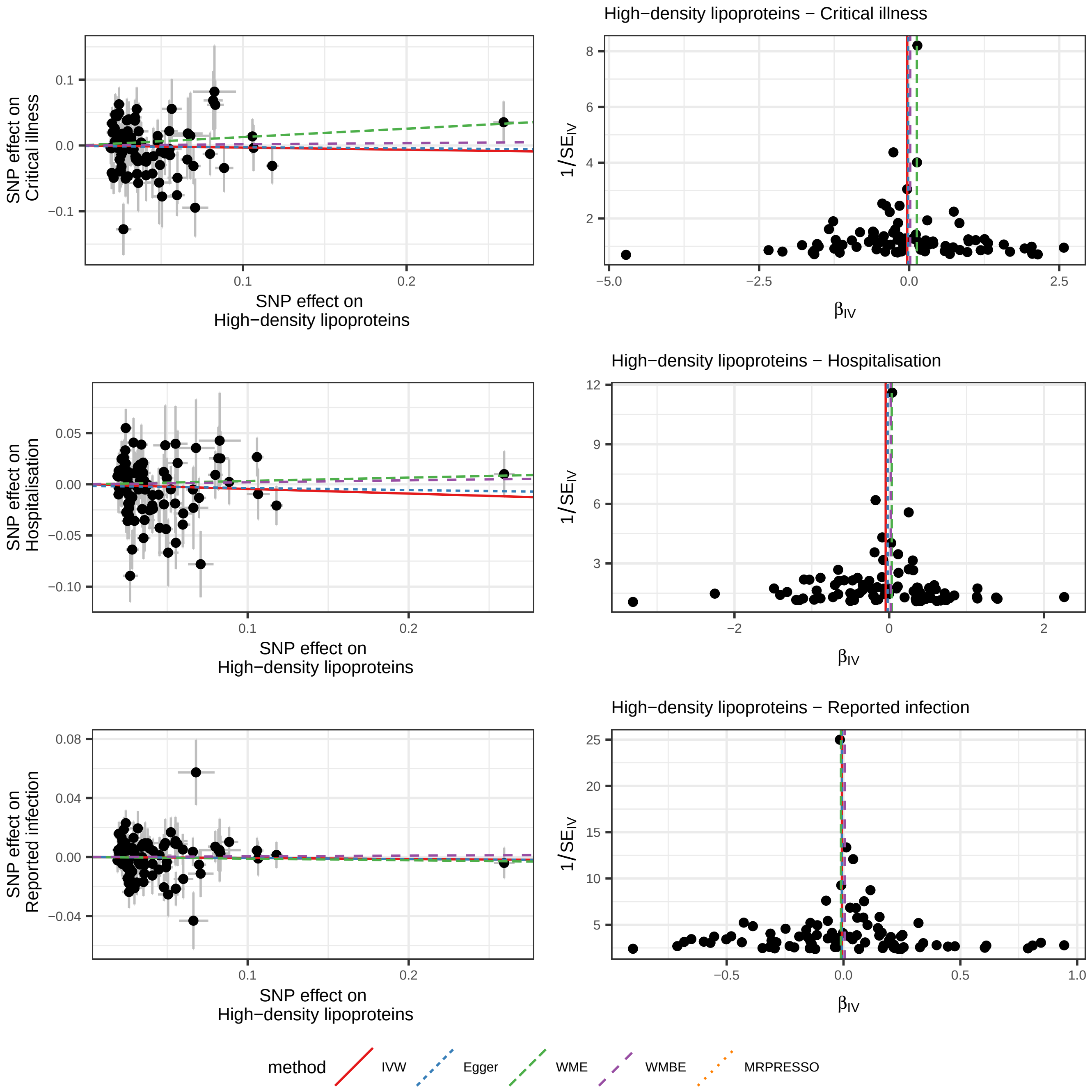
High−density lipoproteins − Critical illness
SNP effect on Critical illness
SNP effect on High−density lipoproteins
1/SE_IV
β_IV
High−density lipoproteins − Hospitalisation
SNP effect on Hospitalisation
SNP effect on High−density lipoproteins
1/SE_IV
β_IV
High−density lipoproteins − Reported infection
SNP effect on Reported infection
SNP effect on High−density lipoproteins
1/SE_IV
β_IV
method
IVW
Egger
WME
WMBE
MRPRESSO

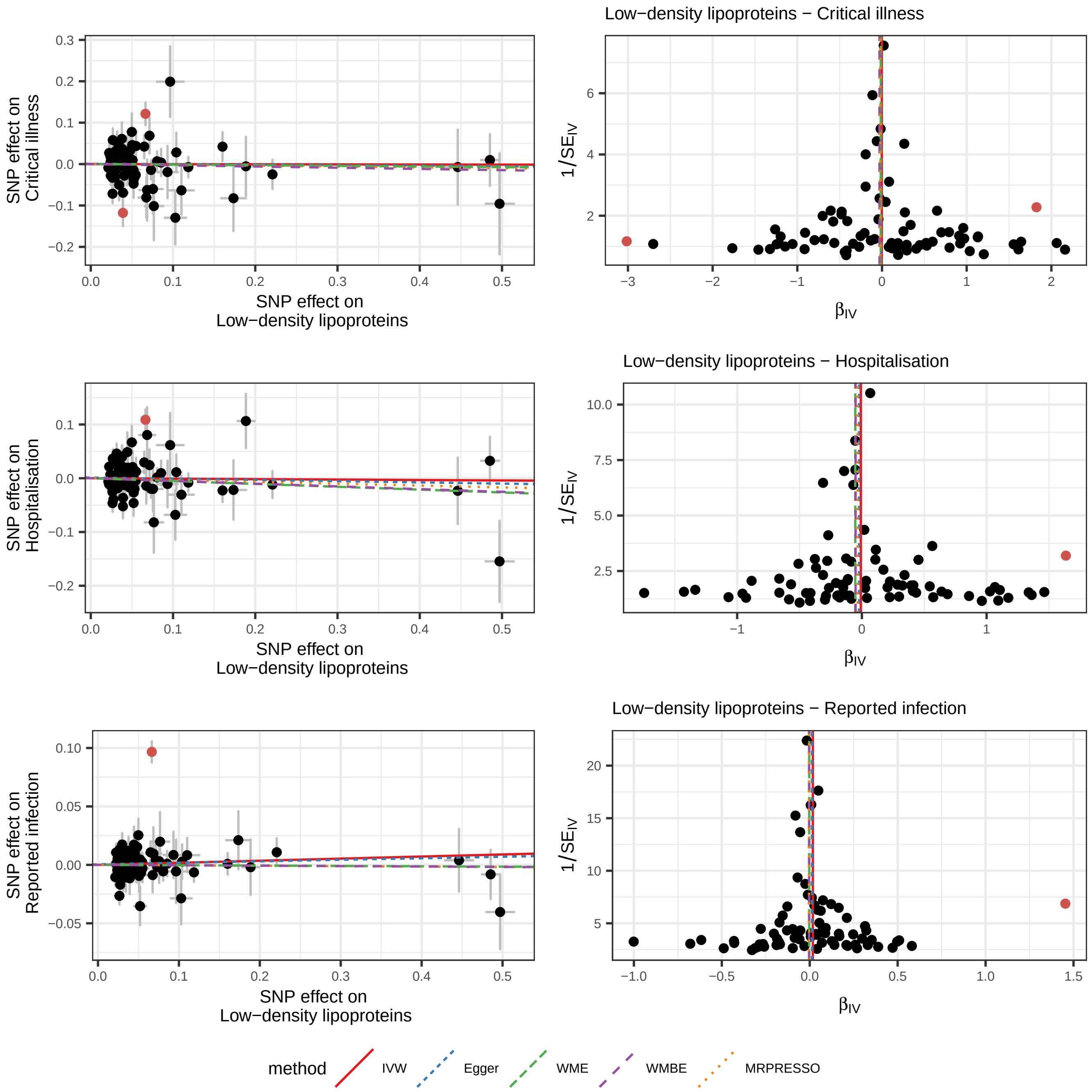
Low−density lipoproteins − Critical illness
SNP effect on
Critical illness
SNP effect on
Low−density lipoproteins
1/SE$_{IV}$
$\beta_{IV}$
Low−density lipoproteins − Hospitalisation
SNP effect on
Hospitalisation
SNP effect on
Low−density lipoproteins
1/SE$_{IV}$
$\beta_{IV}$
Low−density lipoproteins − Reported infection
SNP effect on
Reported infection
SNP effect on
Low−density lipoproteins
1/SE$_{IV}$
$\beta_{IV}$
method
IVW
Egger
WME
WMBE
MRPRESSO

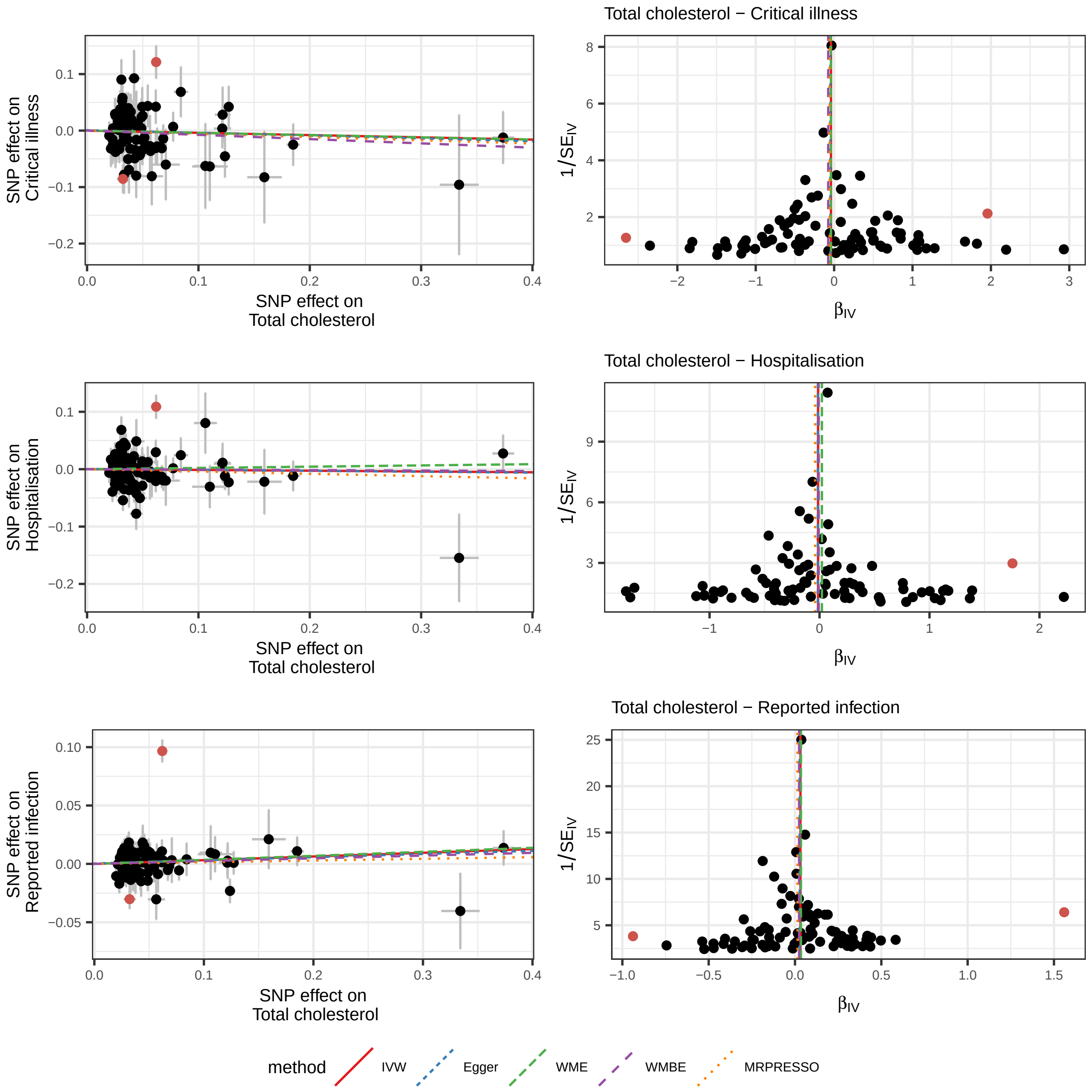

Total cholesterol − Critical illness
Total cholesterol − Hospitalisation
Total cholesterol − Reported infection
SNP effect on Critical illness
SNP effect on Hospitalisation
SNP effect on Reported infection
SNP effect on Total cholesterol
1/SE$_{IV}$
β$_{IV}$
method
IVW
Egger
WME
WMBE
MRPRESSO

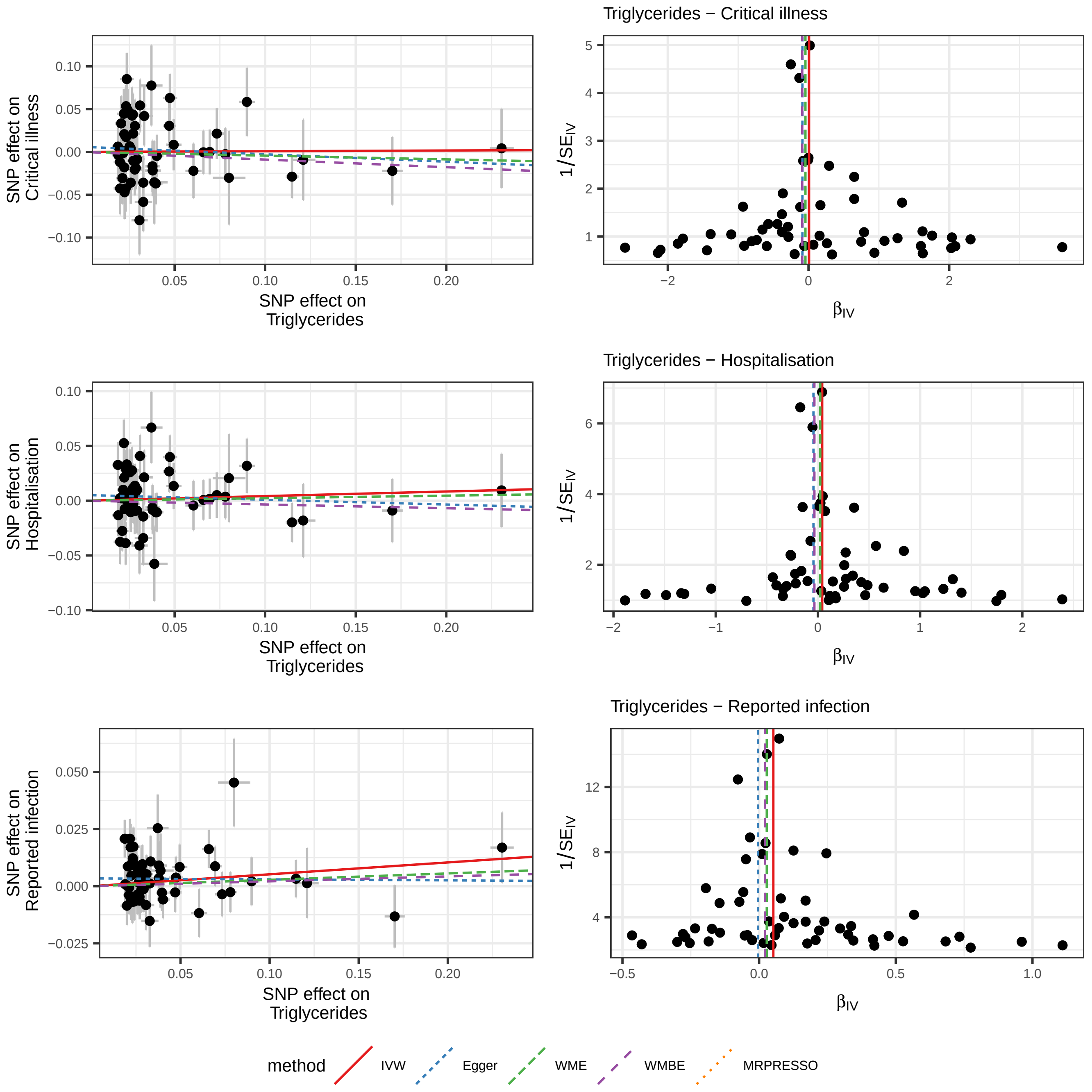

Triglycerides − Critical illness
Triglycerides − Hospitalisation
Triglycerides − Reported infection
SNP effect on Critical illness
SNP effect on Hospitalisation
SNP effect on Reported infection
SNP effect on Triglycerides
1/SE_IV
β_IV
method
IVW
Egger
WME
WMBE
MRPRESSO

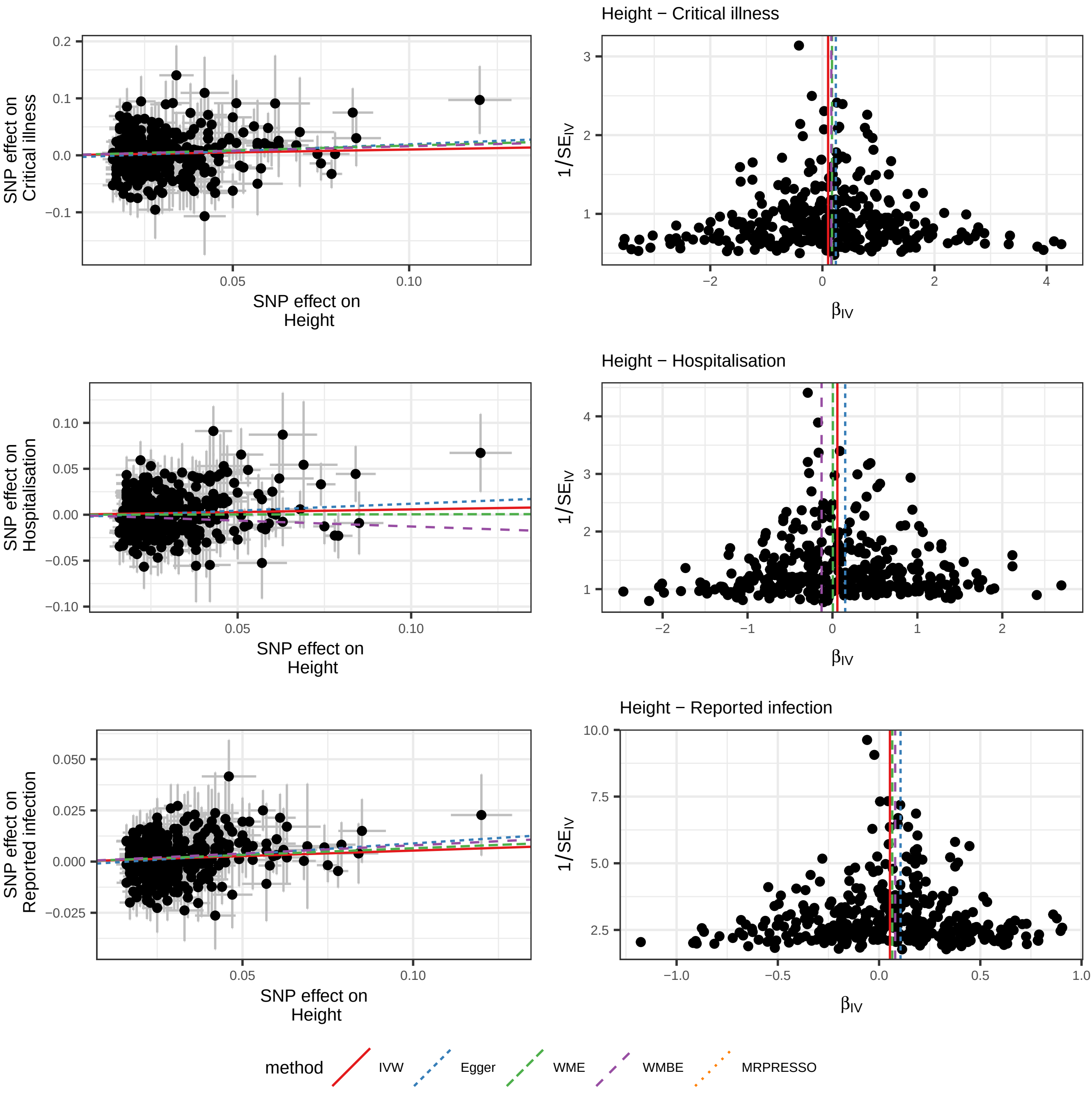

Height − Critical illness
SNP effect on Critical illness
SNP effect on Height
$1/SE_{IV}$
$\beta_{IV}$
Height − Hospitalisation
SNP effect on Hospitalisation
SNP effect on Height
$1/SE_{IV}$
$\beta_{IV}$
Height − Reported infection
SNP effect on Reported infection
SNP effect on Height
$1/SE_{IV}$
$\beta_{IV}$
method
IVW
Egger
WME
WMBE
MRPRESSO

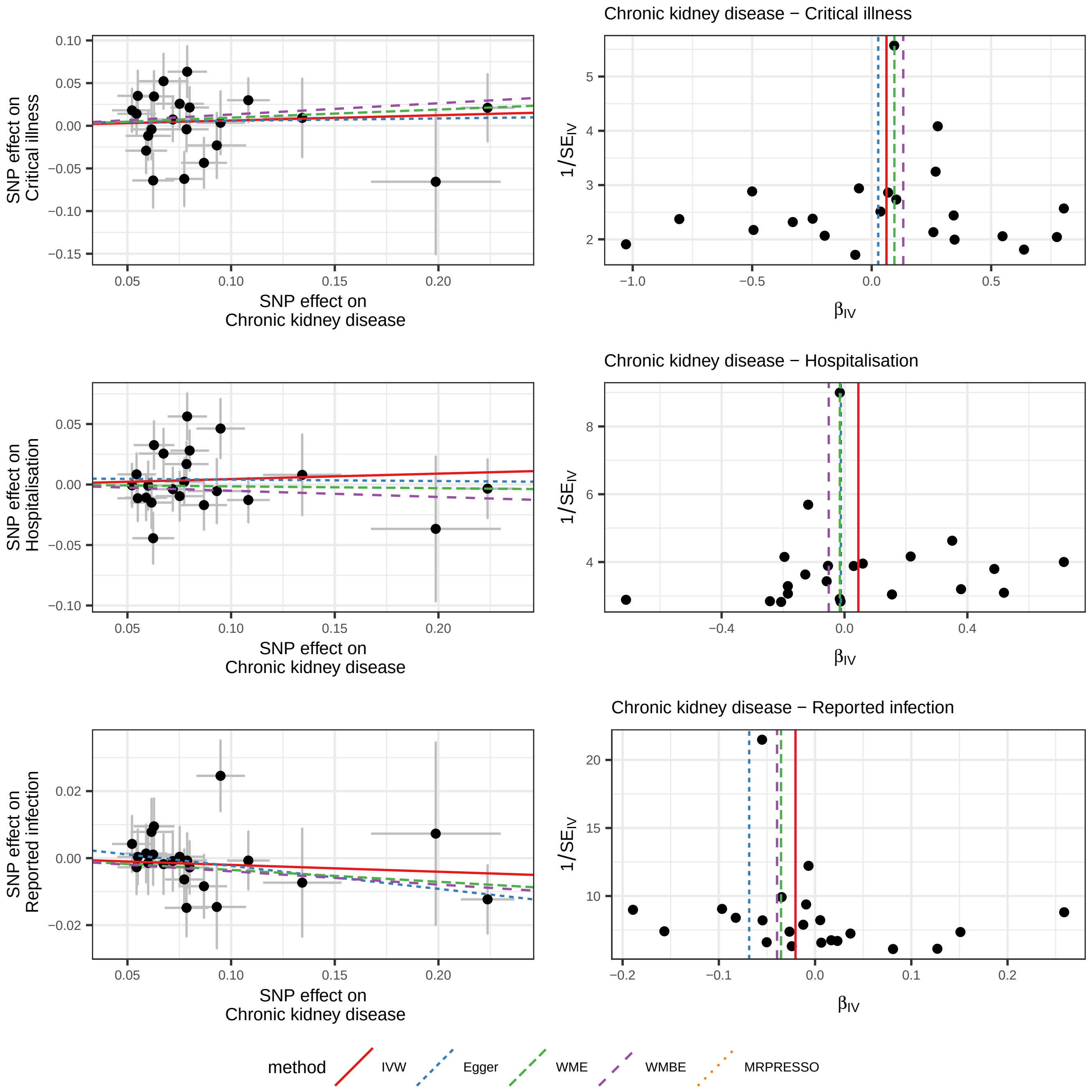

Chronic kidney disease − Critical illness
Chronic kidney disease − Hospitalisation
Chronic kidney disease − Reported infection
SNP effect on Critical illness
SNP effect on Hospitalisation
SNP effect on Reported infection
SNP effect on Chronic kidney disease
$1/SE_{IV}$
$\beta_{IV}$
method
IVW
Egger
WME
WMBE
MRPRESSO

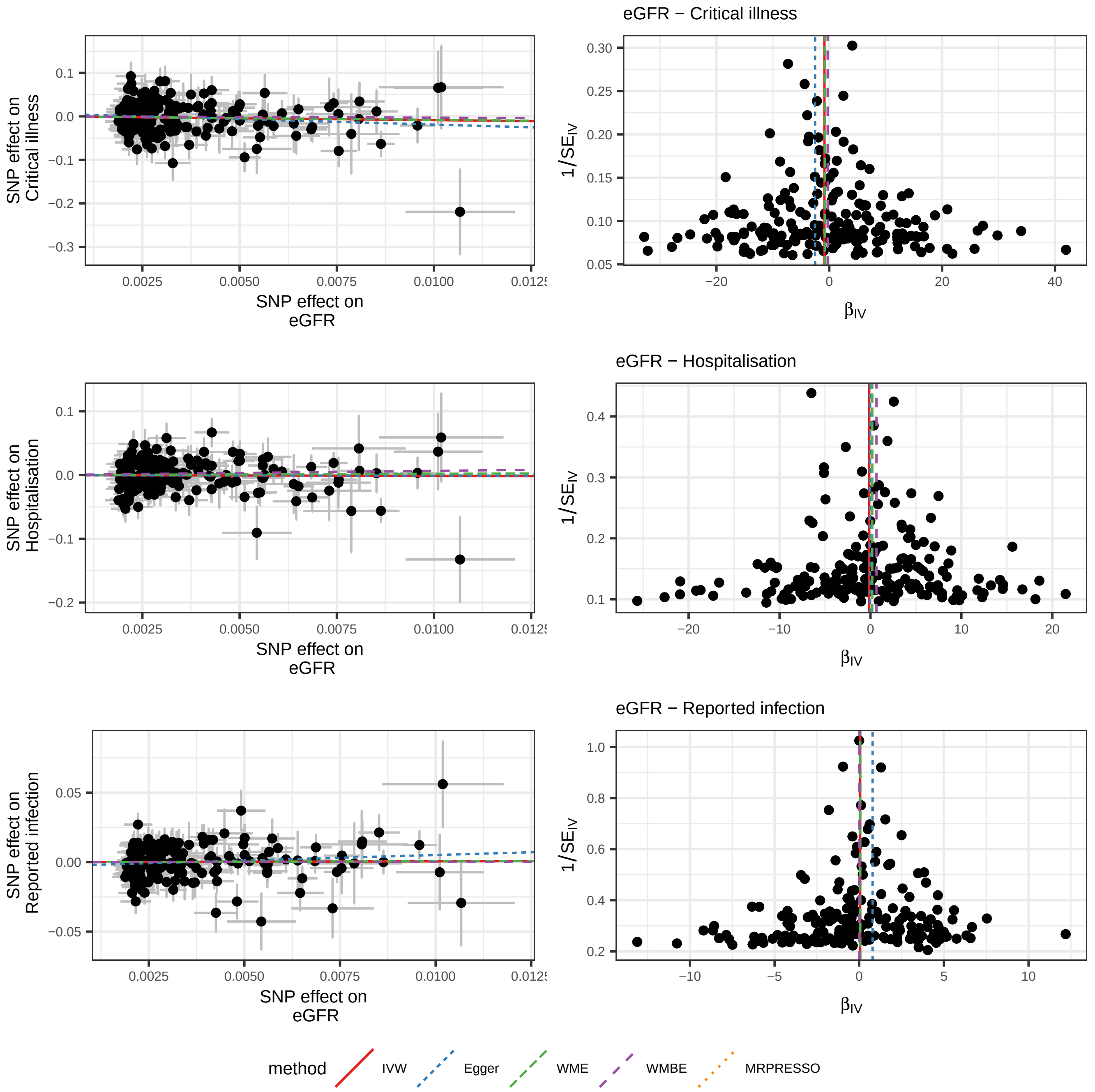

eGFR − Critical illness
eGFR − Hospitalisation
eGFR − Reported infection
SNP effect on Critical illness
SNP effect on Hospitalisation
SNP effect on Reported infection
SNP effect on eGFR
1/SE_IV
β_IV
method
IVW
Egger
WME
WMBE
MRPRESSO

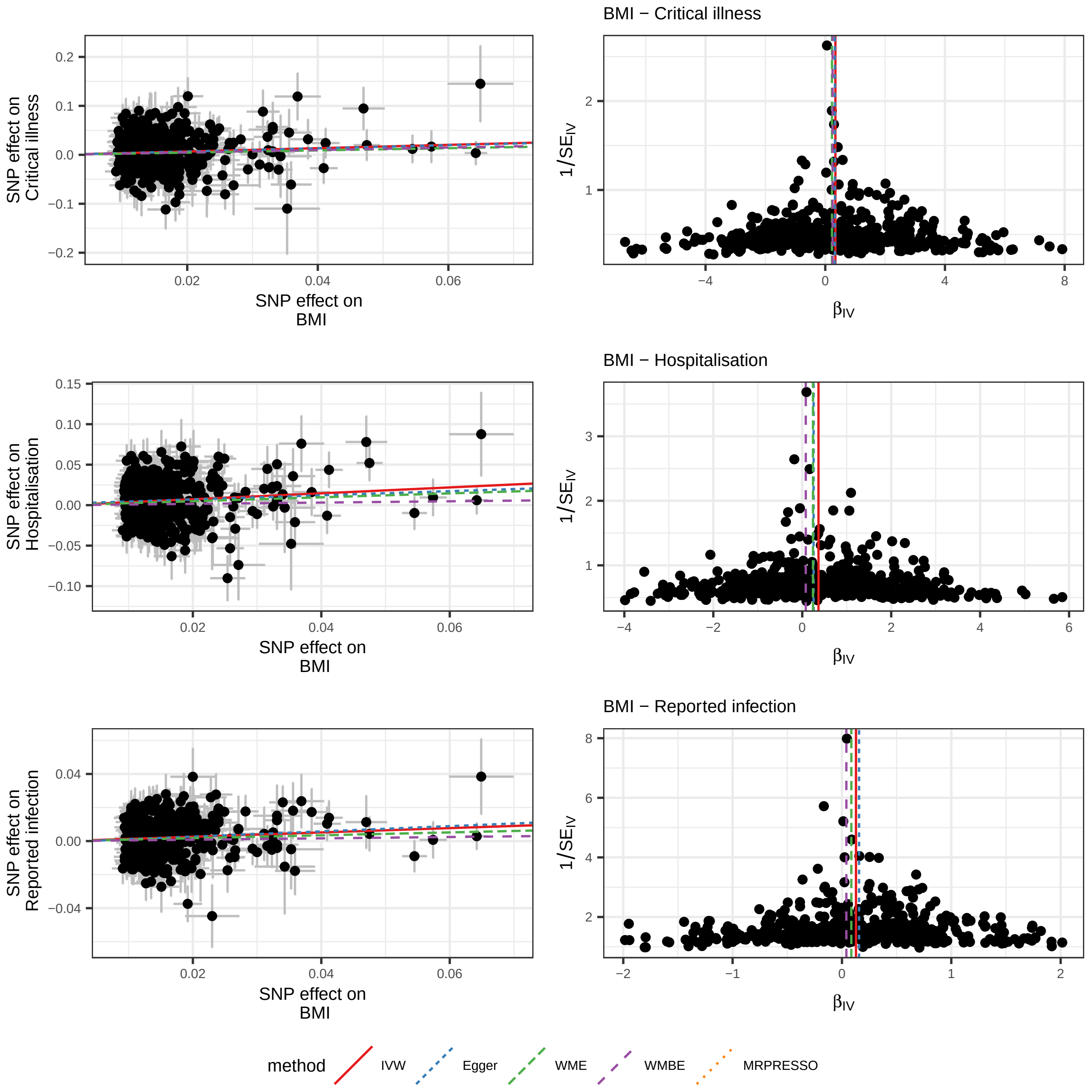

BMI − Critical illness
SNP effect on Critical illness
SNP effect on BMI
1/SE$_{IV}$
$\beta_{IV}$
BMI − Hospitalisation
SNP effect on Hospitalisation
SNP effect on BMI
1/SE$_{IV}$
$\beta_{IV}$
BMI − Reported infection
SNP effect on Reported infection
SNP effect on BMI
1/SE$_{IV}$
$\beta_{IV}$
method
IVW
Egger
WME
WMBE
MRPRESSO
