## Extended data and supplement for "Mapping the human genetic architecture of COVID-19 by worldwide meta-analysis": Extended_data_figures_and_legends.pdf

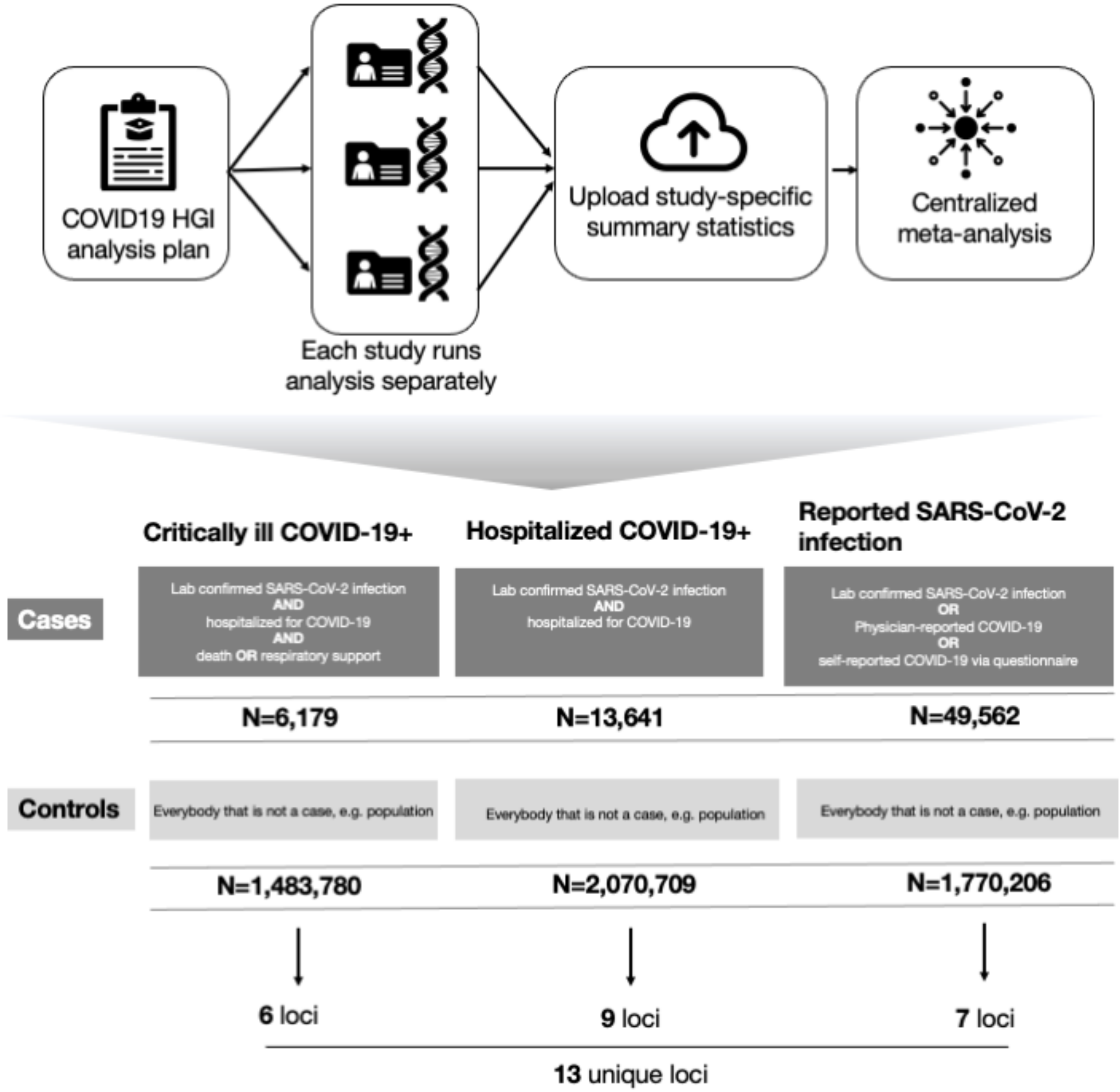


**New Extended Data Figure 1**. **Analytical summary of the COVID-19 HGI worldwide meta-analysis.** Using the analytical plan set by the COVID-19 HGI, each individual study runs their analyses and uploads the results to the Initiative, who then runs the meta-analysis. There are three main analyses that each study can contribute summary statistics to; critically ill COVID-19, hospitalized COVID-19 and reported SARS-CoV-2 infection. The phenotypic criteria used to define cases are listed in the dark grey boxes, along with the numbers of cases (N) included in the final all ancestries meta-analysis. Controls were defined in the same way across all three analyses; as everybody that is not a case e.g. population controls (light grey box). Sensitivity analyses, not reported in this Figure, also used mild/asymptomatic

COVID-19 cases as controls. Sample number (N) of controls differed between the analyses due to the difference in number of studies contributing data to these.

*Please find the corresponding figure in the PDF "Extended Data Figure 2"*

**Extended Data Figure 2. QQ-plots from contributing studies.**

QQ-plots showing the expected $-\log_{10}$(P-values) on the x-axis and the observed values on the y-axis (red line showing no deviation from the expected) for each study contributing data to the analyses. Sample size of cases and controls is listed for each study in the plot title, as well as the median lambda value.

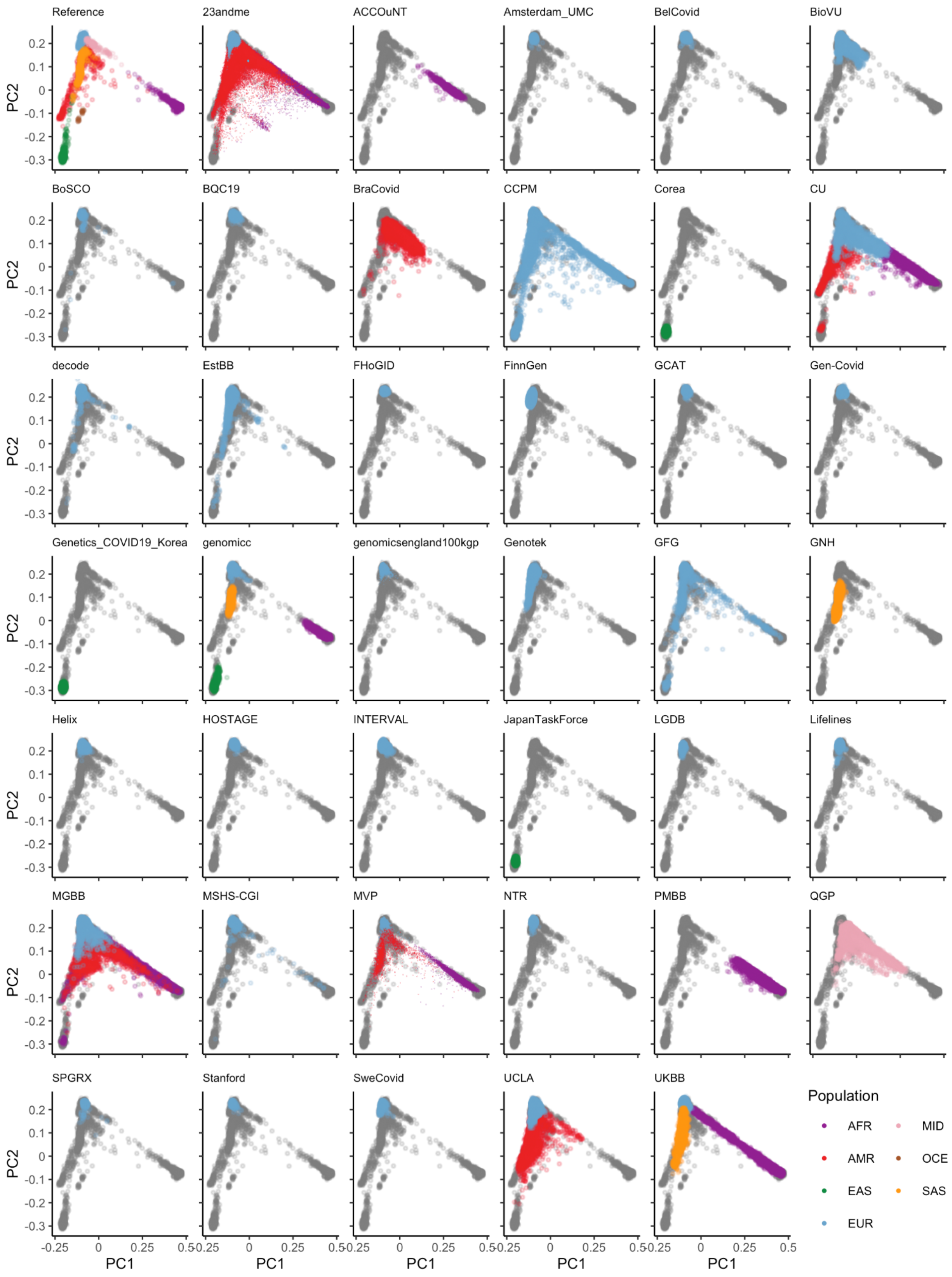


**Extended Data Figure 3. Projection of contributing studies samples into the same PC space.**

We asked participating to studies to perform PC projection using the 1000 Genomes Project and Human Genome Diversity Project as a reference, with a common set of variants. For each panel (except for the reference), colored points correspond to participated samples from each cohort, whereas gray points correspond to reference samples. Color represents a genetic population that each cohort specified. Since 23andme, genomicsengland100kgp, and MVP only submitted PCA images, we overlaid their submitted transparent images using the same coordinates, instead of directly plotting them.

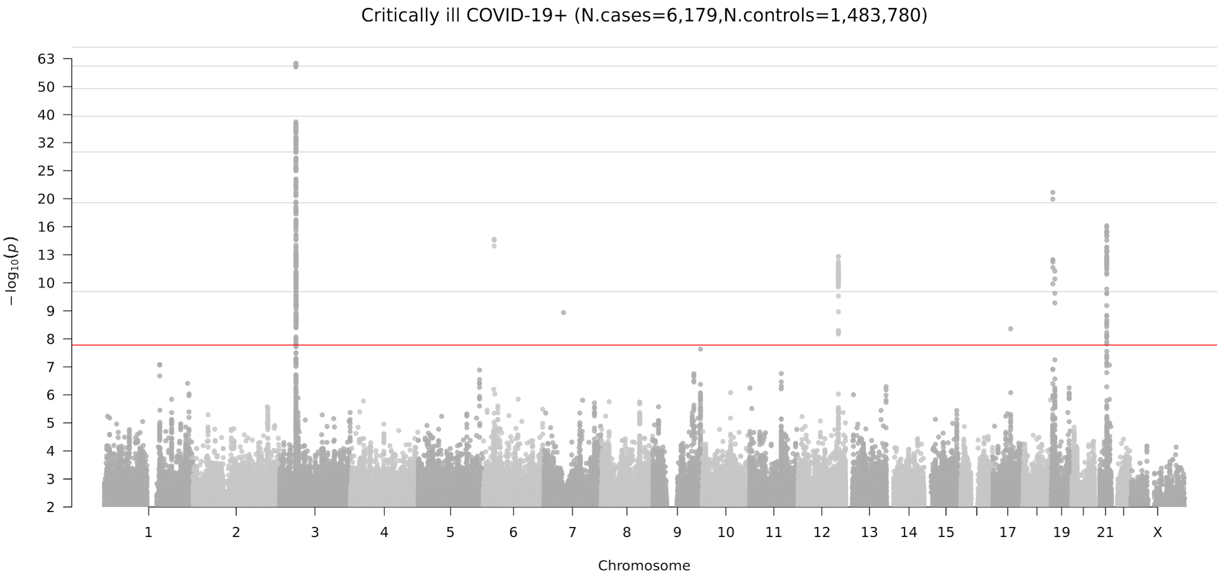


**Extended Data Figure 4**. **Genome-wide meta-analysis association results for critical illness due to COVID-19**.

The locus on chromosome 6 is the HLA locus, which was removed from the list of reported loci in Table 1 due to the high heterogeneity in effect size estimated between studies included in the analysis. The locus on chromosome 7 was also not reported in Table 1 due to missingness across studies, i.e. the high number of studies in the meta-analysis that did not report summary statistics for this region. There are two association peaks on chromosome 19.

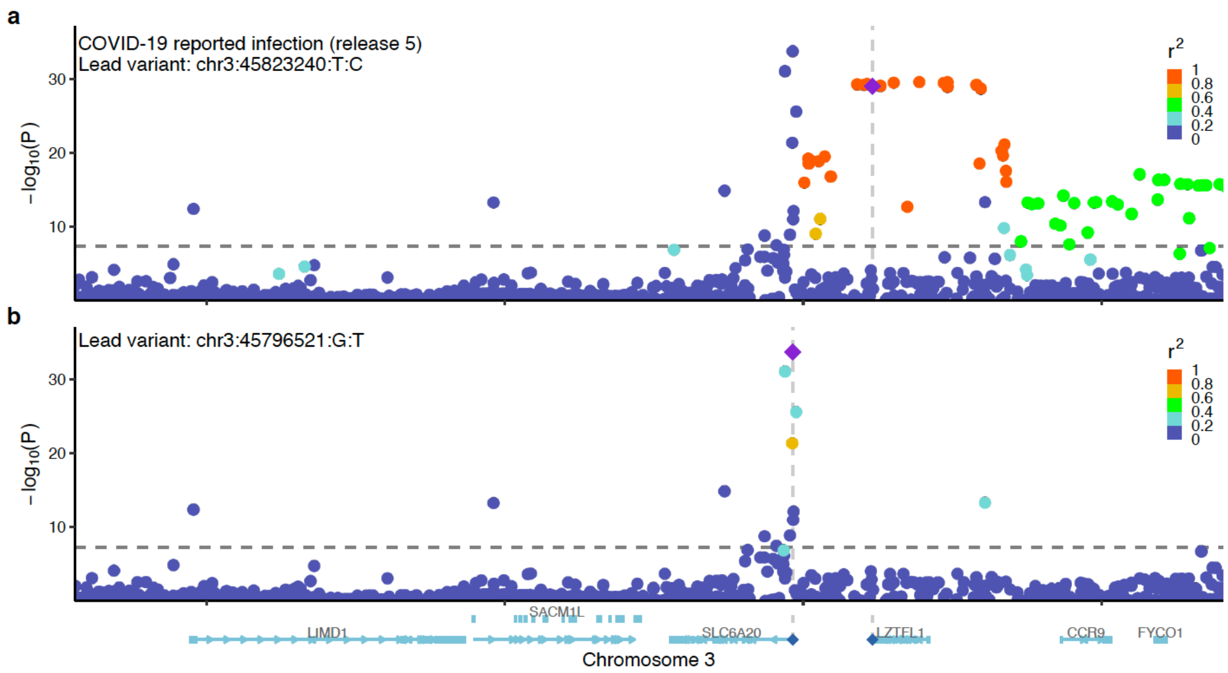


**Extended Data Figure 5. Locuszoom plots of the 3p21.31 region for reported infection.**
a. A standard plot without exclusion. Here, the severity lead variant rs10490770 (chr3:45823240:T:C) is shown as a lead variant. b. Additional independent susceptibility signal(s) after excluding variants with r2 > 0.05 with rs10490770. The susceptibility lead variant rs2271616 (chr3:45796521:G:T) is highlighted.

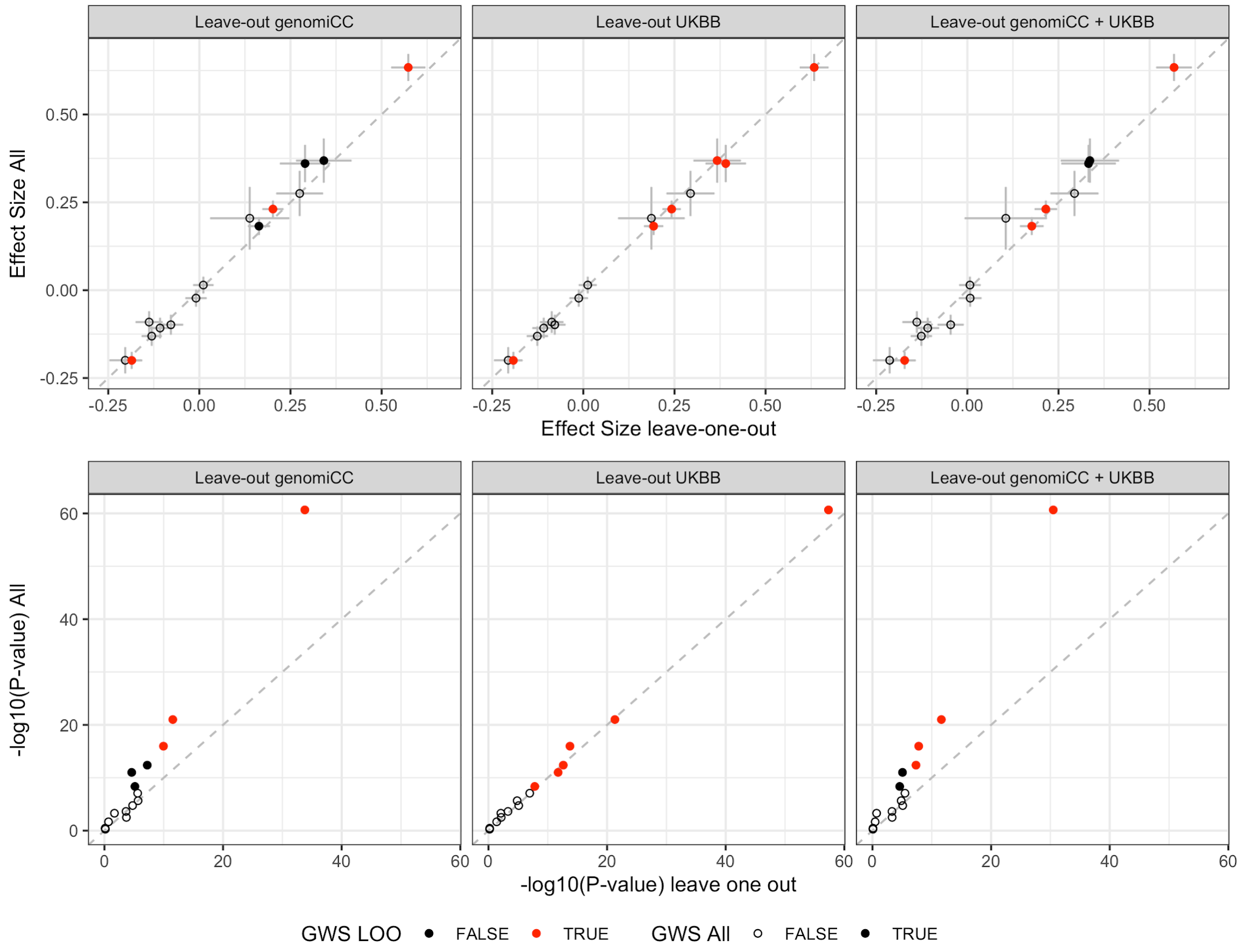


**Extended Data Figure 6. Sensitivity analyses for overlapping controls in genomiCC and UK Biobank.**

Comparison of the effect sizes and *P*-values of the 15 lead variant, using data from the COVID-19 critical illness meta-analysis in all the cohorts (y-axis) to leaving out genomiCC, leaving out UK Biobank (UKBB) and leaving out genomiCC + UKBB, respectively (x-axis). Dots represent the effect size estimates (top panels) and *P*-values (bottom panels), and bars represent the standard error. Filled dots indicate variants that were significant in the full meta-analysis of critical illness due to COVID-19, and empty dots represent variants that were not significant for critical illness but were significant for either hospitalization due to COVID-19 or SARS-CoV-2 reported infection. Red dots represent variants that were significant in leave-one-out analysis for genomiCC, UKBB or genomiCC + UKBB.

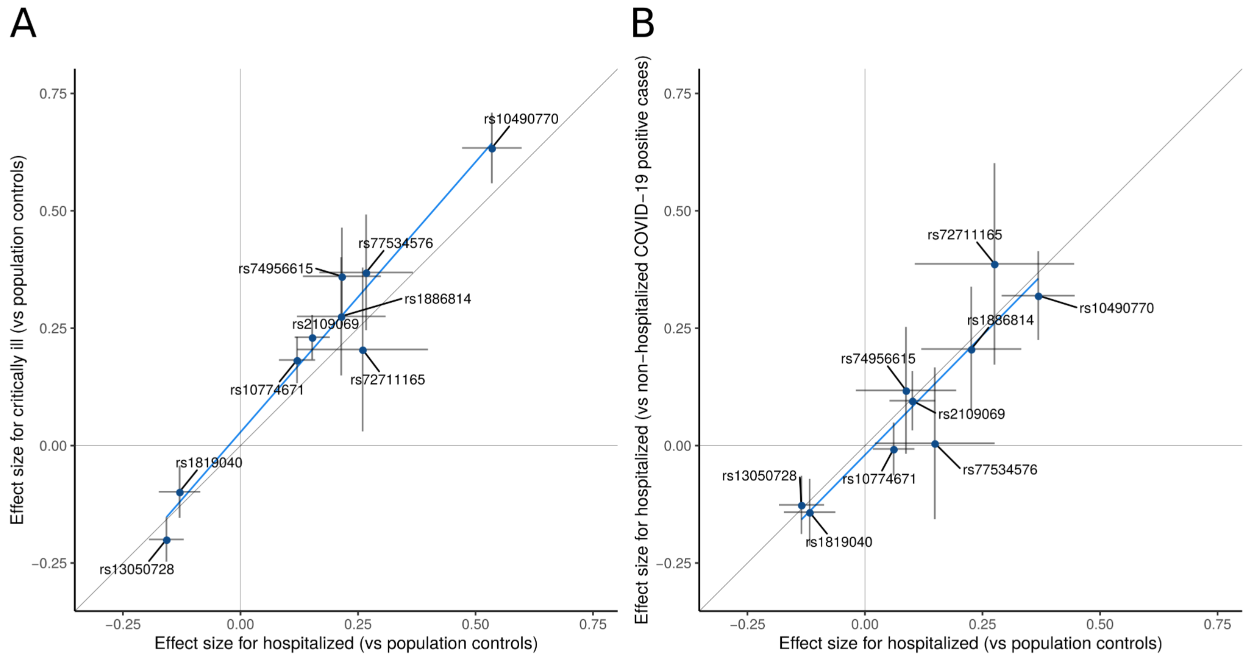


**Extended Data Figure 7. Comparison of lead variant effect sizes between pairs of COVID-19 meta-analyses.**

Comparison of effect sizes for the nine variants associated with severity of COVID-19 disease. A. Comparing hospitalized COVID-19 cases vs population controls (x-axis, n=10,428 cases and n=1,483,270 controls) and critically ill COVID-19 cases vs population controls (y-axis, n=6,179 cases and n=1,483,780 controls). B. hospitalized COVID-19 cases vs population controls (x-axis,n=5,806 cases and n=1,144,263 controls)and hospitalized COVID-19 cases vs non-hospitalized COVID-19 cases (y-axis, n=5,773 and n=15,497 controls). Sample sizes for hospitalized COVID-19 cases vs population controls differ between panels A and B due to difference in the sampling of studies selected for the analysis. This selection included all studies that were able to contribute data to the respective analysis that the data were compared to (on the y-axis) in each panel. Dots represent the effect size estimates, bars represent the confidence interval of the estimates. Effect size estimates and *P*-values for heterogeneity test are reported in Supplementary Table 3.

*Please find the corresponding figure in the PDF "Extended Data Figure 8"*

**Extended Data Figure 8. LocusZoom plots for each COVID-19 locus in three meta-analyses.**

For each genome-wide significant locus in three meta-analyses: critical illness (labelled as Analysis A2), hospitalization (labelled as Analysis B2), and reported infection (labelled as Analysis C2), we showed 1) a manhattan plot of each locus where a color represents a weighted-average $r^2$ value (see **Methods**) to a lead variant; 2) r2 values to a lead variant across gnomAD v2 populations, i.e., African/African-American (AFR), Latino/Admixed American (AMR), Ashkenazi Jewish (ASJ), East Asian (EAS), Estonian (EST), Finnish (FIN), Non-Finish Europeans (NFE), North-Western Europeans (NWE), and Southern Europeans (SEU); 3) genes at a locus; and 4) genes prioritized by each gene prioritization metric where a size of circles represents a rank in each metric. Note that the COVID-19 lead variants were chosen across all the meta-analyses (**Table 1**; see **Methods**) and were not necessarily a variant with the most significant *P*-value in each meta-analysis.

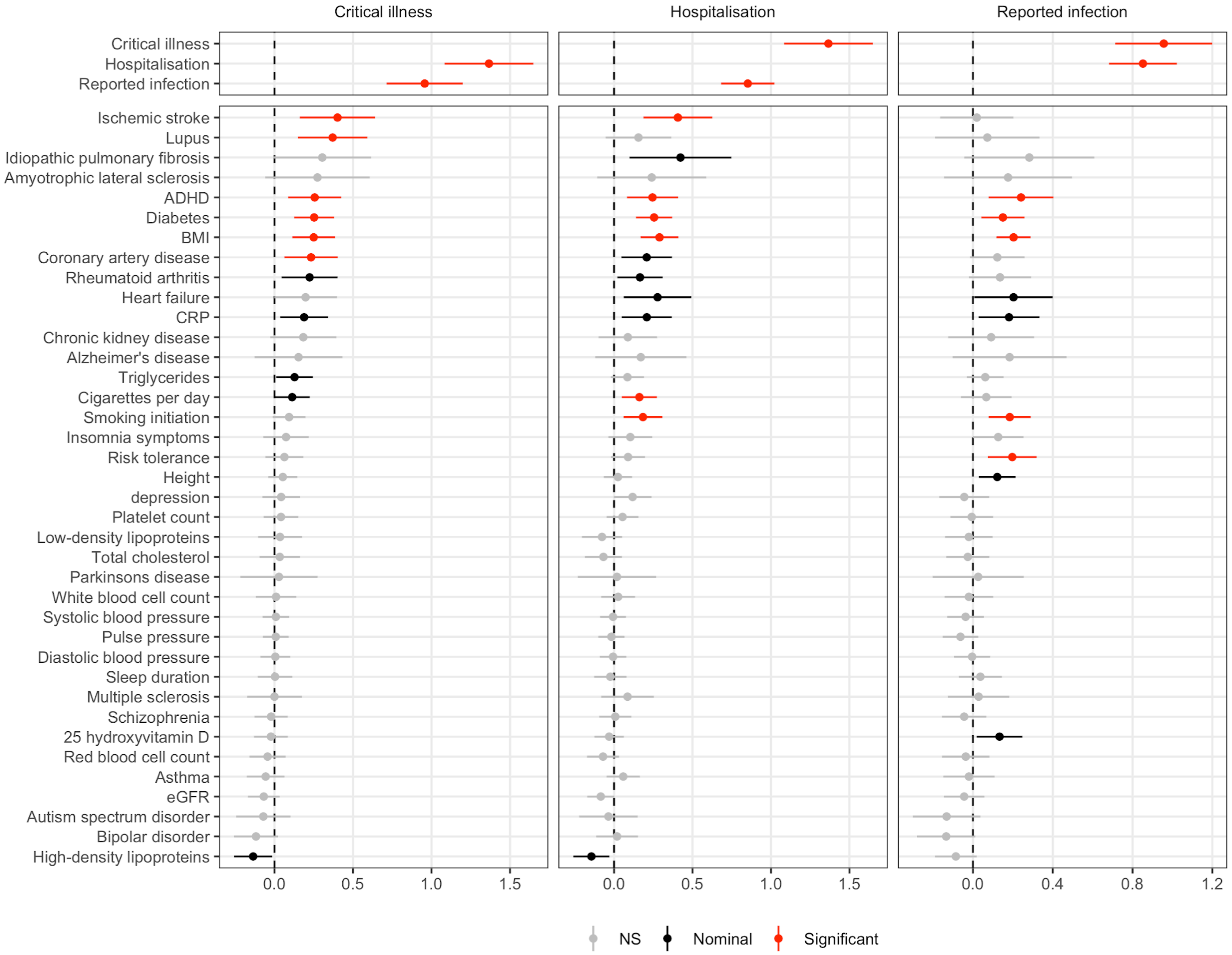


**Extended Data Figure 9. Genetic correlation results.**

Each column shows genetic correlation results for the three COVID-19 phenotypes (European ancestry analyses only): critical illness, hospitalization and reported infection. The traits the genetic correlation is run against are listed on the left. Significant correlations (FDR<0.05) are shown with their 95% confidence intervals in red, nominally significant (P<0.05) in black and non-significant in grey.

*Please find the figure in PDF "Extended Data Figure 10"*

**Extended Data Figure 10. Scatter and funnel plots for each for exposure - COVID-19 outcome pair.**

Scatter plots show the exposure variant effect size against the COVID-19 outcome variant effect size and corresponding standard errors. Funnel plots show the Mendelian randomization (MR) causal estimates for each variant against their precision, with asymmetry in the plot indicating potential violations of the assumptions of MR. Regression lines show the corresponding causal estimates fixed effect inverse-weighted (IVW, red-solid line) meta-analysis; MR-Egger regression (blue-dashed); Weighted median estimator (WME, green-dashed); weighted mode based estimator (WMBE, purple dashed); and Mendelian Randomization Pleiotropy RESidual Sum and Outlier corrected (MR-PRESSO, orange dashed). Variants highlighted in red were flagged as outliers by MR-PRESSO.

*Please find the corresponding figure in the PDF "Extended Data Figure 11"*

**Extended Data Figure 11. Standard error as a function of effective sample size for participating studies.**

The figure panels show the standard error plotted as a function of effective sample size in each study that contributed data to the meta-analysis of the three phenotypes (from the left): critical illness due to COVID-19, hospitalization, and reported infection. Because allele frequency can impact the relationship between standard error and effective sample size, we have also colored the different studies by ancestry group. These plots were used in initial phases of quality control before meta-analysing studies. Outlier studies were identified by eye and removed from the meta-analysis until the issues were fixed, e.g. erroneous reporting of standard errors. These issues were reported to the respective investigators for follow-up.

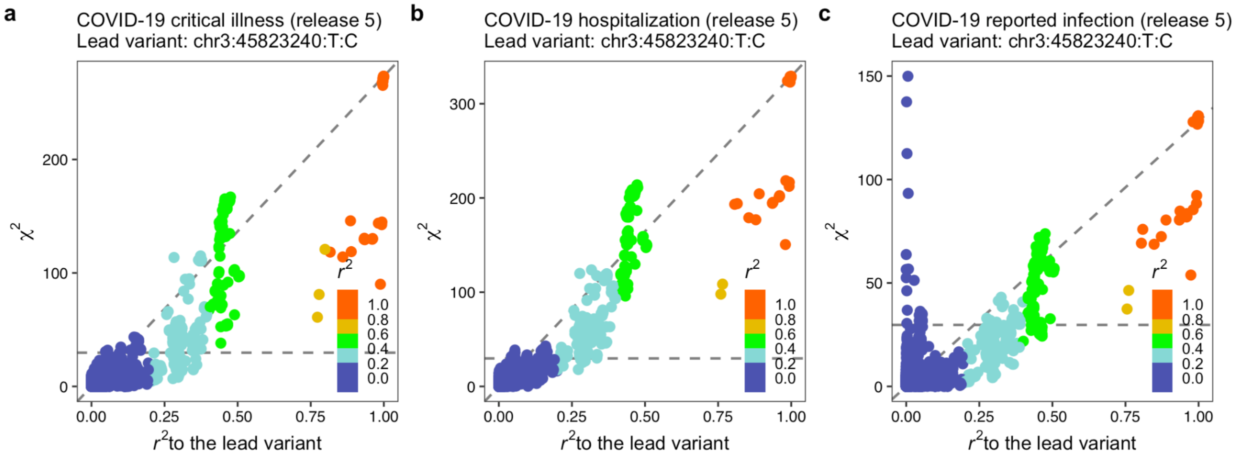


**Extended Data Figure 12. Comparison of chi-squared statistics vs r2 values to the lead variant in the 3p21.31 region**.

For **a**. critical illness **b**. hospitalization, and **c**. reported infection. The left blue peak in panel **c**, which is uncorrelated with the lead variants in the region, indicates that there are independent signals.
