## Extended data and supplement for "Mapping the human genetic architecture of COVID-19 by worldwide meta-analysis": revised_manuscript_COVID19-HGI_Flagship.pdf

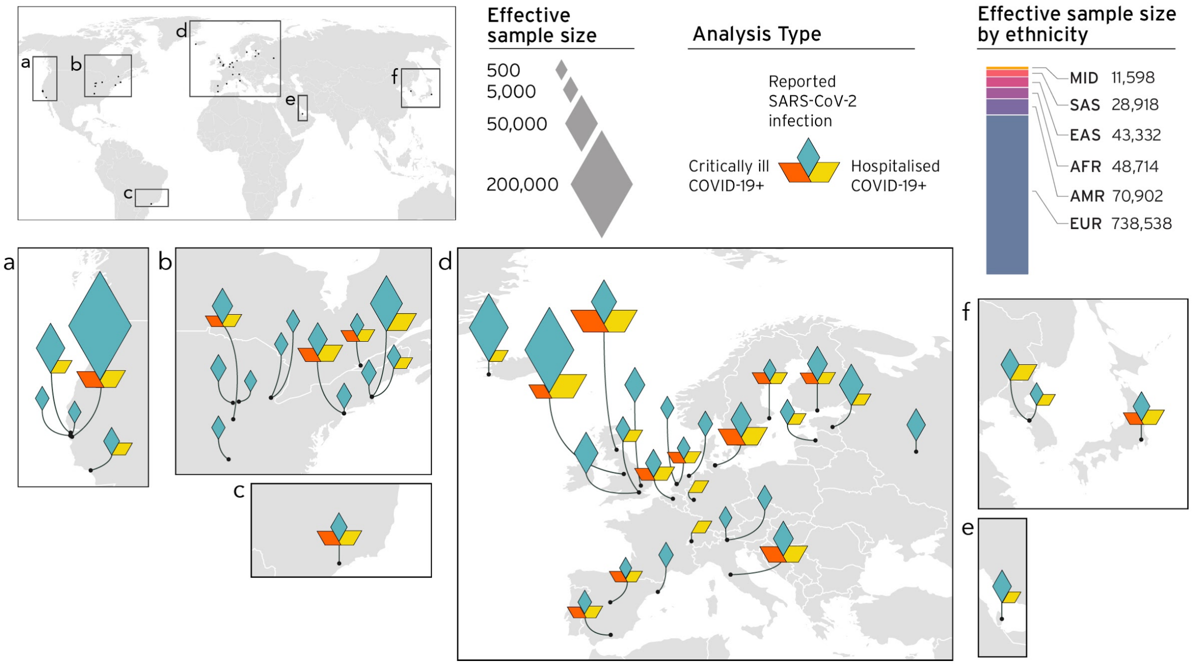


***Figure 1.*** *Geographical overview of the contributing studies to the COVID-19 HGI and composition by major ancestry groups. Middle Eastern (MID), South Asian (SAS), East Asian (EAS), African (AFR), Admixed American (AMR), European (EUR).*

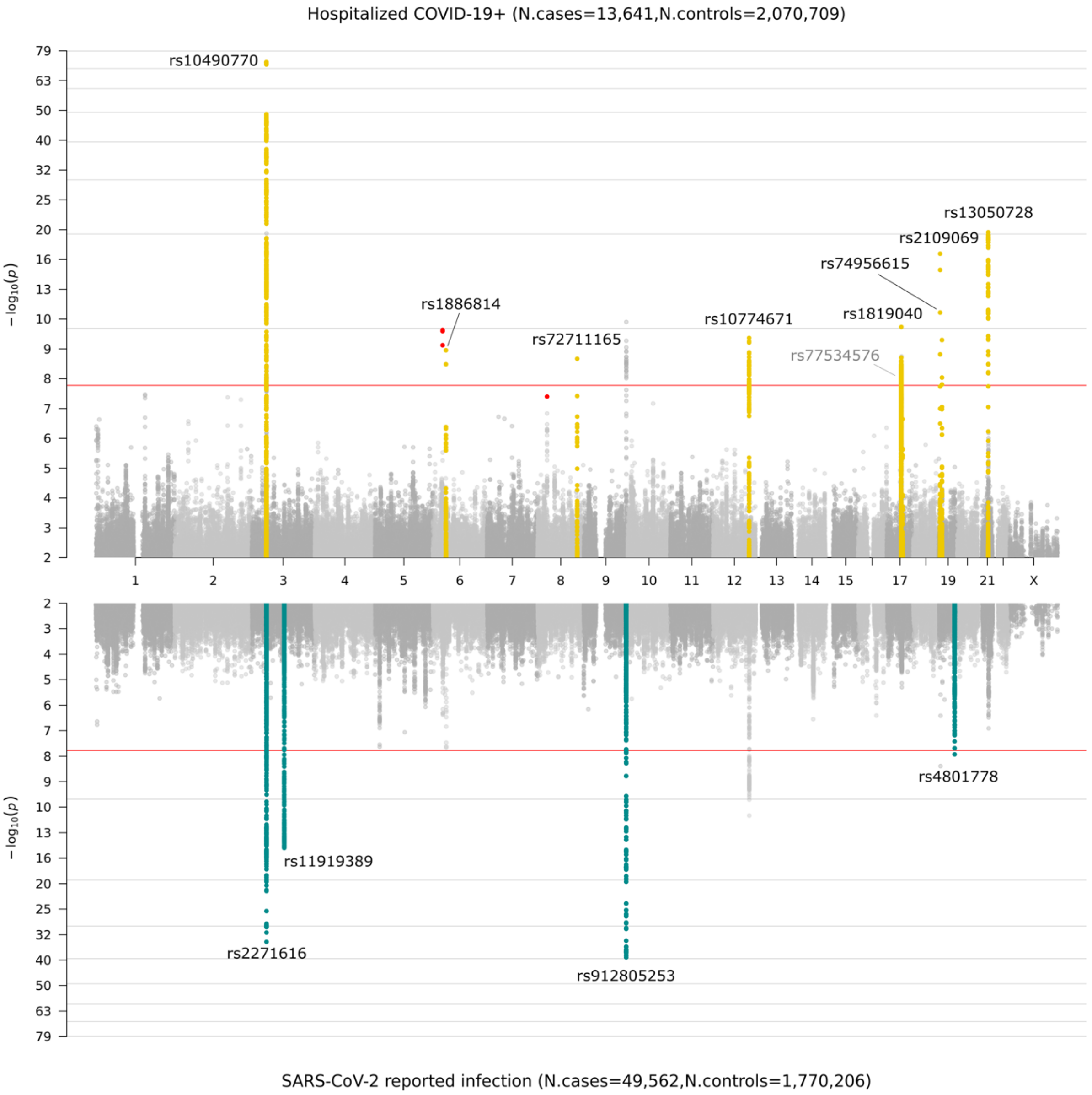


***Figure 2. Genome-wide association results for COVID-19.*** *Top panel shows results of genome-wide association study of hospitalized COVID-19 and controls, and bottom panel the results of reported SARS-CoV-2 infection and controls. Loci highlighted in yellow (top panel) represent regions associated with severity of COVID-19 manifestation i.e. increasing odds for more severe COVID-19 phenotypes, where loci highlighted in green (bottom panel) are regions associated with reported SARS-CoV-2 infection, i.e. the effect is the same across mild and severe COVID-19 phenotypes. A window of +/- 500kb region from the lead variant (**Table 1**) was used to highlight these loci. We highlight only loci that were specific to each analysis. That is, some genome-wide significant loci are not colored in yellow or green if they are not specific to severity (top panel) or susceptibility (bottom panel). Variants highlighted in red represent genome-wide significant variants that had high heterogeneity across studies that contributed data, and which were therefore not considered in the final analyses. Note that top panel regions on chromosomes 17 and 19 contain two statistically independent loci within proximity of each other, and on the bottom panel*

*the first signal on chromosome 3 contains two peaks within the same region. The locus tagged by lead variant rs77534576 on chromosome 17 was discovered in the critically ill COVID-19 analysis (**see Table 1**).*

Our phenotype definitions include population controls without known SARS-CoV-2 infection. This is not an optimal control group because some individuals, if exposed to SARS-CoV-2 could develop a severe form of COVID-19 disease and should be classified as cases. To better understand the effect of such potential misclassification, we conducted a new meta-analysis, including only the studies that compared hospitalized COVID-19 cases with controls with laboratory-confirmed SARS-CoV-2 infection but who had mild symptoms or were asymptomatic ($n$ = 5,773 cases and $n$ = 15,497 controls). We then compared the effect sizes obtained from this analysis with those from the main phenotype definition (hospitalized cases *vs.* controls without known SARS-CoV-2 infection, if that information was available) using only studies that reported results for both analyses (**Methods**). We found that across the nine loci that had reached genome-wide significance in our main hospitalized COVID-19 analysis, the ORs were not significantly different in the analysis with better refined controls (**Extended Data Fig. 7B, Supplementary Table 3**). These results indicate that using population controls can be a valid and powerful strategy for host genetic discovery of infectious disease.

| rsid | Chr:pos (b38) | Ref allele | Effect allele | Effect allele frequency | COVID-19 phenotype | OR | CI lower | CI upper | *P*-value (association) | *P*-value (het) | Suggested phenotypic impact | Genes in LD region (closest gene in bold) | Genes with coding variants | eGenes |
|---|---|---|---|---|---|---|---|---|---|---|---|---|---|---|
| rs2271616 | 3:45796521 | G | T | 0.119 | Critical illness | 1.14 | 1.05 | 1.23 | 1.91E-03 | 0.156 | infection susceptibility | ***SLC6A20*** | ***SLC6A20*** | |
| | | | | 0.117 | Hospitalized | 1.12 | 1.07 | 1.18 | 1.05E-05 | 0.159 | | | | |
| | | | | 0.118 | Reported infection | 1.15 | 1.13 | 1.18 | 1.79E-34 | 0.306 | | | | |
| rs10490770 | 3:45823240 | T | C | 0.075 | Critical illness | 1.89 | 1.75 | 2.03 | 2.20E-61 | 0.051 | disease severity | ***LZTFL1*** | | |
| | | | | 0.081 | Hospitalized | 1.65 | 1.56 | 1.74 | 1.44E-73 | 2.09E-03 | | | | |
| | | | | 0.085 | Reported infection | 1.16 | 1.13 | 1.19 | 9.72E-30 | 7.24E-25 | | | | |
| rs11919389 | 3:101705614 | T | C | 0.344 | Critical illness | 0.98 | 0.93 | 1.03 | 3.51E-01 | 0.082 | infection susceptibility | *ZBTB11*, ***RPL24***, *CEP97*, *NXPE3* | | |
| | | | | 0.348 | Hospitalized | 0.95 | 0.92 | 0.98 | 8.13E-04 | 0.304 | | | | |
| | | | | 0.352 | Reported infection | 0.94 | 0.93 | 0.96 | 3.46E-15 | 0.714 | | | | |
| rs1886814 | 6:41534945 | A | C | 0.038 | Critical illness | 1.32 | 1.16 | 1.49 | 1.79E-05 | 0.815 | disease severity | ***FOXP4*** | | *FOXP4* |
| | | | | 0.042 | Hospitalized | 1.26 | 1.17 | 1.36 | 1.11E-09 | 0.348 | | | | |
| | | | | 0.047 | Reported infection | 1.11 | 1.07 | 1.15 | 2.41E-08 | 0.216 | | | | |
| rs72711165 | 8:124324323 | T | C | 0.011 | Critical illness | 1.23 | 1.03 | 1.46 | 2.14E-02 | 0.267 | disease severity | ***TMEM65*** | | |
| | | | | 0.013 | Hospitalized | 1.37 | 1.24 | 1.52 | 2.13E-09 | 0.551 | | | | |
| | | | | 0.018 | Reported infection | 1.08 | 1.02 | 1.14 | 9.43E-03 | 0.177 | | | | |
| rs912805253 | 9:133274084 | C | T | 0.653 | Critical illness | 0.88 | 0.83 | 0.93 | 2.16E-06 | 0.008 | infection susceptibility | ***ABO*** | *ABO* | *ABO* |
| | | | | 0.655 | Hospitalized | 0.90 | 0.87 | 0.93 | 5.37E-10 | 0.007 | | | | |
| | | | | 0.651 | Reported infection | 0.91 | 0.89 | 0.92 | 1.45E-39 | 0.014 | | | | |
| rs10774671 | 12:112919388 | G | A | 0.652 | Critical illness | 1.20 | 1.14 | 1.26 | 4.08E-13 | 0.923 | disease severity | ***OAS1***, *OAS3*, *OAS2* | *OAS1,OAS3* | |
| | | | | 0.664 | Hospitalized | 1.11 | 1.07 | 1.14 | 6.14E-10 | 0.550 | | | | |
| | | | | 0.669 | Reported infection | 1.06 | 1.04 | 1.08 | 1.61E-11 | 0.200 | | | | |
| rs1819040 | 17:46142465 | T | A | 0.197 | Critical illness | 0.91 | 0.86 | 0.96 | 5.02E-04 | 0.076 | disease severity | *ARHGAP27*, *PLEKHM1*, *LINC02210-CRHR1*, *CRHR1*, *SPPL2C*, *MAPT*, *STH*, ***KANSL1***, *LRRC37A*, *ARL17B*, *LRRC37A2*, *ARL17A*, *NSF*, *WNT3* | *ARHGAP27*, *KANSL1*, *MAPT*, *SPPL2C*, *STH* | *ARL17B*, *KANSL1*, *MAPT*, *WNT3* |
| | | | | 0.186 | Hospitalized | 0.88 | 0.84 | 0.91 | 1.83E-10 | 0.327 | | | | |
| | | | | 0.184 | Reported infection | 0.96 | 0.94 | 0.98 | 5.05E-06 | 0.828 | | | | |
| rs77534576 | 17:49863303 | C | T | 0.033 | Critical illness | 1.45 | 1.28 | 1.64 | 4.37E-09 | 0.508 | disease severity | *KAT7*, ***TAC4*** | | *DLX3* |
| | | | | 0.033 | Hospitalized | 1.26 | 1.15 | 1.37 | 2.26E-07 | 0.009 | | | | |
| | | | | 0.037 | Reported infection | 1.08 | 1.04 | 1.13 | 1.08E-04 | 0.016 | | | | |
| rs2109069 | 19:4719431 | G | A | 0.316 | Critical illness | 1.26 | 1.20 | 1.32 | 9.68E-22 | 0.318 | disease severity | ***DPP9*** | | |
| | | | | 0.312 | Hospitalized | 1.15 | 1.12 | 1.19 | 2.76E-17 | 0.157 | | | | |
| | | | | 0.315 | Reported infection | 1.05 | 1.03 | 1.07 | 4.08E-09 | 6.71E-05 | | | | |
| rs74956615 | 19:10317045 | T | A | 0.048 | Critical illness | 1.43 | 1.29 | 1.59 | 9.71E-12 | 0.223 | disease severity | *ICAM1*, *ICAM4*, *ICAM5*, *ZGLP1*, *FDX2*, ***RAVER1***, *ICAM3*, *TYK2* | *TYK2* | |
| | | | | 0.048 | Hospitalized | 1.27 | 1.18 | 1.36 | 5.05E-10 | 1.94E-04 | | | | |
| | | | | 0.052 | Reported infection | 1.06 | 1.02 | 1.09 | 3.68E-03 | 0.002 | | | | |
| rs4801778 | 19:48867352 | G | T | 0.176 | Critical illness | 0.91 | 0.86 | 0.97 | 3.12E-03 | 0.579 | infection susceptibility | ***PLEKHA4***, *PPP1R15A*, *TULP2*, *NUCB1* | *PPP1R15A* | |
| | | | | 0.176 | Hospitalized | 0.96 | 0.92 | 1.00 | 2.90E-02 | 0.908 | | | | |
| | | | | 0.180 | Reported infection | 0.95 | 0.93 | 0.96 | 1.18E-08 | 0.901 | | | | |
| rs13050728 | 21:33242905 | T | C | 0.662 | Critical illness | 0.82 | 0.78 | 0.86 | 1.05E-16 | 0.642 | disease severity | ***IFNAR2*** | | *IFNAR2* |
| | | | | 0.651 | Hospitalized | 0.86 | 0.83 | 0.89 | 2.72E-20 | 0.194 | | | | |
| | | | | 0.651 | Reported infection | 0.97 | 0.95 | 0.98 | 1.28E-05 | 0.001 | | | | |

***Table 1: Genome-wide significant results for each worldwide meta-analysis.***

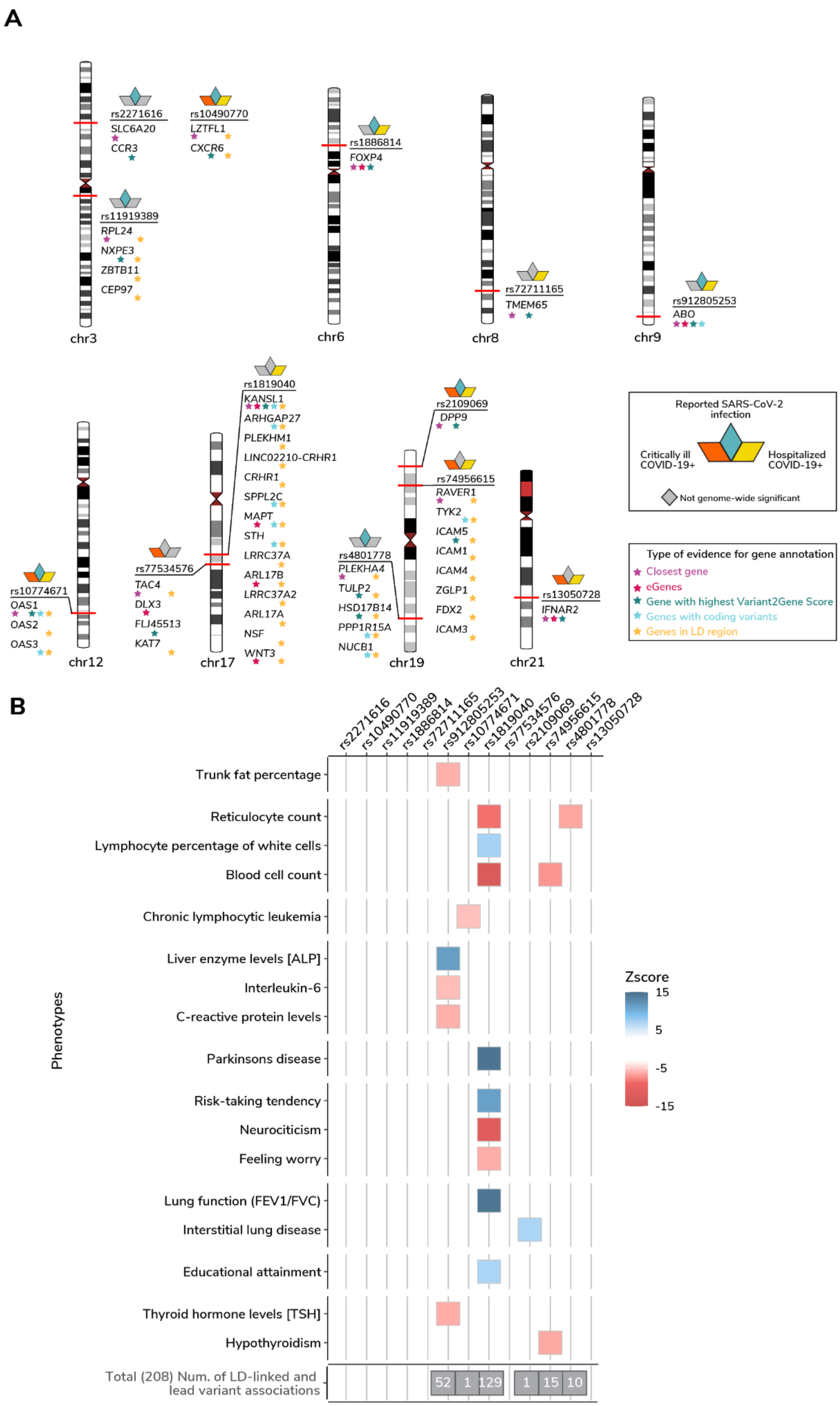
A
rs2271616
SLC6A20
CCR3
rs10490770
LZTFL1
CXCR6
rs11919389
RPL24
NXPE3
ZBTB11
CEP97
chr3
rs1886814
FOXP4
chr6
rs72711165
TMEM65
chr8
rs912805253
ABO
chr9
rs10774671
OAS1
OAS2
OAS3
chr12
rs77534576
TAC4
DLX3
FLJ45513
KAT7
rs1819040
KANSL1
ARHGAP27
PLEKHM1
LINC02210-CRHR1
CRHR1
SPPL2C
MAPT
STH
LRRC37A
ARL17B
LRRC37A2
ARL17A
NSF
WNT3
chr17
rs4801778
PLEKHA4
TULP2
HSD17B14
PPP1R15A
NUCB1
rs2109069
DPP9
rs74956615
RAVER1
TYK2
ICAM5
ICAM1
ICAM4
ZGLP1
FDX2
ICAM3
chr19
rs13050728
IFNAR2
chr21
Reported SARS-CoV-2 infection
Critically ill COVID-19+
Hospitalized COVID-19+
Not genome-wide significant
Type of evidence for gene annotation
Closest gene
eGenes
Gene with highest Variant2Gene Score
Genes with coding variants
Genes in LD region
B
rs2271616
rs10490770
rs11919389
rs1886814
rs72711165
rs912805253
rs10774671
rs1819040
rs77534576
rs2109069
rs74956615
rs4801778
rs13050728
Phenotypes
Trunk fat percentage
Reticulocyte count
Lymphocyte percentage of white cells
Blood cell count
Chronic lymphocytic leukemia
Liver enzyme levels [ALP]
Interleukin-6
C-reactive protein levels
Parkinsons disease
Risk-taking tendency
Neurociticism
Feeling worry
Lung function (FEV1/FVC)
Interstitial lung disease
Educational attainment
Thyroid hormone levels [TSH]
Hypothyroidism
Total (208) Num. of LD-linked and lead variant associations
52
1
129
1
15
10
Zscore
15
5
-5
-15

***Figure 3.*** **Gene prioritization and PheWas.** ***A)*** *Gene prioritization using different evidence measures of gene annotation. The ideogram shows the position of the COVID-19 index SNPs and associated genes with type of gene annotation highlighted as stars. The squares above the SNP rsid represent COVID-19 sub-phenotypes for which the SNP is genome-wide significant. Closest gene: A gene with a minimum distance from each lead variant to gene body. Genes in linkage disequilibrium (LD) region: Genes that overlap with a genomic range that contains any variants in LD ($r^2 > 0.6$) with each lead variant. Genes with coding variants: Genes with a loss-of-function or missense variant in LD with a lead variant ($r^2 > 0.6$). eGenes: Genes with a fine-mapped cis-eQTL variant (PIP > 0.1) in GTEx Lung that is in LD with a lead variant ($r^2 > 0.6$) (see* ***Supplementary Table 4*** *for a complete list). V2G: Highest gene prioritized by OpenTargetGenetics' V2G score.* ***B)*** *Selected phenotypes associated with genome-wide significant COVID-19 variants (see* ***Supplementary Table 5*** *for a complete list). We report those associations for which a lead variant from a prior GWAS results was in high LD ($r^2 > 0.8$) with the index COVID-19 variants. The colour represents the Z-scores of correlated risk increasing alleles for the trait. The total number of associations for each COVID-19 variant is highlighted in the grey box.*

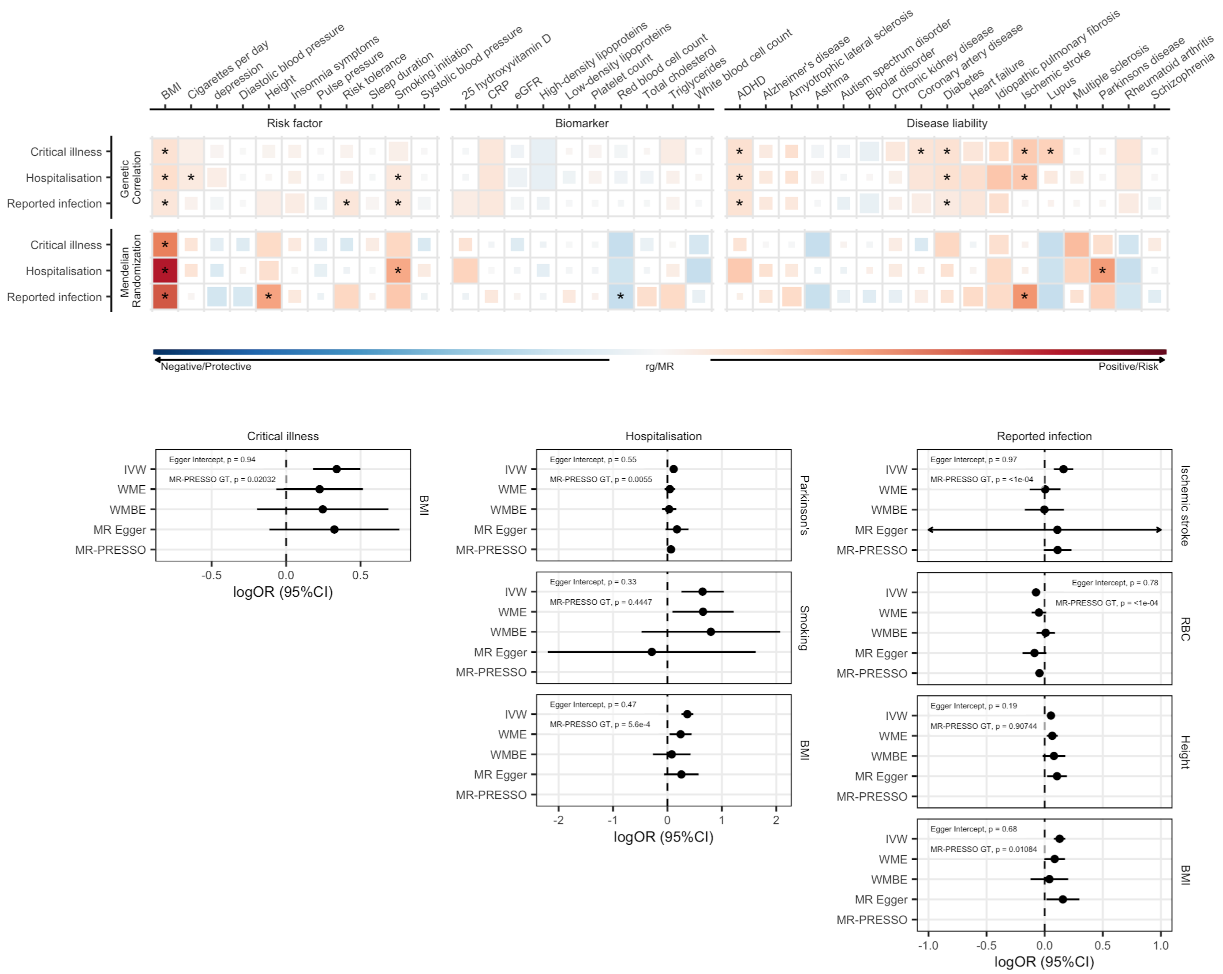


***Figure 4.*** *Genetic correlations and Mendelian randomization causal estimates between 38 traits and COVID-19 hospitalization, critical illness and SARS-CoV-2 reported infection. Blue, negative genetic correlation and protective Mendelian randomization (MR) causal estimates; red, positive genetic correlation and risk MR causal estimates. Larger squares correspond to more significant P values, with genetic correlations or MR causal estimates significantly different from zero at a P < 0.05 shown as a full-sized square. Genetic correlations or causal estimates that are significantly different from zero at a false discovery rate (FDR) of 5% are marked with an asterisk. Forest plots display the causal estimates for each of the sensitivity analyses used in the MR analysis for trait pairs that were significant at an FDR of 5%. Individual scatter and funnel plots for each pair of traits are available in* ***Extended Data Fig. 10****.*
*IVW: Inverse variance weighted analysis; WME: Weighted median estimator; WMBE: weighted mode based estimator; MR-PRESSO: Mendelian Randomization Pleiotropy RESidual Sum and Outlier. RBC: Red blood cell count*
