## Extended data and supplement for "Mapping the human genetic architecture of COVID-19 by worldwide meta-analysis": supplementary_note_COVID19-HGI.pdf

**Additional independent susceptibility signals at the 3p21.31 locus**

To identify loci that potentially have multiple causal variants, we compared marginal chi-squared association statistics and r2 values to the lead variant for each locus. In theory, marginal chi-squared statistics should correlate with r2 values to a lead variant (that is tagged by a single causal variant), and any deviation might be a sign of additional causal variants in a locus, or existence of missing causal variant(s) that are not properly tagged by a lead variant. We found the 3p21.31 locus has multiple independent signals only for reported infection (**Supplementary Fig 12**), and confirmed an additional susceptibility lead variant rs2271616:G>T is independent from the severity lead variant rs10490770:T>C ($r^2$ = 0.0).

Unfortunately, we found that a proper statistical fine-mapping or conditional analysis is extremely challenging for our current meta-analysis due to the concerns over inter-cohort heterogeneity (difference in phenotyping, genotyping, and imputation, etc) and a lack of appropriate in-sample LD reference. Instead, we took a closer look at rs2271616:G>T and its proximal variants rs2271615:G>C (r2 = 0.79 to rs2271616), rs73062389:G>A (r2 = 0.34), and rs73062394:A>T (r2 = 0.30); We observed a clear deviation from an expected relationship between their chi-squared statistics and r2 values to rs2271616, suggesting that there might be missing causal variants or complex structural variants in this region. We note that this deviation holds true even when restricting to the studies that have all the four variants available.

Given the potential sign of missing causal variants in the region, we caution further disentangling the observed susceptibility association with the present result. Indeed, by using the latest release of UK Biobank (number of cases = 13,256 in European samples for reported infection; released Apr 9, 2021), we observed rs73062389:G>A showed more significant association ($P = 2.2 \times 10^{-13}$) than the current lead variant rs2271616:G>T ($P = 2.4 \times 10^{-12}$) (unpublished data), which re-emphasizes the crucial needs to collect more data to dissect this signal.
