## Supplementary figures and images for "Mapping the human genetic architecture of COVID-19 by worldwide meta-analysis"

### Extended_Data_Figure_11.pdf

# 3:45947552:T:G

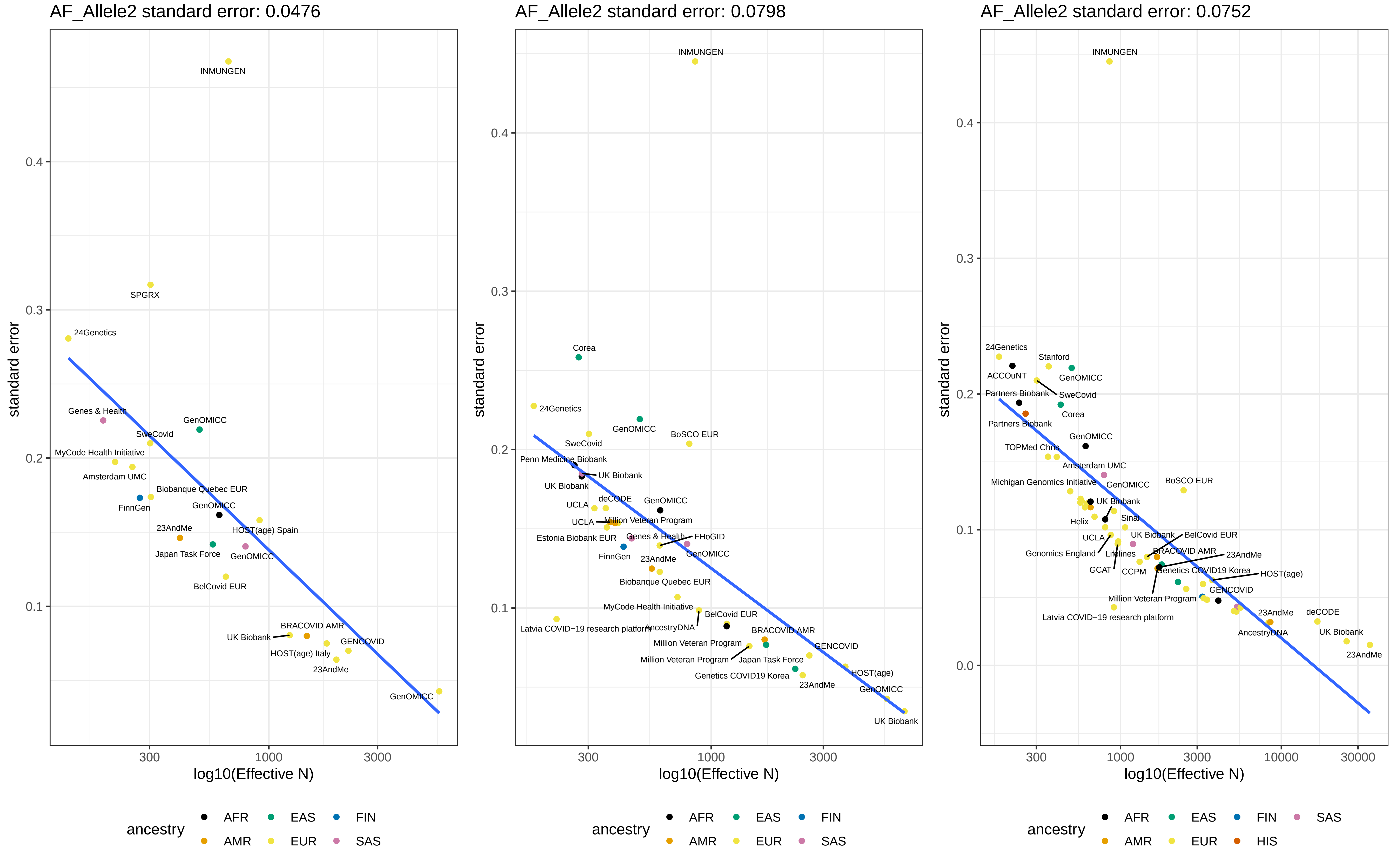

# 3:45933601:A:C

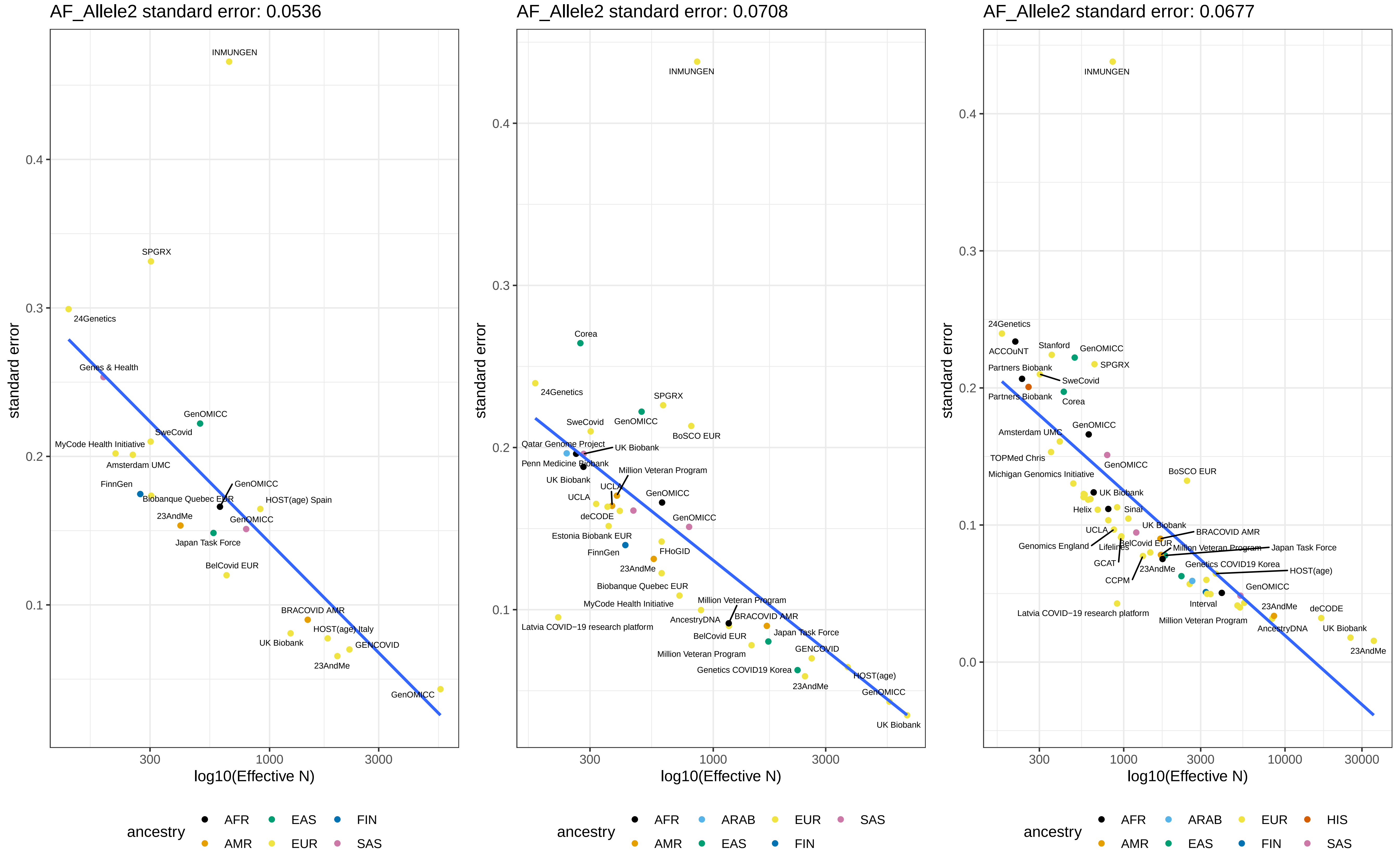

# 9:133273813:C:T

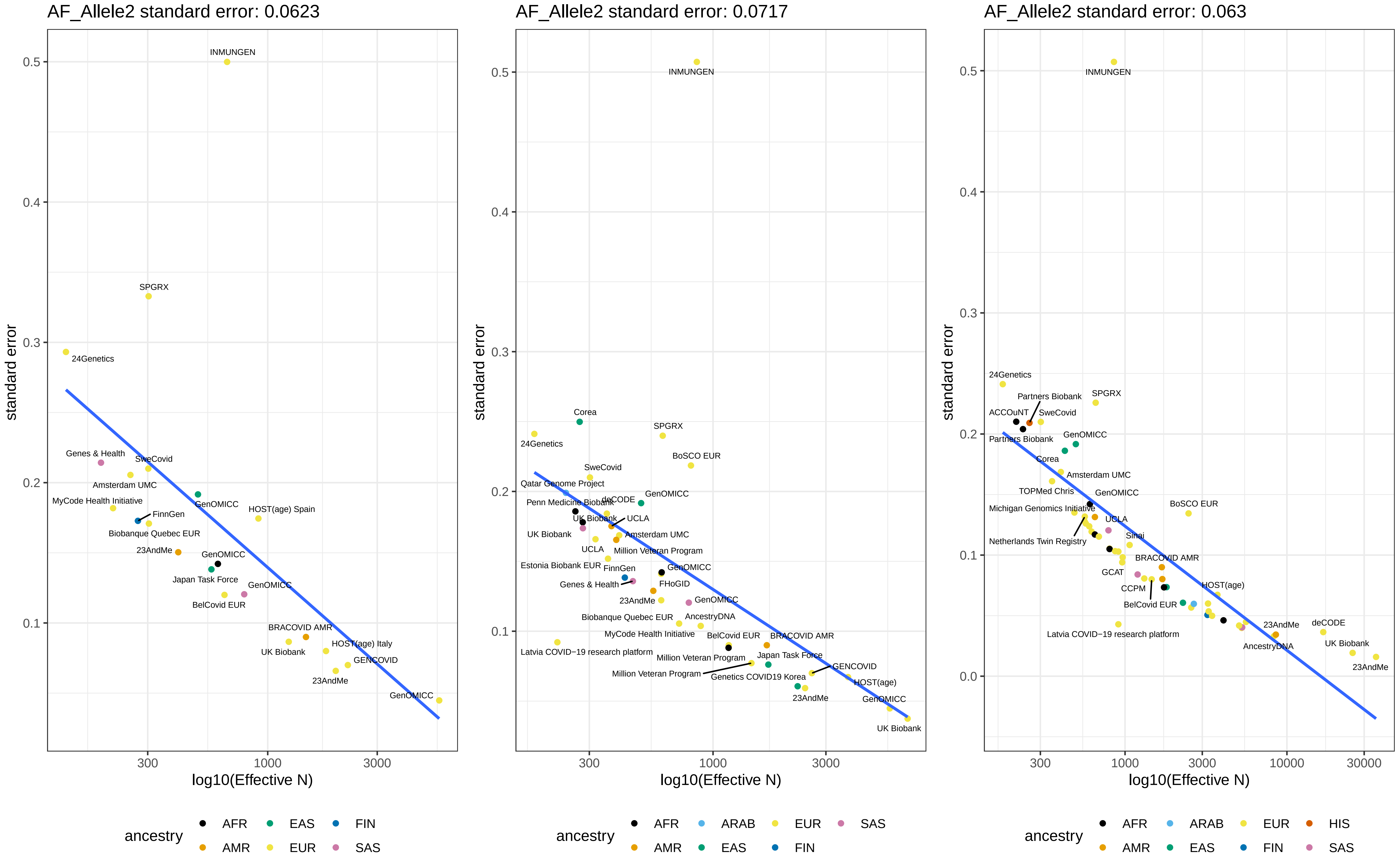

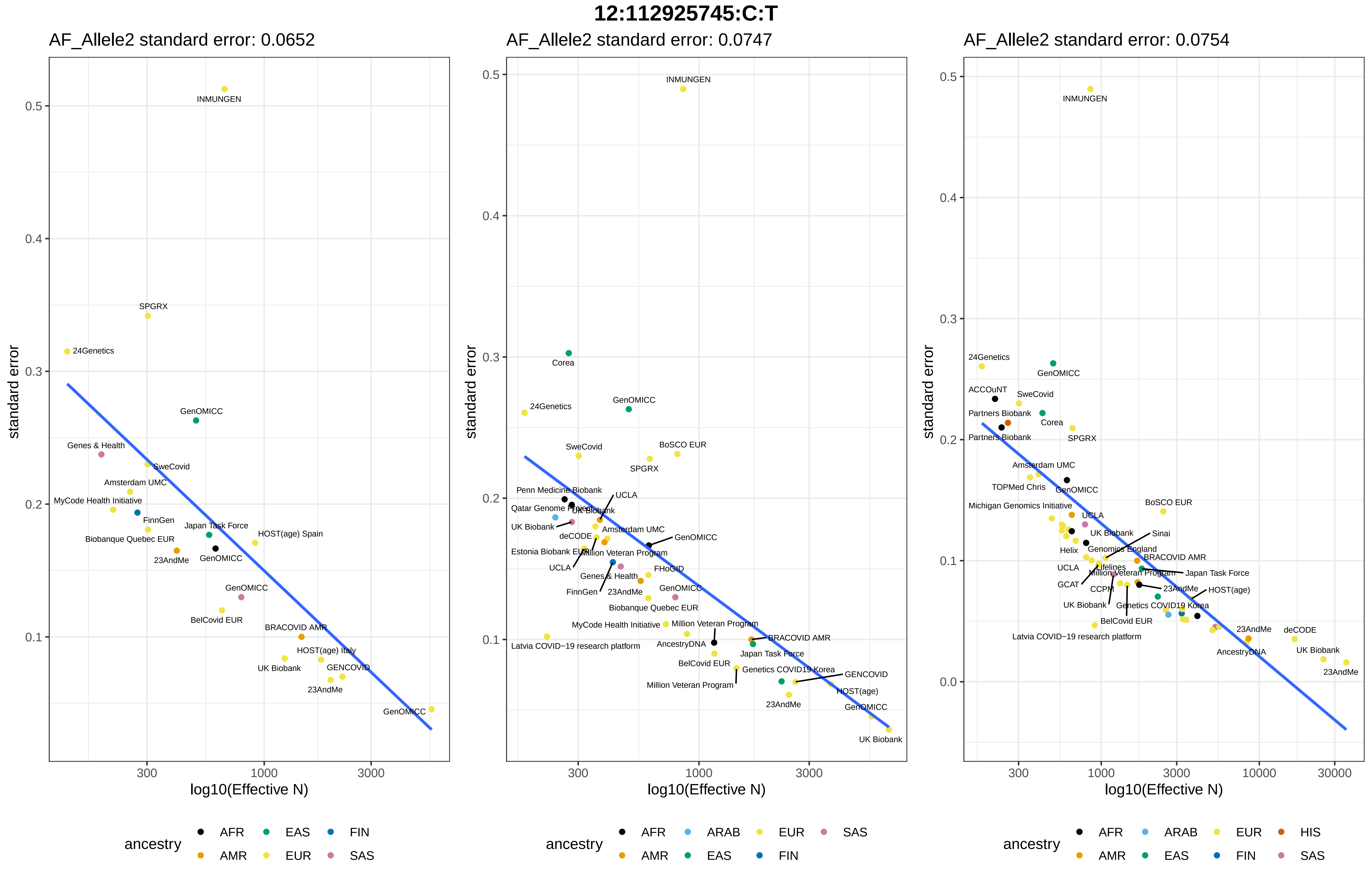

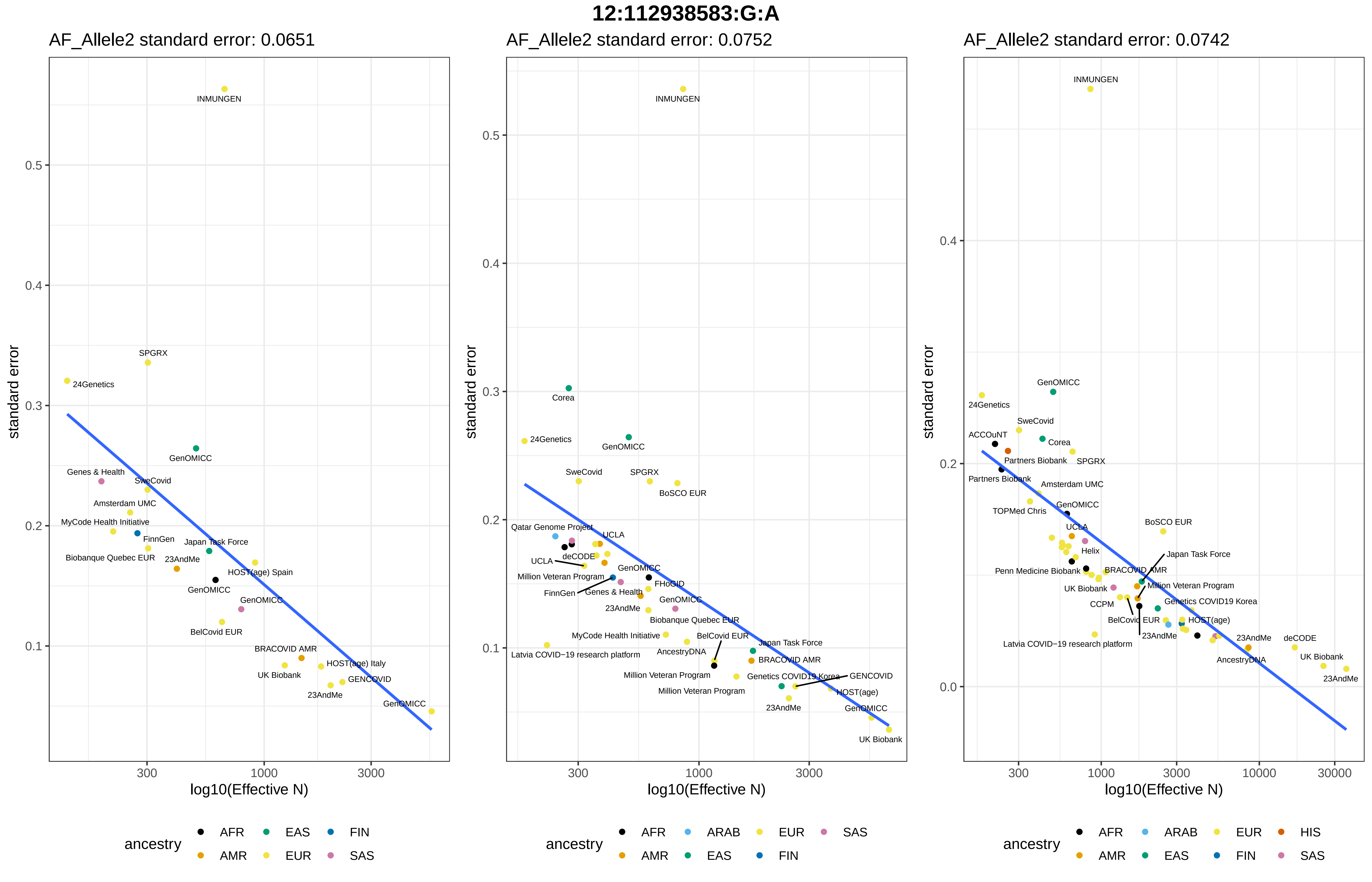

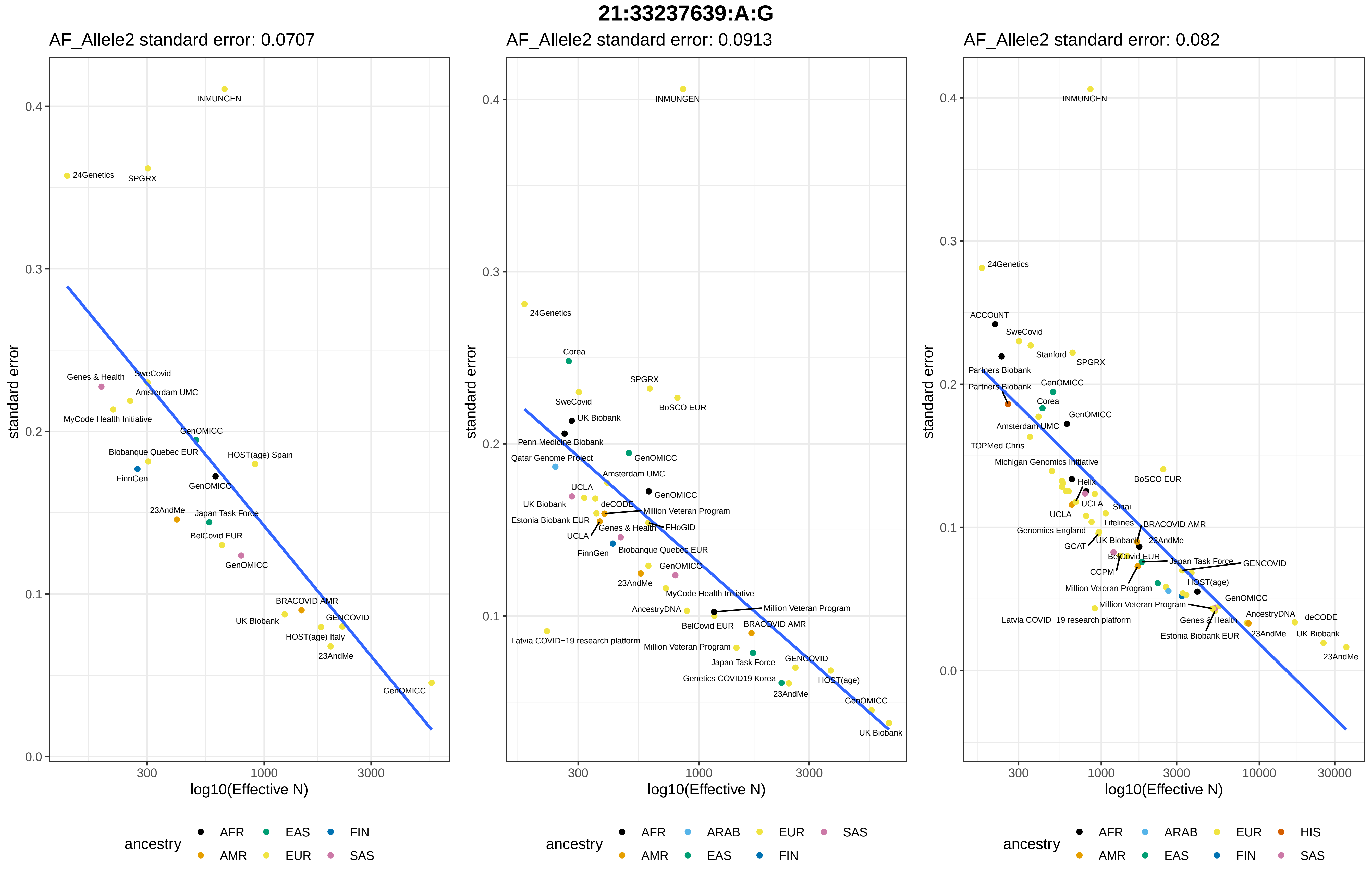

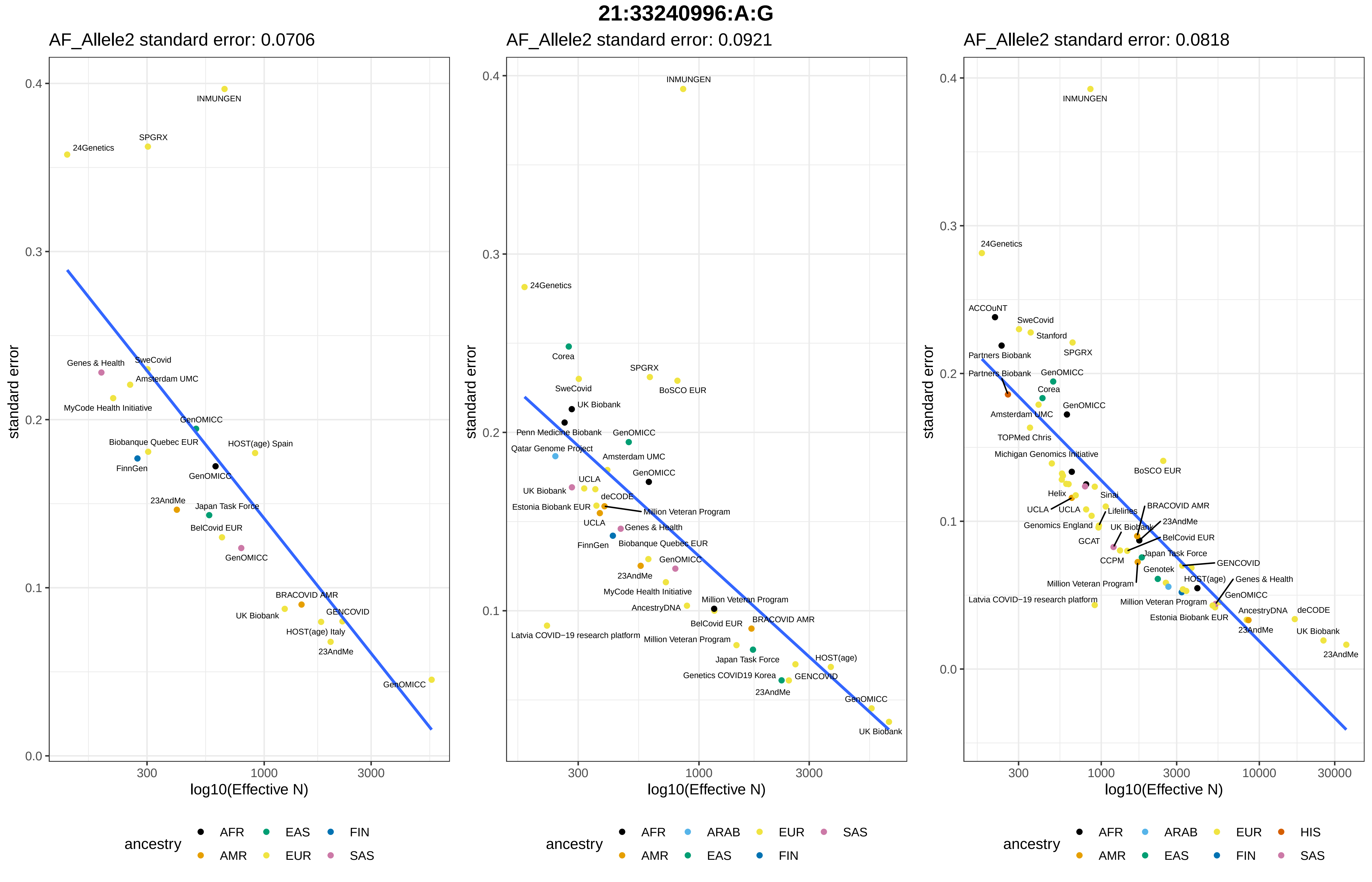
